# A Large-Scale Proteomics Resource of Circulating Extracellular Vesicles for Biomarker Discovery in Pancreatic Cancer

**DOI:** 10.1101/2023.03.13.23287216

**Authors:** Bruno Bockorny, Lakshmi Muthuswamy, Ling Huang, Marco Hadisurya, Christine Maria Lim, Leo L. Tsai, Ritu R. Gill, Jesse L. Wei, Andrea J. Bullock, Joseph E. Grossman, Robert J. Besaw, Supraja Narasimhan, W. Andy Tao, Sofia Perea, Mandeep S. Sawhney, Steven D. Freedman, Manuel Hidalgo, Anton Iliuk, Senthil K. Muthuswamy

## Abstract

Pancreatic cancer has the worst prognosis of all common tumors. Earlier cancer diagnosis could increase survival rates and better assessment of metastatic disease could improve patient care. As such, there is an urgent need to develop biomarkers to diagnose this deadly malignancy. Analyzing circulating extracellular vesicles (cEVs) using ‘liquid biopsies’ offers an attractive approach to diagnose and monitor disease status. However, it is important to differentiate EV-associated proteins enriched in patients with pancreatic ductal adenocarcinoma (PDAC) from those with benign pancreatic diseases such as chronic pancreatitis and intraductal papillary mucinous neoplasm (IPMN). To meet this need, we combined the novel EVtrap method for highly efficient isolation of EVs from plasma and conducted proteomics analysis of samples from 124 individuals, including patients with PDAC, benign pancreatic diseases and controls. On average, 912 EV proteins were identified per 100µL of plasma. EVs containing high levels of PDCD6IP, SERPINA12 and RUVBL2 were associated with PDAC compared to the benign diseases in both discovery and validation cohorts. EVs with PSMB4, RUVBL2 and ANKAR were associated with metastasis, and those with CRP, RALB and CD55 correlated with poor clinical prognosis. Finally, we validated a 7-EV protein PDAC signature against a background of benign pancreatic diseases that yielded an 89% prediction accuracy for the diagnosis of PDAC. To our knowledge, our study represents the largest proteomics profiling of circulating EVs ever conducted in pancreatic cancer and provides a valuable open-source atlas to the scientific community with a comprehensive catalogue of novel cEVs that may assist in the development of biomarkers and improve the outcomes of patients with PDAC.

## INTRODUCTION

Pancreatic ductal adenocarcinoma (PDAC) has the worst prognosis of all common tumors, with a 5-year survival of 10%^1^. With rising incidence, it is expected that PDAC will become the second leading cause of cancer-related deaths by 2030^2^. A critical factor for this dismal development is the late diagnosis, with less than 20% of patients presenting with a potentially resectable and curable tumor^3-5^. Earlier cancer diagnosis could increase the survival rates by an estimated 5-fold, and more reliable and real-time assessment of treatment effects in patients with cancer could improve quality of life and reduce healthcare costs^6,7^. Unfortunately, there are no credentialed serologic biomarkers with high enough performance to assist in the early detection of PDAC. The best-established biomarker for PDAC, carbohydrate antigen 19-9 (CA19-9), is fraught with poor sensitivity and specificity and is only used for monitoring disease on treatment or after surgical resection^8,9^.

Extracellular vesicles (EVs), including exosomes and microvesicles, are nanosized particles released by most cell types and can be detected in the circulation^10^. EVs play important roles in transmission of oncogenic and inflammatory signals^11^, communications between cells and their microenvironment^12^. In addition, exoDNA, exoRNA and protein profiles highly reflect parental cells, therefore offering an attractive strategy for diagnosing cancers non-invasively by analyzing EVs in the circulation^11,13^. Previous studies employed EVs to discover biomarkers for PDAC^13-16^, however those discovery proteomics experiments were carried out using cell lines or tumor tissue, which are not representative of the heterogeneity of human PDAC and are unable to recapitulate the systemic responses to cancer^14-16^. In addition, the EV biomarkers discovered in those studies have been compared only against healthy controls^14-16^. It is unclear how they would perform in subjects with underlying benign diseases of the pancreas, which is highly desirable from the clinical standpoint as many patients with PDAC have underlying chronic pancreatitis and cysts.

To meet this need, we conducted a large EV proteomics study from peripheral blood across a range of patients with pancreatic cancer, benign pancreatic diseases such as chronic pancreatitis and intraductal papillary mucinous neoplasm (IPMN), and healthy controls. Circulating EV (cEV) proteins detected included those involved in metabolism and immune regulation, in addition to proteins involved in protein binding, exocytosis, endocytosis and regulation of cellular protein localization that have been identified in previous studies^17,18^. We subsequently discovered multiple biomarker candidates for cancer diagnosis and verified several of them in an independent cohort of patients with the potential to aid in diagnosing pancreatic cancer. In addition, we identified a set of cEV proteins associated with metastasis which could provide a valuable resource for future biomarker studies.

## RESULTS

### Proteomics Characterization of Circulating EVs

In this study, we sought to identify proteins in extracellular vesicles in the blood that may be used as biomarkers for the diagnosis and prognosis of pancreatic cancer. With the approval of our institutional review board, we enrolled a total of 124 patients to the discovery cohort of this biomarker study (**Methods and Supplementary Table 1**). Subjects in the pancreatic cancer group (N=93) had a mean age of 66.5 years (range, 37-91), and 48.4% were female. All subjects had biopsy-proven disease. Thirty subjects had early-stage disease (stages I-II) and 63 had advanced disease (stages III-IV). Patients with benign pancreatic diseases included chronic pancreatitis (N=12) with a mean age of 57.5 years (range 37-78) and with 50% females, whereas IPMN included individuals with main duct and side branch IPMNs (N=8) with a mean age of 68.2 years (range 50-89) and with 87.5% being females. Subjects in the healthy control group (N=11) had a mean age of 53.4 years (range, 31-83) with 54.5% females (**Supplementary Table 1**).

We employed the novel EVtrap method (Extracellular Vesicles Total Recovery And Purification) to capture EVs from plasma samples and overcome the traditional laborious techniques for EV isolation, which are not scalable for large clinical studies. As described in recent reports, EVtrap is a magnetic bead-based isolation method that enables highly efficient capture of EVs from biofluids, confirmed by multiple common EV markers^19-23^. Previous analyses using electron microscopy and nanoparticle tracking also confirmed that the vast majority of particles isolated by EVtrap had diameters between 100-200 nm, consistent with exosomes^19^. In addition, EVtrap isolates demonstrates higher abundance of CD9, a common exosome marker, as compared to isolates from other traditional EV isolation methods such as size exclusion chromatography and ultracentrifugation^19^. Over 95% recovery yield can be achieved by EVtrap with less contamination from soluble proteins, a significant improvement over current commercially available methods as well as ultracentrifugation^19,21,24^.

Following EV isolation, samples were digested in-solution and analyzed by liquid chromatography-tandem mass spectrometry (nanoLC-MS/MS) on a high-resolution mass spectrometer (Q-Exactive HF-X). The workflow for cEVs isolation and enrichment and subsequent cEV mass spectrometry analysis is illustrated in **Figure 1A**.

**Figure 1.**
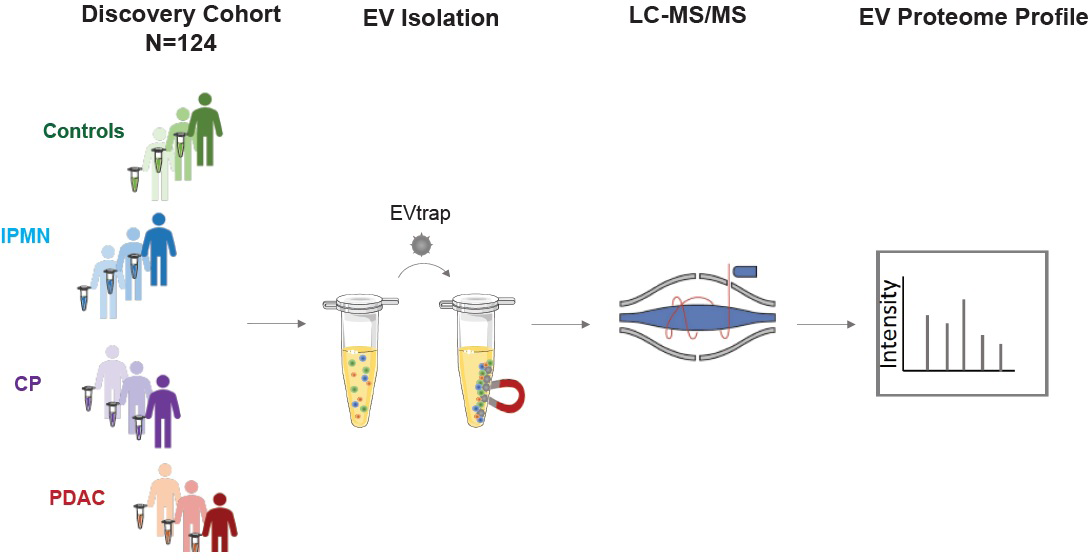
Study Design. The discovery cohort was comprised of 124 individuals, including pancreatic ductal adenocarcinoma (PDAC, N=93), chronic pancreatitis (CP, N=12), intraductal papillary mucinous neoplasm (IPMN, N=8) and healthy controls (N=11). Plasma samples were processed for EV isolation using EVtrap and analyzed by liquid chromatography-tandem mass spectrometry (LC-MS/MS).

First, to confirm that EVtrap can efficiently isolate extracellular vesicles from plasma, a test plasma sample was processed to remove platelets and other large particles and enriched for EVs using EVtrap beads (see **methods** for details). Transmission electron microscopy (TEM) analysis of the EV pellet showed cup-shaped extracellular vesicles (exosomes and microvesicles) (**Supplementary Figure 1A**), and nanoparticle tracking analysis (NTA) using ZetaView instrument (Particle Metrix) demonstrated that the isolated EVs were in the 100−200 nm diameter range, with a mean diameter of 152 nm (**Supplementary Figure 1B**). Second, to assess the technical reproducibility of the EV proteomics approach, the test plasma sample was processed in six replicates and Pearson correlation analysis revealed a very high correlation (r2 > 0.97) between replicates (**Supplementary Figure 2A, Supplementary Table 2**). These results provided the confidence to proceed with the analysis of our discovery set of plasma from 124 subjects. In this cohort, we identified 1,708 unique proteins (**Supplementary Table 3**). The number of unique EV proteins detected per 100µL of plasma sample varied from 817 to 1,128, with an average of 912 unique proteins per sample (**Supplementary Figure 2B**). We did not observe differences between non-tumor and tumor samples regarding the overall number of EV proteins identified. Within the PDAC group, we did not observe significant differences in the average number of EV proteins detected for different disease stages. Collectively, these data demonstrate high reproducibility of EV isolation and robust label-free MS quantification of cEVs.

### Diseases of the Pancreas Express Distinct Circulating EV Proteome Compared to Controls

Next, we aimed at identifying specific cEV proteins associated with clinical parameters with the potential to serve as diagnostic biomarkers. We first compared the proteomics profile of individuals with underlying pancreatic diseases (PDAC, chronic pancreatitis and IPMN) against healthy controls. We selected EV proteins expressed in at least 50% of subjects in the disease group with a fold change of expression ≥2 or ≤2 compared to controls and p-value ≤0.01 after adjusting for multiple testing. A total of 207 proteins were identified that met the criteria, with the largest number of differentially expressed markers in PDAC (176), followed by chronic pancreatitis (55) and IPMN (3) (**Supplementary Table 4**). Principal component analysis (PCA) of these markers showed control samples as a tight cluster segregated away from PDAC samples but closer to IPMN and chronic pancreatitis patients (**Figure 2A**).

**Figure 2.**
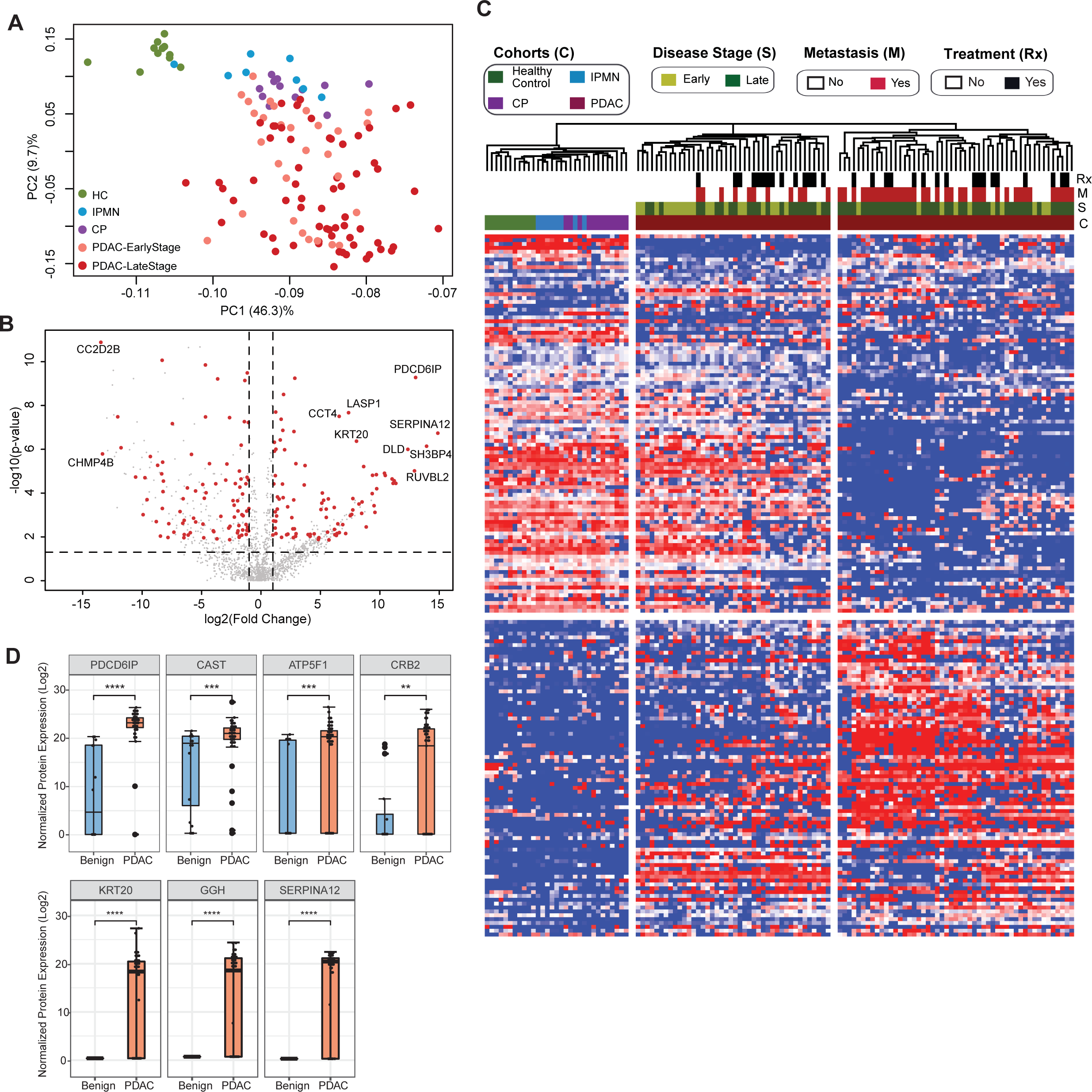
Identification of cEV Proteins Differentially Expressed in Disease Groups. **(A)** Principal component analysis of cEV proteins differentially expressed in the plasma of patients with pancreatic diseases compared to controls. Each dot indicates one individual enrolled in the study: green, controls; blue, patients with intraductal papillary mucinous neoplasm (IPMN); purple, patients with chronic pancreatitis (CP); salmon, early stage (stages I and II) pancreatic ductal adenocarcinoma (PDAC); red, late stage (stages III and IV) PDAC. **(B)** Volcano plot of circulating EV proteins enriched in the plasma of patients with PDAC versus benign pancreatic diseases. X-axis, log base 2 of fold changes; Y-axis, negative of the log base 10 of p values **(C)** Heatmap of cEV proteins differentially expressed in the plasma of patients with pancreatic diseases compared to controls. Designations of clinical parameters were indicated at the top of the heatmap. **(D)** Expression of enriched cEV proteins in patients with PDAC versus benign pancreatic diseases. Each dot indicates the target protein signal from one patient. Y-axis, normalized log base 2 of protein signals detected by mass spectrometry; Error bars, min and max values; lines in boxes, median values. * p ≤ 0.05, ** p ≤ 0.01, *** p ≤ 0.001, **** p ≤ 0.0001.

### Circulating EV Proteome Discriminates Pancreatic Cancer from Benign Pancreatic Diseases

To further assess the potential of cEV proteins for cancer detection, we compared proteomic profiles of cEVs between patients with PDAC with those with underlying benign diseases of the pancreas (chronic pancreatitis and IPMN). We identified 182 differentially expressed proteins in malignant cases (92 over-expressed and 90 with reduced expression) (**Supplementary Table 5)**. Several of those markers had remarkable overexpression in PDAC (greater than 10-fold), including PDCD6IP, SERPINA12, RUVBL2, among others, as shown in the volcano plot (**Figure 2B**). Unsupervised clustering showed a clear separation between PDAC and benign pancreatic diseases. Individuals with IPMN were more closely related to controls, whereas chronic pancreatitis cases were more related to PDAC **(Figure 2C)**. In addition, the PDAC cohort was separated into two subgroups: the first, enriched for early-stage tumors and more closely related to the other pancreatic diseases (chronic pancreatitis and IPMN); the second, enriched for advanced and metastatic cases with expression profiles further apart from early-stage cancer and pancreatic diseases (**Figure 2C**). We further noticed that some proteins such as PDCD6IP, SERPINA12, KRT20 showed statistically significant population-wise enrichment in pancreatic cancer compared to benign pancreatic diseases (**Figure 2D, Supplementary Figure 3**). Together, these data indicate the existence of EV markers that can separate controls, benign and malignant pancreatic diseases, as well as proteins that separate early versus late-stage PDAC, suggesting their potential to serve as diagnostic biomarkers.

### Functional and Systems Biology of cEV Proteome

To gain molecular insight into the functions of the 182 proteins differentially expressed in pancreatic cancer as compared to benign pancreatic diseases, we conducted pathway analysis using the Gene Ontology (GO) and REACTOME databases (**Supplementary Table 5**). We identified protein modules in protein localization, biomolecule binding/docking, peptidase activities among changes enriched in PDAC compared to benign diseases (**Supplementary Figure 4**). Interestingly, KRT20 (keratin 20), a gastrointestinal epithelia-associated keratin, was increased in PDAC patient EVs, while keratins associated basal cells, KRT4, KRT15, and KRT3, were reduced. KRT20 overexpression is frequently found in pancreatic tumor tissues and correlates with poor prognosis^25^, suggesting a biological basis for their high levels in the cEVs of PDAC patients.

Interestingly, proteins associated with immunological functions showed complex regulation with increased representation of leukocyte mediated immunity (GO:0002443), leukocyte degranulation (GO:0043299), myeloid leukocyte activation (GO:0002274), and decrease in Fc receptor signaling (GO: 0038093), regulation of complement activation (GO:0030449), and immune effector process (GO:0002252) (**Supplementary Table 5**). These data suggest that direct profiling of cEVs from patient plasma provided unique insights into systemic changes in immune biology during pancreatic cancer development, which is lacked in analysis restricted to tissue or cell models.

### Circulating EV Proteomics Reveal Markers Associated with Metastasis and Worse Prognosis

We then investigated whether cEV proteins can assist in the distinction of metastatic versus non-metastatic pancreatic cancer. We compared the cEV proteome profiles of individuals with metastatic cancer to those without metastasis and identified 85 proteins differentially expressed between the two groups (**Supplementary Table 6)**. Supervised clustering between metastatic and non-metastatic diseases showed a clear separation with two distinct expression patterns (**Figure 3A**). In particular, PSMB4, RUVBL2 and ANKAR (**Figure 3B**) EV protein levels were increased in patients with metastatic disease, whereas RAP2B, SERPINA12 and IGLV4-69 abundance levels were decreased in the cEVs of patients with metastasis (**Figure 3C**). Together, these findings suggest the presence of a core set of cEV proteins with the potential to distinguish early versus metastatic pancreatic cancer.

**Figure 3.**
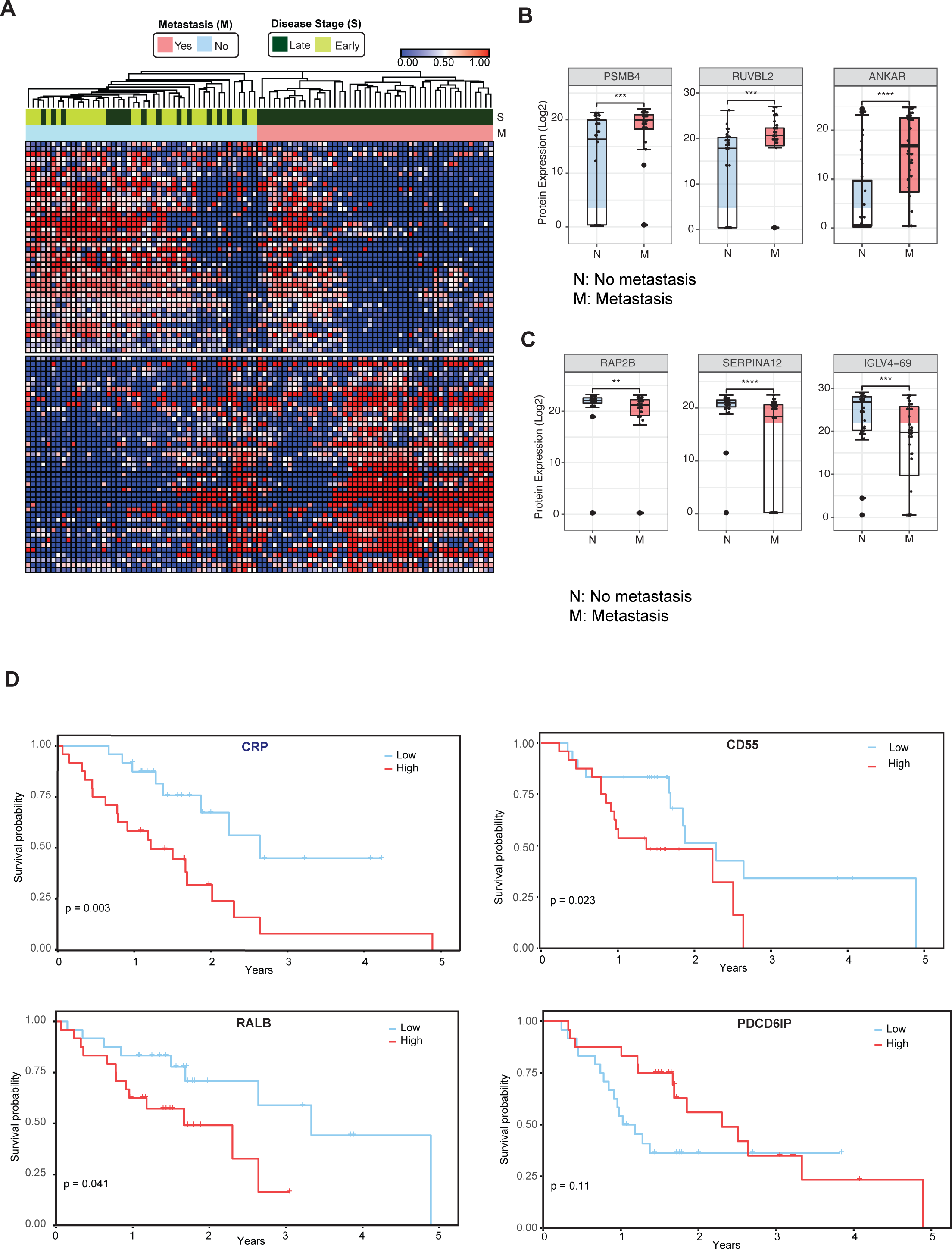
Circulating EV Proteomics Reveal Markers Associated with Metastasis and Worse Prognosis. **(A)** Heatmap showing EV proteins differentially expressed in the plasma of metastatic versus non-metastatic PDAC. Designations of clinical parameters are indicated at the top of the heatmap. **(B)** Expression patterns of cEV proteins associated with metastatic disease. Y-axis, normalized log base 2 of protein signals detected by mass spectrometry; N, non-metastatic PDAC group; M, metastatic PDAC group. Each dot indicated the target protein signal from one patient. Error bars, min and max values; lines in boxes, median values. * p ≤ 0.05, ** p ≤ 0.01, *** p ≤ 0.001, **** p ≤ 0.0001. **(C)** As is **(B)**, except for cEV markers with increased expression in non-metastatic PDAC. **(D)** Correlation of cEV marker expression with survival. Kaplan–Meier curves and log-rank test P values of representative survival cEV markers quantified in the discovery cohort.

We further analyzed whether the expression of certain cEV proteins had prognostic relevance in our cohort. We first classified individuals with PDAC as having low or high expression of any given markers based on each marker’s first and third quartile. Survival was estimated by the Kaplan Meier method. We identified that the cEV expression of RALB, CRP, and CD55 had a significant correlation with overall survival, with a trend for PDCD6IP (**Figure 3D**).

### Validation of cEV Markers Using Parallel Reaction Monitoring and Identification an EV Protein Signature for Pancreatic Cancer Diagnosis

Because pancreatic cancer is extremely heterogeneous, the chance of identifying a single biomarker with sufficient diagnostic performance is likely low. Instead, the identification of a panel of candidate markers may have enhanced diagnostic performance.

To identify a signature that shows the most discriminatory power between ‘benign diseases’ and ‘PDAC,’ we employed a binary classification approach using Support Vector Machines (SVM). Classification models, built based on a large number of proteins, contain irrelevant markers that can reduce the predictive accuracy. Hence, we implemented a consensus feature selection method based on two algorithms: one using recursive feature elimination (RFE) algorithm (SVM-RFE)^26^ and second, RFE combined with a non-parametric Wilcox rank test (sigFeature)^27^. The top 16 markers were selected whose classification performance can be tested in the independent validation cohort (**Supplementary Table 7**). A summary of selection process is shown in **Supplementary Figure 5**. The classification performance of these 16 markers, individual and in all combinations, were tested using 80% training data and evaluated in the remaining 20% test data. The quality of training was assessed using five repetitions of 10-fold cross-validation. The optimal kernel parameters were estimated by tuning over a wide range of values. Receiver operating characteristic (ROC) analysis was used as the metric to assess the performance of the classifier model. We found a set of 7-EV protein signature comprised of RUVBL2, PDCD6IP, ATP5F1, DLD, KRT20, CCT4, and SERPINAI2, that gave 100% accuracy when tested in the discovery cohort (**Supplementary Table 7, Supplementary Figure 6**). Recurrence of these putative markers in our dataset varied from 55% to 97%.

The model was further validated on an independent validation cohort whose proteome was obtained using an alternate technology, parallel reaction monitoring (PRM) mass spectrometry. The markers chosen for validation included 16 markers selected for SVM classification model and an additional 9 markers to result in top 25 markers that are significantly differentially expressed in the discovery cohort with a fold change increase in PDAC ≥ 5.5 and p-value ≤0.01 (**Methods**, **Supplementary Table 8**). The independent validation cohort consisted of 36 new subjects (24 with PDAC, 6 with chronic pancreatitis, and 6 with IPMN) (**Supplementary Table 9**). A total of 10 proteins, including all 7 signature proteins, showed a significant difference (p < 0.05) in patients with PDAC as compared to benign pancreatic diseases (**Figure 4A**). The performance of individual validated markers according to the specific underlying disease in the validation cohort is presented in **Supplementary Figure 7**. The performance of 7-EV protein signature was further tested using SVM model, in our independent validation cohort, yielding an 89% prediction accuracy (**Figure 4B, Supplementary Figure 8**). As expected, we observed that no single marker achieved sufficiently high sensitivity and specificity as the combined model for the diagnosis of pancreatic cancer.

**Figure 4.**
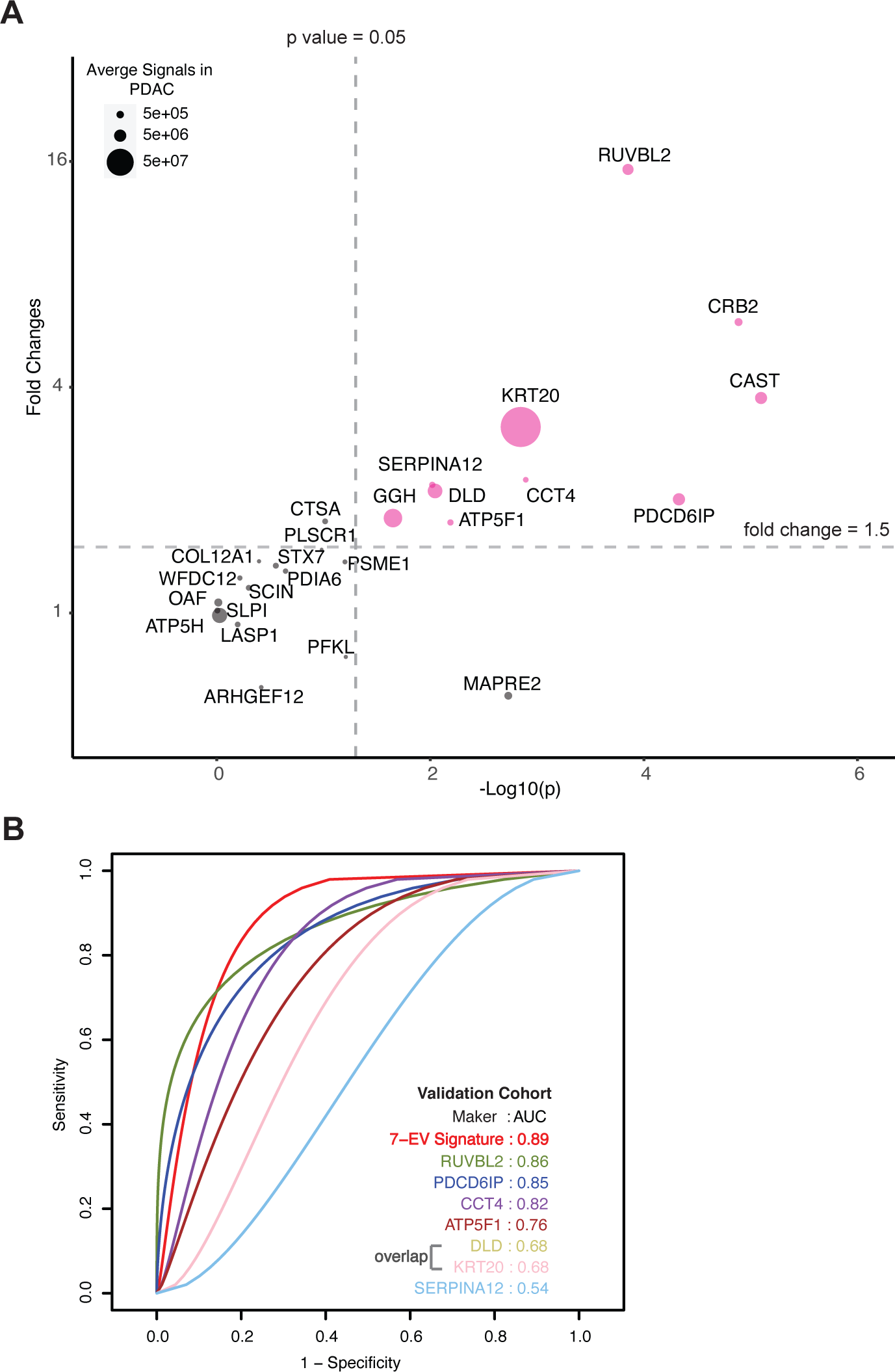
Validation of cEV Markers and Identification of 7-EV Protein Signature for PDAC Diagnosis. **(A)** Differences of cEV protein abundances between patients with PDAC (n=24) and benign pancreatic diseases (chronic pancreatitis and IPMN) (n=12). x axis, minus log *p* values of protein abundance differences between PDAC and benign groups; y axis, average fold changes of proteins in PDAC group compared to benign group. Size of bubbles indicate average protein abundances in PDAC group. Pink color, proteins that had at least two-fold enrichment in PDAC group (p<0.05). **(B)** ROC curves were calculated for individual cEV markers as well as for the 7-EV protein PDAC signature combination to determine optimum diagnostic performance. AUC, area under the curve.

## DISCUSSION

Extracellular vesicles hold a great promise as a source of potential biomarkers, making them attractive candidates for liquid biopsy tests. Previously, we reported that organoid cultures of pancreatic cancer could serve as models to discover tissue-derived EV proteins with high specificity for PDAC, as opposed to chronic pancreatitis and other benign gastrointestinal diseases^28^. A shortcoming of these tissue-based studies is the inability to discover markers associated to the systemic responses to cancer. Here, we performed a large-scale, comprehensive analysis of circulating EV proteomes directly from plasma samples of 124 patients, with subsequent validation in a separate cohort of 36 patients. To our knowledge, this represents the largest proteomics profiling dataset of circulating EVs conducted in pancreatic cancer to date. In this study, we identified and validated new EV markers from plasma that distinguish patients with pancreatic cancer from subjects with benign pancreatic diseases. Furthermore, we discovered several cEV proteins associated with metastatic disease and poor prognosis. In contrast to the prior studies of experimental cell models or tissues extracts that were examined only against healthy subjects^14-16^, we report the identification of EV proteins in plasma of patients with pancreatic cancer compared to patients with underlying pancreatic diseases, which is clinically relevant as many patients with pancreatic cancer have underlying chronic inflammation and premalignant cystic lesions.

In addition, our study demonstrated the feasibility of using the novel EVtrap method^19,20^ for discovery of hundreds of EV proteins directly from a small volume (100uL) of plasma samples. This methodological advance can be adopted for biomarker discovery in other cancer types. Other workflows traditionally employed for EV isolation from blood samples require laborious techniques including lengthy ultracentrifugation steps which are unsuitable for large scale studies^29^.

We identified several EV proteins as significantly associated with metastasis or survival. For instance, PSMB4 and RUVBL2 levels were increased in cEVs of patients with metastatic PDAC. Notably, PSMB4 (proteasome subunit beta type-4), a protein of the ubiquitin-proteasome degradation pathway, has been identified as the first proteasomal subunit with oncogenic properties and associated to poor prognosis in several tumors including melanoma, breast and ovarian cancers^30-33^. As expected, the EV proteomic profiles of PDAC patients exhibited significant heterogeneity. While the above-mentioned markers exhibited strong association with disease states at population levels, their abundances in individual patients varied significantly. Those observations highlight the need to develop multi-protein panels for pancreatic cancer diagnosis and prognosis.

We also discovered RALB, CRP and CD55 expression on EVs to have a significant correlation with poor survival, while PDCD6IP expression was associated with improved outcomes. Interestingly, PDCD6IP (programmed cell death 6-interacting protein), was also identified as a PDAC-enriched protein in the tissue-based proteomics studies from Le Large *et.al*^34^ and Hoshino *et.al*^18^. In line with our findings, its tissue expression in liver metastasis of pancreatic cancer has been found to also correlate with improved prognosis in patients with PDAC in the study of Law *et.al*^35^. Collectively, these data suggest that some tissue-specific proteins can be isolated from circulating EVs and their quantifiable levels in the blood may have the potential to serve as diagnostic or prognostic biomarkers in pancreatic cancer.

In our validation studies, all seven putative markers identified from the model were significantly enriched in the plasma of PDAC patients. Based on the top seven markers, we derived a 7-EV protein panel that yielded an 89% prediction accuracy for diagnosing pancreatic cancer. A recent modeling study showed that a new diagnostic assay for PDAC would have to perform with a minimum sensitivity of 88% and a specificity of 85% to reduce healthcare expenditure and prolong survival^6^. Serum CA19-9, the best-established blood test for PDAC, has a pooled sensitivity of 75.4% and a specificity of 77.6%^36^. It commonly rises late in the disease and may be elevated in nonmalignant conditions such as biliary obstruction and pancreatitis, making it unsuitable as a diagnostic biomarker for PDAC^37^. As such, our 7-EV protein signature with 89% prediction accuracy serves as a proof-of-concept and has the potential to facilitate the further development of biomarker tests for pancreatic cancer. We anticipate that for clinical use application, an even higher diagnostic performance is needed. Future studies are warranted to investigate if combining our validated cEV proteins with other biomarkers such as cell free DNA, serum proteins or metabolites, as a multi-analyte biomarker assay, would yield higher accuracy in diagnosing pancreatic cancer.

While our work involved a large cohort with 160 patients, the single-center nature is an inherent limitation of our study. Also, it would be ideal to perform validation with a larger cohort of controls to achieve greater statistical power. To balance this limitation, we increased the rigor of our validation by selecting controls with underlying benign diseases of pancreas as opposed to healthy volunteers, and an alternate quantitative technology for measuring protein abundance (Parallel Reaction Monitoring Mass Spectrometry) instead of MS-LC. While this approach increased the generalizability, it marginally reduced model prediction. Thus, the performance of our 7-EV protein PDAC panel should be cross validated in larger and multicenter populations. In addition, in this work we only used EVtrap as EV isolation method and mass spectrometry for protein quantification, and it is possible that there was some degree of heterogeneity in the extracellular vesicles analyzed. The clinical impact of biomarkers identified in our study will need to be cross validated using other methods.

With no major treatment breakthrough for pancreatic cancer in the last decade, every effort should be made to diagnose this deadly cancer at earlier stages and to discover new proteins involved in tumorigenesis. Our study provides a valuable open resource to the scientific community with a comprehensive catalog of novel proteins packaged inside circulating EVs that may assist in the development of novel biomarkers and improve the outcomes of patients with pancreatic cancer.

## METHODS

### Study Design and Patient Demographics

We conducted this study at Beth Israel Deaconess Medical Center with the approval of the Harvard Cancer Center Institutional Review Board. All subjects provided written informed consent. Clinical data and blood samples were prospectively collected from 2017 to 2019 from patients with pancreatic cancer, chronic pancreatitis, intraductal papillary mucinous neoplasms (IPMN), and age-matched controls. A total of 124 patients, including PDAC (N=93), chronic pancreatitis of different etiologies (N=12), IPMN (N=8), and controls (N=11), were included in the discovery cohort. PDAC diagnosis was established by histology or cytology, and staging was performed according to the American Joint Committee on Cancer guidelines (8th Edition 2016)^38^(**Supplementary Table 1**). For the independent validation cohort, a total of 36 patients were enrolled, including PDAC (N=24), IPMN (N=6), and chronic pancreatitis (N=6) (**Supplementary Table 9**).

### Plasma Sample Collection and Processing

All blood samples were collected and processed following the same standard operating procedure optimized for EV analysis and included the following steps: (i) whole blood was collected into one 10ml yellow-top tube containing acid citrate dextrose; (ii) blood was mixed by gently inverting the tube five times; (iii) vacutainer tubes were stored upright at room temperature (RT); (iv) samples were centrifuged at 1,300g for 15 min in RT; (v) plasma was removed from the top carefully avoiding cell pellet; (vi) repeat centrifugation of plasma at 2,500g for 15 min in RT; (vii) again, plasma was removed from the top carefully avoiding cell pellet; (viii) third centrifugation at 2,500g for 15 min in RT, then samples were aliquoted to be stored at -80°C.

### Extracellular Vesicle Isolation from Plasma

We employed EVtrap for EV isolation from plasma samples^19^. EVtrap beads were provided by Tymora Analytical (West Lafayette, IN) as a suspension in water and were used as previously described in more details^19,20^. Briefly, 100 μL plasma samples were diluted 20 times in the diluent buffer, the EVtrap beads were added to the samples in a 1:2 v/v ratio, and the samples were incubated by end-over-end rotation for 30 min according to the manufacturer’s instructions. After supernatant removal using a magnetic separator rack, the beads were washed with PBS, and the EVs were eluted by a 10 min incubation with 200 mM triethylamine (TEA, Millipore-Sigma). The samples were fully dried in a vacuum centrifuge.

### Preparation of EV samples

The isolated and dried EV samples were lysed to extract proteins using the phase-transfer surfactant (PTS) aided procedure. The proteins were reduced and alkylated by incubation in 10 mM TCEP and 40 mM CAA for 10 min at 95°C. The samples were diluted fivefold with 50 mM triethylammonium bicarbonate and digested with Lys-C (Wako) at 1:100 (wt/wt) enzyme-to-protein ratio for 3 h at 37°C. Trypsin was added to a final 1:50 (wt/wt) enzyme-to-protein ratio for overnight digestion at 37°C. To remove the PTS surfactants from the samples, the samples were acidified with trifluoroacetic acid (TFA) to a final concentration of 1% TFA, and ethyl acetate solution was added at a 1:1 ratio. The mixture was vortexed for 2 min and then centrifuged at 16,000 × g for 2 min to obtain aqueous and organic phases. The organic phase (top layer) was removed, and the aqueous phase was collected. This step was repeated once more. The samples were dried in a vacuum centrifuge and desalted using Top-Tip C18 tips (Glygen) according to the manufacturer’s instructions. The samples were dried completely in a vacuum centrifuge and stored at -80°C.

### LC-MS Analysis of Plasma EV Proteome

Approximate 1 μg of each dried peptide sample was dissolved in 10.5 μL of 0.05% trifluoroacetic acid with 3% (vol/vol) acetonitrile containing spiked-in indexed Retention Time Standard containing 11 artificially synthetic peptides (Biognosys). The spiked-in 11-peptides standard mixture was used to account for any variation in retention times and to normalize abundance levels among samples. 10 μL of each sample was injected into an Ultimate 3000 nano UHPLC system (Thermo Fisher Scientific). Peptides were captured on a 2-cm Acclaim PepMap trap column and separated on a heated 50-cm Acclaim PepMap column (Thermo Fisher Scientific) containing C18 resin. The mobile phase buffer consisted of 0.1% formic acid in ultrapure water (buffer A) with an eluting buffer of 0.1% formic acid in 80% (vol/vol) acetonitrile (buffer B) run with a linear 60-min gradient of 6–30% buffer B at a flow rate of 300 nL/min. The UHPLC was coupled online with a Q-Exactive HF-X mass spectrometer (Thermo Fisher Scientific). The mass spectrometer was operated in the data-dependent mode, in which a full-scan MS (from m/z 375 to 1,500 with the resolution of 60,000) was followed by MS/MS of the 15 most intense ions (30,000 resolution; normalized collision energy - 28%; automatic gain control target (AGC) - 2E4, maximum injection time - 200 ms; 60sec exclusion).

### EV Proteome Data Processing

The raw files were searched directly against the human Swiss-Prot database with no redundant entries using Byonic (Protein Metrics) and Sequest search engines loaded into Proteome Discoverer 2.3 software (Thermo Fisher Scientific). MS1 precursor mass tolerance was set at 10 ppm, and MS2 tolerance was set at 20ppm. Search criteria included a static carbamidomethylation of cysteines (+57.0214 Da) and variable modifications of oxidation (+15.9949 Da) on methionine residues and acetylation (+42.011 Da) at the N terminus of proteins. The search was performed with full trypsin/P digestion and allowed a maximum of two missed cleavages on the peptides analyzed from the sequence database. The false-discovery rates of proteins and peptides were set at 0.01. All protein and peptide identifications were grouped, and any redundant entries were removed. Only unique peptides and unique master proteins were reported.

All data were quantified using the label-free quantitation node of Precursor Ions Quantifier through the Proteome Discoverer v2.3 (Thermo Fisher Scientific). For the quantification of proteomic data, the intensities of peptides were extracted with initial precursor mass tolerance set at 10 ppm, a minimum number of isotope peaks as 2, maximum ΔRT of isotope pattern multiplets – 0.2 min, PSM confidence FDR of 0.01, with hypothesis test of ANOVA, maximum RT shift of 5 min, pairwise ratio-based ratio calculation, and 100 as the maximum allowed fold change. The abundance levels of all peptides and proteins were normalized to the spiked-in internal iRT standard. For calculations of fold-change between the groups of proteins, total protein abundance values were added together, and the ratios of these sums were used to compare proteins within different samples.

The abundances of EV proteins were normalized using indexed retention time (iRT) in Proteome Discoverer (ThermoFisher Scientific). Abundances were categorized into four different categories: Control, Chronic Pancreatitis, IPMN, and PDAC. Protein abundances were then log2 transformed and quantile normalized for further analysis.

A non-parametric Wilcox Rank Sum test was performed to test the null hypothesis that the distributions of two groups of the patient population are the same, and the fold change and p-values for each protein were estimated for the following comparisons: IPMN vs. Control, CP vs. Control, PDAC vs. Control, Benign Pancreatic Diseases (CP, IPMN) vs. PDAC. Multiple testing correction was done using Benjamini-Hochberg method to control for the false discovery rate^39^. Volcano plots were created using those p values and fold change. Heatmaps visualization and clustering of statistically significant proteins, with adjusted p-value ≤ 0.05 and absolute fold change ≥ 2, were created in R using the pheatmap package. Euclidean distance and average cluster method were used. The values were row-scaled for normalization. Both rows and columns were allowed to cluster.

### Pathways Enrichment and Protein Network Analysis

Pathway enrichment analysis was performed on statistically significant genes using g:Profiler^40^, a web-based tool that searches for pathways whose genes are significantly enriched in our dataset compared to a collection of genes representing Gene Ontology (GO) terms and Reactome pathways. We further used EnrichmentMap^41^, a Cytoscape, v3.8.2^42^ application to create a visual network of connected pathways that helps to identify relevant pathways and theme^43^. A Protein-Protein interaction network was generated using a stringApp, a Cytoscape app. This application allows to import STRING networks into Cytoscape and enables to perform complex network analysis and visualization of networks^44^.

### Parallel Reaction Monitoring and Data Analysis

Parallel reaction monitoring mass spectrometry (PRM-MS) was employed for validation experiments. Twenty-five cEV markers were selected for validation based on fold change increase ≥5.5, p-value ≤0.01, and technical aspects (number of unique peptides and coverage) (**Supplementary Table 8**). Thirty-six plasma samples from a new cohort were used for the validation (24 PDAC, 6 IPMN and 6 chronic pancreatitis samples). The EVs were isolated from plasma and the proteins processed as described before. Peptide samples were dissolved in 10.8 μL 0.05% TFA & 2% ACN, and 10 μL injected into the UHPLC coupled with a Q-Exactive HF-X mass spectrometer (Thermo Fisher Scientific). The mobile phase buffer consisted of 0.1% formic acid in HPLC grade water (buffer A) with an eluting buffer containing 0.1% formic acid in 80% (vol/vol) acetonitrile (buffer B) run with a linear 60-min gradient of 5–35% buffer B at a flow rate of 300 nL/min. Each sample was analyzed under PRM with an isolation width of ±0.8 Th. In these PRM experiments, an MS2 level at 30,000 resolution relative to m/z 200 (AGC target 2E5, 200 ms maximum injection time) was run as triggered by a scheduled inclusion list. Higher-energy collisional dissociation was used with 28 eV normalized collision energy. PRM data were manually curated within Skyline-daily (64-bit) 20.2.1.404 (32d27b598)^45^.

### Identification of EV signature for pancreatic cancer diagnosis

To identify a biomarker signature demonstrating the highest discriminatory power between ‘benign’ and ‘PDAC’ diseases, we adopted a binary classification approach utilizing Support Vector Machines (SVM). Recognizing that classification models built on an extensive array of proteins may incorporate irrelevant markers, which can diminish the predictive accuracy, we started with a list of significantly differentially expressed set of 91 proteins between ‘benign’ and ‘PDAC’ patients and further employed a consensus feature selection strategy using two algorithms, ‘Recursive Feature Elimination’ (SVM-RFE), and ‘Integrated RFE with a non-parametric Wilcox rank test (sigFeature). Subsequently, we selected the top 16 markers, the classification performance of which was subjected to testing in an independent validation cohort. The classification performance evaluation of these markers, both individually and in various combinations, involved a rigorous assessment utilizing 80% of the data for training and the remaining 20% for internal-validation. To ensure the quality of the training process, we employed five repetitions of a 10-fold cross-validation approach. The optimal kernel parameters were determined through tuning across a broad range of values. Receiver operating characteristic (ROC) analysis served as the metric to gauge the performance of the classifier model. All algorithms for identifying the EV signature predictive of pancreatic cancer diagnosis were implemented in R. We used Support Vector Machine (SVM) using CRAN package, e107^46^. Ranking of genes was achieved using packages ‘sigFeature’ and ‘SVM-RFE’. An R package, ‘pROC’^47^ was used to build a receiver operating characteristic curve (ROC) and to calculate the area under the curve (AUC).

### Survival Analysis

The prognostic value of every protein was estimated by dividing patients into two groups: group 1, patients with expression below the 25^th^ percentile, and group 2, patients with expression values greater than 75^th^ percentile. The Kaplan-Meier estimator was used to estimate the survival function associating survival with EV protein expression, and the log-rank test was used to compare survival curves of two groups. ‘survival’ R package was used for the analysis.

### Statistical Analysis

All statistical analyses were performed using the statistical software R. Statistical significance was calculated by two-tailed Student’s t-test or Wilcoxon rank-sum test unless specified otherwise in the figure legend. Data are expressed as mean ± SEM. A p-value < 0.05 in biological experiments or FDR < 0.05 after multiple comparison corrections in proteomics data analysis was considered statistically significant.

## Data Availability

All data presented all be made available upon request

## Acknowledgments

We thank the patients and their families for their participation in this study. B.B. was supported in part through UM1 (CA186709-06). S.D.F. was supported in part through the Barbara Janson and Arthur Hilsinger Pancreatology Fellowship. Institutional startup funds and UO1 (CA224193) to S.K.M., and seed grant from Hirschberg Foundation for Pancreatic Cancer Research to L.H. We thank members of the Muthuswamy laboratory for their critical input throughout the development of this project.

## SUPPLEMENTARY MATERIAL

**Supplementary Table 1**. Baseline characteristics of patients enrolled on the discovery cohort.

**Supplementary Table 2.** Plasma EV analysis reproducibility

**Supplementary Table 3.** LC-MS results of EV analysis of plasma from patients with PDAC (PA), IPMN, Chronic Pancreatitis (CP) and Control individuals.

**Supplementary Table 4.** List of EV proteins that met the eligibility criteria for principal component analysis.

**Supplementary Table 5**. List of 182 proteins differentially expressed in PDAC compared to benign diseases.

**Supplementary Table 6**. List of EV proteins that are significantly altered in patients with metastatic versus non-metastatic diseases.

**Supplementary Table 7. Table A:** Support Vector Machine Prediction model output for the 16 individual markers included in the in External Validation Cohorts. Table B: The contingency table for 7-biomarker signature, offering insights into model accuracy for both the Internal-Discovery and External Validation cohorts.

**Supplementary Table 8**. List of 25 cEV proteins that met the eligibility criteria for validation studies.

**Supplementary Table 9**. Baseline characteristics of patients enrolled in the validation cohort.

**Supplementary Figure 1. EVtrap isolation of extracellular vesicles**

**(A)** Transmission electron microscopy (TEM) images collected of a single EV and multiple EVs captured from plasma by EVtrap. TEM imaging of EVs was carried out on a HITACHI H-8100 electron microscope (Hitachi, Tokyo, Japan) with an accelerating applied potential of 200 kV.

**(B)** Nanoparticle tracking analysis (NTA) of EVs after elution off EVtrap beads. NTA was carried out using ZetaView instrument (Particle Metrix) after calibration with 100 nm polystyrene particles.

**Supplementary Figure 2. EV proteomics analytical performance**

**(A)** Reproducibility of the method. A standard plasma sample was processed in six replicates and performed a Pearson correlation analysis that revealed a very high correlation between replicates.

**(B)** Number of quantified EV proteins per sample according to different patient cohort.

**Supplementary Figure 3.** Heatmap of abundance of 25 proteins enriched and 25 proteins reduced in EVs from PDAC patients compared to EVs from patients without cancer. Protein abundances were normalized across patients for each protein.

**Supplementary Figure 4. Network Analyses of cEV Proteins Differentially Expressed in PDAC Compared to Benign Pancreatic Diseases.**

**(A)** Functional association of proteins identified by STRING database. Red, cEV proteins enriched in PDAC patients as compared to benign pancreatic diseases. Green, cEV proteins decreased in patients with PDAC as compared to benign pancreatic diseases. Red, cEV proteins increased in PDAC as compared to benign pancreatic diseases. Thickness of lines indicate confidence of association.

**(B, C)** Clustering of cEV protein pathways enriched **(B)** or downregulated **(C)** in PDAC cohorts. Pathways were identified using Gene Ontology database and REACTOME database.

**Supplementary Figure 5. Summary of selection process to develop EV signature for pancreatic cancer diagnosis.**

**Supplementary Figure 6.** Diagnostic performance of 7-EV protein signature compared to performance of each of the 7 individual marker.

**Supplementary Figure 7. Validation of individual cEV proteins in an independent cohort of patients.**

Expression of biomarker candidates detected by Parallel Reaction Monitoring (PRM) analyses. A total of 25 cEV proteins with significant overexpression in PDAC in the discovery cohorts were quantified by PRM in a separate validation cohort of patients.

**Supplementary Figure 8.** Performance of PDAC EV Signature in both Discovery and Validation cohorts.

## REFERENCES

1. Siegel RL, Miller KD, Jemal A: Cancer statistics, 2020. CA Cancer J Clin 70:7-30, 2020

2. Rahib L, Smith BD, Aizenberg R, et al: Projecting cancer incidence and deaths to 2030: the unexpected burden of thyroid, liver, and pancreas cancers in the United States. Cancer Res 74:2913–21, 2014

3. Kamisawa T, Wood LD, Itoi T, et al: Pancreatic cancer. Lancet 388:73–85, 2016

4. Rahib L, Fleshman JM, Matrisian LM, et al: Evaluation of Pancreatic Cancer Clinical Trials and Benchmarks for Clinically Meaningful Future Trials: A Systematic Review. JAMA Oncol 2:1209–16, 2016

5. Bockorny B, Grossman JE, Hidalgo M: Facts and Hopes in Immunotherapy of Pancreatic Cancer. Clin Cancer Res 28:4606–4617, 2022

6. Ghatnekar O, Andersson R, Svensson M, et al: Modelling the benefits of early diagnosis of pancreatic cancer using a biomarker signature. Int J Cancer 133:2392–7, 2013

7. Matsuno S, Egawa S, Fukuyama S, et al: Pancreatic Cancer Registry in Japan: 20 years of experience. Pancreas 28:219–30, 2004

8. Locker GY, Hamilton S, Harris J, et al: ASCO 2006 update of recommendations for the use of tumor markers in gastrointestinal cancer. J Clin Oncol 24:5313–27, 2006

9. Galli C, Basso D, Plebani M: CA 19-9: handle with care. Clin Chem Lab Med 51:1369–83, 2013

10. Chen IH, Xue L, Hsu CC, et al: Phosphoproteins in extracellular vesicles as candidate markers for breast cancer. Proc Natl Acad Sci U S A 114:3175–3180, 2017

11. Costa-Silva B, Aiello NM, Ocean AJ, et al: Pancreatic cancer exosomes initiate pre-metastatic niche formation in the liver. Nat Cell Biol 17:816–26, 2015

12. van Niel G, Carter DRF, Clayton A, et al: Challenges and directions in studying cell-cell communication by extracellular vesicles. Nat Rev Mol Cell Biol 23:369–382, 2022

13. Melo SA, Luecke LB, Kahlert C, et al: Glypican-1 identifies cancer exosomes and detects early pancreatic cancer. Nature 523:177–82, 2015

14. Madhavan B, Yue S, Galli U, et al: Combined evaluation of a panel of protein and miRNA serum-exosome biomarkers for pancreatic cancer diagnosis increases sensitivity and specificity. Int J Cancer 136:2616–27, 2015

15. Yang KS, Im H, Hong S, et al: Multiparametric plasma EV profiling facilitates diagnosis of pancreatic malignancy. Sci Transl Med 9, 2017

16. Castillo J, Bernard V, San Lucas FA, et al: Surfaceome profiling enables isolation of cancer-specific exosomal cargo in liquid biopsies from pancreatic cancer patients. Ann Oncol 29:223–229, 2018

17. Fahrmann JF, Mao X, Irajizad E, et al: Plasma-Derived Extracellular Vesicles Convey Protein Signatures that Reflect Pathophysiology in Lung and Pancreatic Adenocarcinomas. Cancers (Basel) 12, 2020

18. Hoshino A, Kim HS, Bojmar L, et al: Extracellular Vesicle and Particle Biomarkers Define Multiple Human Cancers. Cell 182:1044–1061 e18, 2020

19. Iliuk A, Wu X, Li L, et al: Plasma-Derived Extracellular Vesicle Phosphoproteomics through Chemical Affinity Purification. J Proteome Res 19:2563–2574, 2020

20. Wu X, Li L, Iliuk A, et al: Highly Efficient Phosphoproteome Capture and Analysis from Urinary Extracellular Vesicles. J Proteome Res 17:3308–3316, 2018

21. Nunez Lopez YO, Iliuk A, Petrilli AM, et al: Proteomics and Phosphoproteomics of Circulating Extracellular Vesicles Provide New Insights into Diabetes Pathobiology. Int J Mol Sci 23, 2022

22. Hinzman CP, Jayatilake M, Bansal S, et al: An optimized method for the isolation of urinary extracellular vesicles for molecular phenotyping: detection of biomarkers for radiation exposure. J Transl Med 20:199, 2022

23. Hinzman CP, Singh B, Bansal S, et al: A multi-omics approach identifies pancreatic cancer cell extracellular vesicles as mediators of the unfolded protein response in normal pancreatic epithelial cells. J Extracell Vesicles 11:e12232, 2022

24. Shuen TWH, Alunni-Fabbroni M, Ocal E, et al: Extracellular Vesicles May Predict Response to Radioembolization and Sorafenib Treatment in Advanced Hepatocellular Carcinoma: An Exploratory Analysis from the SORAMIC Trial. Clin Cancer Res 28:3890–3901, 2022

25. Schmitz-Winnenthal FH, Volk C, Helmke B, et al: Expression of cytokeratin-20 in pancreatic cancer: an indicator of poor outcome after R0 resection. Surgery 139:104–8, 2006

26. Guyon I, Weston J, Barnhill S, et al: Gene Selection for Cancer Classification using Support Vector Machines. Machine Learning 46:389–422, 2002

27. Das P, Roychowdhury A, Das S, et al: sigFeature: Novel Significant Feature Selection Method for Classification of Gene Expression Data Using Support Vector Machine and t Statistic. Front Genet 11:247, 2020

28. Huang L, Bockorny B, Paul I, et al: PDX-derived organoids model in vivo drug response and secrete biomarkers. JCI Insight, 2020

29. LeBleu VS, Kalluri R: Exosomes as a Multicomponent Biomarker Platform in Cancer. Trends Cancer 6:767–774, 2020

30. Liu R, Lu S, Deng Y, et al: PSMB4 expression associates with epithelial ovarian cancer growth and poor prognosis. Arch Gynecol Obstet 293:1297–307, 2016

31. Zhang X, Lin D, Lin Y, et al: Proteasome beta-4 subunit contributes to the development of melanoma and is regulated by miR-148b. Tumour Biol 39:1010428317705767, 2017

32. Zheng P, Guo H, Li G, et al: PSMB4 promotes multiple myeloma cell growth by activating NF-kappaB-miR-21 signaling. Biochem Biophys Res Commun 458:328-33, 2015

33. Lee GY, Haverty PM, Li L, et al: Comparative oncogenomics identifies PSMB4 and SHMT2 as potential cancer driver genes. Cancer Res 74:3114–26, 2014

34. Le Large TY, Mantini G, Meijer LL, et al: Microdissected pancreatic cancer proteomes reveal tumor heterogeneity and therapeutic targets. JCI Insight 5, 2020

35. Law HC, Lagundzin D, Clement EJ, et al: The Proteomic Landscape of Pancreatic Ductal Adenocarcinoma Liver Metastases Identifies Molecular Subtypes and Associations with Clinical Response. Clin Cancer Res 26:1065–1076, 2020

36. Zhang Y, Yang J, Li H, et al: Tumor markers CA19-9, CA242 and CEA in the diagnosis of pancreatic cancer: a meta-analysis. Int J Clin Exp Med 8:11683-91, 2015

37. Duffy MJ, Sturgeon C, Lamerz R, et al: Tumor markers in pancreatic cancer: a European Group on Tumor Markers (EGTM) status report. Ann Oncol 21:441–7, 2010

38. Amin MB, Greene FL, Edge SB, et al: The Eighth Edition AJCC Cancer Staging Manual: Continuing to build a bridge from a population-based to a more “personalized” approach to cancer staging. CA Cancer J Clin 67:93–99, 2017

39. Benjamini Y, Hochberg Y: Controlling the False Discovery Rate: A Practical and Powerful Approach to Multiple Testing. Journal of the Royal Statistical Society: Series B (Methodological) 57:289–300, 1995

40. Raudvere U, Kolberg L, Kuzmin I, et al: g:Profiler: a web server for functional enrichment analysis and conversions of gene lists (2019 update). Nucleic Acids Res 47:W191–W198, 2019

41. Merico D, Isserlin R, Stueker O, et al: Enrichment map: a network-based method for gene-set enrichment visualization and interpretation. PLoS One 5:e13984, 2010

42. Shannon P, Markiel A, Ozier O, et al: Cytoscape: a software environment for integrated models of biomolecular interaction networks. Genome Res 13:2498–504, 2003

43. Reimand J, Isserlin R, Voisin V, et al: Pathway enrichment analysis and visualization of omics data using g:Profiler, GSEA, Cytoscape and EnrichmentMap. Nat Protoc 14:482–517, 2019

44. Szklarczyk D, Gable AL, Lyon D, et al: STRING v11: protein-protein association networks with increased coverage, supporting functional discovery in genome-wide experimental datasets. Nucleic Acids Res 47:D607–D613, 2019

45. MacLean B, Tomazela DM, Shulman N, et al: Skyline: an open source document editor for creating and analyzing targeted proteomics experiments. Bioinformatics 26:966-8, 2010

46. Meyer D, Dimitriadou E, Hornik K, et al: Misc Functions of the Department of Statistics, ProbabilityTheory Group (Formerly: E1071), TU Wien, 2015

47. Robin X, Turck N, Hainard A, et al: pROC: an open-source package for R and S+ to analyze and compare ROC curves. BMC Bioinformatics 12:77, 2011

