## Supplementary figures and images for "A Large-Scale Proteomics Resource of Circulating Extracellular Vesicles for Biomarker Discovery in Pancreatic Cancer"

### Supplementary Figure 1

#### Supplementary Figure 1

A

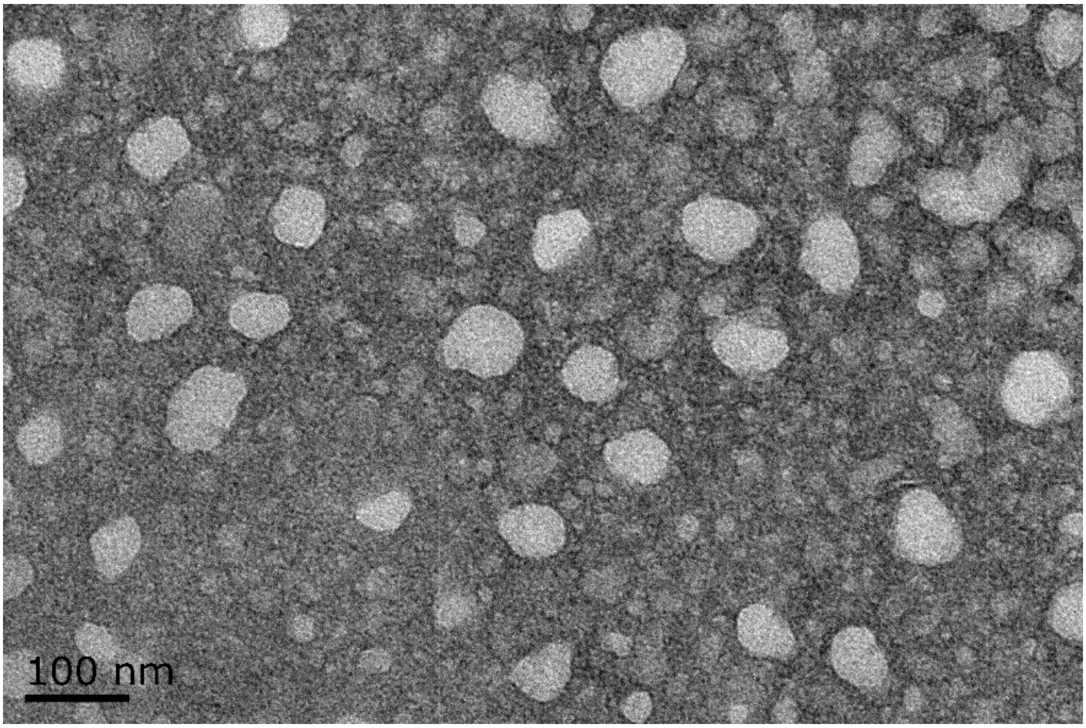


B

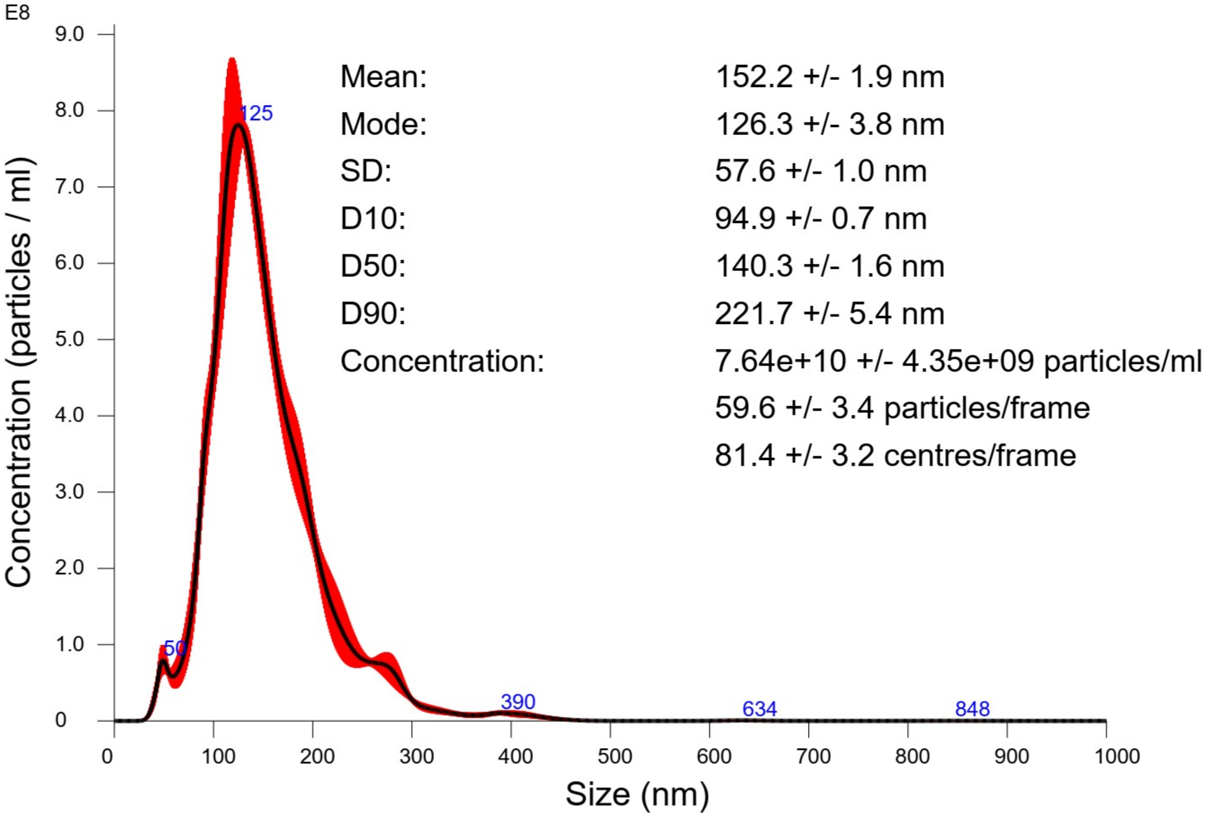

### Supplementary Figure 2

##### Supplementary Figure 2

**A**

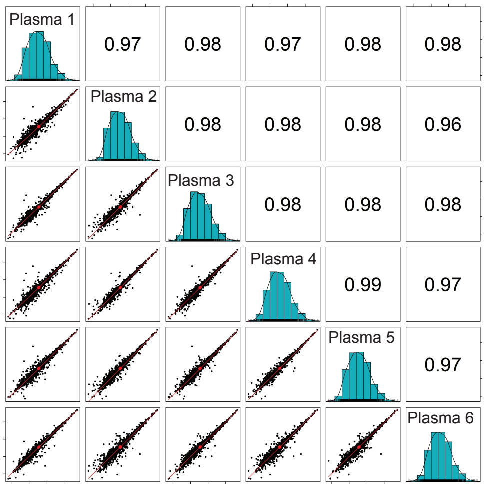


**B**

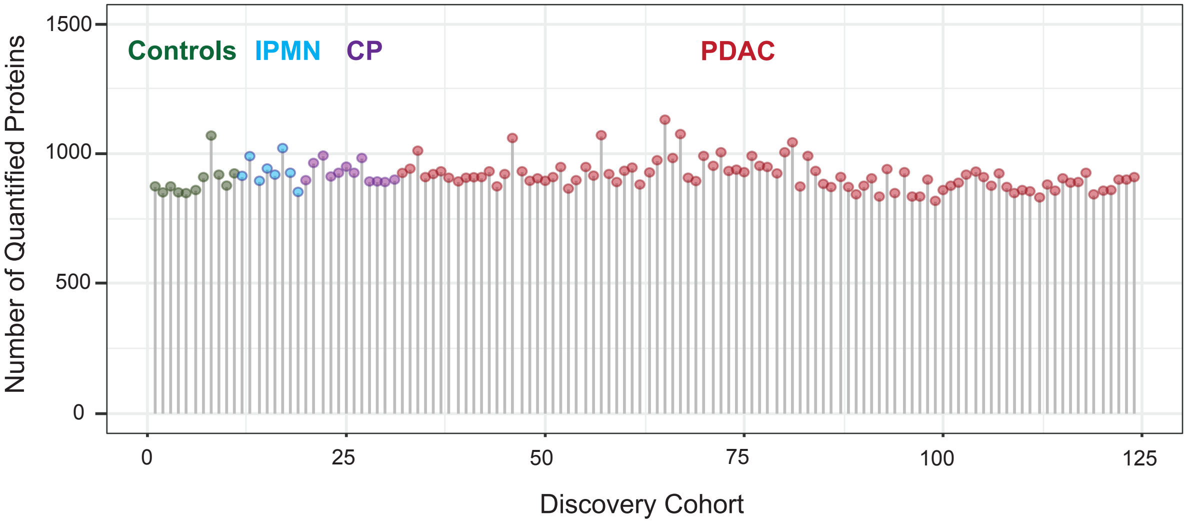

### Supplementary Figure 3

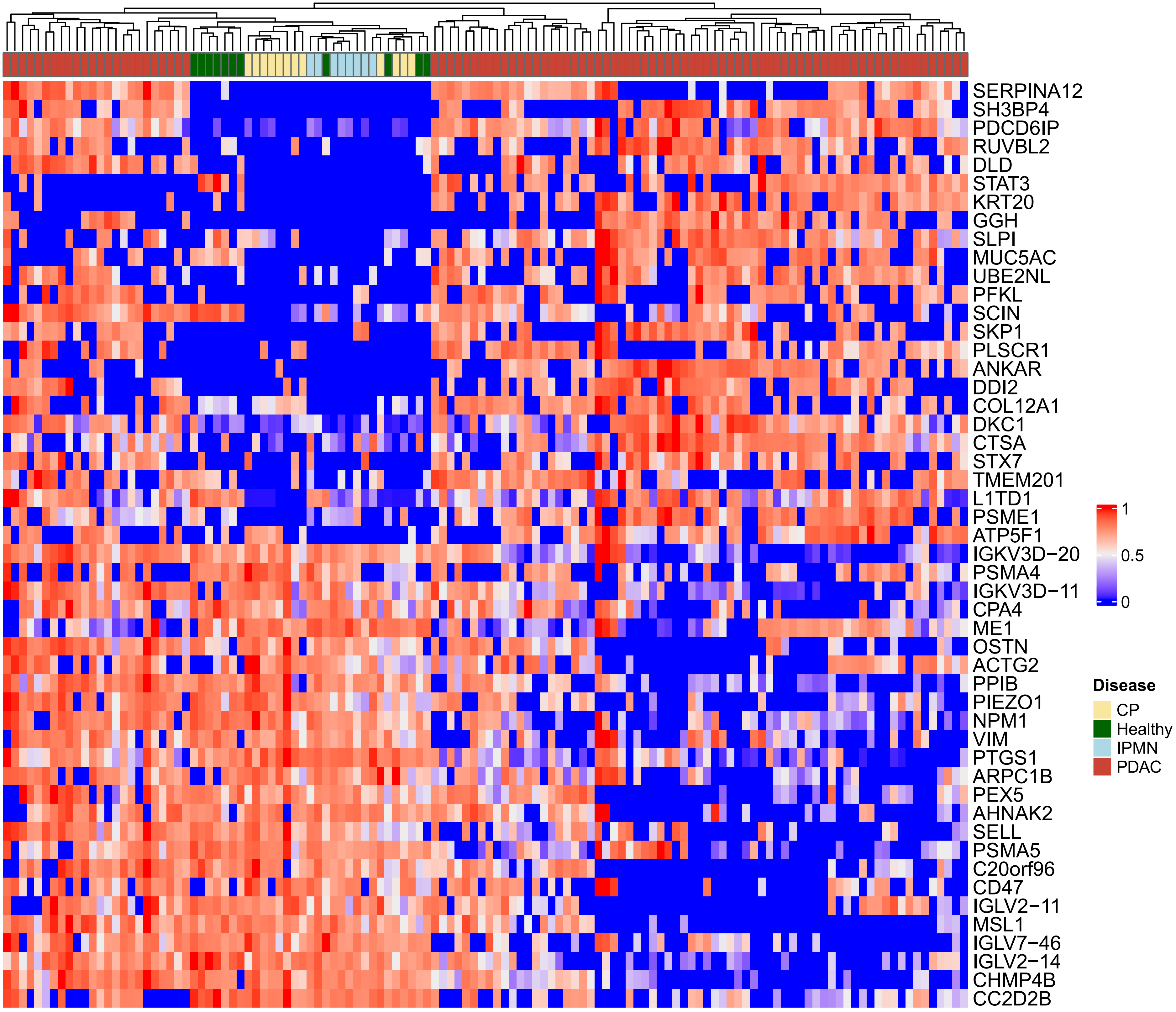

### Supplementary Figure 4

A

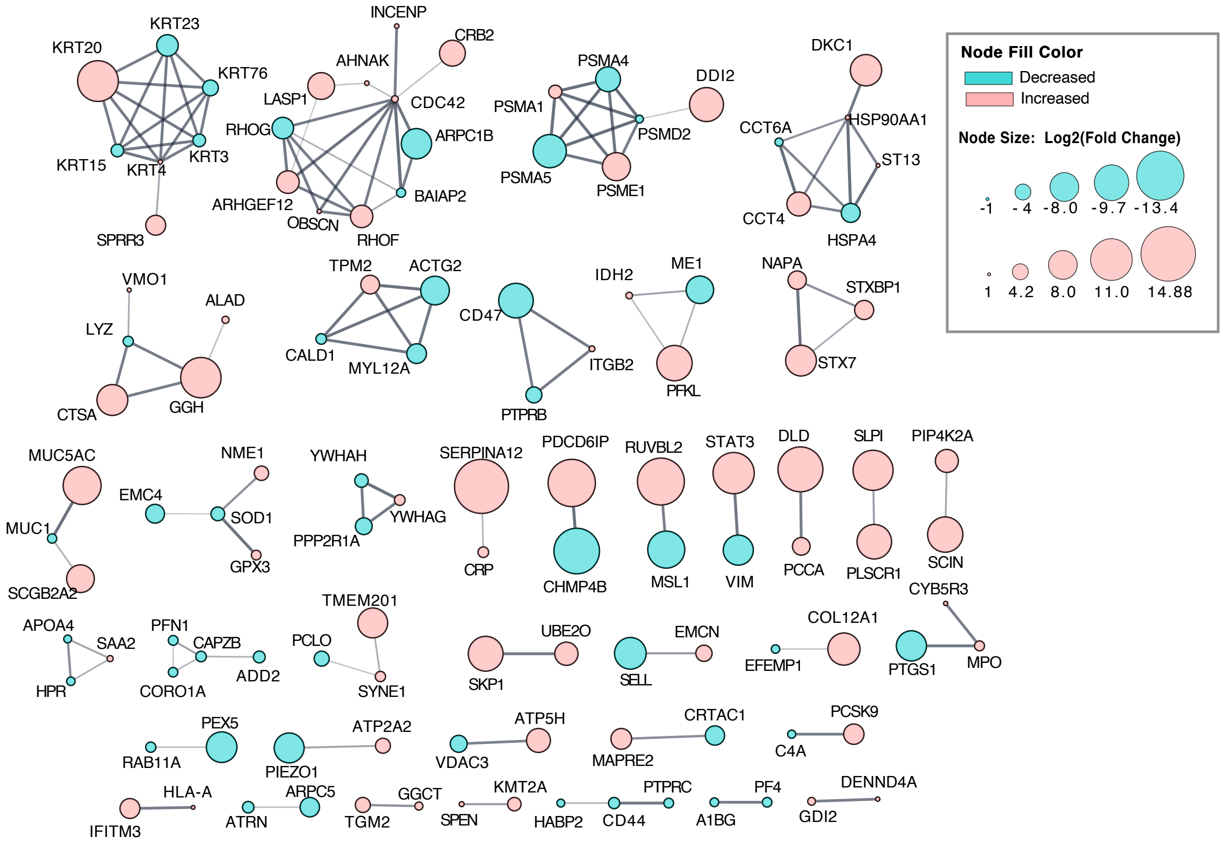


B

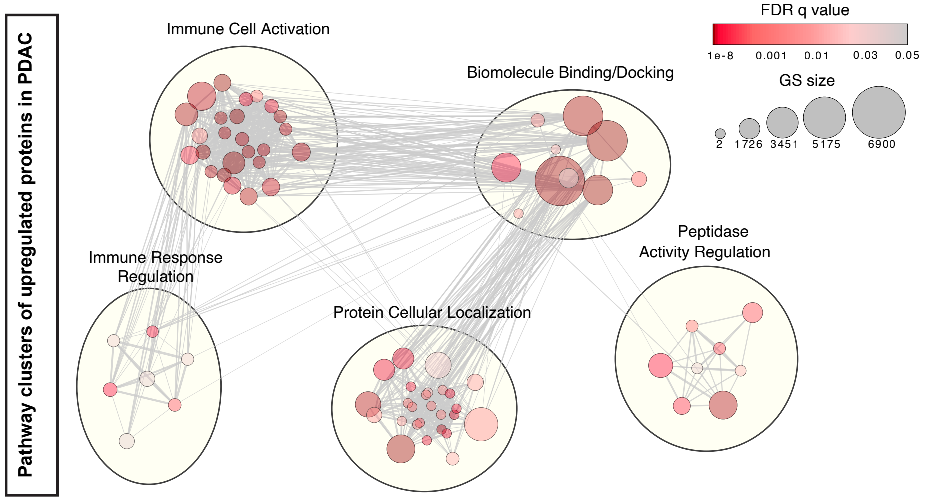


C

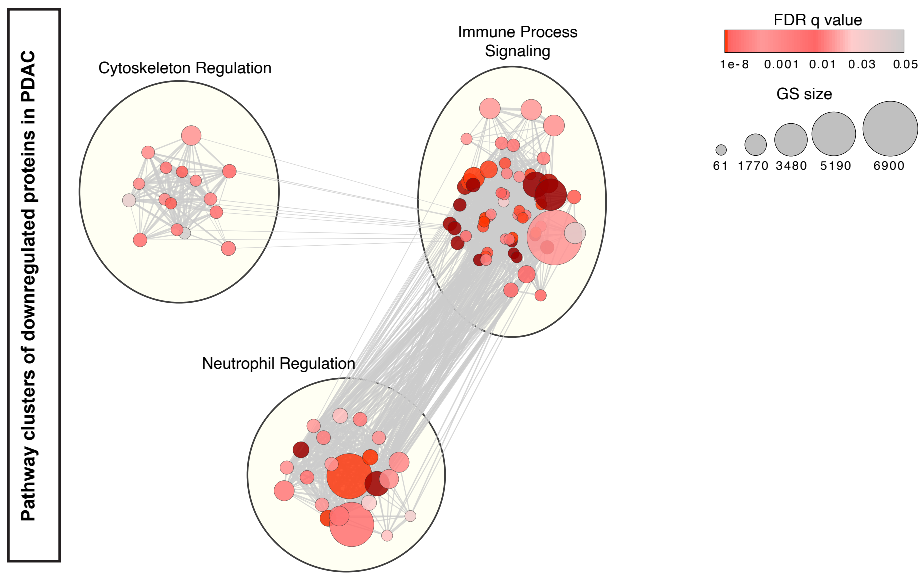


**Suppl. Figure 4**

### Supplementary Figure 5

**Supplementary Figure 5**

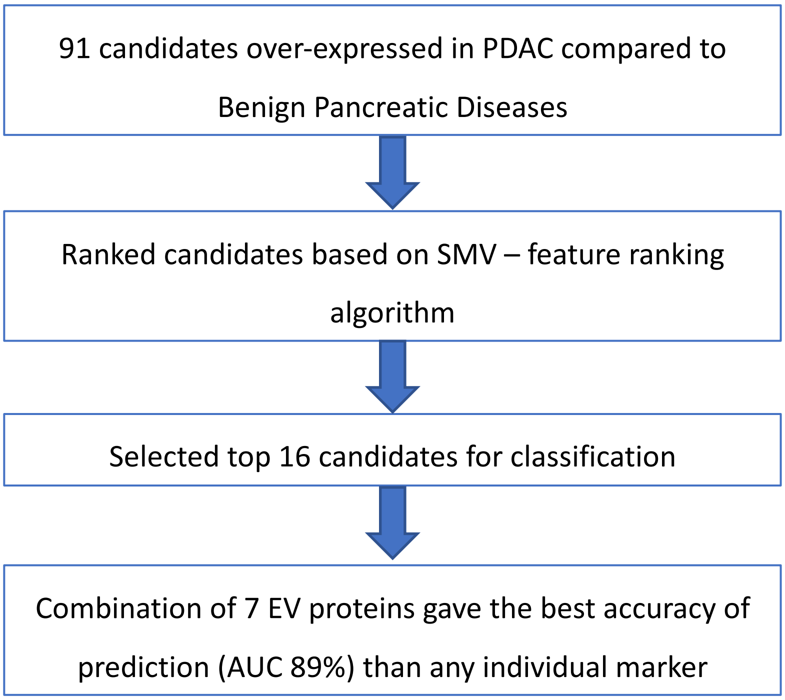

### Supplementary Figure 6

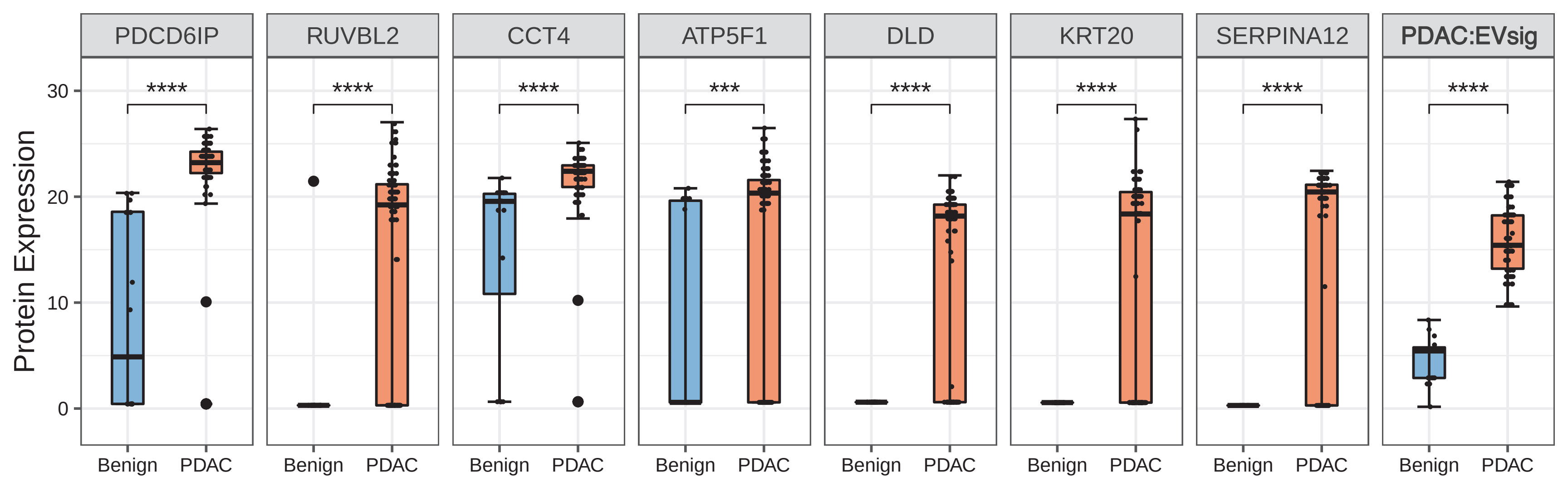

### Supplementary Figure 7

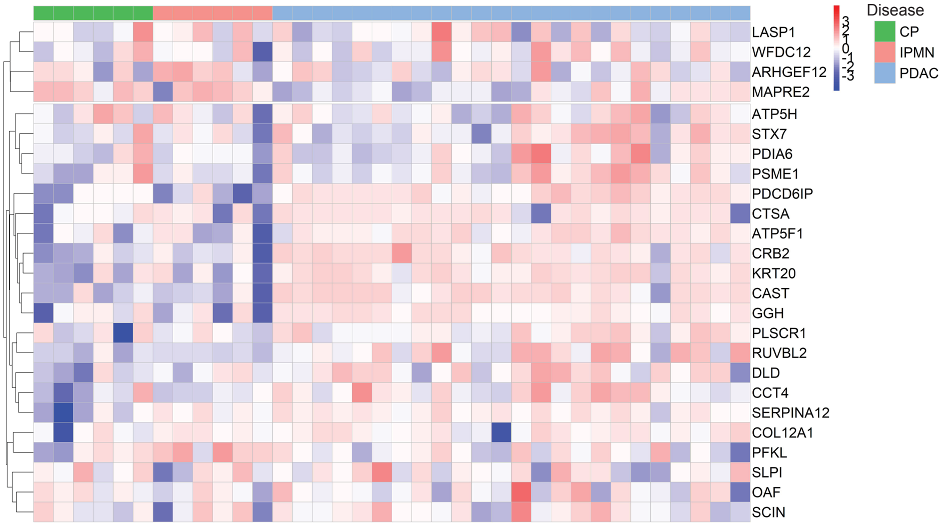


**Supplementary Figure 7**

### Supplementary Figure 8

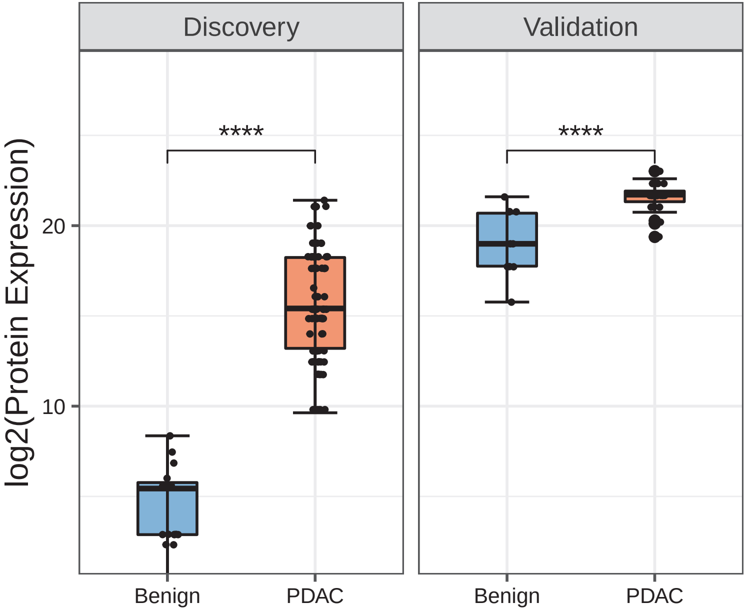
