## Supplementary Table 1 for "A Large-Scale Proteomics Resource of Circulating Extracellular Vesicles for Biomarker Discovery in Pancreatic Cancer"

| **Supplementary Table 1. Baseline characteristics of patients enrolled on the discovery cohort** | | | | |  |  |
| --- | --- | --- | --- | --- | --- | --- |
|  | **Pancreatic Cancer** | | | **Chronic Pancreatitis** | **IPMN*** | **Controls** |
|  | **I-II** | **III-IV** | **All Stages** |  |  |  |
| Number of subjects | 30 | 63 | 93 | 12 | 8 | 11 |
| Age (mean, range) | 72.36 (58-91) | 63.6 (37-82) | 66.5 (37-91) | 57.5 (37-78) | 68.2 (50-89) | 53.4 (31-83) |
| Gender (F) | 43.30% | 52.30% | 48.40% | 50% | 87.50% | 54.50% |
| Baseline CA19-9 (mean, range) | 246.96 (4- 2,421) | 24,054 (4-700,000) | 16,118 (4-700,000) | - | - | - |
| * IPMN group was comprised of two patients with main duct lesions and six patients with side-branch lesions. | | | | | | |
