## Supplementary Table 2 for "A Large-Scale Proteomics Resource of Circulating Extracellular Vesicles for Biomarker Discovery in Pancreatic Cancer"

**Supplementary Table 2. Plasma EV analysis reproducibility**

| Abundances (Normalized): Pl | Abundances (Normalized): Pl | Abundances (Normalized): Pl |
|---|---|---|
| 21.10292053 | 28.44822693 | 20.43869972 |
| 19.20351982 | 20.22241783 | 26.01729584 |
| 18.48998833 | 21.95749664 | 16.26365852 |
| 24.47357178 | 23.13701057 | 22.53872108 |
| 20.94659996 | 16.30039215 | 21.74411392 |
| 22.67775536 | 22.46438789 | 22.70429993 |
| 18.61883545 | 25.01817131 | 24.62578201 |
| 17.09104919 | 22.03594208 | 19.84741592 |
| 21.9788723 | 18.03815079 | 19.86117744 |
| 18.05012131 | 13.84064198 | 17.37805939 |
| 18.90679932 | 17.65001678 | 18.02108002 |
| 19.2427578 | 18.12082863 | 18.46420288 |
| 21.04858208 | 20.31843185 | 17.03453255 |
| 18.39983749 | 21.68955612 | 18.30734634 |
| 14.72741795 | 17.31340218 | 14.02591133 |
| 20.53605461 | 16.2827282 | 20.53548813 |
| 22.30321503 | 20.81468201 | 17.64860344 |
| 16.31697273 | 17.60038757 | 16.46922874 |
| 15.67556 | 19.71768188 | 17.50234795 |
| 18.53872871 | 18.75877953 | 18.26347733 |
| 24.95194244 | 21.63393593 | 22.27240753 |
| 19.72587395 | 20.16625786 | 19.75801849 |
| 20.49734879 | 21.4585762 | 21.08137321 |
| 19.01573563 | 15.58484459 | 19.63193321 |
| 20.4367485 | 20.30464554 | 20.56299591 |
| 18.44562721 | 21.88730621 | 18.62530899 |
| 27.11043739 | 23.14153099 | 27.00409317 |
| 19.05642509 | 18.6377964 | 19.91100502 |
| 22.47563934 | 22.51219368 | 22.38379288 |
| 25.56875038 | 26.35604668 | 22.63398933 |
| 20.14299011 | 20.70135498 | 20.13190079 |
| 19.52144814 | 18.18267441 | 19.11607742 |
| 21.30612755 | 22.0608387 | 19.96650887 |
| 19.90321541 | 17.22467995 | 19.2022419 |
| 22.74886131 | 22.71250916 | 25.50106239 |
| 18.21022797 | 21.72729874 | 19.68479919 |
| 19.95934868 | 22.23920059 | 20.88112068 |
| 18.96846199 | 19.87121964 | 19.638937 |
| 19.16464233 | 19.67747116 | 19.49303055 |
| 19.26055717 | 17.56649208 | 17.5868187 |
| 21.45511627 | 17.89232254 | 20.94699097 |
| 23.94357681 | 21.91524696 | 21.99703598 |
| 17.67493248 | 15.25628948 | 18.22766495 |
| 19.23129463 | 20.00166702 | 18.82384682 |

| | | |
|---|---|---|
| 16.84575272 | 19.88234138 | 19.29631805 |
| 23.20229912 | 23.36773872 | 24.30758476 |
| 23.14934731 | 24.33117676 | 22.90717316 |
| 18.84384537 | 16.97755814 | 18.85171127 |
| 18.49448776 | 15.6914835 | 17.70629883 |
| 19.67020416 | 19.64443207 | 19.74754715 |
| 22.68821907 | 19.67525864 | 22.20681572 |
| 20.54520416 | 22.12301445 | 21.01124382 |
| 17.28112602 | 18.7717762 | 18.51916885 |
| 19.86084747 | 17.19166374 | 19.87664986 |
| 25.08874512 | 22.45386314 | 22.73286057 |
| 19.26724052 | 18.87102699 | 18.73014641 |
| 15.70520592 | 13.65357113 | 14.24940872 |
| 19.54604721 | 18.83798218 | 18.99269676 |
| 21.69579124 | 19.68637657 | 18.2651844 |
| 19.42674255 | 16.79570389 | 19.24985886 |
| 19.59301186 | 21.11605072 | 19.4900074 |
| 17.14294815 | 19.76805878 | 19.02619934 |
| 20.80435753 | 24.36296272 | 20.9279213 |
| 19.39093208 | 17.25701332 | 18.01500893 |
| 17.43137932 | 15.08681583 | 17.07252884 |
| 16.38567924 | 19.54303932 | 19.2723484 |
| 16.90377808 | 18.65580559 | 17.9756546 |
| 19.01913643 | 19.97206879 | 18.84394455 |
| 14.85355377 | 15.3103857 | 14.70685768 |
| 24.4854393 | 21.26295853 | 21.09917068 |
| 19.17593765 | 19.75653648 | 18.43221283 |
| 16.38912773 | 18.95663261 | 16.46190834 |
| 20.94968796 | 20.8393631 | 20.9376564 |
| 19.87433624 | 22.825737 | 22.69854355 |
| 22.01216316 | 20.54215813 | 21.41410637 |
| 19.70679474 | 22.62734032 | 21.68969727 |
| 17.2851162 | 17.03212738 | 19.41326523 |
| 21.67163086 | 23.86614037 | 21.54693985 |
| 23.64892578 | 21.43955994 | 20.50188637 |
| 15.02447701 | 16.45342827 | 15.64505768 |
| 20.56401634 | 20.38579941 | 20.25194931 |
| 19.38983727 | 17.15729141 | 20.06565475 |
| 17.95934677 | 20.773592 | 20.74568558 |
| 21.11170197 | 22.68492508 | 22.57698631 |
| 24.68005371 | 21.52497673 | 24.78059578 |
| 17.92732239 | 16.93915939 | 16.27341652 |
| 17.86384773 | 17.70866394 | 15.68539143 |
| 19.49520874 | 19.87230682 | 21.37083435 |
| 16.7379818 | 18.54576492 | 16.15345764 |
| 22.87246704 | 19.94389534 | 20.35476112 |
| 16.40218163 | 14.81329441 | 15.75652599 |

| | | |
|---|---|---|
| 25.98865509 | 22.69498825 | 25.8673172 |
| 19.39677238 | 20.66121674 | 19.79176521 |
| 22.42378044 | 24.21517754 | 22.59063339 |
| 19.10835075 | 18.04616165 | 19.46960068 |
| 20.72237396 | 20.32219696 | 19.04111862 |
| 18.68190384 | 20.28398514 | 19.21009445 |
| 18.58485413 | 17.82939911 | 17.95550919 |
| 16.82444572 | 16.03363419 | 16.93095207 |
| 21.83984375 | 24.64079094 | 23.87934685 |
| 18.45473099 | 16.35136414 | 18.351408 |
| 17.66550636 | 18.8262558 | 17.9396019 |
| 19.69779396 | 18.19826889 | 18.53886795 |
| 18.37022018 | 20.7764225 | 20.09128952 |
| 22.62380981 | 24.03448105 | 22.73296356 |
| 20.51350021 | 22.98869324 | 20.62390518 |
| 21.38891602 | 19.05922318 | 21.48781967 |
| 21.99757385 | 22.55972862 | 22.29468346 |
| 17.86493492 | 19.63310814 | 19.4012661 |
| 19.27510452 | 20.12433815 | 19.94294167 |
| 23.41430855 | 21.29524231 | 22.12908173 |
| 20.31629562 | 18.12437439 | 20.16700935 |
| 18.57012558 | 18.62133408 | 18.36756706 |
| 18.24254417 | 18.36239243 | 18.40299797 |
| 22.16347694 | 19.72130203 | 21.33789825 |
| 26.48061562 | 23.89249611 | 25.78944588 |
| 19.63998795 | 20.56671333 | 20.29436684 |
| 16.49072838 | 18.77104187 | 18.18795586 |
| 19.15557289 | 19.78436089 | 19.58929443 |
| 24.08522034 | 22.78694153 | 22.88711357 |
| 17.93762779 | 18.17747498 | 19.56604958 |
| 20.47182274 | 21.42894554 | 20.8802166 |
| 19.10735512 | 21.19185638 | 21.00255394 |
| 19.14774704 | 19.20859528 | 19.81671143 |
| 20.4622097 | 20.53937531 | 19.04551315 |
| 19.20852661 | 18.73428917 | 20.17641258 |
| 15.61035728 | 17.02580261 | 16.90720367 |
| 20.86717606 | 19.6327877 | 20.2976284 |
| 21.10283661 | 21.96624184 | 22.54637718 |
| 19.42390823 | 19.60161018 | 19.50391197 |
| 25.71436882 | 25.05393791 | 23.71487617 |
| 19.51463699 | 21.46639633 | 21.14829636 |
| 20.97956276 | 19.54347229 | 19.34103203 |
| 18.69488335 | 19.05100441 | 19.71998596 |
| 20.38823509 | 22.54148102 | 21.9952774 |
| 22.01431274 | 20.47018814 | 20.38432312 |
| 21.23548126 | 21.325634 | 21.45308685 |
| 17.4387989 | 18.11164665 | 18.91612816 |

| | | |
|---|---|---|
| 19.51581192 | 19.18554878 | 19.29194069 |
| 20.98707008 | 19.46549416 | 19.66733932 |
| 22.07969093 | 22.51817322 | 21.78955841 |
| 18.67176056 | 19.11767006 | 19.42836571 |
| 23.11468887 | 24.88202095 | 22.1704998 |
| 19.30068588 | 18.26227951 | 20.65751648 |
| 19.1815052 | 20.76857185 | 19.0508461 |
| 18.03624153 | 18.92562485 | 18.68789864 |
| 23.5596981 | 23.42808533 | 23.28265572 |
| 20.77586937 | 19.76727676 | 21.39601707 |
| 19.33563805 | 19.38822365 | 21.04786873 |
| 15.96446991 | 17.55018425 | 15.90424633 |
| 19.27072716 | 18.17570305 | 18.40216255 |
| 20.32805634 | 21.02336884 | 20.87864304 |
| 20.39996147 | 18.2751503 | 19.60524368 |
| 19.39895439 | 18.00490952 | 17.58098221 |
| 23.12890053 | 24.8731823 | 23.74012756 |
| 19.70098495 | 19.11989021 | 19.71108246 |
| 20.46757889 | 19.51379585 | 18.49919701 |
| 16.98498726 | 17.2642231 | 15.81793308 |
| 19.65303421 | 19.42858124 | 20.80589104 |
| 16.75207329 | 18.10959435 | 17.08347893 |
| 18.62246513 | 20.75998116 | 19.09223938 |
| 19.6228447 | 21.73952866 | 20.55864143 |
| 22.07154274 | 20.19100952 | 21.58702469 |
| 20.47127342 | 22.26995659 | 20.6514473 |
| 19.74328613 | 19.77495384 | 19.52267456 |
| 23.84472466 | 22.95518875 | 22.29860878 |
| 21.97691727 | 22.80240631 | 20.82170296 |
| 22.04627419 | 20.35285759 | 20.35160065 |
| 18.84181595 | 19.11507416 | 17.36700821 |
| 19.932024 | 19.85745049 | 18.53578949 |
| 18.2332325 | 19.51493454 | 19.16806793 |
| 22.01777267 | 22.07638359 | 22.14569473 |
| 18.94684792 | 17.38502121 | 18.96213913 |
| 20.6803093 | 20.58796883 | 20.55626106 |
| 19.52166176 | 18.94104385 | 19.72813416 |
| 20.55323219 | 21.2515831 | 19.47087479 |
| 22.13467407 | 20.19514275 | 21.57809448 |
| 18.549963 | 19.11881065 | 18.24668503 |
| 19.52329636 | 17.52198601 | 18.31591606 |
| 20.99603844 | 19.68448257 | 19.76277924 |
| 16.82178688 | 17.99111366 | 17.09101105 |
| 23.69613838 | 22.54550171 | 23.58760262 |
| 20.40871048 | 19.73857498 | 19.77872658 |
| 18.45321083 | 18.40466118 | 17.59263802 |
| 21.4846859 | 21.77094841 | 21.4953022 |

| | | |
|---|---|---|
| 23.43185234 | 21.52738762 | 22.47687721 |
| 25.3731823 | 22.97160912 | 24.53712845 |
| 22.31406975 | 20.49860001 | 22.60056496 |
| 19.42886162 | 19.25518799 | 21.23187447 |
| 17.49962425 | 19.30347252 | 18.37694168 |
| 18.20436096 | 20.0982399 | 19.54897881 |
| 24.03940201 | 23.43851852 | 25.00896072 |
| 20.55631828 | 21.46000481 | 20.17201233 |
| 17.46273613 | 18.95013046 | 17.83211899 |
| 18.3756752 | 20.43216324 | 19.67980576 |
| 23.12296295 | 21.97126389 | 22.8609848 |
| 14.24872112 | 14.74816704 | 13.73875141 |
| 22.23862839 | 20.89084816 | 20.8905468 |
| 21.09629822 | 22.97764397 | 21.77778053 |
| 24.33372307 | 22.76827049 | 22.85188103 |
| 20.01904488 | 21.52412987 | 20.89256096 |
| 20.68566322 | 22.62137604 | 21.46245384 |
| 20.01751518 | 20.641325 | 19.88189697 |
| 23.35076141 | 23.2112999 | 25.14001465 |
| 20.08519363 | 19.67292213 | 18.5118618 |
| 21.1944828 | 22.64659882 | 21.33725166 |
| 19.72209167 | 19.99172974 | 20.55485725 |
| 23.02985001 | 24.73955345 | 22.74574471 |
| 19.99865341 | 19.23302269 | 19.42526436 |
| 20.20017433 | 20.45580101 | 18.98408699 |
| 17.14822388 | 17.76709366 | 17.92937469 |
| 21.26266098 | 20.33013725 | 20.54230118 |
| 22.75292206 | 24.28208542 | 24.15383339 |
| 23.19397354 | 21.72652245 | 22.72591972 |
| 21.45161819 | 20.46282387 | 20.36314011 |
| 20.53889465 | 19.85865211 | 18.63189507 |
| 18.06895828 | 19.72446442 | 18.06869888 |
| 17.23560524 | 18.52143097 | 19.15975189 |
| 17.06348228 | 18.48939323 | 17.90445709 |
| 22.55368233 | 23.70544624 | 23.00706673 |
| 21.71416664 | 22.55736732 | 21.07494164 |
| 22.5378933 | 23.15826225 | 23.213871 |
| 20.61115456 | 20.16324043 | 18.81323051 |
| 17.49323654 | 18.87150383 | 18.99982834 |
| 20.65093613 | 20.03381729 | 21.21253204 |
| 19.54408836 | 18.10961914 | 19.66662788 |
| 19.58072662 | 19.32647324 | 20.42673302 |
| 16.46549225 | 15.30872154 | 16.85375404 |
| 21.34814072 | 22.19668198 | 20.60802269 |
| 21.17317581 | 21.64278221 | 21.04663849 |
| 21.65317917 | 19.88368607 | 21.17573357 |
| 20.07636452 | 19.20054054 | 19.22994232 |

| | | |
|---|---|---|
| 21.10942841 | 20.9013195 | 19.26900291 |
| 22.01329994 | 20.42406273 | 22.19731712 |
| 25.11427689 | 26.50839615 | 26.48898888 |
| 21.73038292 | 21.66280746 | 21.92661667 |
| 19.32283592 | 18.30340385 | 19.78180504 |
| 17.74346733 | 18.852108 | 18.39803505 |
| 20.88479996 | 20.97221947 | 20.77101517 |
| 18.80208778 | 19.73223495 | 19.40366936 |
| 20.62627602 | 20.63700104 | 20.72814751 |
| 21.11445427 | 20.0279541 | 21.41976547 |
| 21.83951759 | 21.16327286 | 21.69609833 |
| 21.13985634 | 20.2258091 | 21.1537056 |
| 18.07967377 | 18.65968895 | 18.51845551 |
| 19.39838219 | 19.16063118 | 19.45859146 |
| 21.65260315 | 19.71717453 | 20.40999222 |
| 20.34759521 | 20.26403618 | 20.26494789 |
| 22.78628922 | 24.4135685 | 23.11336136 |
| 20.03301811 | 18.96876526 | 20.12714767 |
| 17.49510574 | 18.40776443 | 17.35833359 |
| 16.83656311 | 17.25930023 | 16.96440125 |
| 21.64611244 | 22.98459435 | 21.52592087 |
| 22.30824852 | 22.17082214 | 22.25549698 |
| 19.05368996 | 17.94001198 | 19.37862778 |
| 18.92637062 | 18.51812363 | 17.76599503 |
| 22.45542526 | 23.64022636 | 22.73791122 |
| 24.78427124 | 23.83865929 | 23.14277077 |
| 18.64609146 | 18.57007027 | 18.86975098 |
| 22.07587242 | 22.26635933 | 20.98582077 |
| 25.35462952 | 24.07826424 | 24.28020859 |
| 27.0484314 | 28.47859764 | 27.49710655 |
| 23.85248566 | 25.49981308 | 23.69732857 |
| 19.73521614 | 19.21774483 | 19.44039917 |
| 22.04583549 | 23.47971153 | 22.13149452 |
| 19.86881828 | 20.41953278 | 20.27147675 |
| 19.94093323 | 20.07599449 | 20.08637047 |
| 22.25308228 | 20.77365303 | 21.32798958 |
| 19.17137718 | 19.4480896 | 18.75066185 |
| 20.01668358 | 20.10443115 | 21.46653366 |
| 19.61227226 | 20.08791924 | 21.07964516 |
| 20.39022255 | 22.23962975 | 22.03327942 |
| 19.7015934 | 18.46635056 | 19.34251976 |
| 20.86410522 | 22.37884521 | 20.88220024 |
| 19.81652641 | 20.79832649 | 20.09937477 |
| 25.22033501 | 24.68093681 | 24.87893677 |
| 24.5513134 | 22.75451088 | 24.14677811 |
| 18.43669319 | 19.80329704 | 18.79954529 |
| 20.94649124 | 21.27517891 | 20.95697403 |

| | | |
|---|---|---|
| 22.17843056 | 21.60629463 | 21.92702484 |
| 18.74232292 | 17.37951088 | 18.77509308 |
| 17.31306458 | 18.15081787 | 17.89978409 |
| 21.98592567 | 23.73227692 | 23.6835022 |
| 19.59289551 | 20.26039124 | 21.22018051 |
| 21.10497093 | 20.16613197 | 20.73237991 |
| 23.89984131 | 23.36967468 | 23.25027275 |
| 20.25483322 | 19.84873581 | 19.06973457 |
| 21.64083481 | 21.73819733 | 22.62604523 |
| 20.82475281 | 21.21351242 | 20.56813049 |
| 17.1350708 | 17.68715477 | 17.44871521 |
| 20.79181862 | 20.31191444 | 21.16911697 |
| 20.60332108 | 19.69609261 | 20.13467407 |
| 21.89788628 | 20.72244835 | 21.95836258 |
| 22.58697891 | 21.57011223 | 21.91783333 |
| 20.18340492 | 21.31818199 | 21.28276825 |
| 19.7169838 | 19.58110046 | 19.39093971 |
| 19.54282379 | 20.04353523 | 18.92096519 |
| 19.41818047 | 20.41205978 | 19.64014244 |
| 19.15390968 | 19.50469971 | 19.52949905 |
| 18.308918 | 18.57273102 | 18.23029709 |
| 19.457901 | 20.66789246 | 20.88853836 |
| 21.09013367 | 22.66541481 | 22.67495155 |
| 22.5295887 | 21.65346718 | 21.80039406 |
| 20.69474792 | 22.15145874 | 20.84630203 |
| 22.15947533 | 23.54918671 | 22.94952393 |
| 22.22905731 | 23.02761269 | 21.41483498 |
| 20.0547924 | 19.75295067 | 20.20862007 |
| 21.34483719 | 22.51280975 | 21.66992188 |
| 21.79413033 | 22.44774055 | 22.01311302 |
| 18.01797676 | 19.26512718 | 19.30725479 |
| 17.16721535 | 17.58139992 | 17.55042839 |
| 18.68560791 | 19.80764389 | 18.82298279 |
| 29.31277657 | 28.20864677 | 28.19367981 |
| 22.18173981 | 20.9145565 | 21.9550705 |
| 21.12247658 | 21.56349945 | 21.39212799 |
| 26.32393456 | 25.07496834 | 25.44530487 |
| 21.06311226 | 20.13930702 | 20.61112213 |
| 20.21958542 | 19.0422287 | 20.242239 |
| 28.97517395 | 27.50387383 | 28.58972931 |
| 18.8627491 | 18.92455482 | 18.80775452 |
| 19.27923203 | 18.09592056 | 19.3331604 |
| 26.4068222 | 28.08600235 | 26.47790337 |
| 20.51801109 | 20.65081978 | 21.32961845 |
| 19.22160912 | 20.20135498 | 19.90010834 |
| 20.11807251 | 21.70795441 | 21.54730606 |
| 24.17510796 | 23.82678032 | 23.70439911 |

| | | |
|---|---|---|
| 20.91274452 | 21.28219223 | 20.71484375 |
| 18.66043472 | 18.63109398 | 17.65463066 |
| 18.92097855 | 19.89833641 | 19.41656303 |
| 17.68119049 | 18.47551537 | 17.330513 |
| 20.77110863 | 21.19856262 | 20.63551712 |
| 19.2067852 | 17.91597366 | 19.09367371 |
| 22.78213692 | 24.10361671 | 23.06999779 |
| 21.99097633 | 23.156744 | 22.44232941 |
| 22.31464386 | 21.38550758 | 21.18241882 |
| 21.71276283 | 22.93836594 | 22.8176384 |
| 21.25389481 | 20.42538071 | 20.08880997 |
| 17.25175667 | 18.14757347 | 17.56115723 |
| 23.20702744 | 21.68749428 | 22.12382889 |
| 18.08070183 | 16.95807457 | 17.68369293 |
| 19.19303894 | 19.24809074 | 19.08808136 |
| 18.32977676 | 19.67326927 | 19.19804001 |
| 21.31043625 | 21.65254211 | 21.91922951 |
| 19.56293678 | 19.662117 | 18.51083374 |
| 20.96810913 | 22.34129524 | 21.21629143 |
| 20.43912697 | 21.0241642 | 20.16851807 |
| 21.98131561 | 21.83692169 | 20.67477608 |
| 20.43048668 | 21.09602356 | 21.48685837 |
| 21.07287598 | 20.97515297 | 21.25139809 |
| 19.43988991 | 19.17460632 | 18.80144691 |
| 17.20256424 | 18.27377129 | 17.41981506 |
| 21.78257751 | 20.94729996 | 21.30846596 |
| 18.05941582 | 17.6960125 | 17.70810509 |
| 21.28065872 | 22.15005684 | 21.64506912 |
| 21.83934784 | 21.23351669 | 20.36440468 |
| 20.86979485 | 21.0885601 | 20.16392708 |
| 23.13848686 | 22.28703117 | 21.93611145 |
| 21.14255714 | 22.12663651 | 21.87682724 |
| 17.67021561 | 18.17383003 | 17.78478432 |
| 22.32118034 | 22.4628067 | 21.76113129 |
| 18.0110321 | 18.60440445 | 18.30834389 |
| 18.07135201 | 17.32628632 | 17.80372238 |
| 19.25819778 | 18.38678169 | 19.1711216 |
| 20.57730484 | 19.8682785 | 20.40452957 |
| 20.90248489 | 21.8210144 | 21.14171982 |
| 20.62234306 | 19.58572578 | 20.99783516 |
| 20.61387253 | 21.35846519 | 19.93640518 |
| 20.28073311 | 19.49238014 | 19.77789497 |
| 18.727005 | 18.79413223 | 18.21529961 |
| 27.88888168 | 26.27811432 | 27.38828278 |
| 19.89039803 | 20.79769707 | 21.30456352 |
| 22.17074394 | 21.10200119 | 21.73649597 |
| 21.47004509 | 22.27606773 | 21.07359123 |

| | | |
|---|---|---|
| 20.30926895 | 21.45667076 | 21.04494476 |
| 24.320261 | 23.77276993 | 22.95632172 |
| 21.26157761 | 19.95856857 | 20.96229744 |
| 20.90492439 | 20.91040421 | 21.16294098 |
| 20.54410172 | 20.48077965 | 20.8476963 |
| 20.00605583 | 20.16317558 | 19.92654419 |
| 18.75066948 | 19.71772385 | 19.1644249 |
| 18.85200691 | 18.78484726 | 18.45450401 |
| 19.83025932 | 19.52705765 | 19.64256859 |
| 22.00468254 | 21.66024971 | 21.17238617 |
| 17.8813591 | 17.76275253 | 17.60064888 |
| 23.35845566 | 24.61794853 | 23.25490379 |
| 21.85643005 | 20.94876862 | 22.21126366 |
| 18.02444267 | 18.30563927 | 17.31930733 |
| 23.49922562 | 23.45883369 | 23.41653252 |
| 21.17026138 | 21.71991158 | 20.87261009 |
| 21.26410294 | 20.91649818 | 21.52276993 |
| 18.99547958 | 18.98594093 | 19.17027092 |
| 17.89612579 | 18.68128586 | 18.34374809 |
| 18.7786293 | 19.45856094 | 18.91096878 |
| 21.79955482 | 20.29042816 | 21.07337952 |
| 20.48710442 | 21.51065636 | 21.59361649 |
| 18.02243805 | 18.15089989 | 18.26003075 |
| 21.29859543 | 21.41887474 | 21.29617882 |
| 19.83934212 | 19.18234253 | 18.96761131 |
| 20.5232048 | 20.87933731 | 21.64773178 |
| 21.25364876 | 20.45267105 | 19.84764671 |
| 26.05726051 | 24.82975197 | 26.09107208 |
| 19.01433563 | 19.78777504 | 19.5472908 |
| 19.60830307 | 19.83079529 | 20.12138939 |
| 19.42499352 | 19.05794525 | 19.40329361 |
| 22.60656738 | 22.86251068 | 22.40386391 |
| 23.97307396 | 24.2562542 | 24.08386803 |
| 21.42081451 | 22.04481506 | 20.85411453 |
| 24.69426918 | 26.17678261 | 26.01891327 |
| 21.58130646 | 20.91056824 | 20.23587036 |
| 20.90661049 | 21.72504044 | 20.87339973 |
| 20.87012863 | 20.23058319 | 20.9335537 |
| 18.20907402 | 18.09083176 | 17.50414085 |
| 21.3328743 | 21.18775177 | 20.26741409 |
| 20.61132431 | 21.307621 | 20.99445915 |
| 22.36647224 | 21.46411133 | 22.51937294 |
| 19.12228203 | 19.22447777 | 18.41747475 |
| 18.19636726 | 19.24161148 | 18.71790123 |
| 22.33256912 | 21.43826294 | 22.84759712 |
| 19.53296089 | 19.81513023 | 19.48208237 |
| 19.65151405 | 19.09653091 | 19.65873909 |

| | | |
|---|---|---|
| 21.09210777 | 20.22652054 | 20.49262238 |
| 21.51196861 | 21.36635971 | 20.33039284 |
| 21.65487862 | 21.14748764 | 21.42580032 |
| 18.09939957 | 17.84185028 | 17.90945625 |
| 19.87187958 | 19.21471786 | 20.08185196 |
| 22.164114 | 20.88835144 | 21.51820564 |
| 26.62322235 | 25.45722198 | 26.54405975 |
| 22.9981308 | 22.13621902 | 22.95876884 |
| 21.75906372 | 21.70793343 | 21.57089043 |
| 21.72052956 | 20.63742065 | 21.71481514 |
| 22.98646736 | 23.73381805 | 23.02676582 |
| 21.28928185 | 22.43639755 | 21.39138031 |
| 21.45777893 | 21.59999275 | 22.16581154 |
| 22.19429016 | 21.46009254 | 21.68533516 |
| 21.61091614 | 22.15598106 | 21.23298645 |
| 21.7878685 | 21.0896244 | 21.68533707 |
| 23.25217438 | 22.46634674 | 22.88313293 |
| 20.08716583 | 19.75529671 | 19.84588814 |
| 20.02121162 | 20.36252213 | 20.96450424 |
| 21.35638809 | 21.84938812 | 20.71653175 |
| 22.24681282 | 21.80927467 | 22.11850929 |
| 21.58023834 | 21.74236488 | 21.26466179 |
| 22.42127991 | 22.60618973 | 22.74663734 |
| 21.33893394 | 20.84021759 | 20.17980957 |
| 21.63856316 | 22.62888718 | 21.70850563 |
| 22.48749542 | 23.0743618 | 22.99138641 |
| 20.63778687 | 20.59926224 | 20.66822815 |
| 19.84517288 | 20.3157196 | 20.25061607 |
| 25.90510559 | 26.6287384 | 25.9883976 |
| 23.77270126 | 23.55209732 | 23.65382957 |
| 16.98162651 | 16.52788544 | 17.62277603 |
| 18.20044708 | 18.03508186 | 17.38484764 |
| 20.58694077 | 21.1245079 | 20.67059135 |
| 20.28287697 | 20.01914024 | 20.6433773 |
| 22.8757534 | 23.02460289 | 22.13511848 |
| 18.67689514 | 18.56936264 | 18.82647514 |
| 21.38747025 | 22.05306435 | 21.47319031 |
| 21.04117584 | 20.33936501 | 21.00290108 |
| 20.38449097 | 21.11662865 | 20.10457802 |
| 20.81769562 | 21.39683151 | 20.78592682 |
| 24.41226959 | 25.2071743 | 25.20682907 |
| 20.17768669 | 21.03167725 | 21.01503754 |
| 21.75734901 | 20.8733654 | 21.77004051 |
| 19.58409691 | 20.00886726 | 19.36908913 |
| 20.6219368 | 20.47816658 | 20.89425278 |
| 23.11774254 | 22.57499504 | 22.5526886 |
| 22.13631248 | 22.71933556 | 22.93053246 |

| | | |
|---|---|---|
| 19.92043304 | 20.41614914 | 20.66047287 |
| 20.7665844 | 20.81005096 | 21.01065636 |
| 23.72357941 | 24.05817604 | 23.9955368 |
| 17.76156235 | 18.69065857 | 17.91918755 |
| 21.66974258 | 22.16888237 | 22.24317932 |
| 19.1539135 | 18.73336792 | 19.25997543 |
| 22.43685341 | 22.98027229 | 22.06184387 |
| 22.25020218 | 23.01441574 | 22.64565277 |
| 24.23444939 | 23.66261482 | 24.18252945 |
| 20.09253311 | 20.76956367 | 20.59184647 |
| 19.91332245 | 20.35252571 | 20.36828995 |
| 27.43546104 | 26.07575226 | 26.96864319 |
| 22.28319168 | 22.04668236 | 21.80861473 |
| 18.64242935 | 18.70583916 | 18.85565186 |
| 20.47199249 | 21.67190552 | 20.93456078 |
| 21.16502762 | 21.66019249 | 21.8143158 |
| 19.20632935 | 18.95409393 | 18.95458412 |
| 17.2892704 | 18.16762352 | 18.14564896 |
| 16.2121048 | 16.26719856 | 15.97057533 |
| 19.88791466 | 18.96186829 | 19.67636108 |
| 18.44734192 | 19.36558342 | 18.99689102 |
| 22.43385887 | 22.70234108 | 22.99913216 |
| 22.39128304 | 21.84189415 | 22.08821106 |
| 24.99876404 | 25.75179482 | 25.34057617 |
| 21.25930214 | 21.28305435 | 20.7054882 |
| 20.97677231 | 21.81854057 | 21.59252357 |
| 16.02091217 | 16.85506058 | 16.49775124 |
| 19.6192894 | 19.89099884 | 19.33903313 |
| 22.8829422 | 22.10087776 | 22.18039513 |
| 20.10884285 | 19.26086998 | 19.37659836 |
| 21.24015808 | 21.9351368 | 21.28572273 |
| 23.40021133 | 24.54370689 | 23.35364532 |
| 19.30249786 | 19.88921547 | 19.79762077 |
| 20.03186226 | 19.02695465 | 19.53189659 |
| 25.81998062 | 26.80185318 | 26.48599815 |
| 19.3498745 | 19.03871536 | 19.29761696 |
| 21.00390625 | 21.7063961 | 20.79888725 |
| 22.64233589 | 22.38905907 | 21.5491581 |
| 18.06822205 | 18.27587891 | 17.92410278 |
| 22.27155495 | 21.47003174 | 21.82452774 |
| 21.10542679 | 20.2351799 | 20.75587654 |
| 16.08060837 | 16.66156006 | 16.18765831 |
| 23.18816185 | 24.02974701 | 23.61535263 |
| 19.85047913 | 20.11278915 | 20.46234703 |
| 18.78196526 | 18.36842155 | 18.63635445 |
| 19.65846252 | 19.93336105 | 18.92235565 |
| 17.56303024 | 18.51992035 | 18.02395439 |

| | | |
|---|---|---|
| 21.60433578 | 21.38140488 | 21.90699959 |
| 17.78322792 | 18.07098198 | 18.09232521 |
| 17.670084 | 18.1770134 | 17.83859825 |
| 18.28801537 | 18.63441658 | 18.4968586 |
| 23.65985489 | 24.24521446 | 24.01428986 |
| 21.33934212 | 22.30350685 | 21.88149452 |
| 21.532547 | 20.95884323 | 21.69874191 |
| 18.67600822 | 18.99646187 | 19.00465584 |
| 17.81517982 | 18.14878082 | 18.25851631 |
| 19.80165482 | 20.8337059 | 20.2654705 |
| 20.94251633 | 21.12100601 | 20.34301567 |
| 20.52611351 | 20.35741425 | 19.96378136 |
| 19.48711205 | 19.35795975 | 18.982687 |
| 22.41451836 | 22.41178894 | 22.5364933 |
| 18.23330307 | 18.08015251 | 17.54405975 |
| 20.84103012 | 20.48828506 | 20.83248711 |
| 18.32995224 | 17.58969116 | 18.21786118 |
| 19.84294319 | 20.164711 | 19.83756065 |
| 24.36648178 | 25.00156212 | 25.36611557 |
| 21.37791443 | 21.36597061 | 21.05096054 |
| 23.55480766 | 23.36586952 | 23.71034431 |
| 22.06995964 | 22.92193222 | 22.3051796 |
| 21.69087029 | 21.87935448 | 21.60668182 |
| 21.14969063 | 21.50886345 | 21.17689133 |
| 22.38863945 | 21.98847008 | 23.05545235 |
| 23.11584663 | 23.35638237 | 23.35851669 |
| 19.51174736 | 20.14608955 | 19.84696198 |
| 23.79047775 | 23.39611053 | 23.64431953 |
| 20.29942131 | 20.02274513 | 19.60096741 |
| 21.6579361 | 21.36534119 | 20.63843918 |
| 25.0306015 | 24.09448814 | 24.94700432 |
| 22.1384697 | 22.43584061 | 22.76984787 |
| 20.39125061 | 21.04063034 | 20.91322136 |
| 23.02525902 | 23.01620102 | 22.80544281 |
| 20.13290215 | 19.52269936 | 20.17969704 |
| 21.34692764 | 21.19437408 | 20.66783524 |
| 21.63331223 | 22.3680706 | 22.28553772 |
| 25.4856205 | 24.64928436 | 24.71110725 |
| 25.15768051 | 25.47888184 | 25.290905 |
| 25.36854744 | 24.34095764 | 25.23975754 |
| 17.97905731 | 18.26911354 | 18.69152069 |
| 19.48322678 | 19.48682022 | 18.73417664 |
| 21.29482269 | 21.72942924 | 21.19537354 |
| 19.21233368 | 18.96157646 | 18.82146835 |
| 17.86882973 | 18.51732826 | 17.71563339 |
| 21.12081337 | 20.35076523 | 21.15208054 |
| 22.27871513 | 21.30756378 | 22.01174927 |

| | | |
|---|---|---|
| 18.89163017 | 18.88219833 | 18.79459 |
| 20.30213165 | 21.13084221 | 20.34242821 |
| 24.80766869 | 23.97055626 | 25.03871346 |
| 20.55424309 | 21.37930489 | 20.531847 |
| 19.29553795 | 18.94128227 | 19.34210396 |
| 24.20125198 | 24.14402008 | 23.26767921 |
| 21.9151783 | 22.34482193 | 21.79192543 |
| 20.01535034 | 19.98553848 | 20.8646946 |
| 19.95387077 | 19.17773438 | 20.00035286 |
| 21.93453217 | 21.86816025 | 21.09755898 |
| 22.21122742 | 22.79038429 | 22.42730141 |
| 21.1486454 | 21.00963783 | 21.57510376 |
| 21.64968681 | 20.89950371 | 21.61542702 |
| 21.42313194 | 20.55924225 | 21.04389381 |
| 22.94516945 | 22.08867455 | 22.14477158 |
| 19.44784355 | 19.76060486 | 19.44655228 |
| 19.93386459 | 20.44805527 | 20.50084305 |
| 22.96164131 | 23.18903923 | 22.35777664 |
| 22.15228462 | 22.13334465 | 22.09323883 |
| 21.81321526 | 21.03980064 | 21.02358055 |
| 26.67323685 | 25.74914169 | 26.25084877 |
| 22.14014244 | 21.36829948 | 21.99047089 |
| 19.77955437 | 18.96839333 | 19.08640862 |
| 22.80620384 | 23.54782295 | 22.92572784 |
| 24.93810654 | 25.10678482 | 25.12075996 |
| 23.44274139 | 23.37534523 | 23.44348145 |
| 19.51918602 | 19.82491875 | 19.31713104 |
| 22.59156227 | 23.02644348 | 22.97873497 |
| 20.83696556 | 21.1269474 | 20.97743988 |
| 20.76550674 | 20.70188141 | 19.94185638 |
| 20.50027847 | 21.01197624 | 20.38814926 |
| 18.76278496 | 19.36368561 | 18.96660233 |
| 21.34030342 | 21.92052269 | 21.40396881 |
| 18.74273682 | 18.44547272 | 18.85374832 |
| 19.96502304 | 20.33156967 | 19.70830154 |
| 22.77474403 | 22.550354 | 22.38895607 |
| 22.47639847 | 22.94518661 | 22.40879822 |
| 24.17460823 | 23.86273956 | 23.74323082 |
| 21.97549057 | 22.43229675 | 22.20680618 |
| 22.98017883 | 22.10354424 | 22.44092369 |
| 18.59960365 | 19.36118698 | 19.01630592 |
| 25.60854721 | 24.8391819 | 25.2843895 |
| 22.06228256 | 22.73773766 | 22.04275322 |
| 22.87575912 | 22.65535545 | 22.11891174 |
| 21.69041634 | 21.29690742 | 21.33790398 |
| 17.53696632 | 18.26060295 | 17.98050117 |
| 21.3315773 | 20.74292755 | 20.89582253 |

| | | |
|---|---|---|
| 18.97387505 | 18.80761719 | 19.30230141 |
| 22.41063309 | 21.52833557 | 21.87294197 |
| 21.32851219 | 22.00610542 | 21.42013931 |
| 21.71960449 | 21.77627754 | 21.43679428 |
| 23.97554207 | 23.97908401 | 23.96232414 |
| 24.68558311 | 23.83766556 | 24.56233597 |
| 23.48496819 | 22.63013268 | 22.90340996 |
| 20.16552925 | 19.48962784 | 20.21412468 |
| 24.19719124 | 23.30839157 | 23.62582016 |
| 20.43029594 | 20.78419685 | 19.9431076 |
| 19.65936661 | 19.63475609 | 20.08268738 |
| 21.68220711 | 20.89441681 | 21.15070152 |
| 19.95616722 | 20.39044189 | 19.6799469 |
| 21.32371521 | 21.5958519 | 21.40591812 |
| 30.23952293 | 29.02241898 | 29.78243446 |
| 24.77715302 | 24.72953987 | 25.11629486 |
| 25.45326996 | 26.26761055 | 25.88600731 |
| 17.52572823 | 17.68569565 | 18.0637455 |
| 18.49554443 | 18.09238815 | 18.11102486 |
| 19.99123383 | 19.81991959 | 19.68097305 |
| 19.52809906 | 19.55386353 | 19.05255318 |
| 21.95432854 | 20.92345047 | 21.50243759 |
| 22.12661171 | 21.37325287 | 22.05051994 |
| 16.95406532 | 17.279953 | 16.97251129 |
| 19.21941566 | 19.49575233 | 19.17785835 |
| 22.00852585 | 21.93270493 | 21.89454842 |
| 18.74862862 | 18.8386898 | 19.38846397 |
| 19.02795982 | 19.54792404 | 19.22952843 |
| 22.26032066 | 21.86143112 | 22.11925697 |
| 19.47458076 | 19.99228096 | 19.70848656 |
| 23.00409508 | 23.54493713 | 22.65463257 |
| 21.53936005 | 21.84267807 | 21.58166504 |
| 17.77876663 | 18.26711464 | 17.57178879 |
| 22.87732887 | 23.52482224 | 22.90229797 |
| 20.34752655 | 20.14487457 | 19.72074509 |
| 21.58906364 | 21.11240578 | 21.5279541 |
| 18.23053551 | 17.94857979 | 18.01891899 |
| 23.37169456 | 23.14414787 | 23.12287331 |
| 22.98664093 | 23.60699081 | 22.857481 |
| 19.15028381 | 18.87817955 | 19.32156181 |
| 18.30458641 | 19.05257225 | 18.31405449 |
| 22.14247894 | 21.76510048 | 22.0678196 |
| 19.40863037 | 18.99344444 | 18.93554688 |
| 21.53998756 | 21.72773933 | 20.98370934 |
| 22.54411316 | 22.80848122 | 22.47560692 |
| 24.63599777 | 24.76058769 | 25.23612785 |
| 20.53165054 | 20.31570435 | 20.34537888 |

| | | |
|---|---|---|
| 22.67131424 | 23.46631813 | 23.26877213 |
| 20.44035912 | 20.40338898 | 20.35440254 |
| 29.6687088 | 28.60385132 | 29.28472519 |
| 22.41357231 | 22.29306412 | 22.47945976 |
| 21.10028458 | 21.17760086 | 21.19351768 |
| 24.9944706 | 25.2543602 | 25.98638344 |
| 20.01734543 | 20.48634338 | 20.7639904 |
| 22.27482605 | 22.89372063 | 22.34730148 |
| 18.87622261 | 19.13341141 | 19.03528595 |
| 22.18634796 | 21.60860634 | 21.46559906 |
| 23.88004875 | 24.13247871 | 23.3210659 |
| 21.50988388 | 21.47849655 | 21.81065369 |
| 26.69708443 | 27.48021317 | 26.74765015 |
| 18.65343094 | 19.00975418 | 19.19409752 |
| 24.28660774 | 24.66280746 | 24.71995354 |
| 18.16769028 | 18.97534943 | 18.48129082 |
| 21.09314346 | 21.8161068 | 21.27838898 |
| 19.57422447 | 20.12848282 | 20.39976883 |
| 23.47994614 | 22.97253799 | 23.56334496 |
| 22.11249542 | 22.68011665 | 22.40504646 |
| 21.25302887 | 21.56395721 | 21.49824333 |
| 19.04212952 | 19.07200813 | 18.93525124 |
| 19.45151138 | 19.88187981 | 19.4507885 |
| 18.57874489 | 17.79849434 | 18.09210587 |
| 22.51619339 | 22.35498047 | 22.90716743 |
| 21.05465889 | 21.22753525 | 21.39852333 |
| 21.56056404 | 21.76382828 | 21.32081795 |
| 16.97774124 | 16.87792206 | 16.90637589 |
| 19.41268349 | 20.16483498 | 19.56747055 |
| 19.19659805 | 19.24632072 | 19.60859489 |
| 22.49786949 | 23.07790375 | 22.20082855 |
| 18.66154099 | 18.80972481 | 18.81162453 |
| 20.66023445 | 20.83348846 | 20.60885048 |
| 23.62364006 | 23.61970139 | 23.66110611 |
| 23.0161705 | 23.96879005 | 23.27105331 |
| 19.17449188 | 18.98440361 | 18.52846909 |
| 17.20747948 | 17.78765678 | 17.94852257 |
| 19.78195381 | 20.08192062 | 19.79099655 |
| 23.14823723 | 23.77402306 | 22.96790886 |
| 21.25678825 | 21.24650764 | 21.10185051 |
| 23.30202675 | 23.62327194 | 23.26382828 |
| 23.24144554 | 23.75708199 | 23.37826157 |
| 23.69256592 | 23.13087273 | 23.89128876 |
| 19.22436523 | 19.10270309 | 19.17402649 |
| 24.49026299 | 25.03812981 | 24.63037109 |
| 25.38721657 | 24.807724 | 25.26304626 |
| 20.07566833 | 19.91095161 | 20.35755348 |

| | | |
|---|---|---|
| 24.9528923 | 24.3726635 | 25.1125164 |
| 23.06031036 | 23.23257828 | 23.12540817 |
| 20.46112442 | 21.12054825 | 21.04579163 |
| 22.75957108 | 22.67698288 | 22.16044807 |
| 22.83771324 | 22.902174 | 22.49259758 |
| 20.83909416 | 20.50222206 | 20.72746086 |
| 17.64861488 | 18.17954636 | 17.75279808 |
| 21.17994308 | 20.87986755 | 20.98954964 |
| 20.33042336 | 20.28854179 | 20.23142624 |
| 22.35482407 | 22.05207443 | 21.5701046 |
| 19.5916481 | 20.1564064 | 20.02606201 |
| 17.93488121 | 18.26118851 | 17.9581871 |
| 19.63836861 | 19.52583313 | 20.16994476 |
| 22.01715279 | 21.30525017 | 21.22017097 |
| 18.38547516 | 18.56522369 | 18.3244915 |
| 20.23431206 | 20.70760155 | 19.89967155 |
| 21.65023613 | 21.71128654 | 21.25181198 |
| 20.57911301 | 20.67718697 | 20.19821358 |
| 24.32488632 | 24.07648087 | 24.69094086 |
| 22.42297554 | 22.5381279 | 21.70508766 |
| 21.54735756 | 21.32940674 | 21.67385292 |
| 25.80959892 | 25.76642036 | 25.01581001 |
| 22.6759491 | 23.13153648 | 22.57589912 |
| 22.48508835 | 22.56624794 | 22.64880371 |
| 21.75072098 | 22.05077553 | 21.58739853 |
| 21.38769913 | 21.72909164 | 22.01780128 |
| 20.76256752 | 20.84387779 | 20.54099274 |
| 18.7165947 | 18.46613121 | 18.53252983 |
| 22.74118614 | 23.08990479 | 23.02519417 |
| 23.26836967 | 22.92413521 | 23.54212379 |
| 22.76435089 | 22.81513405 | 22.66562843 |
| 20.09986687 | 20.02817726 | 19.87399673 |
| 21.56085777 | 21.18406868 | 21.33348465 |
| 20.89946747 | 20.84592056 | 20.87529373 |
| 21.40341759 | 21.65131569 | 22.00494957 |
| 20.75090599 | 20.88192368 | 20.17923546 |
| 19.94776917 | 20.09848022 | 19.6217289 |
| 19.24606323 | 19.29258919 | 19.0626297 |
| 20.87018204 | 21.4359169 | 21.04410553 |
| 22.00248528 | 22.77764702 | 22.77228355 |
| 20.99403763 | 20.34941673 | 20.87338638 |
| 21.41758347 | 20.8361454 | 21.37989998 |
| 23.11462784 | 22.68918991 | 22.84911156 |
| 22.87469292 | 23.20266151 | 23.1388607 |
| 24.43134117 | 23.89638519 | 23.62220764 |
| 23.85442543 | 24.36165237 | 23.47807693 |
| 22.30841446 | 21.57264137 | 22.16099548 |

| | | |
|---|---|---|
| 20.59836388 | 21.32566452 | 20.76185799 |
| 24.67432404 | 23.98435211 | 24.73409462 |
| 19.57827377 | 19.24656677 | 19.8190403 |
| 23.75711632 | 24.30924034 | 24.17991257 |
| 18.3185215 | 18.86142921 | 18.76695824 |
| 19.86731339 | 19.38552856 | 20.10128021 |
| 21.11578369 | 21.25467491 | 21.29367828 |
| 20.68396378 | 20.89272499 | 20.22975731 |
| 23.14799118 | 22.46789742 | 23.03763008 |
| 20.86317253 | 20.79771423 | 20.72924232 |
| 27.54364777 | 27.52947807 | 27.6410141 |
| 21.75264168 | 22.3707428 | 21.64723015 |
| 21.06028748 | 21.15041733 | 21.20214462 |
| 21.8099308 | 22.26828003 | 21.67001915 |
| 29.2915287 | 28.23448944 | 28.96630478 |
| 24.37034988 | 23.39453125 | 23.92533493 |
| 18.76110268 | 18.97765732 | 18.37243462 |
| 18.60292053 | 18.74462318 | 19.12407684 |
| 21.31754494 | 21.14881706 | 21.31921959 |
| 22.4440136 | 22.86822319 | 23.00125885 |
| 20.41774368 | 20.74865913 | 20.78082657 |
| 24.15305328 | 23.84908104 | 24.09456444 |
| 27.48052979 | 28.15312767 | 27.63928986 |
| 18.60497284 | 18.80791092 | 18.67965317 |
| 19.93384361 | 20.52427292 | 20.11375809 |
| 23.64088821 | 24.20311165 | 23.93727875 |
| 25.85716057 | 25.10707474 | 25.38211632 |
| 18.20569038 | 18.46025276 | 18.65090179 |
| 19.39958 | 18.85871124 | 19.08318138 |
| 21.5812664 | 22.03577805 | 21.98534584 |
| 21.39850235 | 21.80753708 | 21.97219658 |
| 22.38695908 | 21.67588997 | 21.99975014 |
| 23.50902367 | 23.22525787 | 22.94179344 |
| 23.2071476 | 23.43147469 | 23.5219841 |
| 18.78891945 | 18.18083763 | 18.44175148 |
| 20.23649406 | 20.64003372 | 19.91039276 |
| 23.13101768 | 23.30051422 | 23.17269707 |
| 16.47783852 | 17.03150558 | 16.85939407 |
| 20.16136932 | 19.93673706 | 19.89426994 |
| 20.37015915 | 21.16443253 | 20.7201767 |
| 21.26218414 | 21.43823624 | 21.56710434 |
| 22.1539669 | 21.63493347 | 21.33547783 |
| 22.00864983 | 21.88745308 | 21.87169647 |
| 21.97908211 | 22.21835327 | 22.52655411 |
| 19.45546532 | 19.89850616 | 19.61351585 |
| 22.49263 | 22.9799099 | 23.04281998 |
| 22.21507645 | 22.52405167 | 22.22725296 |

| | | |
|---|---|---|
| 26.63606834 | 27.40813065 | 27.27080154 |
| 23.98545265 | 24.66564941 | 24.09658623 |
| 20.57149506 | 20.11226845 | 20.5111351 |
| 21.57794952 | 21.875 | 21.45885277 |
| 24.07276535 | 23.75422096 | 23.76881218 |
| 21.90768051 | 21.8730793 | 22.08480453 |
| 25.83515549 | 26.36255455 | 26.16389275 |
| 21.53581047 | 21.56717491 | 22.18096924 |
| 19.58481407 | 19.31723213 | 19.52442551 |
| 21.62376595 | 22.27462769 | 21.94866753 |
| 22.03990746 | 21.55973434 | 21.69991875 |
| 19.88751602 | 19.75390816 | 19.88704681 |
| 20.94668579 | 20.43544197 | 21.1260376 |
| 19.91899109 | 19.94043922 | 20.0522213 |
| 23.54006958 | 24.10453606 | 23.6594677 |
| 23.4917717 | 23.37068939 | 23.47737885 |
| 21.85073471 | 21.74632072 | 21.70486832 |
| 22.44620514 | 22.66630363 | 22.71333885 |
| 18.49650574 | 19.17470551 | 18.83255196 |
| 18.58565712 | 18.21626854 | 18.1673317 |
| 23.36655998 | 23.93366814 | 23.56282043 |
| 22.39240074 | 21.82721329 | 22.19211578 |
| 27.03600311 | 26.22762108 | 26.83629417 |
| 23.18126488 | 22.5791111 | 23.10712433 |
| 21.03042603 | 20.31563187 | 20.74264908 |
| 18.314785 | 18.42068863 | 18.67275238 |
| 22.48812866 | 22.89315987 | 22.07465553 |
| 21.54510689 | 21.57712555 | 21.72424507 |
| 22.4554882 | 22.79281998 | 22.6252346 |
| 19.21401787 | 19.68803596 | 19.31097412 |
| 19.11954117 | 18.72034836 | 18.53209686 |
| 22.54043579 | 22.67902565 | 22.93979263 |
| 25.27529144 | 25.3854084 | 25.60225868 |
| 20.35502052 | 20.8849926 | 20.46761131 |
| 21.31733513 | 21.56993866 | 21.5603981 |
| 19.80093384 | 20.02653885 | 19.445755 |
| 23.20697594 | 23.73932648 | 23.5605526 |
| 19.12561035 | 19.57925224 | 19.37248039 |
| 19.20517349 | 19.43454361 | 19.22827339 |
| 27.77211571 | 27.07948112 | 27.13673973 |
| 20.46092796 | 20.68023872 | 20.70407677 |
| 21.32510948 | 21.76234245 | 21.74332047 |
| 21.25238609 | 21.15832138 | 21.69402504 |
| 19.8969574 | 20.25138474 | 19.70415688 |
| 23.27778244 | 23.46562958 | 23.33938789 |
| 25.84592819 | 25.19965172 | 25.8120575 |
| 24.44744301 | 25.04941368 | 24.63235092 |

| | | |
|---|---|---|
| 21.71917915 | 21.13970947 | 21.55348969 |
| 23.98274231 | 23.29733467 | 23.76217651 |
| 20.59537697 | 20.15779114 | 20.70982933 |
| 20.38386917 | 20.43905258 | 20.37435913 |
| 20.23563194 | 20.94374847 | 20.47242165 |
| 19.59159088 | 19.40501022 | 19.92433167 |
| 24.19129944 | 24.56921196 | 24.35744667 |
| 22.61759949 | 23.22010231 | 22.72879601 |
| 25.75434303 | 25.5483799 | 25.0698204 |
| 23.16054153 | 22.90011597 | 22.61282158 |
| 18.86643028 | 18.41979027 | 18.73752785 |
| 22.60964966 | 23.05955887 | 22.79131889 |
| 22.25715446 | 21.88212013 | 21.99314308 |
| 20.84511948 | 20.40286064 | 20.86244202 |
| 20.14663506 | 20.65277863 | 20.53126335 |
| 23.53804207 | 23.92990112 | 23.36968994 |
| 23.46935272 | 23.06395721 | 23.17069054 |
| 21.67711258 | 21.51283836 | 21.89428139 |
| 23.51153755 | 23.06800842 | 23.12924194 |
| 24.83489227 | 24.24164391 | 24.18950462 |
| 18.94480515 | 18.75058174 | 19.28440475 |
| 22.6967144 | 22.98534775 | 23.07629776 |
| 21.22975731 | 20.91651344 | 21.19453239 |
| 18.72802734 | 18.84521675 | 18.6943779 |
| 22.78467941 | 22.99033165 | 22.92133904 |
| 22.45732117 | 22.58663559 | 22.6670723 |
| 25.1331768 | 24.94894028 | 25.23258209 |
| 20.06446838 | 20.11618233 | 20.21636009 |
| 18.65936661 | 18.95433998 | 18.78686523 |
| 19.27861595 | 19.14875793 | 18.97946167 |
| 23.32207298 | 23.94582558 | 23.64793587 |
| 30.986866 | 30.06046295 | 30.70681763 |
| 24.35680962 | 24.46386909 | 24.55784416 |
| 21.34788322 | 21.33905792 | 21.73700523 |
| 20.55998039 | 21.04407883 | 20.41703033 |
| 23.40532112 | 22.88547707 | 22.98353958 |
| 20.3087883 | 20.74042511 | 20.57017517 |
| 20.5483532 | 20.26288414 | 20.6961441 |
| 23.76098442 | 23.72241974 | 23.37843323 |
| 19.01443291 | 18.99586105 | 19.00307274 |
| 18.56977081 | 18.40666771 | 18.62175179 |
| 24.15798187 | 23.63782692 | 23.97736549 |
| 21.79982376 | 22.31827545 | 22.50419807 |
| 19.58251572 | 19.52381706 | 19.08808708 |
| 21.91542435 | 22.32086372 | 21.96501732 |
| 21.13071442 | 21.63710213 | 21.47131538 |
| 22.79892349 | 22.54174423 | 23.02005768 |

| | | |
|---|---|---|
| 21.20082092 | 21.28863144 | 21.37060547 |
| 21.98955345 | 22.30200768 | 22.1010952 |
| 23.67239571 | 24.05070114 | 23.83083344 |
| 23.27545738 | 22.59058571 | 22.89674759 |
| 23.40372276 | 24.10937309 | 23.79989243 |
| 20.72519302 | 21.16670418 | 21.21472168 |
| 21.35883522 | 21.37416458 | 21.52272987 |
| 22.007061 | 22.65677261 | 22.29859734 |
| 22.50788879 | 22.7597847 | 22.66001129 |
| 23.69796371 | 23.17230034 | 23.52447128 |
| 27.95998573 | 27.0982132 | 27.46161842 |
| 21.12878227 | 21.26998711 | 21.36081696 |
| 24.31007004 | 24.41435051 | 24.40745926 |
| 21.4655304 | 21.87814522 | 21.36493301 |
| 21.62569237 | 21.33999825 | 21.35194778 |
| 22.34490585 | 21.9309082 | 22.08756065 |
| 22.01584244 | 22.04484558 | 22.49133873 |
| 19.82958412 | 19.94363403 | 19.68497658 |
| 17.63157272 | 18.10967445 | 17.73995209 |
| 20.27928162 | 20.65076256 | 20.49493599 |
| 21.82065392 | 22.17281532 | 22.24760628 |
| 22.72040749 | 22.262146 | 22.81941795 |
| 23.42548561 | 23.65789986 | 23.55611992 |
| 21.28117561 | 21.50997162 | 21.28814125 |
| 22.72363472 | 23.24843025 | 22.71735764 |
| 19.52135277 | 19.44103432 | 19.39098167 |
| 25.32780075 | 25.12514687 | 25.13740158 |
| 20.1361351 | 20.12257385 | 20.3745575 |
| 22.48942184 | 22.14416313 | 22.4440403 |
| 19.88968849 | 19.82370567 | 19.89492035 |
| 24.9131012 | 25.37119484 | 25.2159729 |
| 20.06963539 | 19.89779472 | 20.22602081 |
| 18.92010117 | 18.66564751 | 19.04146385 |
| 23.02503204 | 23.27019691 | 23.29039955 |
| 24.63466072 | 25.31510544 | 24.83189201 |
| 20.68630409 | 20.36016273 | 20.25056648 |
| 22.08532524 | 21.58473396 | 21.48322105 |
| 25.03763962 | 24.78082275 | 24.55889702 |
| 20.69325638 | 20.83712196 | 20.99158096 |
| 27.16279602 | 27.47354889 | 27.10181046 |
| 20.51890373 | 20.53096008 | 20.93663216 |
| 19.50180817 | 19.71815872 | 19.38186264 |
| 22.31317329 | 22.71968842 | 22.85967255 |
| 23.26406097 | 23.4734478 | 22.88862991 |
| 20.21696091 | 20.64876366 | 20.01678658 |
| 24.7031765 | 24.2941761 | 24.80895424 |
| 21.56833267 | 22.04683113 | 21.86699104 |

| | | |
|---|---|---|
| 24.4966774 | 23.80360603 | 24.15301514 |
| 19.45388794 | 19.53299904 | 19.31253242 |
| 21.82295418 | 22.12169456 | 21.8137722 |
| 19.13829231 | 19.30426598 | 19.43927383 |
| 24.91683769 | 25.27522278 | 24.9076252 |
| 19.32014275 | 18.93895721 | 19.44674873 |
| 22.8896637 | 23.02226639 | 23.1526413 |
| 21.36932755 | 21.38852692 | 21.46087646 |
| 21.86297226 | 21.62317467 | 21.81332397 |
| 24.91602707 | 25.06485558 | 24.70642281 |
| 22.00113106 | 22.13276291 | 22.53300667 |
| 24.99317169 | 25.34791183 | 25.35025978 |
| 23.60940361 | 23.82139015 | 23.40960884 |
| 19.34047508 | 19.55648613 | 19.30383873 |
| 18.06148148 | 18.49242401 | 17.98779106 |
| 18.27441978 | 18.29142189 | 18.11764908 |
| 25.15032768 | 25.12354279 | 24.91991043 |
| 20.87436867 | 20.82572556 | 21.20713806 |
| 22.54030609 | 22.67210388 | 22.52088356 |
| 25.73919296 | 25.27694702 | 25.48133278 |
| 23.92835236 | 24.07051849 | 23.62998962 |
| 22.83080292 | 23.14254951 | 22.83551788 |
| 18.54144096 | 18.99063301 | 18.45917892 |
| 23.94669724 | 24.01320457 | 23.49637032 |
| 22.46953773 | 21.90253448 | 22.27733803 |
| 18.39054871 | 18.70830917 | 18.66339874 |
| 20.94124603 | 21.18408775 | 20.64829063 |
| 22.61423683 | 22.62372589 | 22.87985039 |
| 22.11850548 | 21.67277336 | 21.87012672 |
| 22.25745392 | 22.22373962 | 21.91165161 |
| 18.72073555 | 18.9129467 | 19.13460541 |
| 22.7368679 | 23.2537117 | 22.83262062 |
| 23.11851311 | 23.04556274 | 23.04344559 |
| 24.62233734 | 24.95773315 | 24.9045639 |
| 24.49727631 | 24.72733688 | 24.04076958 |
| 20.65023041 | 20.49887466 | 20.59244156 |
| 23.07343292 | 22.50257683 | 22.79722977 |
| 21.08289146 | 21.43039703 | 20.83650017 |
| 20.46061897 | 19.9913826 | 20.41999626 |
| 19.47910881 | 19.77861786 | 19.40859032 |
| 18.49293518 | 18.64511299 | 18.87899399 |
| 20.21861458 | 20.08593369 | 20.29381371 |
| 23.20300102 | 23.86381531 | 23.59429169 |
| 21.10069847 | 21.37574768 | 21.57944679 |
| 24.65140915 | 25.13820267 | 24.74904823 |
| 21.98976707 | 21.87942123 | 21.9331646 |
| 18.70549583 | 18.88887787 | 18.87291718 |

| | | |
|---|---|---|
| 24.01968002 | 24.42421722 | 24.10416222 |
| 24.50450897 | 24.89515877 | 24.81747246 |
| 26.35853195 | 26.79694366 | 26.8784008 |
| 22.68009186 | 22.63941574 | 22.39413261 |
| 23.58803558 | 23.21417999 | 23.40033913 |
| 21.14824295 | 21.50738525 | 21.41944122 |
| 18.84545517 | 18.77009583 | 18.84995651 |
| 22.25790787 | 22.6368103 | 22.62371826 |
| 22.18451118 | 21.98125648 | 22.3287468 |
| 23.05204582 | 23.44662476 | 23.28105354 |
| 23.20121384 | 23.49728012 | 23.26571274 |
| 21.06976318 | 21.14179039 | 21.14085388 |
| 20.89028358 | 21.20504951 | 20.91701126 |
| 21.45931625 | 21.74346161 | 21.58182716 |
| 18.27513504 | 18.59357071 | 18.58734894 |
| 23.88770676 | 24.30934143 | 24.2835865 |
| 19.59856606 | 19.34781075 | 19.33833504 |
| 20.29382896 | 20.25534439 | 20.21665192 |
| 21.12504387 | 20.60322762 | 20.96557045 |
| 21.92401886 | 21.74670029 | 21.95990753 |
| 23.4011364 | 23.12197685 | 23.41537476 |
| 23.23093605 | 23.67140579 | 23.57672882 |
| 22.46007347 | 22.65895653 | 23.04115295 |
| 18.17578316 | 18.44643402 | 18.28109932 |
| 23.50085068 | 23.09540367 | 23.31814384 |
| 23.690382 | 23.88150406 | 23.96067429 |
| 22.1865139 | 22.61804771 | 22.31550217 |
| 26.9596405 | 26.95277786 | 27.19338226 |
| 19.75054932 | 19.55227661 | 19.49421692 |
| 23.61515427 | 23.15950012 | 23.58333397 |
| 21.70803642 | 21.79623604 | 21.9577961 |
| 23.03059959 | 23.56118584 | 23.19895935 |
| 22.2432003 | 21.86567497 | 22.1751194 |
| 23.26070786 | 23.07727814 | 23.23287964 |
| 24.2130928 | 23.97505569 | 24.07062721 |
| 21.95409966 | 21.63472557 | 21.36002159 |
| 19.89339638 | 19.76953888 | 19.42597771 |
| 24.38516808 | 24.43869591 | 24.36571884 |
| 22.31956863 | 22.34499931 | 22.12842178 |
| 22.89357948 | 22.56497955 | 22.51265144 |
| 24.81435776 | 25.15915489 | 25.0492878 |
| 21.44449615 | 21.31801987 | 21.54280663 |
| 18.48229599 | 18.58512497 | 18.46559143 |
| 23.7621727 | 23.24179459 | 23.57385254 |
| 19.38026428 | 19.56336212 | 19.08714104 |
| 20.82187653 | 21.17163849 | 21.01467896 |
| 22.34072495 | 22.61364746 | 22.33238792 |

| | | |
|---|---|---|
| 22.19863701 | 22.49007797 | 22.71737099 |
| 26.54815674 | 26.59017372 | 26.4226017 |
| 24.87503052 | 24.72257996 | 24.51924896 |
| 24.4987812 | 24.84662819 | 24.43781281 |
| 24.00449944 | 24.62115669 | 24.30075073 |
| 18.98355484 | 19.26110268 | 19.01022911 |
| 24.34192657 | 24.18922043 | 24.46852112 |
| 20.38978386 | 20.46299553 | 19.94478226 |
| 24.7844696 | 25.08670425 | 24.84043312 |
| 21.81258965 | 22.26866531 | 21.9824028 |
| 21.91212082 | 22.25889778 | 22.3645401 |
| 20.92993164 | 20.85089493 | 20.83847809 |
| 20.64589119 | 20.9647789 | 20.92577171 |
| 27.17335129 | 26.59682274 | 27.02541161 |
| 21.30163002 | 20.77901459 | 21.11332893 |
| 25.30603218 | 25.6246624 | 25.64602089 |
| 24.79112244 | 24.51880646 | 24.80396271 |
| 21.28150177 | 21.7160511 | 21.55912209 |
| 21.56480026 | 21.59381294 | 21.37468147 |
| 23.67110443 | 23.35140228 | 23.88103294 |
| 22.64342308 | 22.47327805 | 22.45476723 |
| 24.12096024 | 24.31602097 | 24.07899475 |
| 21.61623764 | 21.809515 | 21.37495041 |
| 21.53809929 | 21.46727753 | 21.47527885 |
| 21.50529671 | 21.47324371 | 21.49608803 |
| 17.34950447 | 17.67561722 | 17.72418785 |
| 18.30106926 | 18.56415939 | 18.54751015 |
| 22.20709038 | 22.40698242 | 22.17253494 |
| 18.89235306 | 19.13939095 | 19.20952415 |
| 22.89846992 | 22.47548676 | 22.5087738 |
| 24.06019783 | 24.13585472 | 24.28612328 |
| 20.95627403 | 20.90180969 | 20.79004478 |
| 19.53547096 | 19.61849213 | 19.58794975 |
| 26.09344101 | 26.03406525 | 26.22760963 |
| 19.6044426 | 19.51004982 | 19.86757469 |
| 22.79310226 | 23.29816055 | 23.00886345 |
| 25.29797554 | 25.78918839 | 25.60772896 |
| 20.85582542 | 21.26457024 | 21.29167747 |
| 24.97608566 | 25.02230263 | 24.91818237 |
| 22.60918427 | 22.20588684 | 22.40763474 |
| 19.27395058 | 19.52685547 | 19.36112785 |
| 24.29083252 | 23.80457497 | 23.99264336 |
| 23.26403809 | 22.82595634 | 22.89108086 |
| 20.86244965 | 20.74966431 | 21.03463173 |
| 22.6367836 | 22.58721161 | 22.91464615 |
| 25.11898041 | 25.3815155 | 24.95065498 |
| 22.64119339 | 23.20786095 | 22.92326164 |

| | | |
|---|---|---|
| 21.37721634 | 21.37890625 | 21.34730911 |
| 19.1326828 | 18.81621742 | 19.0848465 |
| 23.034132 | 23.37504196 | 23.11123276 |
| 22.84631348 | 23.05273056 | 22.96449852 |
| 20.38089561 | 19.92788315 | 19.97509193 |
| 20.93002701 | 20.85720634 | 20.83310509 |
| 23.67554092 | 23.72363472 | 23.64142609 |
| 28.30678558 | 28.45390892 | 28.08986855 |
| 25.56050682 | 26.04374123 | 25.89746666 |
| 19.0530262 | 18.88216591 | 19.29606247 |
| 19.42462158 | 19.52994919 | 19.29887962 |
| 22.55195999 | 22.69182587 | 22.8704071 |
| 22.35775566 | 22.29424286 | 22.28291321 |
| 20.94047737 | 20.95081711 | 20.9447937 |
| 26.48612213 | 26.98715591 | 26.43789101 |
| 18.86781693 | 19.14547348 | 19.33870506 |
| 19.38394356 | 19.38705826 | 19.75372696 |
| 20.39632797 | 20.22060394 | 20.39761353 |
| 20.07839012 | 20.3731575 | 20.18517876 |
| 29.43590546 | 29.39849472 | 29.30697632 |
| 22.31516266 | 22.29579163 | 22.45953751 |
| 18.51944351 | 18.88315582 | 18.53876114 |
| 25.64540863 | 25.67216873 | 25.62237167 |
| 24.33745766 | 24.48804092 | 24.1765976 |
| 21.11807251 | 20.90700912 | 20.87349129 |
| 21.51207352 | 21.38002205 | 21.79433823 |
| 21.49728584 | 21.18895149 | 21.52119446 |
| 23.72103882 | 23.74802589 | 24.05219841 |
| 19.88449478 | 19.70254135 | 19.66671371 |
| 26.80784988 | 26.98689842 | 27.08391571 |
| 21.95628166 | 22.41163063 | 22.26966286 |
| 17.69642067 | 17.41724777 | 17.66139984 |
| 20.69895172 | 20.64657021 | 20.26552773 |
| 25.77054977 | 26.10639572 | 25.87124443 |
| 20.55204391 | 20.47480774 | 20.42540932 |
| 20.01561737 | 20.2673893 | 19.90574265 |
| 22.20852852 | 22.69448853 | 22.25134468 |
| 21.52726936 | 21.83738327 | 21.95777893 |
| 22.68439102 | 23.0333519 | 22.71935654 |
| 18.64005852 | 18.82452774 | 18.39557457 |
| 23.16540527 | 22.9633503 | 23.11859894 |
| 19.34877396 | 19.38622284 | 19.5757618 |
| 22.18222809 | 22.57116127 | 22.65748596 |
| 24.06494522 | 24.50134277 | 24.4133873 |
| 23.87290001 | 23.98042488 | 23.73739052 |
| 24.58095932 | 24.65297699 | 24.96331596 |
| 23.78882027 | 23.30284119 | 23.55749321 |

| | | |
|---|---|---|
| 25.32954788 | 25.93740273 | 25.70379448 |
| 21.25912666 | 21.42786598 | 21.3617878 |
| 21.20258331 | 21.09662437 | 21.14433479 |
| 25.51935768 | 25.84403419 | 25.71239281 |
| 24.46735191 | 24.98255539 | 24.79829407 |
| 20.76924706 | 21.23501205 | 20.93273163 |
| 21.47826195 | 21.78347397 | 21.44941902 |
| 24.06861115 | 23.89641571 | 24.4530201 |
| 24.72925758 | 24.41400909 | 24.35753822 |
| 26.00616074 | 26.41936684 | 26.13355637 |
| 19.87339783 | 19.51079178 | 19.4797821 |
| 24.18457222 | 24.31416893 | 24.17335701 |
| 25.40111351 | 25.18015671 | 25.00433922 |
| 22.8538723 | 23.22331047 | 22.97291565 |
| 22.92392731 | 22.93657875 | 22.84892654 |
| 19.04846573 | 19.37466049 | 18.96192741 |
| 20.67087555 | 20.37870216 | 20.63001251 |
| 21.65680695 | 21.44501305 | 21.48412323 |
| 23.12866592 | 23.45551682 | 22.92259979 |
| 29.0576973 | 29.07012367 | 29.03057861 |
| 24.0334034 | 23.94118118 | 23.95052719 |
| 23.93244362 | 23.84057999 | 23.51065254 |
| 22.89126968 | 22.63991928 | 22.53357124 |
| 23.57233047 | 23.69064903 | 24.0361557 |
| 23.59937096 | 24.04516029 | 23.84694481 |
| 19.02535057 | 19.36188507 | 19.26472855 |
| 22.31024742 | 22.5729847 | 22.55839729 |
| 26.49387741 | 26.60808372 | 26.23676872 |
| 23.75028038 | 23.76360512 | 23.62733269 |
| 23.49492836 | 23.40342522 | 23.27206802 |
| 22.84932899 | 22.9992485 | 22.8601017 |
| 24.44831085 | 24.99869728 | 24.6931839 |
| 20.59017181 | 20.42888832 | 20.490448 |
| 23.60731506 | 23.18544769 | 23.61671829 |
| 20.96593094 | 20.79319763 | 20.94487953 |
| 19.79564095 | 19.69800377 | 20.00150871 |
| 22.90929031 | 22.61611366 | 22.65906334 |
| 24.3291378 | 24.77191544 | 24.41571045 |
| 21.78129196 | 21.49225426 | 21.50965118 |
| 23.67284012 | 23.97627258 | 23.954813 |
| 23.08662796 | 23.39180756 | 23.35678101 |
| 27.8268013 | 28.0870018 | 28.42263985 |
| 22.97832489 | 22.75675774 | 23.05238152 |
| 23.9252243 | 24.22916794 | 24.22408676 |
| 21.00749969 | 20.67033768 | 20.80798149 |
| 23.1627636 | 23.302351 | 23.49739838 |
| 22.21305847 | 22.48327637 | 22.08805275 |

| | | |
|---|---|---|
| 29.70678902 | 30.14489365 | 29.76222992 |
| 21.85084534 | 21.68795204 | 21.63849068 |
| 23.41908455 | 23.26712799 | 23.29023743 |
| 18.59186172 | 18.84722328 | 18.79059219 |
| 21.27783966 | 21.28339577 | 21.52541733 |
| 24.01614571 | 23.84511185 | 23.95397568 |
| 23.18212318 | 23.49760437 | 23.58382225 |
| 23.96952629 | 24.21407509 | 23.9729805 |
| 20.3574276 | 20.13693428 | 20.31995392 |
| 23.80604744 | 24.25678635 | 23.93579292 |
| 23.54813194 | 23.78478241 | 23.78207779 |
| 23.79063225 | 23.56130791 | 23.62124062 |
| 19.79372406 | 19.70195007 | 20.03175354 |
| 23.64920044 | 24.16930389 | 23.73627472 |
| 19.59346771 | 19.88680077 | 19.74400711 |
| 25.51558495 | 25.26002502 | 25.32010841 |
| 21.30391312 | 21.70148849 | 21.26662064 |
| 23.09579659 | 23.38619041 | 23.29555893 |
| 24.50223732 | 24.98620033 | 24.90953827 |
| 20.84332085 | 21.07915115 | 20.63707733 |
| 24.5296402 | 24.98776245 | 24.76187706 |
| 22.83008003 | 23.30412483 | 23.25967026 |
| 27.40584564 | 26.9126339 | 27.0205307 |
| 23.30109215 | 23.55852318 | 23.58164024 |
| 22.94733429 | 22.77324867 | 23.14759064 |
| 22.3187027 | 22.10817528 | 22.32803917 |
| 24.73235893 | 25.03756523 | 24.84742355 |
| 25.50007248 | 25.60399437 | 25.59625816 |
| 23.74743843 | 23.68525505 | 24.2007618 |
| 25.49091339 | 25.29558945 | 25.32057762 |
| 20.17556572 | 20.37001991 | 20.01436424 |
| 21.92767715 | 22.2443428 | 22.36020088 |
| 23.00487518 | 23.08175087 | 22.77670479 |
| 25.48475647 | 25.33504295 | 25.58163261 |
| 21.46306419 | 21.3305378 | 21.71809006 |
| 24.30127907 | 24.34365273 | 24.34830284 |
| 23.75470352 | 23.85738945 | 23.75856209 |
| 20.90599442 | 21.13659477 | 20.94514847 |
| 22.57927895 | 22.75969696 | 22.45096397 |
| 28.07585907 | 27.93109131 | 28.20156097 |
| 22.09865761 | 21.81480408 | 22.08411026 |
| 21.79133034 | 21.9066143 | 22.12743759 |
| 21.96247101 | 22.08741379 | 21.99013138 |
| 19.82392311 | 19.98653603 | 19.96156883 |
| 25.86929893 | 25.64813232 | 25.80156517 |
| 23.30989075 | 23.28898621 | 23.29813194 |
| 24.39871788 | 24.28970909 | 24.2776947 |

| | | |
|---|---|---|
| 26.11512184 | 26.17527008 | 26.30018234 |
| 28.72652054 | 28.89901733 | 29.25956345 |
| 23.26914215 | 23.35146523 | 22.86201477 |
| 21.09144783 | 20.97972679 | 21.13755035 |
| 28.57752037 | 29.01029968 | 28.62400627 |
| 21.1308918 | 20.96442223 | 20.84150124 |
| 25.16554642 | 25.38647079 | 25.08890724 |
| 24.12114143 | 24.03728485 | 24.33107948 |
| 24.57071495 | 24.8454628 | 24.39594269 |
| 29.3823452 | 29.26946449 | 29.19558525 |
| 23.46670341 | 23.82435989 | 23.56951141 |
| 21.93856621 | 21.86379242 | 21.76676369 |
| 21.52902412 | 21.80199814 | 21.76542282 |
| 26.16054153 | 26.06767082 | 26.01083374 |
| 23.97503853 | 23.75163269 | 23.77211189 |
| 20.58246803 | 20.84919167 | 20.9640274 |
| 23.77390862 | 23.74370003 | 23.90841484 |
| 20.66425514 | 20.30395126 | 20.26774025 |
| 22.94528008 | 23.15182304 | 22.98534012 |
| 22.48180008 | 22.6328373 | 22.39366341 |
| 24.69214821 | 24.94085693 | 24.56672287 |
| 25.2142849 | 25.0156765 | 25.05943871 |
| 28.66586304 | 28.82177925 | 28.61738014 |
| 24.41604233 | 24.87857246 | 24.41752243 |
| 22.90423965 | 22.8547554 | 23.17213058 |
| 20.79549217 | 20.991045 | 20.86352158 |
| 18.52126694 | 18.63280869 | 18.82782364 |
| 22.11751938 | 22.24862289 | 22.11283112 |
| 23.90645218 | 24.39758301 | 24.04071808 |
| 21.3586216 | 21.73582268 | 21.64789391 |
| 24.77080536 | 24.82720566 | 24.69799805 |
| 24.56525993 | 24.73583984 | 24.84067726 |
| 24.62432861 | 24.40055656 | 24.71416283 |
| 24.16067123 | 24.28851128 | 24.4237957 |
| 24.30513573 | 24.71168518 | 24.58089447 |
| 25.254673 | 25.44483185 | 25.39970779 |
| 25.51341248 | 25.19950867 | 25.66020775 |
| 25.58579826 | 25.60250282 | 25.47998238 |
| 20.83475494 | 21.00597954 | 21.24422455 |
| 24.33943367 | 24.00632858 | 24.06759453 |
| 23.49212265 | 23.5508213 | 23.25986862 |
| 23.46225357 | 23.43919754 | 23.38059235 |
| 26.21184349 | 26.08199501 | 26.23711586 |
| 21.14986038 | 20.91180801 | 21.34423637 |
| 26.65283775 | 26.19115448 | 26.55937767 |
| 22.95866394 | 23.11247063 | 23.02637291 |
| 24.33276558 | 24.76046944 | 24.40812492 |

| | | |
|---|---|---|
| 20.306633 | 20.11069489 | 20.43005371 |
| 20.53430748 | 20.43054199 | 20.25667191 |
| 24.79451561 | 24.45041275 | 24.87458611 |
| 22.17173195 | 22.27511978 | 21.96258354 |
| 21.58839989 | 21.78336525 | 21.40568924 |
| 28.41954613 | 28.52068138 | 28.59325409 |
| 26.17252159 | 25.761549 | 26.03325272 |
| 25.4902916 | 25.49106789 | 25.42864418 |
| 23.1274147 | 23.064785 | 23.0970726 |
| 23.73600197 | 23.72067833 | 23.92022514 |
| 21.64000893 | 21.79010963 | 21.79877663 |
| 20.87693214 | 21.1282444 | 21.01928139 |
| 25.24626923 | 25.70150566 | 25.48532295 |
| 23.00446892 | 23.12145805 | 22.70616722 |
| 21.44809341 | 21.2920475 | 21.56885147 |
| 27.29341316 | 27.64202118 | 27.32829094 |
| 23.04946518 | 22.80565643 | 23.02087784 |
| 24.19087791 | 24.66382217 | 24.29665756 |
| 23.68629265 | 23.64626694 | 23.5213356 |
| 22.95667267 | 23.13640594 | 23.15437698 |
| 20.04789162 | 20.25224304 | 20.25185204 |
| 22.15280533 | 22.24990273 | 22.04042625 |
| 23.6710453 | 23.88155556 | 23.64024162 |
| 23.17384148 | 23.25336647 | 23.28092003 |
| 23.52118874 | 23.52353477 | 23.79937744 |
| 22.46968842 | 22.69892502 | 22.71897507 |
| 24.44121933 | 24.40924644 | 24.48323822 |
| 23.09336662 | 22.98316956 | 23.18084335 |
| 18.98675537 | 18.76005173 | 18.91896248 |
| 23.75112343 | 23.90967369 | 23.42677498 |
| 20.30818939 | 20.1993103 | 20.42853165 |
| 21.4970417 | 21.4389267 | 21.74554825 |
| 22.89166832 | 23.02171516 | 23.19686127 |
| 25.76256371 | 26.07193184 | 26.10676193 |
| 22.96570396 | 23.12735558 | 23.30430984 |
| 24.64610481 | 24.9273262 | 24.64299583 |
| 24.00520706 | 23.81400681 | 23.98433876 |
| 25.83889198 | 25.85349846 | 25.86011124 |
| 22.30292702 | 22.49006271 | 22.16120338 |
| 25.56605911 | 26.03161812 | 25.96844101 |
| 19.48479843 | 19.31583595 | 19.3006134 |
| 21.47819519 | 21.68364334 | 21.53333092 |
| 24.02086067 | 24.15155792 | 24.18938065 |
| 25.78623962 | 26.05388069 | 26.00903893 |
| 25.84841347 | 26.36147499 | 26.05522346 |
| 21.07959938 | 21.17976952 | 21.48806381 |
| 27.88775253 | 28.28614426 | 28.23545837 |

| | | |
|---|---|---|
| 23.74178314 | 23.98880386 | 23.67638016 |
| 23.0551033 | 23.25508308 | 23.12952423 |
| 20.9724369 | 21.13519478 | 21.24117851 |
| 26.98605728 | 26.78892136 | 26.96120262 |
| 28.43652344 | 28.32720566 | 28.1388092 |
| 24.51044273 | 24.40015411 | 24.41932869 |
| 20.96031761 | 21.06654167 | 20.78087616 |
| 21.82837486 | 21.8174057 | 21.96848488 |
| 22.15228653 | 22.30852509 | 22.23828506 |
| 23.40744591 | 23.3806572 | 23.46449471 |
| 24.31707954 | 24.41545296 | 24.36037254 |
| 22.83247757 | 23.07773018 | 23.00823021 |
| 22.13098526 | 22.24608231 | 22.22397804 |
| 20.14064217 | 20.16010284 | 19.84591293 |
| 25.93120193 | 25.88148689 | 26.01326942 |
| 21.40620232 | 21.3252449 | 21.31108856 |
| 30.2046566 | 30.19913673 | 30.38235283 |
| 21.71856689 | 21.55030823 | 21.51714897 |
| 23.69879341 | 23.99725342 | 23.8283596 |
| 24.98167038 | 25.18645287 | 25.32574463 |
| 23.12757683 | 23.29980087 | 22.8860817 |
| 28.83059502 | 28.28509331 | 28.62137032 |
| 23.7953968 | 23.6166153 | 23.52520943 |
| 24.64684677 | 24.70837975 | 24.70216179 |
| 22.60570526 | 22.24822044 | 22.24185753 |
| 22.25741768 | 22.34289551 | 22.41337967 |
| 26.10160446 | 25.93812561 | 25.9198246 |
| 23.37330246 | 23.7883358 | 23.46753693 |
| 24.81070137 | 24.79294777 | 24.87638664 |
| 24.88942909 | 24.65624809 | 24.84184647 |
| 23.95908356 | 23.79131699 | 23.95168686 |
| 22.94688606 | 22.85499191 | 22.76739311 |
| 25.42417526 | 25.63923836 | 25.55044746 |
| 23.99378204 | 24.37775993 | 24.22526169 |
| 21.77870178 | 21.67033005 | 21.43706703 |
| 23.6409893 | 23.62764931 | 23.41115379 |
| 23.45973969 | 23.39016724 | 23.39266396 |
| 23.14407349 | 22.90189552 | 23.06626511 |
| 27.83709717 | 27.62353325 | 27.6944046 |
| 25.50065041 | 25.59010124 | 25.48986626 |
| 23.73983383 | 23.53969765 | 23.79597855 |
| 20.27315903 | 20.4928112 | 20.39566231 |
| 25.25495148 | 25.39644051 | 25.34276009 |
| 23.40474129 | 23.2231636 | 23.39848328 |
| 19.55694008 | 19.80233574 | 19.82841301 |
| 22.05739975 | 21.97057533 | 22.1059227 |
| 19.99799728 | 19.96461296 | 20.03557396 |

| | | |
|---|---|---|
| 23.73273468 | 23.742239 | 23.60389519 |
| 22.31721687 | 22.12081718 | 22.07356453 |
| 26.00628471 | 26.05892372 | 26.06353569 |
| 24.14790344 | 23.95127678 | 24.09123993 |
| 20.94130707 | 21.26484299 | 21.06916809 |
| 23.53030014 | 23.41021919 | 23.72355652 |
| 23.76037979 | 23.56468201 | 23.70707512 |
| 25.96241951 | 26.32805443 | 26.12093925 |
| 28.38490677 | 27.97588348 | 28.08580971 |
| 18.4161644 | 18.34334183 | 18.25351334 |
| 24.68203545 | 24.67190361 | 24.83790398 |
| 23.19050789 | 23.16359329 | 23.18324852 |
| 24.1262989 | 24.20046425 | 24.02617836 |
| 21.7598877 | 21.6858902 | 21.67144203 |
| 23.86746979 | 24.10547447 | 24.04061127 |
| 22.13494492 | 22.52286148 | 22.4367981 |
| 25.93519783 | 26.20120239 | 26.17378426 |
| 22.53351402 | 22.41563988 | 22.52055359 |
| 25.12363052 | 25.11585045 | 25.40541458 |
| 23.69705582 | 24.07814407 | 23.97725105 |
| 27.07248116 | 27.35328484 | 27.14438438 |
| 23.96658707 | 24.37948418 | 24.25172234 |
| 24.80528259 | 24.96099091 | 24.85382271 |
| 23.98414612 | 24.16461372 | 24.20056343 |
| 21.85676956 | 22.2056942 | 22.11306 |
| 25.6154232 | 25.86543274 | 25.47814941 |
| 24.95927048 | 25.27279282 | 25.19846916 |
| 22.13185883 | 21.87316132 | 22.20090485 |
| 22.40920639 | 22.29904556 | 22.38656235 |
| 23.72236252 | 23.72938156 | 24.03900146 |
| 23.52904129 | 23.28853989 | 23.68603325 |
| 18.97171783 | 18.96860123 | 19.22781944 |
| 24.27812195 | 24.44678497 | 24.38210106 |
| 25.38936234 | 25.56497383 | 25.4375515 |
| 20.66071129 | 20.92248344 | 20.77169418 |
| 20.95761871 | 21.11273003 | 21.25509262 |
| 27.03092194 | 26.98757553 | 26.92024612 |
| 25.22464752 | 25.28619766 | 25.31949615 |
| 21.54262161 | 21.35387993 | 21.30129433 |
| 25.47258377 | 25.52847099 | 25.41129684 |
| 25.46926498 | 25.74475479 | 25.74598694 |
| 20.97417641 | 20.99303246 | 21.02771187 |
| 27.2047081 | 27.46960831 | 27.47638893 |
| 23.2836647 | 23.61161804 | 23.2966671 |
| 24.76792145 | 24.96797943 | 24.66964531 |
| 20.68200302 | 20.51590729 | 20.8465786 |
| 28.65746498 | 28.53819084 | 28.66897583 |

| | | |
|---|---|---|
| 23.09599686 | 23.31157684 | 23.31760216 |
| 22.44055939 | 22.22524834 | 22.15660095 |
| 26.75658417 | 27.21439934 | 26.98970413 |
| 24.80771637 | 24.87476921 | 24.73260689 |
| 24.61365891 | 24.28802109 | 24.53336906 |
| 23.25963211 | 23.06280327 | 23.07049179 |
| 19.5926609 | 19.46523476 | 19.29044151 |
| 26.52622414 | 26.52610016 | 26.49341583 |
| 21.58345795 | 21.40071297 | 21.40129852 |
| 25.98268127 | 26.21281815 | 25.99295425 |
| 19.25143814 | 19.36083031 | 19.10252571 |
| 27.55537605 | 27.57837868 | 27.28834724 |
| 21.152668 | 21.38962364 | 21.15822411 |
| 25.76701164 | 25.90093803 | 25.56072235 |
| 23.93029976 | 24.10632896 | 23.98521042 |
| 22.66588402 | 22.59594917 | 22.49331856 |
| 24.23856735 | 24.16423988 | 24.27665329 |
| 23.1233902 | 22.99940109 | 23.12108803 |
| 25.01151848 | 25.27043915 | 25.14494133 |
| 23.02082062 | 22.87212372 | 23.12590218 |
| 26.62376595 | 26.78109169 | 26.43626404 |
| 21.67344856 | 21.91671944 | 21.60625839 |
| 24.77728844 | 24.55818939 | 24.40337944 |
| 20.08208275 | 20.32336807 | 20.34255028 |
| 21.21740723 | 21.24640083 | 21.26743889 |
| 23.61941719 | 23.93100166 | 23.75833321 |
| 25.0639782 | 25.40942192 | 25.29041862 |
| 22.77798843 | 23.07991791 | 22.78679657 |
| 24.37622261 | 24.33705902 | 24.46227646 |
| 21.06604576 | 20.94788742 | 20.92840767 |
| 25.71909904 | 25.77521515 | 25.64017868 |
| 27.05762482 | 27.21045876 | 27.1162014 |
| 25.86810303 | 26.17750549 | 25.94943237 |
| 22.99009895 | 23.15590668 | 22.88154221 |
| 21.67599106 | 21.52106476 | 21.73078537 |
| 21.44646835 | 21.44977188 | 21.49972534 |
| 19.79688835 | 19.90306282 | 19.66149712 |
| 26.55389023 | 26.20126534 | 26.61306953 |
| 24.24599838 | 24.60396576 | 24.50289536 |
| 25.45824432 | 25.73624611 | 25.60155678 |
| 24.88247871 | 24.94119835 | 24.93554878 |
| 19.71928978 | 19.80919266 | 19.64728355 |
| 28.64529419 | 28.83795166 | 28.82679939 |
| 24.80225945 | 24.99907875 | 24.77303314 |
| 28.36878967 | 28.57950401 | 28.6177845 |
| 22.78283501 | 22.77514458 | 22.64913559 |
| 24.23978806 | 24.54530144 | 24.28018379 |

| | | |
|---|---|---|
| 22.08022118 | 22.33966446 | 22.28356552 |
| 24.51673508 | 24.81149483 | 24.56114578 |
| 24.64125633 | 24.8153801 | 24.7205677 |
| 23.64182854 | 23.46841812 | 23.61618233 |
| 23.94249725 | 24.12779045 | 24.00512123 |
| 25.09529305 | 25.29160118 | 25.41421318 |
| 26.24476051 | 26.3775425 | 26.11802292 |
| 28.23272705 | 28.40135002 | 28.14451981 |
| 24.4154129 | 24.68221092 | 24.50190353 |
| 23.75276566 | 23.4394722 | 23.66859245 |
| 25.46132469 | 25.1124649 | 25.29754829 |
| 22.63490677 | 22.90190506 | 22.7682457 |
| 26.36762619 | 25.98804283 | 26.18937302 |
| 23.56487465 | 23.38729095 | 23.31293106 |
| 22.67859268 | 22.72511101 | 22.66754532 |
| 22.39339638 | 22.20750237 | 22.21907616 |
| 26.28580093 | 26.66663742 | 26.35211182 |
| 21.10703087 | 21.11916161 | 21.27454376 |
| 25.93330383 | 26.28349304 | 26.26964188 |
| 24.35597038 | 24.19610214 | 24.39984894 |
| 23.3389225 | 23.15207291 | 23.41826439 |
| 23.20880699 | 23.10445595 | 23.18026733 |
| 28.16529846 | 28.57182693 | 28.40788078 |
| 27.51847649 | 27.15205383 | 27.47463036 |
| 25.10583878 | 24.98711777 | 25.31408691 |
| 24.4326973 | 24.58956146 | 24.39229584 |
| 22.79615784 | 23.0361042 | 22.84591866 |
| 21.75379372 | 21.63750076 | 21.55512619 |
| 25.99339867 | 25.75036812 | 25.88383102 |
| 24.01997185 | 24.19729233 | 24.15714836 |
| 22.68045807 | 22.90879631 | 22.90238953 |
| 22.58768082 | 22.80342484 | 22.87891197 |
| 23.05601692 | 23.06387901 | 23.17972755 |
| 22.96836281 | 22.85625267 | 22.88493156 |
| 24.59806442 | 24.89476967 | 24.74585152 |
| 20.07711601 | 20.18216133 | 20.35965729 |
| 28.70379448 | 28.89357185 | 28.90520859 |
| 23.9616642 | 23.93045998 | 24.16415787 |
| 25.37017822 | 25.14614677 | 25.01913834 |
| 23.43891716 | 23.25998497 | 23.25810814 |
| 24.78163719 | 24.98031235 | 24.87221718 |
| 24.3966713 | 24.75730324 | 24.49010849 |
| 23.58601189 | 23.86686325 | 23.61211395 |
| 25.29855728 | 25.38545227 | 25.47686005 |
| 22.63223076 | 22.53389549 | 22.52577972 |
| 22.50811577 | 22.28257179 | 22.23912621 |
| 25.55736542 | 25.43556213 | 25.4003315 |

| | | |
|---|---|---|
| 30.40433693 | 30.10739517 | 30.5755043 |
| 18.21835327 | 18.39811707 | 18.23182678 |
| 27.44241714 | 27.4208889 | 27.71248245 |
| 25.81181335 | 26.03621101 | 25.70503426 |
| 27.52356529 | 27.52616882 | 27.63705254 |
| 21.68161964 | 22.01049042 | 21.83938789 |
| 23.26530647 | 23.06178856 | 23.21364212 |
| 19.98827171 | 20.10948753 | 20.0966568 |
| 23.92841339 | 23.74723625 | 23.86796379 |
| 23.60647774 | 23.71779633 | 23.64476013 |
| 24.44427299 | 24.30721664 | 24.3866024 |
| 24.98163033 | 24.95074272 | 25.14048958 |
| 26.14199829 | 26.3305397 | 26.55303383 |
| 20.5120697 | 20.71405029 | 20.59664726 |
| 20.9671936 | 20.87096596 | 20.8889389 |
| 24.69812584 | 24.75735855 | 24.66932487 |
| 26.18269157 | 26.37043953 | 26.54045677 |
| 20.81261826 | 21.04059219 | 21.05254745 |
| 29.01096344 | 29.1182785 | 29.0849247 |
| 20.77649307 | 21.0092926 | 20.89430428 |
| 25.09716988 | 24.89009476 | 24.7992382 |
| 26.15903091 | 26.10805511 | 25.95175934 |
| 23.89663315 | 24.18505859 | 23.95982552 |
| 24.66428185 | 24.67628479 | 24.65151978 |
| 23.50881767 | 23.35657501 | 23.46235847 |
| 23.4707737 | 23.61466789 | 23.74879456 |
| 26.50816154 | 26.88802528 | 26.74039841 |
| 25.02594185 | 25.1402626 | 25.01535797 |
| 27.84152412 | 27.59650993 | 27.53617477 |
| 28.42711639 | 28.77630043 | 28.73617363 |
| 23.69095993 | 23.94011497 | 23.67248535 |
| 25.28728676 | 25.1533432 | 25.1252327 |
| 28.2714901 | 28.5801239 | 28.23389244 |
| 25.45221138 | 25.56727219 | 25.54901314 |
| 24.74593353 | 24.83701706 | 24.92265129 |
| 24.57421494 | 24.80522728 | 24.63992882 |
| 22.03178024 | 22.16592598 | 22.09103203 |
| 24.32336807 | 24.43680763 | 24.23857307 |
| 24.88718796 | 24.80737877 | 24.96364021 |
| 24.45164108 | 24.7855587 | 24.58046722 |
| 24.19749069 | 24.28435135 | 24.10637474 |
| 23.69851494 | 23.80174637 | 23.78403282 |
| 24.23323441 | 24.41770744 | 24.31014442 |
| 22.02945518 | 21.96746063 | 21.79860687 |
| 23.15824127 | 23.38101387 | 23.26445007 |
| 26.04870224 | 26.38876534 | 26.17561722 |
| 24.10276985 | 23.86933517 | 24.04290581 |

| | | |
|---|---|---|
| 21.65488434 | 21.62838554 | 21.74046326 |
| 26.65213776 | 26.52237511 | 26.90073776 |
| 18.96031761 | 18.91260338 | 18.77293015 |
| 23.39646912 | 23.44147873 | 23.35591888 |
| 26.54426575 | 26.50758743 | 26.46372604 |
| 21.74137115 | 21.93509865 | 21.83945465 |
| 25.75426292 | 25.78182602 | 25.86387634 |
| 27.11271477 | 27.03744888 | 27.04580498 |
| 24.67851257 | 24.95054626 | 24.8217411 |
| 19.62477303 | 19.61525726 | 19.45150566 |
| 29.13446236 | 29.10660362 | 29.01117897 |
| 26.48280334 | 26.32430267 | 26.55415535 |
| 26.58224678 | 26.69695663 | 26.87590218 |
| 24.59442711 | 24.65813255 | 24.56699371 |
| 25.70947456 | 25.60870361 | 25.58778954 |
| 26.55481339 | 26.87173462 | 26.74404335 |
| 21.95928192 | 21.92961693 | 22.20142174 |
| 25.81417465 | 25.83157349 | 25.75597763 |
| 25.55638695 | 25.52235794 | 25.68753433 |
| 24.72769547 | 24.7399044 | 24.58422661 |
| 25.88793755 | 25.74658585 | 26.07095337 |
| 23.76581955 | 23.85249901 | 23.86075401 |
| 24.16551208 | 24.4798317 | 24.21464539 |
| 22.997715 | 22.94857597 | 22.80609322 |
| 24.87866402 | 25.18457031 | 24.98430824 |
| 25.4848671 | 25.57711029 | 25.51162338 |
| 23.86374092 | 23.9197464 | 23.87280655 |
| 28.37560463 | 28.39551353 | 28.29490471 |
| 22.73173523 | 22.83964539 | 22.66433716 |
| 25.70070457 | 25.65562248 | 25.57878494 |
| 22.45378494 | 22.41364861 | 22.44696426 |
| 25.38132286 | 25.20392609 | 25.31658745 |
| 27.78336906 | 27.61831284 | 27.82582283 |
| 23.29385567 | 23.46816635 | 23.39196777 |
| 24.67341042 | 24.42572784 | 24.57111549 |
| 24.65969276 | 24.63090324 | 24.82370186 |
| 27.37753868 | 27.4731884 | 27.51667213 |
| 22.02776527 | 21.97636032 | 21.81774712 |
| 27.37468147 | 27.49774933 | 27.3941803 |
| 26.10766792 | 26.18839073 | 26.26028061 |
| 27.48808289 | 27.55441666 | 27.5918541 |
| 21.8570137 | 22.02251625 | 21.96057892 |
| 25.3990078 | 25.19680214 | 25.31428719 |
| 20.29736137 | 20.54367638 | 20.39395905 |
| 23.85768127 | 23.76902199 | 23.86403656 |
| 21.74877548 | 21.77273178 | 21.71850395 |
| 22.19705582 | 22.3899231 | 22.19557953 |

| | | |
|---|---|---|
| 24.52935028 | 24.6486969 | 24.82629585 |
| 24.39259338 | 24.39180183 | 24.36948967 |
| 27.55286598 | 27.45651054 | 27.54389 |
| 23.61738968 | 23.75180817 | 23.5248394 |
| 22.00181198 | 22.09683037 | 21.89321327 |
| 25.32588387 | 25.60519028 | 25.53799057 |
| 28.4541111 | 28.57208824 | 28.57803917 |
| 21.17673683 | 21.31145287 | 21.22660255 |
| 25.94447899 | 26.25522041 | 26.03549576 |
| 23.34205055 | 23.21729851 | 23.25428581 |
| 25.51057053 | 25.4864521 | 25.43312645 |
| 19.75375557 | 19.84251976 | 19.86289978 |
| 26.50161934 | 26.65778351 | 26.66828346 |
| 28.43174934 | 28.30016327 | 28.19300079 |
| 24.01904869 | 24.18458557 | 24.07607841 |
| 27.55375862 | 27.6456604 | 27.5947628 |
| 28.48875999 | 28.44267845 | 28.45311737 |
| 25.37605667 | 25.69130325 | 25.63841057 |
| 24.78057861 | 24.96491623 | 24.85578918 |
| 25.76613617 | 26.00382042 | 25.92424011 |
| 23.41241646 | 23.53789711 | 23.38576317 |
| 24.17042732 | 24.3582077 | 24.41594124 |
| 28.87450409 | 28.99200439 | 28.72589111 |
| 26.11543274 | 26.35176849 | 26.15635681 |
| 23.50648689 | 23.48978043 | 23.50860596 |
| 25.87162209 | 25.88568497 | 25.95937157 |
| 24.41875648 | 24.39258385 | 24.13754463 |
| 26.92777634 | 27.03006744 | 26.92137527 |
| 24.87026405 | 25.16946411 | 24.948349 |
| 24.1012001 | 24.1111908 | 24.05809975 |
| 25.38829231 | 25.25741577 | 25.20236206 |
| 26.65561867 | 26.55318451 | 26.73736572 |
| 24.11007881 | 24.13187408 | 24.13020515 |
| 24.04100418 | 24.26278687 | 23.98887062 |
| 29.38304138 | 29.19512558 | 29.19618225 |
| 24.07446289 | 24.15472221 | 24.22828484 |
| 20.99225044 | 21.07108307 | 20.95155144 |
| 25.96617317 | 25.95993614 | 25.93259048 |
| 25.83918571 | 25.81166077 | 25.6076889 |
| 21.89167404 | 21.93692589 | 22.09526253 |
| 25.9520874 | 26.13310051 | 26.26080894 |
| 24.56304169 | 24.79652786 | 24.81262589 |
| 27.52947998 | 27.28447723 | 27.29585075 |
| 22.71525955 | 22.95764923 | 22.78288078 |
| 24.1075325 | 24.05711555 | 23.94591522 |
| 29.80043411 | 29.60463142 | 29.52157593 |
| 24.65340424 | 24.44427299 | 24.59890747 |

| | | |
|---|---|---|
| 24.3146553 | 24.36725044 | 24.48605156 |
| 25.50709724 | 25.57685089 | 25.66096497 |
| 24.67078018 | 24.77095413 | 24.6272316 |
| 24.74575043 | 24.87388802 | 25.01353836 |
| 21.0628376 | 21.22055244 | 21.03648758 |
| 23.8711071 | 23.79911041 | 23.95025826 |
| 23.3920269 | 23.27036095 | 23.25080872 |
| 32.99484634 | 33.02206421 | 32.92900085 |
| 21.48092651 | 21.56409454 | 21.49813843 |
| 28.89570999 | 28.65685081 | 28.85130119 |
| 25.86431313 | 25.96568871 | 25.82826424 |
| 24.25048065 | 24.25915146 | 24.33537483 |
| 23.86188889 | 23.86149788 | 23.84508705 |
| 29.04764175 | 28.72938156 | 28.83805275 |
| 24.43527985 | 24.36751556 | 24.40353203 |
| 26.49205971 | 26.67136765 | 26.46821022 |
| 26.35913658 | 26.32658577 | 26.4164753 |
| 22.7602272 | 22.93657112 | 22.79727554 |
| 27.04635048 | 27.01211166 | 26.84023476 |
| 24.65716934 | 24.75039101 | 24.75528717 |
| 22.62861443 | 22.6852932 | 22.67382813 |
| 27.13378525 | 27.14612961 | 27.03105736 |
| 21.96777725 | 22.15572739 | 22.03528595 |
| 27.43517876 | 27.66487122 | 27.75180244 |
| 25.57272911 | 25.60693741 | 25.78593636 |
| 22.14958191 | 22.34213257 | 22.12082863 |
| 24.12237358 | 24.02153015 | 23.92562103 |
| 25.32859612 | 25.45348167 | 25.40895271 |
| 28.50093269 | 28.5395546 | 28.34126282 |
| 27.7018795 | 27.52095604 | 27.6533699 |
| 29.22408676 | 29.28455162 | 29.27149963 |
| 28.58813858 | 28.50370216 | 28.70779991 |
| 24.22682953 | 24.39171982 | 24.25387383 |
| 27.18773651 | 27.37416077 | 27.28402138 |
| 22.70519066 | 22.64801979 | 22.72150803 |
| 25.86194801 | 25.82905006 | 25.94112396 |
| 23.52739906 | 23.48212433 | 23.35511971 |
| 25.2547245 | 25.33405495 | 25.27499199 |
| 31.9516716 | 31.89536285 | 32.04027176 |
| 22.01594353 | 21.9753437 | 21.99442482 |
| 24.28042984 | 24.48064041 | 24.47607994 |
| 28.96137238 | 29.29431343 | 29.04187012 |
| 23.46269035 | 23.49390221 | 23.36188316 |
| 25.41253281 | 25.48396301 | 25.32691765 |
| 26.86078072 | 27.00820732 | 27.08424187 |
| 25.69304466 | 25.87026978 | 25.85504913 |
| 24.80224609 | 24.70494843 | 24.94924164 |

| | | |
|---|---|---|
| 23.57016182 | 23.60328293 | 23.45212173 |
| 27.46194649 | 27.47847557 | 27.55093193 |
| 27.30432129 | 27.54240036 | 27.41769028 |
| 23.67879295 | 23.67951012 | 23.88635063 |
| 24.150177 | 24.33062172 | 24.30563927 |
| 24.45735359 | 24.69004059 | 24.53495598 |
| 19.74163246 | 19.56380081 | 19.60164452 |
| 27.4293232 | 27.56307411 | 27.58275795 |
| 30.91817093 | 30.7367382 | 30.86490059 |
| 26.72916985 | 26.85336113 | 26.83041573 |
| 25.71000671 | 25.90520477 | 25.91442871 |
| 25.0270462 | 24.90577888 | 24.77556419 |
| 27.80577087 | 27.85334969 | 28.00800514 |
| 19.20752907 | 19.13331985 | 19.16044617 |
| 19.83014107 | 19.76929665 | 19.82877922 |
| 25.94507027 | 26.16467094 | 26.12335968 |
| 26.42697334 | 26.43045807 | 26.45333481 |
| 23.48189926 | 23.55685806 | 23.54650116 |
| 25.82479858 | 26.04586792 | 26.014534 |
| 23.59973335 | 23.84910202 | 23.74299812 |
| 24.34767532 | 24.42810822 | 24.58354759 |
| 24.97557068 | 24.75355721 | 24.87991524 |
| 27.12360573 | 27.32095909 | 27.15708733 |
| 22.38056564 | 22.37252998 | 22.19612503 |
| 24.41837883 | 24.19207382 | 24.2596817 |
| 23.81064034 | 23.7326088 | 23.73780441 |
| 19.93068695 | 20.00105476 | 19.8515358 |
| 24.3441124 | 24.29330444 | 24.24774742 |
| 26.61339951 | 26.49749374 | 26.62728882 |
| 23.33532143 | 23.30569839 | 23.43340302 |
| 26.49918747 | 26.60066223 | 26.70088959 |
| 26.15149689 | 26.10176468 | 25.92607117 |
| 25.03270912 | 25.10422897 | 25.13850212 |
| 23.67899323 | 23.4841938 | 23.67679024 |
| 25.13759995 | 25.10130882 | 25.23033524 |
| 25.2952919 | 25.20292473 | 25.22862244 |
| 24.28195763 | 24.10045052 | 24.20357895 |
| 27.05788994 | 27.23411369 | 27.04616356 |
| 25.33842468 | 25.43621635 | 25.42669296 |
| 24.99697685 | 24.96583939 | 25.15674782 |
| 25.9870472 | 26.1604023 | 25.9966507 |
| 24.4901638 | 24.31161118 | 24.45141983 |
| 25.04925728 | 25.19071388 | 25.26709175 |
| 26.46660423 | 26.19386673 | 26.40906334 |
| 24.91656113 | 24.83592796 | 24.87224388 |
| 27.14778137 | 27.24262238 | 27.15943718 |
| 19.72163963 | 19.8097744 | 19.72450066 |

| | | |
|---|---|---|
| 24.00804901 | 24.02797318 | 23.91907501 |
| 24.14003372 | 24.11084175 | 24.20431519 |
| 24.85007668 | 24.90639687 | 24.66609001 |
| 25.64055824 | 25.69813538 | 25.66046906 |
| 22.66075325 | 22.74733543 | 22.52647972 |
| 26.64438438 | 26.89345741 | 26.69749641 |
| 25.82257271 | 25.80768394 | 25.68384361 |
| 29.30773735 | 29.20165062 | 29.35181236 |
| 23.75684929 | 23.70039749 | 23.73043633 |
| 27.13476372 | 26.99843025 | 27.05713844 |
| 26.53547287 | 26.41206932 | 26.38298225 |
| 25.22228813 | 25.29951096 | 25.30164719 |
| 24.01337814 | 24.14003944 | 24.14470291 |
| 23.74723816 | 23.74102974 | 23.70412254 |
| 24.44003105 | 24.4398613 | 24.56114197 |
| 29.54458618 | 29.59806252 | 29.43784332 |
| 23.57094574 | 23.60154915 | 23.44895363 |
| 26.50123405 | 26.52178001 | 26.4648571 |
| 27.02294731 | 26.96837425 | 26.98109436 |
| 19.87218666 | 19.72516251 | 19.75913811 |
| 24.92090988 | 25.03611755 | 25.12382507 |
| 26.43885231 | 26.5191803 | 26.47579575 |
| 30.55718613 | 30.35489082 | 30.43411255 |
| 25.90156555 | 25.81129456 | 25.93414879 |
| 26.57255173 | 26.74062729 | 26.65703392 |
| 25.43271446 | 25.42603493 | 25.45964813 |
| 24.18684196 | 24.03427315 | 24.23502731 |
| 23.58859825 | 23.72880173 | 23.66694832 |
| 27.58474922 | 27.58295059 | 27.51792336 |
| 24.93863106 | 24.88706017 | 24.887743 |
| 23.3583107 | 23.25821304 | 23.37919235 |
| 24.94579315 | 25.07362556 | 24.95939445 |
| 29.83751106 | 29.9241848 | 29.91485023 |
| 28.06422806 | 28.09489632 | 28.00605583 |
| 29.67731667 | 29.80396843 | 29.73439789 |
| 26.92365837 | 26.94565964 | 27.0116024 |
| 23.98594093 | 23.97236252 | 24.07691193 |
| 26.68536568 | 26.52348709 | 26.6107769 |
| 25.8562088 | 26.03095055 | 26.03348732 |
| 25.44638824 | 25.34775734 | 25.4516468 |
| 25.02898979 | 25.05923653 | 25.05429077 |
| 24.33598709 | 24.37556839 | 24.45453262 |
| 28.1636982 | 27.98296928 | 28.15125656 |
| 29.7978878 | 29.99887657 | 29.96186066 |
| 27.30646324 | 27.19623375 | 27.16726112 |
| 20.59814262 | 20.60333443 | 20.54965401 |
| 29.24036026 | 29.44969749 | 29.27758217 |

| | | |
|---|---|---|
| 26.82127953 | 26.85297966 | 26.87458992 |
| 25.59700775 | 25.78176498 | 25.70270729 |
| 27.28522873 | 27.17598152 | 27.15389252 |
| 27.47666931 | 27.58804321 | 27.56167984 |
| 25.40305901 | 25.27693367 | 25.4351368 |
| 25.46457672 | 25.59380341 | 25.4753952 |
| 25.82682991 | 26.03121567 | 25.96577835 |
| 25.44198799 | 25.24786949 | 25.4428196 |
| 24.32159996 | 24.43832397 | 24.36273766 |
| 29.18200111 | 29.39988708 | 29.33751106 |
| 24.33695221 | 24.23521042 | 24.31564713 |
| 25.47892761 | 25.5144043 | 25.39463425 |
| 25.83667374 | 25.98429298 | 25.8891449 |
| 29.59441185 | 29.5866909 | 29.4969101 |
| 26.04083824 | 25.97691345 | 25.86157608 |
| 27.55372047 | 27.70485306 | 27.60256004 |
| 24.60190201 | 24.54030228 | 24.58840752 |
| 24.3841114 | 24.52429008 | 24.3417263 |
| 27.93131447 | 27.91022682 | 27.87304306 |
| 26.55334473 | 26.61580467 | 26.45893097 |
| 32.44777679 | 32.23809433 | 32.43126678 |
| 26.12486076 | 26.15803528 | 26.22823906 |
| 25.31832504 | 25.41979599 | 25.48912239 |
| 30.67499352 | 30.84039688 | 30.70761681 |
| 27.21763992 | 27.4028244 | 27.28949356 |
| 25.43957329 | 25.26314354 | 25.31161308 |
| 24.96367073 | 25.12760925 | 24.99342537 |
| 27.55330467 | 27.34630775 | 27.4858017 |
| 24.86500359 | 25.01434326 | 24.9725914 |
| 21.73313332 | 21.71024132 | 21.66141891 |
| 22.14702034 | 22.19963264 | 22.24670601 |
| 27.88961601 | 27.77758026 | 27.69112587 |
| 21.71974564 | 21.56508064 | 21.72730637 |
| 27.29298019 | 27.12768173 | 27.31414413 |
| 27.5447731 | 27.64101219 | 27.6838913 |
| 27.48922729 | 27.57632637 | 27.47400093 |
| 25.91350365 | 26.06567383 | 25.90358734 |
| 23.12632751 | 23.21023369 | 23.05079651 |
| 24.20693207 | 24.37650871 | 24.30640221 |
| 31.5318985 | 31.71930885 | 31.56295395 |
| 21.86183929 | 21.96529579 | 21.82639503 |
| 25.6835804 | 25.64399529 | 25.63296318 |
| 22.12012863 | 22.23500061 | 22.16574097 |
| 24.27955246 | 24.39494133 | 24.35263252 |
| 24.12216377 | 24.02163315 | 24.13745117 |
| 28.11216545 | 28.03986359 | 28.02784348 |
| 26.24516678 | 26.18783569 | 26.31181717 |

| | | |
|---|---|---|
| 28.74172211 | 28.709198 | 28.86273384 |
| 28.47870255 | 28.65806389 | 28.477108 |
| 26.07523155 | 26.13774109 | 26.13773537 |
| 25.53555107 | 25.45991516 | 25.65779877 |
| 23.73919296 | 23.67107773 | 23.71883011 |
| 30.05571556 | 30.03512764 | 29.92227936 |
| 24.8651123 | 24.9750843 | 24.95667076 |
| 28.71046257 | 28.77185249 | 28.65896988 |
| 26.08876038 | 26.01618767 | 26.00564384 |
| 24.79636192 | 24.88176155 | 24.88331032 |
| 26.82046318 | 26.974823 | 27.00733185 |
| 25.00967789 | 24.96356392 | 25.07704353 |
| 31.85207939 | 31.86904526 | 31.91414833 |
| 29.99201775 | 29.92416954 | 29.80002213 |
| 28.44799232 | 28.31015778 | 28.492239 |
| 23.69210815 | 23.71037483 | 23.75085831 |
| 24.50380898 | 24.40932846 | 24.47054482 |
| 25.6890316 | 25.84730911 | 25.86972809 |
| 24.54990005 | 24.46705246 | 24.52400017 |
| 26.54022217 | 26.69918633 | 26.61212158 |
| 24.22307587 | 24.24643517 | 24.16420937 |
| 27.01180077 | 27.10450172 | 27.06759071 |
| 23.15575218 | 23.19258881 | 23.07520676 |
| 27.16455269 | 27.24051476 | 27.1395359 |
| 29.3391571 | 29.33545303 | 29.45760155 |
| 24.74490738 | 24.8652153 | 24.81912422 |
| 30.30109215 | 30.16938972 | 30.31639862 |
| 27.08228111 | 27.15761375 | 27.12585258 |
| 22.94199371 | 22.84156036 | 22.97345734 |
| 26.25318718 | 26.18963051 | 26.18689919 |
| 27.69386292 | 27.85303879 | 27.76128578 |
| 24.82431221 | 24.87736702 | 24.73260117 |
| 27.14619255 | 27.2134552 | 27.21585846 |
| 25.70020103 | 25.68415451 | 25.57739449 |
| 26.28143692 | 26.35422707 | 26.31345749 |
| 26.01738358 | 25.98715019 | 26.05756378 |
| 23.84956551 | 23.83843422 | 23.9231205 |
| 27.24970627 | 27.36449242 | 27.31805611 |
| 28.11458397 | 28.18675995 | 28.28986931 |
| 24.97351265 | 24.95715523 | 25.09666061 |
| 27.3290596 | 27.45317078 | 27.3178997 |
| 17.49340439 | 17.38231277 | 17.45854187 |
| 27.35758591 | 27.4796505 | 27.49459839 |
| 23.83114052 | 23.79681206 | 23.87803841 |
| 33.39237213 | 33.40798187 | 33.31847382 |
| 24.95013428 | 25.00040817 | 24.98842621 |
| 21.9355793 | 21.91461372 | 21.82965851 |

| | | |
|---|---|---|
| 26.77093315 | 26.71199989 | 26.71055412 |
| 27.7358532 | 27.88463402 | 27.83275223 |
| 26.13093758 | 25.97466087 | 26.05393219 |
| 31.8837471 | 32.01119995 | 32.0490799 |
| 25.4874115 | 25.55748367 | 25.41195869 |
| 28.13130569 | 28.18737602 | 28.3014698 |
| 31.07017899 | 31.14289474 | 31.14206696 |
| 25.85268211 | 25.84026909 | 25.88268852 |
| 31.61580086 | 31.73653221 | 31.65403366 |
| 24.9002018 | 25.01025009 | 24.91377068 |
| 25.0453167 | 24.94130135 | 24.94311714 |
| 26.60438728 | 26.61029243 | 26.58305931 |
| 23.31098366 | 23.36452484 | 23.42379761 |
| 29.87214851 | 30.02424622 | 29.95716858 |
| 28.48101425 | 28.43676567 | 28.39547348 |
| 29.07816124 | 29.04423332 | 29.06464767 |
| 26.6796627 | 26.6748867 | 26.65262222 |
| 29.42464638 | 29.40028191 | 29.48120689 |
| 30.0467186 | 30.07424164 | 30.05741501 |
| 24.15993118 | 24.1406517 | 24.21488953 |
| 24.85437393 | 24.96792412 | 24.86324501 |
| 28.7203083 | 28.72353554 | 28.81303215 |
| 29.82754898 | 29.85763741 | 29.86203384 |
| 27.55706024 | 27.48552322 | 27.54606438 |
| 24.5794487 | 24.60572624 | 24.61771393 |
| 28.15130424 | 28.27818108 | 28.18910027 |
| 23.29708672 | 23.2993927 | 23.26763344 |
| 28.23941612 | 28.30267906 | 28.31415939 |
| 27.42699814 | 27.35360527 | 27.45292854 |
| 25.39649963 | 25.39485168 | 25.42245483 |
| 27.13417244 | 27.08301926 | 27.15220451 |
| 25.19580841 | 25.2337532 | 25.16093826 |
| 24.25192642 | 24.25693893 | 24.32284546 |
| 24.1571312 | 24.08452034 | 24.07480812 |
| 31.18713379 | 31.05920792 | 31.08463669 |
| 27.90921402 | 27.94599152 | 27.93894768 |
| 29.1380024 | 29.14372826 | 29.16675377 |
| 26.58031845 | 26.6595192 | 26.600914 |
| 23.24152565 | 23.23727036 | 23.24031067 |
| 28.29101181 | 28.38837433 | 28.38704109 |
| 27.03526497 | 27.00340462 | 27.04955864 |
| 26.56774902 | 26.61325264 | 26.56444168 |
| 24.37675858 | 24.41724205 | 24.38485146 |
| 32.14653778 | 32.26925278 | 32.194664 |
| 32.61891937 | 32.65224075 | 32.70304489 |
| 26.82176781 | 26.83865356 | 26.82039833 |
| 25.12938118 | 25.10700035 | 25.08565521 |

| | | |
|---|---|---|
| 27.64090538 | 27.68531799 | 27.6918869 |
| 29.54473495 | 29.57518768 | 29.6039753 |
| 26.87564087 | 26.88134003 | 26.89457512 |
| 31.1518898 | 31.19151115 | 31.13637161 |

| Abundances (Normalized): Plasma | Abundances (Normalized): Plasma | Abundances (Normalized): Plasma6 |
|---|---|---|
| 28.80784607 | 28.93073082 | 20.67742157 |
| 26.74071121 | 21.94113731 | 20.05984116 |
| 20.07814407 | 19.86816216 | 14.76391792 |
| 21.37379074 | 19.68588829 | 17.26603127 |
| 19.85795212 | 17.98435974 | 21.82536507 |
| 22.49059677 | 16.92278671 | 22.5857563 |
| 24.78528976 | 24.77171135 | 24.34285927 |
| 21.85672569 | 22.22202873 | 19.11735344 |
| 18.95773888 | 20.09311676 | 23.53466606 |
| 18.08384514 | 18.31354523 | 17.27458191 |
| 17.61215591 | 14.27220821 | 18.74282837 |
| 14.78991985 | 18.68354988 | 19.6296711 |
| 20.2556324 | 22.76198578 | 20.02262115 |
| 21.94633484 | 21.81523323 | 18.67264938 |
| 14.0180521 | 14.75776768 | 13.69000721 |
| 18.98191452 | 20.44325447 | 20.4478302 |
| 19.67697906 | 18.99807358 | 18.43890572 |
| 17.57164192 | 16.77690315 | 13.89086723 |
| 19.21852303 | 19.30200005 | 18.5437603 |
| 18.68781662 | 14.94720078 | 18.46276474 |
| 22.1438446 | 23.06771851 | 26.57468414 |
| 15.9737072 | 19.03561592 | 19.44337463 |
| 21.06823349 | 20.57036972 | 17.0261364 |
| 19.16587067 | 19.13325691 | 18.85902405 |
| 16.84831429 | 18.52599144 | 20.60084534 |
| 21.1239109 | 18.71823883 | 18.51262093 |
| 23.85270691 | 23.57406998 | 23.04523277 |
| 18.56543922 | 18.21829414 | 15.72574711 |
| 25.82000732 | 26.1562252 | 22.52134323 |
| 22.64396095 | 25.71987534 | 22.30038261 |
| 20.1759758 | 20.12495804 | 16.62488937 |
| 16.40302849 | 20.11629105 | 20.1619072 |
| 19.36093712 | 19.2742939 | 17.90153885 |
| 16.67198563 | 19.34387589 | 19.91466141 |
| 27.04954338 | 22.83931923 | 24.16801834 |
| 21.68059731 | 21.87742996 | 21.5837841 |
| 21.14703941 | 19.64896965 | 17.96611404 |
| 19.74504662 | 19.44782257 | 16.27041435 |
| 19.35812759 | 16.15043259 | 19.40401268 |
| 17.14888763 | 17.5133934 | 20.40154457 |
| 20.40160561 | 20.59233475 | 22.13010025 |
| 21.98722076 | 22.49217415 | 25.93498802 |
| 16.44039917 | 17.90498543 | 18.22148895 |
| 16.26184082 | 19.15040207 | 19.23199844 |

| | | |
|---|---|---|
| 18.59564972 | 20.17658043 | 20.3907299 |
| 23.65500641 | 23.62626457 | 19.86070633 |
| 22.66367149 | 21.11872864 | 20.05523491 |
| 16.147089 | 18.16710854 | 19.30175972 |
| 15.53725433 | 16.87222672 | 17.36347771 |
| 18.93428421 | 18.23054695 | 16.48324394 |
| 20.43505859 | 22.25481987 | 23.58443832 |
| 21.89893341 | 21.31403351 | 18.25395584 |
| 18.7183609 | 21.16767693 | 18.74864197 |
| 20.78084755 | 20.72878838 | 19.99211693 |
| 22.20741463 | 22.88736534 | 25.90706635 |
| 18.90429688 | 15.97525692 | 18.77305984 |
| 13.93716717 | 15.53450489 | 15.8060236 |
| 19.23543739 | 19.43275452 | 16.30068588 |
| 18.84802818 | 19.12102127 | 18.45919991 |
| 19.75302124 | 20.02293968 | 20.18134499 |
| 20.04384804 | 20.11108017 | 17.29738045 |
| 20.25100899 | 20.30072021 | 18.39032555 |
| 21.33324814 | 21.11869621 | 20.92661476 |
| 17.31670761 | 17.97623444 | 20.13119507 |
| 17.37684059 | 18.30097389 | 17.52919388 |
| 18.67821503 | 18.42220497 | 19.33631325 |
| 17.56680298 | 17.30148888 | 15.46503639 |
| 18.14129639 | 16.49408913 | 18.59715652 |
| 15.11969185 | 14.9719305 | 17.2446785 |
| 21.31075287 | 21.47026253 | 21.12057304 |
| 16.63527679 | 19.57612801 | 18.98461914 |
| 16.77621269 | 17.07432175 | 16.13557816 |
| 17.91476059 | 21.1727562 | 20.67878151 |
| 20.45504951 | 20.44070816 | 22.04018593 |
| 21.25272751 | 21.4977932 | 24.29179001 |
| 22.94105339 | 22.73194122 | 20.77158928 |
| 17.27030182 | 19.13721085 | 18.68856621 |
| 23.95224762 | 24.32972527 | 21.83574486 |
| 22.00225258 | 21.81794548 | 20.20802689 |
| 15.52344704 | 16.23886681 | 17.61838913 |
| 20.01693344 | 19.89604759 | 17.54865456 |
| 20.10523796 | 19.47861099 | 19.34853363 |
| 20.53643227 | 20.86346436 | 20.73844337 |
| 23.18248367 | 23.21620369 | 20.19342422 |
| 24.72681808 | 25.01943207 | 24.84596062 |
| 16.12721443 | 15.89454269 | 18.02060509 |
| 16.34646797 | 17.86747932 | 17.68611526 |
| 19.25151253 | 18.13876724 | 20.15811348 |
| 18.04787636 | 18.26226997 | 17.15060616 |
| 20.17970276 | 20.09447861 | 20.23477745 |
| 15.27849293 | 16.47167778 | 17.16257668 |

| | | |
|---|---|---|
| 26.05235291 | 26.32060623 | 25.92516708 |
| 20.92116737 | 22.23874283 | 19.43897057 |
| 24.78530884 | 25.21978188 | 22.52250671 |
| 19.5982151 | 18.62689972 | 17.03969955 |
| 20.09104538 | 20.89877701 | 18.26435661 |
| 19.96443367 | 19.68806839 | 17.56194496 |
| 19.30343056 | 20.43770027 | 18.9592514 |
| 17.94158173 | 18.25028992 | 16.39546585 |
| 24.48372459 | 24.66522217 | 22.56481743 |
| 18.32289314 | 18.03312683 | 19.04528809 |
| 18.7610569 | 18.46042824 | 16.41956711 |
| 17.88359833 | 18.93586922 | 20.39126015 |
| 20.81445503 | 19.97289085 | 18.97719765 |
| 24.90150642 | 25.12144661 | 22.62364769 |
| 21.73167229 | 20.22165489 | 21.03877068 |
| 20.60680199 | 21.60645103 | 21.59645844 |
| 22.24475861 | 22.59716034 | 19.83930016 |
| 18.5177784 | 19.23296165 | 17.44821167 |
| 20.84667587 | 20.59717941 | 18.33526421 |
| 21.15055656 | 22.84015846 | 23.55870056 |
| 19.56719017 | 20.14530563 | 20.73157883 |
| 18.28025818 | 18.06694984 | 16.39893532 |
| 18.21741676 | 16.27707291 | 18.23973846 |
| 20.27433968 | 21.62867928 | 21.94492531 |
| 24.44696808 | 25.5949192 | 26.9552784 |
| 20.38372993 | 20.48413467 | 18.20941925 |
| 18.44778252 | 18.5541954 | 18.11790848 |
| 17.37415504 | 18.71261215 | 18.99941826 |
| 22.55274391 | 22.90414619 | 25.28643799 |
| 18.15642166 | 19.81385612 | 18.03184128 |
| 22.06078911 | 23.23507881 | 21.38931274 |
| 19.6709919 | 19.61606789 | 19.23903847 |
| 20.12799072 | 19.45424843 | 17.6434536 |
| 18.84212685 | 18.66862297 | 20.31671143 |
| 20.85828972 | 20.97468948 | 19.77347183 |
| 16.09490585 | 15.41466618 | 15.50137615 |
| 18.91497993 | 18.65638161 | 18.99832344 |
| 21.38908005 | 23.63274765 | 21.21149635 |
| 19.34589767 | 17.36904335 | 19.15126991 |
| 23.35976601 | 23.66975021 | 25.73948288 |
| 21.37750435 | 21.20996857 | 19.60146141 |
| 19.10085106 | 20.74983788 | 20.98147392 |
| 20.58958244 | 18.19827652 | 19.17469215 |
| 22.67315865 | 22.5559063 | 21.04350471 |
| 20.27580452 | 20.39095688 | 22.22596931 |
| 19.51770973 | 21.48772049 | 19.80348778 |
| 18.10279274 | 19.51644516 | 19.24072075 |

| | | |
|---|---|---|
| 17.29099846 | 18.88932037 | 19.17965126 |
| 21.34157372 | 21.13689613 | 21.47583008 |
| 23.08671951 | 24.43489075 | 22.20286369 |
| 17.18662453 | 18.33526993 | 18.28535271 |
| 22.75151443 | 22.52882767 | 22.56969643 |
| 19.99180412 | 19.71229172 | 20.16199684 |
| 20.8018074 | 19.50655365 | 19.0984993 |
| 18.72619438 | 18.9753418 | 16.9682045 |
| 23.61098289 | 24.30150795 | 21.41666412 |
| 19.27029991 | 21.19368935 | 21.04284668 |
| 20.74209404 | 19.14048767 | 19.43911362 |
| 15.92165661 | 16.48023605 | 15.7858429 |
| 18.46941185 | 18.673069 | 20.26960564 |
| 19.37581062 | 19.11000824 | 20.88416481 |
| 19.76404381 | 20.43221092 | 20.18288803 |
| 17.30884171 | 17.72802162 | 17.87866783 |
| 24.47400093 | 24.81341743 | 22.47345352 |
| 18.48233223 | 20.65672493 | 20.38716698 |
| 20.55737686 | 20.48338509 | 20.20059204 |
| 16.20425797 | 17.54432106 | 16.30787086 |
| 20.88513565 | 19.02035522 | 20.7200222 |
| 17.90023422 | 17.84499741 | 16.39705086 |
| 19.42066956 | 20.3441124 | 19.83556175 |
| 21.16860962 | 21.95857239 | 21.14142418 |
| 20.20033073 | 21.43871117 | 22.03513527 |
| 21.52932167 | 21.57194138 | 19.995821 |
| 18.26119232 | 19.0931282 | 18.08571434 |
| 22.0848999 | 21.26102257 | 22.1876049 |
| 22.19097519 | 22.47617531 | 23.38031769 |
| 21.98712158 | 22.01696777 | 21.75660515 |
| 19.28731537 | 19.22258949 | 19.08515739 |
| 18.95708656 | 19.02503777 | 18.04675484 |
| 19.52985191 | 19.09165573 | 17.74821281 |
| 20.22403908 | 21.9084549 | 22.6984787 |
| 19.33870316 | 19.07260132 | 19.2399807 |
| 21.40436172 | 22.5558567 | 20.54243469 |
| 19.25512505 | 17.777771 | 19.64602661 |
| 19.21726036 | 20.51490021 | 20.61044502 |
| 20.52996063 | 21.51178551 | 22.11190414 |
| 19.05489731 | 17.42307472 | 19.30411148 |
| 18.52038574 | 19.06617546 | 19.01646042 |
| 20.58680916 | 21.62386513 | 21.11253929 |
| 17.35348511 | 17.04911613 | 16.04438591 |
| 21.93810463 | 24.11870003 | 22.44698715 |
| 19.58429337 | 20.54439926 | 18.47635651 |
| 18.23799706 | 18.08033562 | 19.64450645 |
| 22.24565506 | 23.59769821 | 21.61523247 |

| | | |
|---|---|---|
| 21.87878418 | 22.73165512 | 23.63155174 |
| 23.39052773 | 24.12501335 | 24.73961639 |
| 22.53161812 | 22.55991364 | 21.91952133 |
| 20.14752388 | 20.52624321 | 19.82583618 |
| 19.09333611 | 19.21545792 | 18.82512665 |
| 20.18958855 | 19.71942139 | 19.65946007 |
| 23.51909637 | 23.58999634 | 22.35751343 |
| 20.77414322 | 20.23048592 | 19.21640205 |
| 17.04636574 | 17.97997665 | 17.5633049 |
| 19.74232674 | 19.78293991 | 19.04512978 |
| 21.37416458 | 22.50989532 | 23.65620804 |
| 14.49457264 | 14.74868107 | 13.52842712 |
| 21.38207817 | 20.87442398 | 19.90797806 |
| 22.38889313 | 22.13147926 | 20.88690948 |
| 22.61656761 | 24.35423088 | 22.67544174 |
| 21.32214737 | 20.99368095 | 19.63820076 |
| 21.30050659 | 20.43391609 | 21.50315285 |
| 21.15047455 | 19.54315758 | 19.18429756 |
| 23.6985321 | 23.4030571 | 24.93917465 |
| 18.98821449 | 20.35084343 | 19.28216934 |
| 22.86359024 | 22.86699295 | 22.75004959 |
| 20.56002617 | 21.34218788 | 21.57989311 |
| 22.46488953 | 22.83931923 | 22.92617607 |
| 18.395504 | 18.8156662 | 20.16676521 |
| 19.37158585 | 20.80175209 | 20.38849831 |
| 17.74873924 | 16.28850937 | 17.43476295 |
| 19.9496479 | 19.63670158 | 21.44070816 |
| 24.1516571 | 24.27238083 | 22.51369667 |
| 22.27770615 | 22.94727707 | 24.03632164 |
| 21.86029434 | 22.0384903 | 21.821455 |
| 19.84908104 | 20.01269531 | 20.46479225 |
| 18.65007973 | 18.58267593 | 19.15602875 |
| 18.4360981 | 18.58707809 | 18.50050735 |
| 17.96644592 | 17.98309708 | 16.91123199 |
| 23.12858391 | 22.99509811 | 21.39917183 |
| 22.08128548 | 22.18388557 | 23.30463409 |
| 23.37221718 | 23.38746262 | 21.38614464 |
| 19.85181236 | 19.70374298 | 20.61200333 |
| 18.67154312 | 19.21529961 | 18.22859001 |
| 19.75609016 | 21.01424217 | 21.56461525 |
| 19.42437935 | 19.98439026 | 19.55836487 |
| 20.7673912 | 19.04595184 | 19.61997223 |
| 16.51761818 | 16.59773827 | 16.54977989 |
| 22.0800457 | 22.16745758 | 22.5914135 |
| 22.03139496 | 21.02915955 | 19.96456718 |
| 21.31926155 | 21.83073616 | 21.53951263 |
| 18.85529327 | 20.12470818 | 20.47120285 |

| | | |
|---|---|---|
| 20.78430748 | 20.40278435 | 20.91201019 |
| 20.85894775 | 20.89559364 | 21.23336601 |
| 26.24205971 | 24.6482029 | 24.9718895 |
| 23.24594116 | 21.60153961 | 21.13403702 |
| 18.23768234 | 19.2574482 | 19.31806755 |
| 18.65214539 | 18.68910027 | 17.35795593 |
| 20.96418381 | 19.32761574 | 21.1551857 |
| 19.88722992 | 18.58923149 | 18.40143204 |
| 19.99772835 | 21.65017891 | 19.72474861 |
| 20.01509857 | 21.33045387 | 21.41484261 |
| 21.16061783 | 21.99645805 | 20.10737991 |
| 19.48404503 | 19.96243477 | 20.32373428 |
| 18.61517334 | 18.26919746 | 17.09133148 |
| 19.55228043 | 19.31957436 | 20.89749718 |
| 20.13861275 | 20.77681351 | 20.33762932 |
| 18.88569641 | 20.40515327 | 19.3432579 |
| 24.4718914 | 23.19570923 | 22.93165016 |
| 19.95255089 | 20.14202499 | 18.76217651 |
| 18.27002335 | 18.80353355 | 17.71822739 |
| 17.14011765 | 16.26144981 | 15.91109753 |
| 23.15869331 | 22.67858124 | 21.9492054 |
| 23.56478882 | 21.72601318 | 21.56461143 |
| 19.38255501 | 19.59177399 | 18.79324913 |
| 19.35429955 | 19.31739998 | 18.91150665 |
| 23.07973671 | 22.79419327 | 21.50233459 |
| 23.03341675 | 24.49865341 | 23.25317192 |
| 18.55651283 | 19.02259254 | 17.38650894 |
| 22.39710617 | 22.55151939 | 21.07844162 |
| 24.06520844 | 23.11136436 | 24.76585579 |
| 26.99272156 | 26.88046265 | 28.90851593 |
| 23.62420273 | 23.66586494 | 23.61743546 |
| 20.58045769 | 18.79755974 | 19.80737305 |
| 23.44851685 | 23.50810051 | 22.47163582 |
| 19.63840675 | 18.76453209 | 19.37716484 |
| 18.50282288 | 19.92802048 | 19.67440987 |
| 21.759058 | 21.76759148 | 22.671175 |
| 19.15031433 | 17.8081646 | 18.81650543 |
| 20.70552254 | 20.38541985 | 19.72126961 |
| 20.81099319 | 19.83618927 | 19.71366692 |
| 21.70678329 | 21.81664085 | 21.47766113 |
| 18.58255386 | 19.80172539 | 19.58406258 |
| 20.54197693 | 21.20995903 | 21.27406311 |
| 21.22001076 | 21.09454918 | 19.93665504 |
| 24.42294502 | 24.99118805 | 26.56621933 |
| 23.24863815 | 24.06982994 | 24.4462986 |
| 19.13731766 | 19.60425949 | 19.82705116 |
| 21.26181984 | 21.15418243 | 19.64079094 |

| | | |
|---|---|---|
| 21.81048584 | 23.43723297 | 22.4866066 |
| 17.86493683 | 18.0970192 | 18.40135193 |
| 17.71842194 | 17.64787483 | 16.66568184 |
| 23.73071098 | 23.50342941 | 23.69559479 |
| 19.72410011 | 20.10539246 | 20.60031891 |
| 19.81894684 | 19.4647541 | 20.37020302 |
| 23.80643654 | 25.1259613 | 23.38446236 |
| 19.51602364 | 20.13294411 | 18.83789635 |
| 22.46149635 | 22.25418091 | 20.91312218 |
| 21.18292999 | 22.16472054 | 20.50290108 |
| 16.66792488 | 17.11648369 | 16.3450737 |
| 21.02140617 | 19.54516983 | 20.58620834 |
| 19.53608513 | 19.1516552 | 20.47686005 |
| 20.6061554 | 21.01212883 | 21.68715096 |
| 22.06612968 | 20.9755497 | 22.59410477 |
| 21.47181511 | 21.34261703 | 20.24246979 |
| 19.41619682 | 19.35795975 | 18.2122879 |
| 20.29505539 | 20.43245506 | 19.75324249 |
| 20.91547585 | 20.43312073 | 20.41910744 |
| 18.2479744 | 19.67132187 | 19.58548927 |
| 17.74404526 | 18.04268837 | 17.16095924 |
| 20.75218201 | 20.59264755 | 19.91033936 |
| 22.34858704 | 22.02499962 | 22.60702324 |
| 20.74863052 | 21.22392464 | 21.67174149 |
| 21.89988708 | 21.69846535 | 21.73759651 |
| 23.02462769 | 22.37423706 | 21.89959526 |
| 22.20442009 | 22.62203598 | 23.02613831 |
| 19.98557663 | 21.2536087 | 19.83867645 |
| 22.77680969 | 21.99258804 | 21.37630463 |
| 20.67593575 | 21.58125496 | 21.81417084 |
| 19.43747139 | 19.09714127 | 19.08556366 |
| 17.09026909 | 17.09084129 | 16.2988224 |
| 18.88757896 | 19.82657814 | 18.89869881 |
| 27.57980537 | 27.19565201 | 27.52410316 |
| 20.91620636 | 21.98204231 | 22.01934814 |
| 21.16911888 | 21.12024879 | 19.9575882 |
| 24.69612885 | 25.63817596 | 26.44889832 |
| 20.74076462 | 21.31162071 | 21.73817444 |
| 19.93130875 | 20.62607193 | 20.15724182 |
| 27.00509262 | 28.02692604 | 28.60030174 |
| 19.75000763 | 19.87772179 | 19.76779366 |
| 19.39154053 | 19.27781487 | 19.39678192 |
| 27.77911758 | 26.95815659 | 26.57496643 |
| 20.23436356 | 20.21158981 | 19.70922279 |
| 19.34015083 | 19.4010067 | 18.74859238 |
| 21.18597031 | 21.25650787 | 21.31092834 |
| 23.84643364 | 24.07675743 | 25.42200089 |

| | | |
|---|---|---|
| 22.05765724 | 21.87836456 | 20.92252541 |
| 19.01420784 | 18.44824219 | 18.8857193 |
| 18.69893074 | 18.66177177 | 19.47967529 |
| 17.75958633 | 17.96159744 | 17.20592308 |
| 20.39325523 | 19.66303635 | 20.17413521 |
| 19.17961884 | 18.96289444 | 18.93506622 |
| 23.03611183 | 23.97070694 | 22.75838661 |
| 23.37017822 | 23.25740623 | 23.43099976 |
| 22.51508141 | 22.34990692 | 22.20710182 |
| 23.22694778 | 22.1239109 | 22.8990612 |
| 21.20337486 | 21.26134872 | 21.33611107 |
| 17.77813148 | 17.91747665 | 16.93217278 |
| 22.6919651 | 22.94756889 | 22.8970871 |
| 17.35824394 | 17.03003693 | 17.7838192 |
| 18.93730927 | 19.154356 | 17.98234367 |
| 19.55699921 | 19.00115776 | 18.974226 |
| 21.18831444 | 21.49857712 | 20.37286758 |
| 19.91801453 | 19.53516006 | 19.55529213 |
| 21.1330452 | 20.90707588 | 21.17096138 |
| 20.5378685 | 21.55606842 | 20.37541199 |
| 21.91597557 | 21.57755661 | 21.04418945 |
| 21.73736572 | 21.77902794 | 20.9265728 |
| 22.35824203 | 21.02521706 | 21.13953018 |
| 19.00902557 | 18.22210121 | 19.47593498 |
| 17.3503933 | 17.21150398 | 17.12313652 |
| 20.42347145 | 20.47550392 | 20.92358589 |
| 18.23893547 | 18.75065804 | 18.61476707 |
| 22.36403465 | 22.65310478 | 21.51699638 |
| 21.1330986 | 21.46038628 | 21.65337753 |
| 20.39448547 | 20.99819183 | 21.57793617 |
| 22.60449982 | 23.30525017 | 23.16619492 |
| 22.39874458 | 22.1323719 | 21.17597008 |
| 18.40084267 | 18.88919258 | 18.09968567 |
| 21.88989449 | 23.2823391 | 22.53289986 |
| 18.23239517 | 18.17407036 | 17.30523491 |
| 17.71716881 | 17.86322594 | 18.62924767 |
| 18.69684601 | 19.47476387 | 19.56876183 |
| 20.07159996 | 20.00633812 | 21.19965553 |
| 21.86801529 | 22.08696747 | 22.17722511 |
| 20.81838226 | 20.64312363 | 20.6417923 |
| 21.12809372 | 20.83701324 | 20.53463364 |
| 19.51113892 | 20.04194641 | 20.72597122 |
| 19.12436867 | 19.4053421 | 19.3751812 |
| 26.57888031 | 27.12303734 | 27.79143524 |
| 20.57455444 | 20.19928932 | 20.64665031 |
| 20.95131111 | 21.82086182 | 22.1172905 |
| 21.58869362 | 21.48626709 | 22.39786339 |

| | | |
|---|---|---|
| 21.7115078 | 21.18939972 | 20.77314758 |
| 23.08972359 | 22.98789597 | 23.12349892 |
| 21.15891075 | 21.13185501 | 21.17964554 |
| 21.20788574 | 20.94016457 | 22.20031166 |
| 20.24722481 | 19.49164391 | 20.00896645 |
| 20.10397339 | 19.27007484 | 19.08153725 |
| 19.62085152 | 19.57571793 | 18.71032524 |
| 18.08707047 | 18.18751526 | 17.74599075 |
| 19.12892723 | 19.66585541 | 18.61833 |
| 21.09938431 | 21.50291443 | 20.58725548 |
| 16.90055275 | 18.05308533 | 17.4232254 |
| 24.01084518 | 24.23464203 | 23.44282722 |
| 21.19286537 | 21.06418419 | 21.61889458 |
| 17.27886391 | 17.70467377 | 17.92555046 |
| 23.57012558 | 23.50758934 | 24.81075478 |
| 21.83136368 | 21.4556427 | 20.59209633 |
| 20.82962036 | 20.33395767 | 21.61058617 |
| 19.71142387 | 19.93733978 | 19.86961174 |
| 18.32374763 | 18.23443222 | 17.49028206 |
| 19.62947273 | 19.67995644 | 19.8639946 |
| 21.12099457 | 21.12332344 | 21.14894295 |
| 21.63460159 | 21.82790756 | 21.14931679 |
| 17.22898865 | 18.28859901 | 18.28042412 |
| 22.27860069 | 22.24972916 | 21.26450729 |
| 19.6611557 | 18.71785545 | 18.98562431 |
| 20.62616348 | 21.55214119 | 20.83348656 |
| 20.71507645 | 20.77710533 | 20.8005085 |
| 25.02219009 | 26.09859467 | 25.88308716 |
| 19.38504982 | 19.51383781 | 18.58097839 |
| 19.48248482 | 19.03244781 | 19.0059166 |
| 18.27228737 | 18.80970573 | 19.06209946 |
| 22.60026741 | 22.38780594 | 23.74895668 |
| 24.19334412 | 25.40975189 | 24.0053215 |
| 22.0090847 | 21.44976616 | 21.09031487 |
| 25.9146862 | 26.27128029 | 25.77624321 |
| 21.22611809 | 20.75944519 | 21.21876907 |
| 21.59257317 | 21.37498665 | 22.08389854 |
| 20.09370804 | 19.99592972 | 20.95202827 |
| 17.98043633 | 18.29450989 | 17.32902336 |
| 21.25432396 | 21.37906647 | 20.5988903 |
| 21.45499039 | 21.67292213 | 20.54452515 |
| 21.47062111 | 22.25763321 | 22.39332008 |
| 19.20457458 | 18.82775688 | 19.64449692 |
| 19.09382439 | 18.86511993 | 18.36773682 |
| 22.43004036 | 22.63626862 | 22.27126122 |
| 20.37157249 | 19.84518242 | 20.52467537 |
| 18.76509094 | 19.4718132 | 19.90266991 |

| | | |
|---|---|---|
| 20.36760521 | 19.78247833 | 20.11409569 |
| 21.04583359 | 21.45660973 | 20.83663559 |
| 21.41692352 | 21.47142029 | 20.37663269 |
| 17.62285805 | 18.51965523 | 18.5974865 |
| 20.26494408 | 20.42763138 | 19.86050797 |
| 20.99092674 | 21.41693497 | 21.49426079 |
| 25.54381752 | 26.58045197 | 26.55620193 |
| 21.96138382 | 21.95739174 | 22.44722176 |
| 21.63312721 | 21.41086388 | 22.70672035 |
| 20.9140625 | 21.50109673 | 21.5200634 |
| 22.84209251 | 22.22442245 | 23.12527466 |
| 22.25800896 | 21.81528473 | 21.67276955 |
| 22.42733955 | 21.42018127 | 21.30013657 |
| 21.75226021 | 22.75787354 | 21.86025238 |
| 22.50306702 | 21.52537918 | 21.84854889 |
| 20.71853256 | 20.81440926 | 21.07005882 |
| 22.85614967 | 23.28639221 | 23.87964439 |
| 19.2340374 | 20.23212051 | 20.40974617 |
| 20.30532265 | 20.98355484 | 20.06616592 |
| 21.51783752 | 21.19652367 | 21.91883659 |
| 21.27216911 | 21.28428268 | 21.27867889 |
| 21.67363739 | 21.34961891 | 20.53765297 |
| 22.21889877 | 22.24917412 | 21.43403435 |
| 20.55292892 | 21.10406303 | 21.15476608 |
| 21.72772598 | 21.59884834 | 21.29455185 |
| 23.17141914 | 23.17618752 | 22.00700378 |
| 20.62626839 | 20.69901657 | 21.69730186 |
| 20.37498665 | 19.32056046 | 20.30952644 |
| 26.60365486 | 26.30763245 | 25.19410133 |
| 24.39043236 | 24.77159691 | 23.67992401 |
| 17.13392258 | 17.09518051 | 17.14862061 |
| 18.44221878 | 17.95710945 | 17.73001862 |
| 21.0653801 | 21.62534714 | 20.48693275 |
| 19.62555885 | 19.55548096 | 20.16321945 |
| 22.89474487 | 22.90888596 | 23.60770798 |
| 17.74971581 | 18.40174866 | 18.58180046 |
| 22.13824081 | 22.2211647 | 21.18515396 |
| 20.20213699 | 20.0483532 | 20.37707329 |
| 20.96490288 | 20.78163719 | 20.1999855 |
| 21.23530006 | 21.52874947 | 20.4040432 |
| 25.26939774 | 25.43498802 | 24.21543884 |
| 21.02272415 | 21.14523697 | 20.30922318 |
| 21.98261833 | 21.95653725 | 22.06759834 |
| 19.68838882 | 19.37255478 | 18.83434296 |
| 20.15094948 | 19.84984589 | 20.87578964 |
| 22.18275452 | 21.9431324 | 23.00815392 |
| 22.17274284 | 22.85146904 | 21.83369637 |

| | | |
|---|---|---|
| 20.51021576 | 21.18273354 | 20.37573814 |
| 21.37493134 | 21.74700546 | 21.65229607 |
| 24.90929413 | 23.51315689 | 24.21694183 |
| 18.18815041 | 18.58061409 | 18.19372559 |
| 21.51718903 | 21.14115715 | 21.44482613 |
| 19.13516426 | 19.27972984 | 18.32860947 |
| 22.52543831 | 22.31224442 | 23.270998 |
| 22.97332382 | 23.10207939 | 22.01396942 |
| 23.63471222 | 24.20323181 | 24.939991 |
| 20.51445198 | 20.59051132 | 19.70720673 |
| 20.7391777 | 20.62788391 | 19.72587204 |
| 26.10575867 | 26.72613716 | 26.73801613 |
| 21.7028656 | 21.97339058 | 22.89832497 |
| 17.99425697 | 18.84539986 | 19.04268837 |
| 21.25492477 | 20.8605175 | 20.87231064 |
| 20.94787979 | 21.89391708 | 21.04410744 |
| 19.08584785 | 18.64249802 | 19.74800491 |
| 17.91955566 | 18.12350655 | 18.17676353 |
| 16.52446556 | 16.32887459 | 15.63937473 |
| 19.09360886 | 19.11764145 | 19.17958069 |
| 19.17929459 | 19.30731964 | 18.68700218 |
| 22.89373589 | 23.69899368 | 22.65930176 |
| 22.09162712 | 21.65317345 | 22.84825897 |
| 25.97312546 | 26.11524391 | 25.01346588 |
| 20.84840965 | 20.68947029 | 21.6942749 |
| 21.15586853 | 20.92017174 | 20.83023643 |
| 16.17323303 | 16.58474922 | 16.21917534 |
| 19.69107628 | 19.81190491 | 20.45698166 |
| 21.77713776 | 21.81064224 | 22.45939064 |
| 19.84393692 | 19.38526726 | 20.02040291 |
| 21.64229202 | 21.20660973 | 20.77238464 |
| 23.90734863 | 23.8064537 | 23.52791214 |
| 19.7745266 | 19.35662842 | 18.96271896 |
| 19.40114594 | 19.35972786 | 19.86369514 |
| 26.86086464 | 26.68699837 | 25.76302147 |
| 18.5188694 | 18.5754528 | 18.89842796 |
| 20.81676865 | 20.8370018 | 20.59396744 |
| 22.62080383 | 22.57658386 | 22.42775917 |
| 17.90585136 | 18.16172409 | 17.33041382 |
| 21.29474449 | 21.93374443 | 21.26097107 |
| 20.15322876 | 20.73756027 | 20.27269173 |
| 16.35054016 | 16.52186012 | 15.83941078 |
| 24.00519562 | 23.80623817 | 22.97790337 |
| 19.64814186 | 20.47711754 | 19.69605064 |
| 19.24468803 | 18.53783226 | 19.1468544 |
| 19.71762276 | 19.73793793 | 19.36778641 |
| 18.27768707 | 17.972332 | 17.85984802 |

| | | |
|---|---|---|
| 21.18759155 | 21.08121681 | 20.80372047 |
| 17.77714729 | 17.67723846 | 18.57346535 |
| 17.5274868 | 17.72210884 | 17.20486641 |
| 18.92604828 | 19.23550606 | 18.92089462 |
| 24.75798988 | 24.77806473 | 24.52707291 |
| 22.08585358 | 22.27537537 | 21.53656197 |
| 20.65383148 | 21.10889435 | 21.27429771 |
| 19.2330761 | 18.51584816 | 18.34305573 |
| 18.04090881 | 18.28229141 | 17.43478203 |
| 20.66989899 | 20.44879532 | 20.20958138 |
| 20.89074898 | 20.66272545 | 20.16090012 |
| 20.17369461 | 20.08021164 | 19.48370171 |
| 20.02554131 | 19.71172523 | 19.60434532 |
| 22.56155014 | 22.12752914 | 23.32765961 |
| 18.30949974 | 18.40972137 | 18.39483261 |
| 20.13683319 | 20.62147522 | 21.23217964 |
| 17.60753059 | 18.07865906 | 17.75084496 |
| 19.84873009 | 19.81757164 | 20.71651268 |
| 25.16400719 | 25.71486473 | 25.04677582 |
| 21.60776329 | 21.57136154 | 22.20075798 |
| 23.20228386 | 22.73546028 | 22.72905159 |
| 22.92056084 | 22.62628174 | 22.04843903 |
| 22.49058914 | 22.0758667 | 21.39687729 |
| 21.98708534 | 21.7443676 | 21.03286552 |
| 22.11541557 | 22.72659492 | 22.36003876 |
| 22.89019012 | 24.04259872 | 23.02214622 |
| 19.26376915 | 19.87417603 | 20.12466431 |
| 24.11131287 | 23.0169239 | 24.05808067 |
| 20.24837303 | 19.80510712 | 19.43621445 |
| 21.55373192 | 21.40846252 | 21.58834267 |
| 24.2081852 | 24.83142853 | 25.07202339 |
| 22.57099915 | 22.41764832 | 21.65460968 |
| 20.88945198 | 20.19956398 | 20.32564163 |
| 22.88680649 | 22.97389984 | 23.90731812 |
| 19.87086487 | 20.02601624 | 19.32839584 |
| 21.19552422 | 21.20042229 | 20.43240356 |
| 22.72595596 | 22.25388336 | 21.86730576 |
| 24.84946632 | 24.89435577 | 25.69788361 |
| 26.27741814 | 25.25569916 | 25.06668854 |
| 24.35167313 | 24.79865837 | 24.8947773 |
| 18.38713646 | 18.529953 | 17.86675072 |
| 19.43758011 | 19.57180214 | 19.67760277 |
| 21.10417557 | 22.04211617 | 21.2161541 |
| 19.21206665 | 18.4862957 | 19.39396667 |
| 17.78245926 | 17.73222542 | 17.75802803 |
| 20.69424629 | 20.73418617 | 21.30957794 |
| 21.35297966 | 21.77069855 | 21.81204987 |

| | | |
|---|---|---|
| 18.45775986 | 18.55410385 | 18.06167793 |
| 20.2505188 | 20.39754677 | 20.81752205 |
| 25.00505257 | 25.13040733 | 24.91735649 |
| 20.66741943 | 21.04099464 | 20.49076462 |
| 18.99769592 | 18.45018005 | 19.17399406 |
| 24.47255135 | 24.20515442 | 24.14843369 |
| 22.67878532 | 22.72174835 | 22.29390526 |
| 20.50929832 | 20.48924255 | 20.64031792 |
| 19.98315048 | 19.45583534 | 19.76958084 |
| 21.66292191 | 21.62447739 | 22.18653107 |
| 23.03104401 | 22.98467255 | 22.16462135 |
| 20.91268349 | 21.66984558 | 21.74641609 |
| 21.76643181 | 21.94352341 | 21.78108978 |
| 21.00331879 | 21.29018211 | 21.55212402 |
| 22.84536552 | 22.71741104 | 22.85596085 |
| 18.77843857 | 19.5197525 | 19.42464638 |
| 20.67305756 | 20.7816143 | 20.02321243 |
| 22.85828209 | 23.15584373 | 22.31494141 |
| 22.44932365 | 23.09451103 | 22.41559029 |
| 21.07902908 | 20.90704346 | 20.83194923 |
| 25.61874962 | 25.80578423 | 25.54950142 |
| 21.45277214 | 21.76761436 | 22.26192093 |
| 19.09713745 | 19.45237732 | 18.98645401 |
| 23.20603561 | 23.14066887 | 22.42307091 |
| 25.10205078 | 25.88492012 | 25.86369896 |
| 23.55545616 | 24.34357071 | 23.25179672 |
| 19.75619698 | 20.2888031 | 19.68345642 |
| 23.21663284 | 23.05598831 | 22.19394493 |
| 20.89832497 | 20.97359467 | 20.14916611 |
| 20.86107826 | 20.76161575 | 20.46059799 |
| 21.06372261 | 21.11494064 | 20.43655586 |
| 19.22582054 | 18.83615303 | 19.56574631 |
| 21.8016243 | 22.15285301 | 21.28203201 |
| 18.10557747 | 18.90580177 | 18.79460144 |
| 20.23615074 | 20.5871067 | 20.50739479 |
| 23.02628326 | 21.97759628 | 22.82102394 |
| 22.24655342 | 22.25564575 | 23.1312027 |
| 23.51290894 | 23.31183434 | 24.35395241 |
| 21.82242966 | 21.73407555 | 21.40938568 |
| 22.0943718 | 22.38904572 | 22.84617805 |
| 19.17008018 | 19.00553894 | 18.57806015 |
| 24.81103516 | 24.42888451 | 24.93376732 |
| 22.72883034 | 22.73089409 | 22.08011055 |
| 22.84087372 | 22.04064941 | 22.7437458 |
| 21.45697594 | 21.92615891 | 22.19175339 |
| 17.95226097 | 17.56632233 | 17.60982323 |
| 20.43238449 | 20.57822227 | 20.4548912 |

| | | |
|---|---|---|
| 19.35319138 | 19.40536499 | 18.66498184 |
| 21.66286278 | 21.8494072 | 22.34254456 |
| 21.93360329 | 21.23586464 | 21.2365303 |
| 22.20246887 | 21.23873711 | 21.3643074 |
| 24.65481567 | 24.81634331 | 24.00997353 |
| 24.95053101 | 24.8635273 | 24.70528221 |
| 22.50959969 | 22.71776199 | 23.1943779 |
| 20.12152672 | 20.47159195 | 20.09507942 |
| 23.32478523 | 23.39256859 | 24.01747322 |
| 20.68586731 | 20.70645332 | 20.80878258 |
| 20.41747856 | 19.76156616 | 19.63747787 |
| 21.5816555 | 21.2456398 | 21.77010536 |
| 20.33510971 | 20.32931519 | 19.73212433 |
| 21.57005692 | 21.19853783 | 20.66674232 |
| 29.02569771 | 29.3867836 | 29.66449356 |
| 24.85239983 | 24.84969711 | 25.78977394 |
| 26.09038353 | 26.18027306 | 25.25310707 |
| 17.62291527 | 17.7690258 | 17.20757294 |
| 18.12323952 | 17.94678307 | 18.70947075 |
| 19.24576187 | 19.21871758 | 19.70113754 |
| 19.5650425 | 19.6638298 | 18.93061638 |
| 21.43429184 | 21.22384262 | 21.36296654 |
| 21.36206436 | 21.89146233 | 22.01955605 |
| 16.53700829 | 17.07081604 | 17.25330162 |
| 19.51413536 | 18.81524277 | 18.84053612 |
| 21.47967339 | 21.12483978 | 21.85779762 |
| 18.59123611 | 18.83944321 | 19.19989777 |
| 19.55108643 | 19.57853889 | 18.87455559 |
| 21.7790184 | 21.82725906 | 22.6848259 |
| 19.87429428 | 19.47574615 | 19.14927864 |
| 23.36587334 | 23.49230957 | 22.88592339 |
| 21.83504677 | 21.70477486 | 22.47774124 |
| 17.47904778 | 17.65665054 | 17.76066017 |
| 23.63455009 | 23.66010475 | 23.13005829 |
| 20.45762825 | 20.29604721 | 20.64204407 |
| 21.13080597 | 21.64662361 | 21.99061584 |
| 17.46069717 | 18.14949417 | 18.14596939 |
| 22.50631523 | 22.53608513 | 23.10271263 |
| 23.63920212 | 23.5414753 | 22.9895134 |
| 19.34418297 | 19.025177 | 19.72871399 |
| 18.71393394 | 18.45151329 | 18.65266037 |
| 22.543396 | 22.60185814 | 22.5718441 |
| 19.56819916 | 19.18800354 | 18.80450249 |
| 21.50223541 | 21.97413826 | 21.42718315 |
| 22.68591881 | 22.50713539 | 21.81761169 |
| 25.24473572 | 25.48651695 | 24.56172752 |
| 20.26968956 | 20.16799736 | 21.03153229 |

| | | |
|---|---|---|
| 23.53497124 | 23.48272324 | 22.91412735 |
| 20.98197556 | 20.8158989 | 20.14562416 |
| 28.8006916 | 29.17106056 | 29.69429588 |
| 21.56445313 | 21.9911232 | 22.16109085 |
| 21.2630024 | 20.40197945 | 20.9325695 |
| 25.13027191 | 25.06206703 | 24.96812057 |
| 19.94968224 | 20.18481064 | 20.18299675 |
| 22.5755024 | 22.68230629 | 21.93081093 |
| 19.47066879 | 19.00848007 | 18.60518074 |
| 21.8571949 | 21.50300598 | 22.18174553 |
| 24.22164345 | 24.19835663 | 23.65591431 |
| 21.56693649 | 21.8202877 | 20.93722153 |
| 27.73791504 | 27.1192646 | 27.11513901 |
| 18.89155388 | 18.39520454 | 18.74222946 |
| 24.65189171 | 24.46888542 | 23.7559185 |
| 18.62895203 | 18.61338425 | 18.34664154 |
| 21.67272186 | 21.4863205 | 21.01990891 |
| 20.16250992 | 20.22646523 | 19.83176994 |
| 22.67210579 | 23.20791245 | 23.44547081 |
| 22.39128876 | 22.57433701 | 21.77210236 |
| 20.89427948 | 20.81285286 | 21.02849007 |
| 18.7755146 | 19.10103989 | 18.36517334 |
| 19.87996864 | 19.78434372 | 19.16993332 |
| 18.17476082 | 18.24714088 | 17.95613861 |
| 22.94231415 | 22.46026421 | 22.07708931 |
| 21.15114403 | 21.41794777 | 21.94021606 |
| 21.15236664 | 22.0212822 | 21.74732208 |
| 16.49722481 | 17.26841927 | 16.83346367 |
| 19.99661827 | 20.03444099 | 19.85777092 |
| 18.90737534 | 18.88746643 | 18.93382454 |
| 22.83661652 | 22.70488167 | 22.3091507 |
| 19.39840889 | 18.7507782 | 18.66968346 |
| 21.1041851 | 21.40906715 | 20.75728607 |
| 23.98560333 | 23.58833122 | 24.46982384 |
| 23.60821342 | 23.59908485 | 23.22566223 |
| 19.28484917 | 19.05434799 | 19.24927711 |
| 17.78338814 | 17.64934921 | 17.81001282 |
| 19.429739 | 20.26500702 | 19.98603821 |
| 23.17983055 | 22.81696892 | 22.9706707 |
| 21.48654747 | 20.89203644 | 21.78314781 |
| 23.42456818 | 22.72771835 | 23.6630249 |
| 23.63923073 | 24.07723808 | 23.1892128 |
| 23.31171227 | 23.79735756 | 23.14261818 |
| 19.34193611 | 18.54889679 | 19.07433701 |
| 25.37849426 | 25.33390808 | 24.99635887 |
| 24.83213806 | 24.743433 | 25.61209679 |
| 20.44210052 | 19.9653492 | 19.66933632 |

| | | |
|---|---|---|
| 24.4208622 | 24.49025536 | 24.22188759 |
| 23.83489037 | 23.52776527 | 23.77959251 |
| 20.76577568 | 21.22543144 | 20.65337944 |
| 22.6498909 | 22.4862709 | 21.95912933 |
| 22.57101059 | 22.69706726 | 22.00748444 |
| 20.38394737 | 20.48357391 | 20.00372887 |
| 17.46605873 | 17.54420662 | 17.65619278 |
| 20.84523773 | 20.85823631 | 21.61414719 |
| 20.87718582 | 20.75240326 | 20.21075821 |
| 21.94268036 | 22.09933662 | 22.44142151 |
| 20.4056015 | 20.03859901 | 19.79938889 |
| 17.73226738 | 17.85946846 | 17.50439835 |
| 19.65009689 | 19.98173523 | 20.13649178 |
| 21.72851372 | 21.80556297 | 21.66329002 |
| 19.0131321 | 18.78074265 | 18.46914291 |
| 20.46736336 | 20.29273987 | 20.07513237 |
| 21.33054543 | 20.91466331 | 21.57721329 |
| 20.65306854 | 19.95477676 | 20.40287018 |
| 24.0789299 | 23.73604012 | 24.47945023 |
| 22.5438385 | 22.31254959 | 22.29573631 |
| 21.72465897 | 21.60501099 | 20.9242878 |
| 25.04413986 | 25.17133713 | 25.29918861 |
| 22.89625168 | 23.4576683 | 23.00535011 |
| 22.65509796 | 22.49759865 | 23.32341003 |
| 21.91399384 | 21.90372086 | 22.47484589 |
| 22.0917778 | 21.96569824 | 21.45300674 |
| 20.16049576 | 20.96433067 | 20.58668518 |
| 18.50314331 | 17.95868874 | 18.41062355 |
| 23.06202698 | 22.66960144 | 22.28243828 |
| 22.98921585 | 22.63236618 | 23.23037529 |
| 23.04387093 | 22.65305328 | 22.11206627 |
| 20.55918503 | 20.51252174 | 20.32771683 |
| 21.34965897 | 20.78466034 | 21.5653019 |
| 21.07502937 | 20.26124954 | 20.93741035 |
| 21.46372414 | 21.22924805 | 21.24759102 |
| 20.64071274 | 20.92452049 | 20.87102509 |
| 20.24303818 | 20.40478897 | 19.97518921 |
| 19.58952713 | 19.64279556 | 19.71871376 |
| 21.05912781 | 21.19949341 | 20.6055336 |
| 22.76067924 | 22.71286392 | 22.67300606 |
| 20.5077076 | 20.31798935 | 20.63180923 |
| 21.25915718 | 21.50568771 | 20.89508438 |
| 22.75620461 | 22.75180435 | 23.4847393 |
| 23.38837242 | 23.57716751 | 22.75398827 |
| 23.87729645 | 23.89169502 | 24.38191032 |
| 24.14647865 | 24.22749138 | 24.13761711 |
| 22.03453827 | 22.04712868 | 22.41282082 |

| | | |
|---|---|---|
| 20.95866203 | 20.69564056 | 20.62247849 |
| 24.69673347 | 24.92010117 | 24.7520752 |
| 19.0832386 | 19.49917984 | 19.4140873 |
| 24.01977539 | 24.16909409 | 23.46162415 |
| 18.3712635 | 18.58652878 | 18.88456154 |
| 19.4864254 | 19.67370796 | 19.75928307 |
| 20.80062675 | 21.54896355 | 20.87852478 |
| 20.78089523 | 20.89667511 | 20.41601181 |
| 23.05315208 | 22.95278931 | 23.38379669 |
| 20.68699646 | 20.97161293 | 20.1919651 |
| 28.37593651 | 27.32284737 | 27.62188721 |
| 21.71664619 | 22.03036308 | 21.65392113 |
| 21.20158577 | 20.6391983 | 21.48493767 |
| 22.32744598 | 22.32555771 | 21.93302727 |
| 28.9775486 | 28.66695595 | 29.13352203 |
| 23.89994621 | 23.84637451 | 23.8006649 |
| 18.55366707 | 18.98661041 | 18.84932327 |
| 18.93759537 | 18.53765297 | 18.51124382 |
| 21.10042 | 21.8461113 | 21.55090141 |
| 23.09964752 | 23.09404755 | 22.48780823 |
| 21.0700779 | 21.03144836 | 21.1388588 |
| 24.34007072 | 24.25333023 | 24.78854942 |
| 28.4140625 | 28.16426277 | 28.17707825 |
| 19.03587914 | 19.07917023 | 18.46883202 |
| 20.40416145 | 20.57567406 | 20.52550697 |
| 24.13272285 | 24.06520462 | 23.4192543 |
| 25.62243843 | 24.99210167 | 25.55619431 |
| 18.76798439 | 18.67640877 | 18.87011528 |
| 18.78988647 | 18.78622437 | 18.82805252 |
| 21.86573601 | 21.52132607 | 21.35151291 |
| 21.91394043 | 22.10096359 | 22.19244766 |
| 22.03620148 | 22.04892159 | 22.44404984 |
| 23.50218582 | 22.78579903 | 23.30213165 |
| 22.87985992 | 22.81599998 | 22.98443985 |
| 18.24202919 | 18.27079391 | 18.19452286 |
| 20.09955406 | 20.01802254 | 20.24160957 |
| 22.7308197 | 23.13579369 | 23.63112259 |
| 16.78228188 | 16.97307968 | 16.6225853 |
| 19.8538723 | 19.99504089 | 20.52609062 |
| 20.75570679 | 20.76013947 | 20.91835022 |
| 21.25275421 | 21.53072739 | 21.98624802 |
| 21.59126854 | 21.54872704 | 21.68376923 |
| 22.07317924 | 21.79362679 | 22.55336571 |
| 21.90528297 | 21.80436707 | 22.31576157 |
| 19.6531086 | 19.83752632 | 19.24337769 |
| 22.82407951 | 23.26639366 | 22.6356945 |
| 22.46497536 | 22.97318459 | 22.40572166 |

| | | |
|---|---|---|
| 27.51143456 | 27.44860077 | 27.00212669 |
| 24.5160923 | 24.60507393 | 24.09916687 |
| 19.98354721 | 20.03728485 | 20.30433464 |
| 21.95723724 | 22.20372391 | 21.83621788 |
| 23.50775909 | 23.52569008 | 24.23433113 |
| 21.38071251 | 21.49862671 | 21.79474831 |
| 26.40056992 | 26.57227135 | 25.78423119 |
| 21.72224808 | 21.72237015 | 21.42769051 |
| 19.6502533 | 19.82018471 | 20.00680923 |
| 21.59538078 | 21.72056389 | 21.64110184 |
| 21.97793007 | 22.16021347 | 22.24420166 |
| 19.51473427 | 19.37978172 | 19.38536453 |
| 21.03115082 | 21.08938217 | 20.98552704 |
| 20.35174942 | 19.99333763 | 20.50683975 |
| 24.12210083 | 23.63636208 | 24.16872406 |
| 23.65440941 | 24.0150795 | 24.02713966 |
| 22.12796021 | 22.30436325 | 21.64165306 |
| 23.05679131 | 23.1618557 | 22.6299305 |
| 18.72901154 | 18.79588699 | 18.67442703 |
| 18.04891396 | 18.25432777 | 17.95279121 |
| 23.47375107 | 23.88729668 | 24.06271744 |
| 22.18856049 | 22.46371269 | 22.57347298 |
| 26.51429939 | 26.28509903 | 26.71343613 |
| 22.97616196 | 22.62663651 | 23.18243408 |
| 20.57400513 | 20.5506382 | 20.80761909 |
| 18.36614609 | 18.61220932 | 18.06763077 |
| 22.58364677 | 22.5440464 | 22.39505959 |
| 21.28848076 | 21.17261505 | 21.84463692 |
| 23.07154465 | 23.16061974 | 22.6951046 |
| 19.44623375 | 19.177948 | 19.04142952 |
| 19.05467033 | 18.78510284 | 18.96076202 |
| 22.52638626 | 22.21450996 | 22.28139687 |
| 25.87416267 | 26.03916168 | 25.41467667 |
| 20.70048332 | 20.31526947 | 20.2853241 |
| 21.70851898 | 21.47385979 | 20.99535179 |
| 20.01275635 | 19.79475212 | 19.57411385 |
| 23.90419579 | 23.95855713 | 23.80886269 |
| 19.13901138 | 18.93468475 | 19.27928162 |
| 18.75702667 | 19.08801842 | 19.14351273 |
| 27.23376465 | 27.15248871 | 27.7428112 |
| 20.73404884 | 20.7677784 | 20.14885712 |
| 21.23435211 | 21.21138191 | 21.54391479 |
| 21.71648979 | 21.53767014 | 21.23266411 |
| 20.12246704 | 20.12328529 | 19.71544647 |
| 23.95145607 | 23.74685287 | 23.32653618 |
| 25.34104729 | 25.8733902 | 25.81212234 |
| 24.95654869 | 24.4440937 | 24.4003849 |

| | | |
|---|---|---|
| 21.11192703 | 21.26655579 | 21.48704338 |
| 23.37054443 | 23.8129921 | 23.7667923 |
| 20.45672417 | 20.3750248 | 20.79286194 |
| 20.40595627 | 20.60144234 | 20.97454643 |
| 20.52145767 | 20.61525536 | 20.46268082 |
| 19.43476868 | 19.28560066 | 19.48816109 |
| 24.53340149 | 24.56482697 | 23.87303543 |
| 22.52038574 | 23.00114441 | 22.77087021 |
| 25.12423515 | 25.4932251 | 25.10923767 |
| 22.82756615 | 23.11309433 | 23.33081055 |
| 18.66736984 | 18.40793991 | 18.88854218 |
| 22.72181511 | 22.43371391 | 22.35638237 |
| 22.21166229 | 22.5576725 | 22.40053368 |
| 20.78355026 | 20.85475349 | 21.1298275 |
| 20.18095207 | 20.37047577 | 20.08654404 |
| 23.86372948 | 23.90781975 | 23.37923431 |
| 23.08077812 | 22.92251015 | 23.59602165 |
| 22.08472824 | 22.16545105 | 21.90409279 |
| 23.32906151 | 23.78323174 | 23.34193039 |
| 24.26838875 | 24.22184753 | 24.6543808 |
| 19.07688713 | 18.71935844 | 19.02535057 |
| 23.35484314 | 23.40122223 | 23.06667519 |
| 21.26364326 | 21.42670441 | 21.62425423 |
| 19.0392971 | 18.40859795 | 18.82888603 |
| 22.99995995 | 23.52812576 | 22.97609711 |
| 23.04216194 | 22.30865097 | 22.71439552 |
| 24.93318558 | 24.50326157 | 24.66839027 |
| 19.92072868 | 19.66232109 | 19.73254395 |
| 18.88288689 | 18.57344055 | 18.39935684 |
| 19.54058647 | 19.50636101 | 19.30014801 |
| 23.68550873 | 23.46997452 | 23.24487114 |
| 30.30971527 | 30.34897614 | 30.67851448 |
| 23.96336555 | 24.29832077 | 23.90482712 |
| 21.41745758 | 21.82661438 | 21.8155117 |
| 20.77214432 | 20.69728088 | 20.50123596 |
| 23.19050789 | 23.55080414 | 23.31982422 |
| 20.859375 | 20.3507309 | 20.40292358 |
| 20.20458984 | 20.27363586 | 20.12226486 |
| 23.60560608 | 23.07249451 | 23.46588135 |
| 19.11230087 | 18.68795204 | 19.32547951 |
| 18.48007393 | 18.34735107 | 18.06306076 |
| 23.82686806 | 24.09389687 | 24.3801403 |
| 22.08574677 | 22.11675453 | 22.07399368 |
| 19.68655968 | 19.40698242 | 19.35003853 |
| 22.0844326 | 22.13850212 | 21.6181488 |
| 21.35159302 | 21.20024872 | 21.01589394 |
| 22.80635071 | 22.35865974 | 22.91510963 |

| | | |
|---|---|---|
| 20.8054924 | 20.95334625 | 21.33389664 |
| 22.31864357 | 22.6414299 | 22.0528183 |
| 24.34136009 | 23.94380188 | 23.66014481 |
| 22.6486721 | 22.94494438 | 22.94555092 |
| 23.97991753 | 23.89327621 | 23.62353706 |
| 21.3016777 | 21.13046455 | 20.85088539 |
| 21.08737373 | 21.57569122 | 21.01142693 |
| 22.46910286 | 22.43436241 | 22.12275124 |
| 22.69936752 | 23.00023079 | 22.28219223 |
| 23.08695984 | 23.10478783 | 23.33748245 |
| 27.37244415 | 27.29438782 | 27.55885696 |
| 20.87523842 | 20.96468353 | 20.79889488 |
| 24.40399933 | 24.29968071 | 23.74822617 |
| 21.79166985 | 21.93313599 | 21.745924 |
| 20.95995522 | 21.21916199 | 21.14411545 |
| 22.34231758 | 22.44461632 | 22.56870079 |
| 22.40235329 | 22.55222511 | 22.14629555 |
| 19.94372559 | 20.25843048 | 19.70859909 |
| 17.80594635 | 17.92620087 | 18.0694294 |
| 20.60645103 | 20.3768959 | 20.07943153 |
| 22.35124779 | 22.15624428 | 22.52019501 |
| 22.46946526 | 22.3819809 | 22.26103783 |
| 23.67720985 | 23.86961937 | 23.15039253 |
| 21.3565197 | 21.83085632 | 21.23641586 |
| 23.15170288 | 23.13412666 | 22.80613136 |
| 19.57722473 | 19.04118919 | 19.20889854 |
| 25.83338928 | 25.43417168 | 25.23176575 |
| 19.76074028 | 20.01278687 | 20.24575043 |
| 21.98542404 | 22.55263138 | 22.13290215 |
| 19.84295464 | 19.46756363 | 20.10475922 |
| 25.34659767 | 25.53012276 | 24.87735176 |
| 20.10334396 | 20.45079422 | 20.40126228 |
| 18.77596474 | 18.48028374 | 18.77237701 |
| 23.20677757 | 23.34696007 | 23.75635529 |
| 24.80746078 | 25.21171188 | 24.91861725 |
| 20.14891052 | 20.49368286 | 20.62852478 |
| 21.77697563 | 21.96799469 | 21.76040077 |
| 24.63309288 | 24.8809967 | 25.2492466 |
| 21.13111877 | 20.73843956 | 20.53193474 |
| 27.36182213 | 27.62411308 | 26.84233856 |
| 20.57954216 | 20.33263206 | 20.38770103 |
| 19.97257042 | 19.61639023 | 19.60998344 |
| 22.68673897 | 22.32009315 | 22.42527771 |
| 23.39083099 | 22.9426918 | 23.26615143 |
| 20.28907967 | 20.35449791 | 20.23537064 |
| 24.8975544 | 24.9858551 | 24.91850853 |
| 21.76988602 | 22.03655624 | 21.5357933 |

| | | |
|---|---|---|
| 23.909235 | 24.01912117 | 24.1937027 |
| 19.65379715 | 19.84301376 | 19.76602364 |
| 22.41412354 | 22.04256058 | 22.12380409 |
| 19.2868824 | 18.9926796 | 18.94142723 |
| 24.92131805 | 24.50476837 | 25.05298805 |
| 19.24008179 | 19.26929665 | 19.00631523 |
| 23.11970901 | 22.65754128 | 22.61626053 |
| 21.10417557 | 21.18105125 | 20.89476967 |
| 21.54871178 | 21.32293129 | 21.87518311 |
| 25.32236862 | 24.85688972 | 25.31466103 |
| 22.19080734 | 22.29081535 | 21.90393448 |
| 25.2973423 | 25.36312485 | 24.75646591 |
| 23.76619148 | 23.82056618 | 23.25494385 |
| 19.79815292 | 19.70475197 | 19.50607491 |
| 18.10383415 | 18.06598663 | 18.07447815 |
| 18.16010094 | 18.60956192 | 18.14562798 |
| 25.35641098 | 25.63075638 | 25.41547203 |
| 20.81197929 | 20.8549099 | 20.55986214 |
| 22.68670082 | 23.14193535 | 22.70648193 |
| 25.3043251 | 25.64419937 | 25.91982079 |
| 24.07263756 | 23.50871086 | 23.73839569 |
| 22.66277885 | 22.58315659 | 23.10194588 |
| 18.70582771 | 18.71023369 | 18.75478172 |
| 23.54000282 | 23.51488304 | 23.57677841 |
| 21.95639038 | 22.30981445 | 22.12854004 |
| 18.5525074 | 18.35799217 | 18.25157547 |
| 20.78970528 | 20.93449593 | 20.6509285 |
| 22.39203262 | 22.69859886 | 22.25689507 |
| 21.79728889 | 21.73674774 | 22.20763588 |
| 22.13119507 | 22.04027748 | 21.68241501 |
| 19.22196579 | 18.8914547 | 18.86496544 |
| 23.21780586 | 23.05367851 | 22.79487228 |
| 22.87690735 | 23.08311081 | 22.52360153 |
| 25.10115433 | 24.94577026 | 24.45377159 |
| 24.38943291 | 24.5603466 | 24.60231972 |
| 20.93120003 | 20.66484261 | 20.32688522 |
| 22.68276978 | 22.73595428 | 23.05391312 |
| 21.19557381 | 20.9652195 | 21.04022026 |
| 20.12178612 | 20.40407372 | 20.13642502 |
| 19.71243858 | 19.56117249 | 19.27292824 |
| 18.99246025 | 18.66607666 | 18.65224075 |
| 20.36668015 | 20.50775528 | 19.96245766 |
| 23.72908592 | 23.50205231 | 23.47553444 |
| 21.215868 | 21.10798073 | 21.51525497 |
| 24.89427185 | 24.92776299 | 24.44909859 |
| 21.48010445 | 22.08081627 | 21.96596527 |
| 18.81443977 | 19.02764702 | 18.50079346 |

| | | |
|---|---|---|
| 24.23642731 | 24.43825912 | 23.84963226 |
| 25.02343941 | 24.95718956 | 24.46187782 |
| 26.42553902 | 26.27175522 | 26.37336731 |
| 22.72916794 | 23.01695251 | 22.48088837 |
| 23.14978981 | 22.94559669 | 23.35717773 |
| 21.42478943 | 21.7384491 | 21.25583458 |
| 19.26100349 | 18.84604836 | 18.81310081 |
| 22.25616264 | 22.63783073 | 22.75377083 |
| 22.33498764 | 22.46829033 | 22.58614349 |
| 23.41333199 | 23.61168671 | 23.0673542 |
| 23.48298836 | 22.96748161 | 23.03347588 |
| 21.52802658 | 21.51669121 | 21.20944786 |
| 21.19770432 | 21.38424492 | 21.29043007 |
| 21.93920135 | 22.00040817 | 21.70781136 |
| 18.8094101 | 18.5557251 | 18.45825768 |
| 24.19869232 | 24.06722641 | 23.74060249 |
| 19.17673492 | 19.50362396 | 19.66769218 |
| 20.35107613 | 20.43719101 | 20.74338531 |
| 21.09362411 | 20.80377197 | 20.82833862 |
| 21.91064262 | 22.1354599 | 22.3435173 |
| 23.72604179 | 23.4711895 | 23.16281891 |
| 23.51515579 | 23.91005325 | 23.5143013 |
| 22.6321373 | 22.45581245 | 22.64435768 |
| 18.46177483 | 18.52909088 | 18.10160255 |
| 22.86393166 | 23.2947464 | 23.25840759 |
| 23.91910172 | 23.75709343 | 24.32922745 |
| 22.51213837 | 22.6172123 | 22.15738869 |
| 26.84973907 | 26.52943802 | 26.58467293 |
| 19.63181877 | 19.97705269 | 19.52093697 |
| 23.2472229 | 23.13850594 | 23.50658798 |
| 22.18882751 | 21.9039135 | 21.61481857 |
| 23.23266602 | 23.14570808 | 22.93640518 |
| 22.22275162 | 22.49447823 | 22.09011841 |
| 23.47650528 | 22.83987999 | 23.08630562 |
| 23.66295052 | 23.66657448 | 23.85851669 |
| 21.75535583 | 21.74112129 | 21.81240273 |
| 19.87537766 | 19.70594978 | 19.90223122 |
| 24.74497986 | 24.49672508 | 24.92728806 |
| 22.67679787 | 22.5935421 | 22.55641365 |
| 22.97286224 | 22.71152115 | 22.99856758 |
| 25.17962456 | 25.44331551 | 24.8626976 |
| 21.78459358 | 21.40638924 | 21.78613091 |
| 18.43280983 | 18.16128349 | 18.65463638 |
| 23.5579567 | 23.19844246 | 23.51166344 |
| 19.1876564 | 19.14018631 | 19.28359032 |
| 20.9048481 | 20.83910751 | 20.60816193 |
| 22.53037071 | 22.66755867 | 22.86942863 |

| | | |
|---|---|---|
| 22.4468956 | 22.48479843 | 22.17324829 |
| 26.30408859 | 26.41434097 | 26.98913193 |
| 24.88228035 | 24.29958344 | 24.66306496 |
| 24.36979675 | 24.30293274 | 24.77484703 |
| 24.31807518 | 24.08859062 | 24.15393257 |
| 19.25408554 | 19.00154686 | 18.81878662 |
| 24.01270866 | 23.87316895 | 24.09581184 |
| 20.23810768 | 20.35112762 | 20.23589134 |
| 24.67605209 | 24.5673275 | 24.45481873 |
| 22.31584358 | 22.27117348 | 22.17228699 |
| 22.00461769 | 21.95041656 | 21.89492416 |
| 21.10370064 | 21.18368721 | 21.29541969 |
| 20.81585503 | 20.73386574 | 20.46582222 |
| 26.5816555 | 26.94488144 | 26.97155952 |
| 21.0474968 | 20.92145157 | 20.87981415 |
| 25.72997665 | 26.01690102 | 25.6885643 |
| 24.28877449 | 24.85783577 | 24.72924042 |
| 21.79475784 | 21.64979362 | 21.42482948 |
| 21.54142189 | 21.74695396 | 21.93169594 |
| 23.55402184 | 23.87280273 | 23.51791 |
| 22.4800415 | 22.54733658 | 22.06393814 |
| 23.98174286 | 23.74511719 | 24.29951286 |
| 21.94133186 | 21.72662926 | 21.70638466 |
| 21.23642921 | 21.11240768 | 21.13542557 |
| 21.4533329 | 21.12057686 | 21.1166687 |
| 17.76190186 | 17.61177826 | 17.49819565 |
| 18.33247757 | 18.26717949 | 18.15712929 |
| 22.37133408 | 22.56859207 | 22.67647934 |
| 19.36194038 | 19.17032814 | 19.33493233 |
| 22.6346817 | 22.74345207 | 22.95599937 |
| 23.90194702 | 23.68741989 | 23.90101051 |
| 20.99915314 | 21.25881767 | 20.7447052 |
| 19.99673271 | 19.80692291 | 19.76581764 |
| 25.92440987 | 25.65155602 | 25.703125 |
| 19.93711281 | 19.89308929 | 19.74855804 |
| 23.00935173 | 23.11447144 | 22.76652908 |
| 25.47440529 | 25.91824532 | 25.54734039 |
| 21.36445808 | 21.30930138 | 21.2444973 |
| 25.08248138 | 24.84337234 | 25.46014977 |
| 22.32170105 | 22.46055984 | 22.06202316 |
| 19.4039669 | 19.54300499 | 19.09525681 |
| 23.74749184 | 23.82242012 | 24.08358192 |
| 23.20281982 | 23.10107231 | 23.30653 |
| 20.78215599 | 21.16871452 | 21.13096428 |
| 22.629879 | 22.86126328 | 22.37790298 |
| 25.2457695 | 25.1367836 | 25.56853104 |
| 23.12073708 | 23.01658249 | 22.90525436 |

| | | |
|---|---|---|
| 21.81271362 | 21.48430061 | 21.64370728 |
| 19.06688881 | 19.32116508 | 19.01439285 |
| 23.29564095 | 23.3839283 | 22.89837837 |
| 22.8611908 | 22.73504257 | 22.49209785 |
| 20.20864296 | 20.10627365 | 20.25602913 |
| 20.89725113 | 21.07910538 | 21.30601501 |
| 23.18251228 | 23.4633522 | 23.63572884 |
| 28.51870918 | 28.49324799 | 27.9247303 |
| 25.66106033 | 25.9709034 | 26.12345695 |
| 19.18313599 | 19.1608429 | 18.90953827 |
| 19.31071472 | 19.24425888 | 19.04616356 |
| 22.44929886 | 22.31580353 | 22.59161377 |
| 22.6755352 | 22.09274673 | 22.37843513 |
| 21.26973343 | 20.93668556 | 21.30784607 |
| 26.60026741 | 26.53240776 | 26.33826637 |
| 19.0376091 | 19.22818756 | 19.08761406 |
| 19.70603561 | 19.59477997 | 19.70893669 |
| 19.96494102 | 20.09676552 | 20.28775978 |
| 20.3108387 | 20.05418968 | 19.91684341 |
| 29.25717545 | 29.30948067 | 28.75690842 |
| 21.94248199 | 22.24160576 | 22.44786263 |
| 18.85933495 | 18.61162758 | 18.67790794 |
| 25.44027519 | 25.10746765 | 25.58733749 |
| 24.47865677 | 24.79636383 | 24.50349236 |
| 20.79173279 | 20.69560242 | 21.13352394 |
| 21.46051407 | 21.50211525 | 21.2530632 |
| 21.33855438 | 21.31316757 | 21.68878174 |
| 24.11066055 | 24.19217682 | 24.08835793 |
| 19.85000038 | 19.88606262 | 20.13050652 |
| 26.80349922 | 26.70973969 | 26.44798088 |
| 22.08650398 | 22.05985641 | 22.36761093 |
| 17.68441582 | 17.75514221 | 17.42963791 |
| 20.43545914 | 20.66453362 | 20.46100807 |
| 26.11599159 | 25.92549706 | 25.55090332 |
| 20.08661079 | 20.3436203 | 20.50432777 |
| 19.84311104 | 20.10365868 | 20.19701576 |
| 22.31066132 | 22.32365608 | 22.21146774 |
| 21.89053154 | 22.05319214 | 21.80477524 |
| 22.73506737 | 22.50062943 | 22.91601372 |
| 18.47592735 | 18.60194969 | 18.49353409 |
| 23.12249565 | 23.32623482 | 23.50811005 |
| 19.6701622 | 19.23566246 | 19.3925724 |
| 22.6266346 | 22.62336159 | 22.41015434 |
| 24.60771751 | 24.43733406 | 24.21106339 |
| 23.50259018 | 23.53804398 | 23.86477089 |
| 24.78215981 | 24.75993156 | 24.37182617 |
| 23.49093819 | 23.72363853 | 23.77869797 |

| | | |
|---|---|---|
| 25.7730484 | 25.667696 | 25.53412628 |
| 21.58823395 | 21.37199593 | 21.0705204 |
| 21.4839344 | 21.29541206 | 21.50172806 |
| 25.7650547 | 25.98683739 | 25.42262459 |
| 24.58515549 | 24.60863113 | 24.48675156 |
| 21.07705307 | 21.17024422 | 21.09832764 |
| 21.44333649 | 21.27530479 | 21.61777306 |
| 24.11080933 | 24.22936058 | 24.30343056 |
| 24.6474781 | 24.87124062 | 24.71516418 |
| 26.13702965 | 26.25639343 | 25.80503845 |
| 19.59213448 | 19.72173691 | 19.48693085 |
| 24.22582245 | 24.21378136 | 23.76307297 |
| 25.48106384 | 25.55129242 | 25.28145981 |
| 22.76517487 | 23.11976433 | 23.17739677 |
| 22.97361183 | 22.56306267 | 22.58845139 |
| 19.03269768 | 19.00246811 | 19.17722702 |
| 20.8942318 | 20.68232918 | 20.69781303 |
| 21.65845108 | 21.77636719 | 21.90466118 |
| 23.33014679 | 23.28536987 | 23.15480042 |
| 28.55148506 | 28.6172924 | 28.83973122 |
| 24.2889328 | 23.69283295 | 23.99314499 |
| 23.93183517 | 24.04536057 | 23.72336388 |
| 22.58702469 | 22.99073982 | 22.78457832 |
| 23.7936573 | 23.98494148 | 23.63802719 |
| 24.13877487 | 23.82222939 | 23.83060837 |
| 19.07814026 | 19.16833687 | 19.40337563 |
| 22.41960335 | 22.15617943 | 22.60811615 |
| 26.38960075 | 26.77563858 | 26.74923515 |
| 23.95306778 | 24.11828041 | 23.65851784 |
| 23.53930855 | 23.82768059 | 23.56765747 |
| 22.86525536 | 23.00802994 | 23.32511902 |
| 24.88524246 | 24.9026947 | 24.7812252 |
| 20.1637516 | 20.29083633 | 20.53202438 |
| 23.47512245 | 23.22680283 | 23.40932655 |
| 20.59115601 | 20.65263176 | 20.94563103 |
| 19.79265785 | 20.03239059 | 20.07613564 |
| 22.56561279 | 22.47935295 | 22.89896774 |
| 24.65715408 | 24.70210266 | 24.36408043 |
| 21.7019577 | 21.92209625 | 21.79468346 |
| 23.92350197 | 23.62957573 | 23.55740929 |
| 23.24619102 | 23.39967346 | 22.9581337 |
| 28.02111626 | 28.18790054 | 27.8680439 |
| 22.63915634 | 22.71505547 | 23.02809906 |
| 23.89420891 | 23.83094215 | 23.83174515 |
| 20.93406677 | 20.56838799 | 20.78517532 |
| 23.10072327 | 23.07225418 | 23.45002556 |
| 22.40026283 | 22.35083771 | 22.55697632 |

| | | |
|---|---|---|
| 29.60574532 | 29.57042694 | 29.50323105 |
| 21.37315941 | 21.74699783 | 21.78368568 |
| 22.97654152 | 23.33898354 | 23.50112724 |
| 18.57307243 | 18.47596741 | 18.73579597 |
| 21.2689209 | 21.06390953 | 21.46528053 |
| 23.67284012 | 24.15049171 | 23.71168518 |
| 23.51933098 | 23.17709351 | 23.48193741 |
| 24.35957336 | 24.20110512 | 23.88555336 |
| 20.22774696 | 19.92918587 | 20.14622307 |
| 24.26721382 | 24.12029457 | 23.99728203 |
| 23.75111198 | 23.76617622 | 23.34253311 |
| 23.73400307 | 23.95723152 | 24.01903534 |
| 20.0035038 | 19.67020226 | 19.82156944 |
| 23.95129776 | 23.89682579 | 23.92827797 |
| 19.56166077 | 19.49880219 | 19.53610611 |
| 25.34716034 | 25.6160183 | 25.75465775 |
| 21.4492588 | 21.47400475 | 21.56767082 |
| 23.64448357 | 23.38192558 | 23.36707878 |
| 25.02734375 | 24.84173203 | 24.90256882 |
| 20.86776352 | 20.81298065 | 21.01984215 |
| 24.92026138 | 24.90451431 | 24.59765625 |
| 23.26864052 | 23.18994331 | 23.19294167 |
| 26.87633514 | 26.92336655 | 27.17074585 |
| 23.71736336 | 23.81500626 | 23.50986862 |
| 22.83863831 | 22.68911743 | 22.71778488 |
| 22.07933807 | 21.882267 | 22.14442635 |
| 25.13575172 | 24.64030838 | 24.81090546 |
| 25.1901207 | 25.17974091 | 25.40307045 |
| 23.87511444 | 23.85325623 | 23.83045006 |
| 25.19667816 | 25.60267067 | 25.08347702 |
| 20.40402985 | 20.37092972 | 20.21443558 |
| 22.12747955 | 22.21678734 | 21.97739029 |
| 22.86136436 | 22.71001053 | 22.64364624 |
| 25.25336266 | 25.42951775 | 25.04854393 |
| 21.73067665 | 21.47251129 | 21.46535492 |
| 24.29229736 | 24.12168121 | 24.67378044 |
| 23.79117584 | 24.14535522 | 23.62353134 |
| 21.25770187 | 21.16599274 | 21.27490807 |
| 22.51070976 | 22.79043388 | 22.86119652 |
| 27.94964981 | 28.28435898 | 28.46889877 |
| 22.2076664 | 21.92248344 | 22.23159218 |
| 21.87073326 | 22.11830139 | 22.17792702 |
| 22.10386467 | 22.41047668 | 22.19127846 |
| 20.12533569 | 20.20407486 | 19.86495781 |
| 25.41482735 | 25.66238022 | 25.4216671 |
| 23.39831924 | 23.60156059 | 23.68696404 |
| 24.61331558 | 24.70783043 | 24.56909561 |

| | | |
|---|---|---|
| 25.80291748 | 25.86876869 | 25.9686985 |
| 28.94597244 | 29.25168991 | 28.93244171 |
| 23.18682289 | 23.23768044 | 23.20981026 |
| 20.77614403 | 21.2026062 | 21.11933517 |
| 28.99197006 | 28.98579407 | 28.64837646 |
| 20.81907082 | 20.73883247 | 21.05974197 |
| 25.41294098 | 25.12469101 | 25.53448677 |
| 24.15785408 | 24.36768913 | 23.90725708 |
| 24.86276817 | 24.61256409 | 24.59494591 |
| 29.13934517 | 29.16501617 | 29.70269203 |
| 23.86773109 | 23.76315308 | 23.51795959 |
| 21.83730125 | 21.48949814 | 21.8450222 |
| 21.7538681 | 21.41043663 | 21.596138 |
| 25.77736282 | 25.92813873 | 25.65991592 |
| 23.78520775 | 23.44532776 | 23.71956635 |
| 20.65397835 | 20.68073463 | 20.62063408 |
| 24.11640358 | 24.1085968 | 24.09082603 |
| 20.33617783 | 20.4497242 | 20.46908188 |
| 23.17286682 | 22.72763634 | 22.92168045 |
| 22.3445034 | 22.22447395 | 22.2125988 |
| 24.88805199 | 25.05420113 | 24.76872063 |
| 24.93319321 | 24.94664764 | 25.3959198 |
| 29.17725754 | 28.86133766 | 28.69001389 |
| 24.64500618 | 24.58052635 | 24.50407028 |
| 22.78396416 | 22.90483475 | 23.15906334 |
| 20.88058281 | 20.81578827 | 20.54925728 |
| 18.46970177 | 18.53171158 | 18.68093109 |
| 22.37890434 | 22.27422523 | 21.93198586 |
| 24.18245125 | 24.20734406 | 24.0456028 |
| 21.59980202 | 21.38609314 | 21.6308136 |
| 24.67475128 | 24.41707802 | 24.42817497 |
| 24.82845497 | 24.67278099 | 25.07592583 |
| 24.71455574 | 24.65629959 | 24.93988609 |
| 24.09473801 | 23.96173859 | 24.03517342 |
| 24.60962677 | 24.56550026 | 24.82224655 |
| 25.67681503 | 25.64437675 | 25.27451706 |
| 25.35020256 | 25.29634476 | 25.22850227 |
| 25.43952179 | 25.32577324 | 25.12678719 |
| 20.8744812 | 20.90337753 | 20.96294212 |
| 23.99286842 | 24.23701477 | 23.88537407 |
| 23.64623451 | 23.65767288 | 23.71658897 |
| 23.38756561 | 23.81912231 | 23.4585247 |
| 25.91507339 | 26.46953392 | 26.18099403 |
| 21.25119209 | 21.12635994 | 21.23095512 |
| 26.36294746 | 26.27511787 | 26.5625 |
| 22.78799629 | 22.70533371 | 22.78988647 |
| 24.66191101 | 24.66278458 | 24.67862511 |

| | | |
|---|---|---|
| 20.13888741 | 20.42339516 | 20.37164688 |
| 20.64295769 | 20.52581596 | 20.62147522 |
| 24.88457298 | 24.91699982 | 24.7804203 |
| 22.43315506 | 22.24185562 | 22.24019241 |
| 21.48018074 | 21.56358528 | 21.37466812 |
| 28.47348595 | 28.09687805 | 28.65585327 |
| 25.82202339 | 25.97766876 | 26.20378685 |
| 25.69893074 | 25.90312576 | 25.6421814 |
| 23.38939095 | 23.40805817 | 23.07778358 |
| 23.64611244 | 23.41620445 | 23.66957855 |
| 21.66913033 | 21.75409889 | 21.40252304 |
| 20.9653492 | 20.77655983 | 20.76408577 |
| 25.66085052 | 25.4655571 | 25.35952759 |
| 23.02871513 | 22.82812119 | 23.04813957 |
| 21.42226028 | 21.25562477 | 21.62351799 |
| 27.2647934 | 27.39882851 | 27.69901085 |
| 22.79137802 | 23.10068512 | 23.16914177 |
| 24.51799583 | 24.44046021 | 24.39920425 |
| 23.85023308 | 23.57814407 | 23.93939209 |
| 22.95515823 | 22.91944504 | 22.7305584 |
| 20.03255844 | 19.98175049 | 19.93592834 |
| 22.18185043 | 21.93018913 | 21.870718 |
| 23.8777771 | 23.49637222 | 23.56349182 |
| 23.41143417 | 22.98130035 | 23.3861599 |
| 23.78513527 | 23.6973114 | 23.41435814 |
| 22.5952301 | 22.32664299 | 22.47305107 |
| 24.75480843 | 24.74132347 | 24.73066139 |
| 23.13868141 | 23.28606224 | 22.84693146 |
| 19.11585808 | 19.05352974 | 18.88660622 |
| 23.62944031 | 23.6897316 | 23.68902016 |
| 20.15262985 | 20.0368824 | 20.19282722 |
| 21.74322128 | 21.70350075 | 21.48080063 |
| 23.19194603 | 23.19933701 | 23.32139778 |
| 26.06329727 | 26.06410217 | 25.72813225 |
| 23.02720833 | 22.86805534 | 22.97871971 |
| 25.04411697 | 24.90819931 | 24.90692902 |
| 24.10149956 | 23.81559753 | 24.21593285 |
| 26.20685196 | 25.7045517 | 25.9730587 |
| 22.11901283 | 22.16540909 | 22.3787117 |
| 25.9942131 | 25.94411469 | 25.89216614 |
| 19.16479492 | 19.16860008 | 19.398983 |
| 21.24850273 | 21.4597435 | 21.48396301 |
| 24.11799812 | 24.01614952 | 23.75539017 |
| 26.16645241 | 26.08149719 | 25.74674797 |
| 26.13012505 | 26.05858994 | 25.99512482 |
| 21.30908775 | 21.28242302 | 21.2223053 |
| 28.39234734 | 28.37560272 | 28.24839211 |

| | | |
|---|---|---|
| 24.00064278 | 23.84486961 | 23.64394379 |
| 22.93114281 | 23.19989014 | 22.8752861 |
| 21.32536507 | 21.25692558 | 21.33523369 |
| 27.03430939 | 27.22857094 | 27.25270844 |
| 28.491045 | 28.52188301 | 28.09209633 |
| 24.41574669 | 24.353302 | 24.7856617 |
| 20.71254349 | 20.91103554 | 20.77568626 |
| 21.66131592 | 21.62164497 | 21.62770462 |
| 22.06646347 | 22.1639595 | 21.90183067 |
| 23.67229652 | 23.58127594 | 23.25035095 |
| 24.03417969 | 24.16479874 | 24.09318924 |
| 23.1250267 | 22.74082947 | 22.9496212 |
| 22.42722702 | 22.2596817 | 22.00225258 |
| 20.04661369 | 20.01467896 | 19.89333344 |
| 26.26597214 | 26.27264214 | 26.08974457 |
| 21.61283302 | 21.61351395 | 21.51504135 |
| 30.43334389 | 30.39131737 | 29.91981316 |
| 21.52232742 | 21.76872444 | 21.82844162 |
| 23.77358818 | 23.5942421 | 23.61322594 |
| 25.30915451 | 25.32383919 | 25.00706863 |
| 23.22470093 | 23.22784996 | 23.09053612 |
| 28.49713898 | 28.47412872 | 28.50865173 |
| 23.69249344 | 23.49927139 | 23.87722778 |
| 24.76964951 | 24.90932846 | 25.06199837 |
| 22.37642097 | 22.24209595 | 22.33315849 |
| 22.04195595 | 22.18256187 | 22.11190224 |
| 25.94279861 | 25.60124016 | 25.91636467 |
| 23.46193695 | 23.4123764 | 23.47363472 |
| 25.0553093 | 25.1985836 | 24.98435211 |
| 24.99441147 | 25.00014687 | 24.64862823 |
| 23.82547951 | 23.8631115 | 24.20794106 |
| 22.55296898 | 22.77435112 | 22.9230423 |
| 25.51826096 | 25.35437775 | 25.19068718 |
| 24.03498459 | 24.18248367 | 24.01103973 |
| 21.6511364 | 21.8182621 | 21.60347557 |
| 23.3947525 | 23.59694862 | 23.77960014 |
| 23.66533852 | 23.66145897 | 23.70624352 |
| 23.1687603 | 23.13949776 | 23.34449196 |
| 27.90315247 | 28.09454346 | 27.95618057 |
| 25.57522011 | 25.2894516 | 25.20539665 |
| 23.84043694 | 23.91544914 | 23.94847488 |
| 20.33980942 | 20.48826981 | 20.17137527 |
| 25.41461754 | 25.60858917 | 25.14813423 |
| 23.4517765 | 23.6661644 | 23.49629211 |
| 19.67245293 | 19.84425354 | 19.61573219 |
| 21.96995544 | 21.71572113 | 21.98850632 |
| 20.1551609 | 20.17335129 | 20.28660774 |

| | | |
|---|---|---|
| 23.74504089 | 23.76476097 | 23.39264107 |
| 22.26399422 | 22.25528717 | 22.4492836 |
| 25.90991211 | 25.65620232 | 26.04860687 |
| 24.11854553 | 24.39523888 | 24.21512985 |
| 21.01443481 | 21.12343788 | 20.91866493 |
| 23.44691086 | 23.4450798 | 23.29969978 |
| 23.95739555 | 23.85929489 | 23.63823891 |
| 26.34576988 | 26.07228851 | 26.03253555 |
| 28.28613281 | 28.2046032 | 28.41254807 |
| 18.23412895 | 18.19484329 | 18.10341644 |
| 24.4750061 | 24.4456768 | 24.54506302 |
| 23.24174309 | 23.06077576 | 23.4789505 |
| 23.7910614 | 23.95998192 | 23.95020485 |
| 21.84468269 | 22.0255127 | 21.84203529 |
| 24.20968246 | 23.9942894 | 23.82890129 |
| 22.33634186 | 22.41042137 | 22.30393219 |
| 26.21150208 | 26.38305473 | 26.02823448 |
| 22.30700111 | 22.61612701 | 22.67670822 |
| 25.04221535 | 24.97035789 | 25.18858719 |
| 23.78579712 | 23.89927673 | 23.99642563 |
| 26.96714211 | 26.8881588 | 27.05713844 |
| 24.07012558 | 24.19347382 | 24.17860985 |
| 25.04086876 | 24.97563362 | 24.63373756 |
| 24.27493668 | 24.14732552 | 23.8958931 |
| 21.90056419 | 22.01127052 | 22.04200745 |
| 25.75178337 | 25.62441063 | 25.48418999 |
| 25.18875694 | 25.02341843 | 24.90810585 |
| 22.01171494 | 21.95545006 | 21.90377998 |
| 22.16558647 | 22.25917053 | 22.06643867 |
| 23.6871109 | 23.65548706 | 23.79354858 |
| 23.43247986 | 23.42819023 | 23.57125664 |
| 19.12205315 | 19.167799 | 19.18663025 |
| 24.6005764 | 24.6123333 | 24.61850739 |
| 25.61657715 | 25.55158997 | 25.2141304 |
| 20.69633484 | 20.56485939 | 20.68964195 |
| 21.12708855 | 21.24411964 | 21.28468513 |
| 27.24921417 | 27.09837341 | 27.32781601 |
| 25.44737053 | 25.41098785 | 25.03844643 |
| 21.60813141 | 21.50071907 | 21.57457733 |
| 25.13715363 | 25.37407684 | 25.2401123 |
| 25.7648983 | 25.55443382 | 25.44544792 |
| 20.9832325 | 21.25488281 | 20.89942169 |
| 27.46225357 | 27.37881851 | 27.10508537 |
| 23.51630783 | 23.30758095 | 23.41483498 |
| 24.95681953 | 25.00594139 | 24.73250771 |
| 20.69831467 | 20.80019951 | 20.7933445 |
| 28.30336761 | 28.34190559 | 28.36367035 |

| | | |
|---|---|---|
| 23.32538033 | 23.38771057 | 23.06977844 |
| 22.43359947 | 22.18461037 | 22.20962906 |
| 26.99552155 | 26.89799881 | 26.87381935 |
| 24.77380371 | 25.12821007 | 24.8038578 |
| 24.55092239 | 24.37555122 | 24.29954338 |
| 22.95824242 | 23.23745537 | 22.95551491 |
| 19.5713501 | 19.49354553 | 19.55224991 |
| 26.58627319 | 26.83527946 | 26.80360031 |
| 21.34301186 | 21.22469521 | 21.47690773 |
| 26.19641113 | 25.83720207 | 25.94865036 |
| 19.38752365 | 19.1822567 | 19.30731392 |
| 27.71704483 | 27.67071915 | 27.45043564 |
| 21.32272148 | 21.24316597 | 21.0671463 |
| 25.83908272 | 25.76813126 | 25.55006981 |
| 23.8384304 | 24.00134277 | 23.72307014 |
| 22.42970276 | 22.78699875 | 22.62733078 |
| 24.19992256 | 24.06004333 | 24.46645546 |
| 22.8219986 | 23.04491234 | 22.86107826 |
| 25.18702888 | 25.30029488 | 24.9470005 |
| 23.01673317 | 22.81151962 | 22.82231522 |
| 26.5403347 | 26.45218086 | 26.75597 |
| 21.57311058 | 21.69620132 | 21.72301483 |
| 24.58734322 | 24.43258858 | 24.47476959 |
| 20.38744736 | 20.33513069 | 20.21539116 |
| 21.38784409 | 21.24292374 | 21.02323723 |
| 23.88660049 | 23.8697319 | 23.64198685 |
| 25.25330162 | 25.37837982 | 25.10734177 |
| 22.92010117 | 22.80418015 | 22.7443676 |
| 24.30309296 | 24.31568718 | 24.06136131 |
| 21.11129761 | 21.22432709 | 21.15267372 |
| 25.4664402 | 25.45721436 | 25.74814224 |
| 27.2772274 | 27.43654251 | 27.04942322 |
| 25.82051277 | 25.7735405 | 25.91631317 |
| 22.98847389 | 22.89169312 | 22.78951263 |
| 21.78253555 | 21.8174572 | 21.84791946 |
| 21.55164719 | 21.69742012 | 21.35150337 |
| 19.88997841 | 19.74595261 | 19.65444183 |
| 26.50207329 | 26.4088192 | 26.48900223 |
| 24.49250031 | 24.51408195 | 24.61813354 |
| 25.62734985 | 25.64832115 | 25.35367775 |
| 24.66854095 | 24.73303604 | 24.64356613 |
| 19.88290405 | 19.85013199 | 19.93859673 |
| 28.71956444 | 28.67649841 | 28.41043091 |
| 25.03173828 | 25.10642815 | 24.86164856 |
| 28.63824081 | 28.66494179 | 28.29780579 |
| 22.69121361 | 22.97719765 | 22.65335274 |
| 24.53385925 | 24.50097084 | 24.42029572 |

| | | |
|---|---|---|
| 22.07201576 | 22.28389549 | 22.11099434 |
| 24.81350899 | 24.80428314 | 24.70451927 |
| 24.66611099 | 24.41355896 | 24.65252876 |
| 23.43227768 | 23.49053574 | 23.7647171 |
| 24.04290199 | 23.85955238 | 23.77135468 |
| 25.27346611 | 25.25893974 | 25.04755783 |
| 26.11007881 | 26.14038086 | 26.43616104 |
| 28.48867226 | 28.26831245 | 28.08917046 |
| 24.7742939 | 24.6298542 | 24.68035889 |
| 23.72676468 | 23.74036217 | 23.54621696 |
| 25.22889519 | 25.35339737 | 25.45581245 |
| 22.89537621 | 22.76527786 | 22.62052345 |
| 26.02433395 | 26.13022804 | 26.05498695 |
| 23.51213264 | 23.63764572 | 23.38172913 |
| 22.79493141 | 22.83385658 | 22.98921967 |
| 22.25229263 | 22.25301933 | 22.5040226 |
| 26.52903366 | 26.49018669 | 26.37566566 |
| 21.04404449 | 21.15087128 | 20.94138336 |
| 26.17336082 | 26.12887573 | 26.02724457 |
| 24.07786179 | 24.24582481 | 24.12311935 |
| 23.505476 | 23.44444847 | 23.3923378 |
| 22.88110733 | 23.17036819 | 23.0688591 |
| 28.41958618 | 28.30005646 | 28.23085785 |
| 27.25606155 | 27.26166916 | 27.38407707 |
| 25.3018589 | 25.21587181 | 25.08478737 |
| 24.71702957 | 24.54317856 | 24.40753365 |
| 22.99321365 | 22.89388657 | 22.72334671 |
| 21.473526 | 21.48549271 | 21.67795372 |
| 25.66772079 | 25.77111053 | 25.98547363 |
| 24.17436028 | 23.99478722 | 23.88511658 |
| 22.75910759 | 22.70440483 | 22.95341301 |
| 22.92622566 | 22.83248901 | 22.83113289 |
| 23.14488792 | 22.93523407 | 22.87486839 |
| 23.07671356 | 23.15015221 | 23.07747078 |
| 24.63751793 | 24.57449532 | 24.58125877 |
| 20.32294083 | 20.18436623 | 20.24223709 |
| 28.6170311 | 28.57017136 | 28.62251282 |
| 23.84047127 | 23.91716766 | 23.82490349 |
| 25.10825539 | 25.18401146 | 25.29954147 |
| 23.17150879 | 23.10378838 | 23.15718269 |
| 25.07185364 | 25.09342957 | 24.84867096 |
| 24.62022018 | 24.49536133 | 24.56197357 |
| 23.5825119 | 23.58589363 | 23.76328087 |
| 25.38302231 | 25.3523941 | 25.09785652 |
| 22.69022942 | 22.82874107 | 22.70874786 |
| 22.42396927 | 22.47494698 | 22.28833389 |
| 25.44774055 | 25.17213249 | 25.45834541 |

| | | |
|---|---|---|
| 30.27903366 | 30.35781479 | 30.35758018 |
| 18.28334427 | 18.19790077 | 18.40818214 |
| 27.32309151 | 27.3488903 | 27.45505524 |
| 26.01067543 | 25.94473267 | 25.81093597 |
| 27.38394928 | 27.28949356 | 27.31072807 |
| 21.79686356 | 21.78733635 | 21.76317406 |
| 23.31807709 | 23.24204636 | 23.41403008 |
| 20.28600693 | 20.13852692 | 20.20470619 |
| 23.60475159 | 23.70302582 | 23.84408188 |
| 23.89863586 | 23.86061668 | 23.81743813 |
| 24.42192841 | 24.39421844 | 24.66892433 |
| 25.14468956 | 25.29598427 | 25.13817406 |
| 26.35437393 | 26.30673409 | 26.37789345 |
| 20.76073456 | 20.74705124 | 20.57384682 |
| 21.01156998 | 21.12518311 | 21.09744263 |
| 24.75748062 | 24.45838165 | 24.80315208 |
| 26.22587585 | 26.25138092 | 26.37383652 |
| 20.89834023 | 21.06432152 | 20.90390205 |
| 29.07265282 | 28.73620224 | 29.10424614 |
| 20.96031761 | 21.07038498 | 20.88059616 |
| 24.74278831 | 24.82188034 | 24.83953476 |
| 25.91957283 | 25.89546394 | 25.82848167 |
| 23.97367477 | 23.94561577 | 23.8346405 |
| 24.616539 | 24.67281342 | 24.95073509 |
| 23.64824295 | 23.64593506 | 23.44747162 |
| 23.60763359 | 23.60169983 | 23.42272568 |
| 26.69708824 | 26.80107117 | 26.64170456 |
| 25.07580566 | 25.31132889 | 24.97044373 |
| 27.5948391 | 27.5116539 | 27.78148651 |
| 28.54123306 | 28.55467033 | 28.48112679 |
| 23.63753319 | 23.63502502 | 23.66727638 |
| 24.94462204 | 24.994627 | 25.05693245 |
| 28.46243095 | 28.53024101 | 28.3979702 |
| 25.49435234 | 25.70454025 | 25.3321991 |
| 25.04068565 | 24.70824623 | 24.87914085 |
| 24.7467804 | 24.89809608 | 24.64993858 |
| 22.05089188 | 21.85202217 | 21.97558212 |
| 24.27827644 | 24.45037842 | 24.14365196 |
| 24.7786808 | 24.94333076 | 25.11264229 |
| 24.60525131 | 24.53166771 | 24.70092773 |
| 24.08491516 | 23.93631363 | 24.11859322 |
| 23.68850899 | 23.791996 | 23.49876785 |
| 24.57165146 | 24.32718277 | 24.32122803 |
| 22.10570335 | 22.01012039 | 22.05098724 |
| 23.46304703 | 23.3905735 | 23.25442886 |
| 26.17956924 | 26.17135429 | 26.04247475 |
| 24.01678276 | 24.06643105 | 24.2226963 |

| | | |
|---|---|---|
| 21.70250702 | 21.52634621 | 21.83124161 |
| 26.66602707 | 26.77268028 | 26.66775894 |
| 18.97830391 | 18.7720108 | 18.86155319 |
| 23.66970634 | 23.47659874 | 23.53359032 |
| 26.55304527 | 26.24518967 | 26.32985687 |
| 21.98674774 | 21.91526604 | 21.73914719 |
| 25.92657661 | 26.04950714 | 25.72537613 |
| 27.16244125 | 26.91638947 | 27.29151344 |
| 24.89954567 | 24.84746361 | 24.66396713 |
| 19.45820618 | 19.4036808 | 19.49536514 |
| 29.11136627 | 29.24388504 | 29.40274048 |
| 26.25470543 | 26.25044823 | 26.35738564 |
| 26.63024139 | 26.81851006 | 26.57974243 |
| 24.74037933 | 24.71216011 | 24.42359734 |
| 25.64760208 | 25.90167046 | 25.79115486 |
| 26.77361488 | 26.64187431 | 26.57131958 |
| 21.92702103 | 22.0132103 | 22.01632118 |
| 26.02877045 | 26.03159142 | 25.80599594 |
| 25.83576775 | 25.74483871 | 25.61115456 |
| 24.7885704 | 24.69886971 | 24.48281097 |
| 26.01774216 | 25.82335854 | 25.91326141 |
| 23.88155746 | 23.71051407 | 23.59649086 |
| 24.35180855 | 24.25741386 | 24.254282 |
| 22.72417068 | 22.83118057 | 22.77108192 |
| 24.97340012 | 24.97260475 | 24.85936356 |
| 25.58763695 | 25.5801487 | 25.27928162 |
| 23.88646889 | 23.62055588 | 23.88437843 |
| 28.55202484 | 28.63646126 | 28.35380363 |
| 22.86925888 | 22.94942856 | 22.87955666 |
| 25.82157326 | 25.48230362 | 25.57796669 |
| 22.21201706 | 22.49201202 | 22.33803749 |
| 25.10264206 | 25.16559601 | 25.10036659 |
| 27.99509811 | 27.70367813 | 27.7793026 |
| 23.53907013 | 23.5138855 | 23.29752922 |
| 24.56540871 | 24.48135948 | 24.72210503 |
| 24.90151787 | 24.86327171 | 24.72336769 |
| 27.66983414 | 27.63191223 | 27.37710762 |
| 22.06162834 | 22.06238747 | 21.89486694 |
| 27.51595497 | 27.53291702 | 27.72074318 |
| 26.11218834 | 26.08909607 | 25.9100666 |
| 27.31916428 | 27.28717804 | 27.44281578 |
| 21.84402657 | 21.74431419 | 21.92794418 |
| 25.40690613 | 25.38018608 | 25.13923454 |
| 20.35764694 | 20.33912849 | 20.46778488 |
| 24.05241966 | 24.01234627 | 23.8744812 |
| 21.96913528 | 21.73759079 | 21.86538124 |
| 22.15274429 | 22.27166748 | 22.36971283 |

| | | |
|---|---|---|
| 24.77918243 | 24.7226696 | 24.62435722 |
| 24.62751007 | 24.54685211 | 24.5457077 |
| 27.44962692 | 27.78448486 | 27.57073784 |
| 23.6654377 | 23.81946945 | 23.63215446 |
| 22.13642883 | 21.92408752 | 22.05967522 |
| 25.53415108 | 25.43070984 | 25.35718727 |
| 28.5716629 | 28.70998955 | 28.8097477 |
| 21.03693771 | 21.23029709 | 21.15537262 |
| 26.13660622 | 26.1861248 | 26.04343987 |
| 23.20376778 | 23.47369957 | 23.3301487 |
| 25.38660812 | 25.54046059 | 25.70599747 |
| 19.63675499 | 19.69884109 | 19.74243736 |
| 26.82552719 | 26.67793655 | 26.53803062 |
| 28.26643372 | 28.20414543 | 28.49230385 |
| 23.95803261 | 23.94686317 | 23.89934349 |
| 27.50547028 | 27.38899422 | 27.34248161 |
| 28.38654518 | 28.3783474 | 28.71188354 |
| 25.51784897 | 25.55722427 | 25.59278297 |
| 25.00813484 | 25.07921982 | 24.93244934 |
| 26.00882912 | 25.99034119 | 25.78468132 |
| 23.6557312 | 23.46147919 | 23.55221558 |
| 24.33191299 | 24.33624649 | 24.16586113 |
| 29.01011848 | 28.99838638 | 28.77866936 |
| 26.3776741 | 26.27671051 | 26.35734558 |
| 23.46383667 | 23.30909538 | 23.61742592 |
| 25.86361313 | 26.02383232 | 25.69797707 |
| 24.38757324 | 24.36944771 | 24.32584572 |
| 27.04293633 | 26.79597855 | 27.11782074 |
| 24.97688484 | 25.08656883 | 25.0267601 |
| 24.05116463 | 24.07922363 | 24.3230648 |
| 25.30996704 | 25.42499352 | 25.48522568 |
| 26.46246147 | 26.47229576 | 26.49031639 |
| 24.24043846 | 24.23392868 | 24.37751198 |
| 24.09558296 | 24.04096603 | 24.17376328 |
| 29.33810043 | 29.47012138 | 29.46456337 |
| 24.03569984 | 24.16539574 | 23.94924545 |
| 21.02968597 | 21.14965248 | 21.17629433 |
| 25.71310997 | 25.76434898 | 25.9117794 |
| 25.92601776 | 25.80680466 | 25.73296738 |
| 21.95238113 | 21.97934532 | 22.12168884 |
| 26.03778648 | 26.08200073 | 26.17973328 |
| 24.73887062 | 24.66521454 | 24.60438347 |
| 27.24352455 | 27.40263176 | 27.46617126 |
| 22.85385895 | 22.83260536 | 22.95196915 |
| 24.19203949 | 24.10012817 | 24.22659111 |
| 29.67630005 | 29.63301659 | 29.44732285 |
| 24.67191887 | 24.46210861 | 24.47957993 |

| | | |
|---|---|---|
| 24.35029221 | 24.45455933 | 24.20507622 |
| 25.36792374 | 25.44524384 | 25.45244026 |
| 24.90372467 | 24.66022301 | 24.75106239 |
| 24.95533562 | 24.7858181 | 24.84089851 |
| 21.07808495 | 21.09420967 | 20.95856094 |
| 23.73924065 | 23.70021248 | 23.72575188 |
| 23.4918232 | 23.41918945 | 23.42793465 |
| 32.80353165 | 32.7759819 | 33.12465286 |
| 21.62902069 | 21.7034359 | 21.50547028 |
| 28.76629829 | 28.99273491 | 28.8948555 |
| 25.89561844 | 25.71227074 | 25.69962692 |
| 24.27416611 | 24.18288231 | 24.05197525 |
| 23.63406372 | 23.81249619 | 23.70113564 |
| 28.78596497 | 28.75153732 | 28.78641701 |
| 24.58753586 | 24.58025742 | 24.38820076 |
| 26.67325974 | 26.65514183 | 26.72474098 |
| 26.43922997 | 26.17909241 | 26.4726162 |
| 22.89103317 | 22.7581501 | 22.69239044 |
| 27.11991501 | 27.05999947 | 27.14352989 |
| 24.86204529 | 24.94250679 | 24.80399895 |
| 22.54114532 | 22.45543098 | 22.54188728 |
| 27.31208992 | 27.11624527 | 27.28616333 |
| 21.98303223 | 21.91464806 | 21.93960762 |
| 27.6864872 | 27.68575287 | 27.64620781 |
| 25.67287445 | 25.8317852 | 25.66146851 |
| 22.26207161 | 22.2472744 | 22.14090729 |
| 24.14800835 | 24.12333107 | 23.96071434 |
| 25.48658371 | 25.46619606 | 25.22796059 |
| 28.65810204 | 28.56533813 | 28.62206459 |
| 27.54327583 | 27.49346161 | 27.40873146 |
| 29.40433502 | 29.29189301 | 29.05703545 |
| 28.78016663 | 28.66505623 | 28.51054382 |
| 24.41897392 | 24.25475883 | 24.43354034 |
| 27.16454887 | 27.26069641 | 27.43817902 |
| 22.72261429 | 22.49083328 | 22.64674377 |
| 25.96926689 | 26.08008385 | 25.81839371 |
| 23.52282143 | 23.61906433 | 23.5757885 |
| 25.24392509 | 25.30276871 | 25.50814247 |
| 32.05003357 | 32.15156937 | 32.23144913 |
| 22.05308342 | 21.81526947 | 22.02607727 |
| 24.34794044 | 24.26768875 | 24.41731453 |
| 29.09313965 | 29.14876556 | 29.13071823 |
| 23.52060318 | 23.33905029 | 23.30392456 |
| 25.42320824 | 25.3951416 | 25.61272049 |
| 27.14968109 | 26.99431992 | 26.93336678 |
| 25.94784546 | 25.75716209 | 25.72458839 |
| 24.73511124 | 24.87034798 | 24.89024353 |

| | | |
|---|---|---|
| 23.69410515 | 23.53981018 | 23.4684391 |
| 27.70711136 | 27.62547302 | 27.4414711 |
| 27.48855019 | 27.58190346 | 27.38254738 |
| 23.76403427 | 23.63693047 | 23.69436646 |
| 24.13539314 | 24.12009048 | 24.24238014 |
| 24.58911514 | 24.45854187 | 24.61785507 |
| 19.58693314 | 19.72067261 | 19.62755203 |
| 27.55540466 | 27.44715309 | 27.31859016 |
| 30.82801628 | 30.59034729 | 30.78244972 |
| 26.91096497 | 26.72369385 | 26.97768974 |
| 25.92188263 | 25.7383461 | 25.7851696 |
| 24.8517952 | 24.78798103 | 24.89119339 |
| 27.9885006 | 27.93034744 | 27.75054741 |
| 19.10387611 | 19.09637833 | 19.2818718 |
| 19.95615387 | 19.93783569 | 19.8231144 |
| 26.11293602 | 25.94478035 | 26.01848602 |
| 26.48387909 | 26.3571949 | 26.64387512 |
| 23.68017769 | 23.69553566 | 23.65393829 |
| 26.05142784 | 25.9907608 | 25.87629318 |
| 23.78859711 | 23.79555702 | 23.7148037 |
| 24.38794899 | 24.36113739 | 24.36761856 |
| 24.74068642 | 24.7880249 | 24.79232597 |
| 27.3734169 | 27.2242527 | 27.17775726 |
| 22.39818573 | 22.41484451 | 22.3838253 |
| 24.25584412 | 24.1706543 | 24.24311829 |
| 23.82538986 | 23.89430046 | 23.65203476 |
| 19.93468475 | 20.02527046 | 19.85737228 |
| 24.2250824 | 24.17727661 | 24.41784477 |
| 26.60708618 | 26.61925507 | 26.39340591 |
| 23.24012947 | 23.2765274 | 23.1941185 |
| 26.64999771 | 26.65512085 | 26.46234894 |
| 26.10337257 | 25.99875069 | 26.16016006 |
| 25.02483559 | 25.02676773 | 24.88143921 |
| 23.50702286 | 23.61996078 | 23.60916328 |
| 25.18470001 | 25.22754478 | 25.35468674 |
| 25.30614662 | 25.0619297 | 25.2426281 |
| 24.08789444 | 24.21880913 | 24.27899933 |
| 27.04329872 | 26.94900131 | 27.02131844 |
| 25.55845451 | 25.48524857 | 25.57438469 |
| 25.14520836 | 24.96714783 | 25.07827187 |
| 25.97644043 | 25.93766403 | 25.89574623 |
| 24.49071693 | 24.55791473 | 24.51709747 |
| 25.28403091 | 25.21196747 | 25.26742363 |
| 26.34011269 | 26.33997154 | 26.36063194 |
| 24.87934494 | 24.68027878 | 24.77823257 |
| 27.19199944 | 27.04390526 | 26.99293709 |
| 19.71569443 | 19.61041832 | 19.65747833 |

| | | |
|---|---|---|
| 24.07164764 | 24.16851616 | 24.05018044 |
| 24.34012794 | 24.14518738 | 24.20589828 |
| 24.80949593 | 24.76659012 | 24.85463715 |
| 25.8410759 | 25.80084991 | 25.63737488 |
| 22.70061111 | 22.70168877 | 22.7023468 |
| 26.78738022 | 26.67916298 | 26.73301888 |
| 25.90130997 | 25.87454033 | 25.92471695 |
| 29.25461578 | 29.31790733 | 29.48971176 |
| 23.54181671 | 23.69194031 | 23.74453926 |
| 27.11075974 | 27.15929222 | 27.26108932 |
| 26.3849926 | 26.55559731 | 26.34501648 |
| 25.23356056 | 25.18321037 | 25.41949463 |
| 24.06355095 | 23.94141006 | 24.10928154 |
| 23.81224251 | 23.59239388 | 23.79300308 |
| 24.42498016 | 24.3091774 | 24.46411896 |
| 29.72264862 | 29.65567017 | 29.61097717 |
| 23.63719368 | 23.65983963 | 23.64207649 |
| 26.63072968 | 26.60108566 | 26.39432144 |
| 27.01222992 | 27.158041 | 26.88892746 |
| 19.7665329 | 19.73206711 | 19.67970848 |
| 25.0521183 | 25.13483047 | 25.12407494 |
| 26.44451523 | 26.34827995 | 26.28665543 |
| 30.35986137 | 30.48161316 | 30.58880806 |
| 25.80299377 | 25.96627808 | 25.75910187 |
| 26.64767647 | 26.51501465 | 26.54128838 |
| 25.59639931 | 25.44522667 | 25.59426308 |
| 24.09796906 | 24.21640205 | 24.15098381 |
| 23.68341064 | 23.79618645 | 23.62193108 |
| 27.45765495 | 27.39164162 | 27.40061951 |
| 24.93230438 | 24.96183968 | 25.09827423 |
| 23.30306816 | 23.18443108 | 23.25244904 |
| 24.96746635 | 24.90290833 | 24.8396759 |
| 29.84633064 | 29.68640137 | 29.75481796 |
| 28.09244728 | 28.20535851 | 28.23257828 |
| 29.77309227 | 29.72563362 | 29.54574776 |
| 26.78884315 | 26.8500576 | 26.83323479 |
| 24.06252098 | 23.96940231 | 23.87501526 |
| 26.70201683 | 26.65828705 | 26.75634384 |
| 26.05550194 | 25.99111366 | 25.91390991 |
| 25.29044533 | 25.36135292 | 25.49187851 |
| 25.15897942 | 25.21147919 | 25.03462982 |
| 24.32374763 | 24.24018097 | 24.39962387 |
| 28.17861748 | 28.23374748 | 28.14583778 |
| 30.02072525 | 30.0214138 | 29.88432693 |
| 27.25765419 | 27.19688988 | 27.06881523 |
| 20.72430229 | 20.61340332 | 20.66999435 |
| 29.42087936 | 29.33038521 | 29.26091194 |

| | | |
|---|---|---|
| 26.69707489 | 26.71800041 | 26.70869064 |
| 25.66488075 | 25.62174225 | 25.77196693 |
| 27.24489975 | 27.15495491 | 27.35262489 |
| 27.66060448 | 27.70796013 | 27.63851738 |
| 25.37239075 | 25.37146568 | 25.24144173 |
| 25.41955757 | 25.3843708 | 25.40819168 |
| 25.88208961 | 25.91043472 | 25.99636078 |
| 25.41750526 | 25.41640472 | 25.4190197 |
| 24.24966049 | 24.25670052 | 24.34448051 |
| 29.2011776 | 29.23806 | 29.30817604 |
| 24.42788696 | 24.41531372 | 24.33696747 |
| 25.56617165 | 25.54533577 | 25.604496 |
| 26.02603912 | 26.01824188 | 25.95051193 |
| 29.73408318 | 29.65973091 | 29.69253159 |
| 26.03793716 | 26.02098274 | 25.90677071 |
| 27.5473671 | 27.56369209 | 27.46330643 |
| 24.46170998 | 24.62787056 | 24.66090012 |
| 24.41080856 | 24.46511459 | 24.49711609 |
| 27.96101189 | 27.91556358 | 28.1015377 |
| 26.67301559 | 26.51790619 | 26.60075569 |
| 32.29265213 | 32.41690826 | 32.4610939 |
| 26.28980827 | 26.13464546 | 26.08720207 |
| 25.45611191 | 25.32481956 | 25.45145226 |
| 30.73686218 | 30.77458954 | 30.58682251 |
| 27.37233925 | 27.32105255 | 27.21956062 |
| 25.4072628 | 25.43527985 | 25.37068176 |
| 25.12865257 | 25.00581741 | 25.04034805 |
| 27.41565514 | 27.36937714 | 27.44800758 |
| 25.05004501 | 25.02769852 | 24.92864609 |
| 21.75212097 | 21.58746719 | 21.65830231 |
| 22.13876152 | 22.2371006 | 22.29811096 |
| 27.80633736 | 27.79479027 | 27.68548393 |
| 21.70701218 | 21.66548729 | 21.67207909 |
| 27.22128296 | 27.28032112 | 27.32789612 |
| 27.58236122 | 27.57230759 | 27.46642113 |
| 27.45562935 | 27.34146118 | 27.46044731 |
| 25.87307358 | 25.96585846 | 25.89057541 |
| 23.09283638 | 23.05003548 | 23.15826035 |
| 24.20709038 | 24.30870438 | 24.29988098 |
| 31.7195816 | 31.71626854 | 31.60939217 |
| 21.88971329 | 21.91020012 | 21.80163574 |
| 25.71619797 | 25.75238609 | 25.55550575 |
| 22.10259628 | 22.12086487 | 22.23561859 |
| 24.40065193 | 24.34894943 | 24.47541237 |
| 24.07176208 | 23.96746635 | 24.03122902 |
| 28.20587158 | 28.19699097 | 28.10139084 |
| 26.12700081 | 26.13261223 | 26.19376373 |

| | | |
|---|---|---|
| 28.79224205 | 28.87556076 | 28.89694023 |
| 28.54973221 | 28.56276131 | 28.45402908 |
| 26.24833679 | 26.24875069 | 26.13709259 |
| 25.57135201 | 25.49917221 | 25.55634689 |
| 23.8386116 | 23.77744484 | 23.81625175 |
| 30.14405441 | 29.95290565 | 30.03075409 |
| 25.06453705 | 24.98372269 | 25.00510025 |
| 28.83427238 | 28.62819099 | 28.73018265 |
| 26.06356049 | 25.9404335 | 26.1312027 |
| 24.92904663 | 24.79926491 | 24.9512043 |
| 27.00975037 | 26.95173264 | 26.94847107 |
| 25.00967789 | 25.08288765 | 25.14163208 |
| 31.74472427 | 31.75180626 | 31.71135139 |
| 29.91178322 | 29.98734665 | 30.00387001 |
| 28.38257027 | 28.31835175 | 28.4125576 |
| 23.65322685 | 23.64134407 | 23.58078766 |
| 24.4709053 | 24.49481392 | 24.59687805 |
| 25.78219223 | 25.79507637 | 25.76029968 |
| 24.57269669 | 24.41787338 | 24.4491272 |
| 26.71033669 | 26.67809868 | 26.69439507 |
| 24.20186234 | 24.27194214 | 24.10547638 |
| 27.1169014 | 27.21100235 | 27.06653595 |
| 23.20983315 | 23.0798111 | 23.17184258 |
| 27.31055069 | 27.17299843 | 27.1455574 |
| 29.30627441 | 29.25158691 | 29.28052139 |
| 24.8892765 | 24.89134979 | 24.78096962 |
| 30.28348923 | 30.37075615 | 30.36708832 |
| 27.06924248 | 27.04732895 | 26.96867371 |
| 22.86186409 | 22.95091057 | 22.95936012 |
| 26.13894844 | 26.27792358 | 26.114851 |
| 27.7143116 | 27.74855995 | 27.6610527 |
| 24.86851883 | 24.86222458 | 24.8887558 |
| 27.27588463 | 27.09526825 | 27.16733742 |
| 25.55168724 | 25.59927177 | 25.63920021 |
| 26.39723969 | 26.35787773 | 26.22988129 |
| 26.07384872 | 25.96689987 | 25.91384315 |
| 23.8227005 | 23.79168701 | 23.92365646 |
| 27.37419319 | 27.24048233 | 27.24575806 |
| 28.24860954 | 28.15525436 | 28.20052147 |
| 25.04924583 | 25.00699997 | 24.9632988 |
| 27.33631325 | 27.31079292 | 27.27655411 |
| 17.45148087 | 17.47882462 | 17.46471214 |
| 27.51965141 | 27.40421295 | 27.45631599 |
| 23.73454475 | 23.7750473 | 23.85015106 |
| 33.47204971 | 33.53274536 | 33.40261459 |
| 25.10891724 | 25.00383759 | 24.97751999 |
| 21.84375572 | 21.86700058 | 21.8178978 |

| | | |
|---|---|---|
| 26.86559486 | 26.75039482 | 26.73106956 |
| 27.77686119 | 27.75405502 | 27.73769569 |
| 26.02026749 | 26.01300621 | 26.08497047 |
| 32.08144379 | 31.97887993 | 31.98190498 |
| 25.51301956 | 25.43484688 | 25.44732475 |
| 28.2115078 | 28.18185425 | 28.14873123 |
| 31.07575417 | 31.05155945 | 30.9663868 |
| 25.78676224 | 25.80652046 | 25.73026657 |
| 31.64620972 | 31.67512512 | 31.53798294 |
| 24.92005539 | 24.87259483 | 24.86958694 |
| 25.03229523 | 25.03087997 | 24.94446564 |
| 26.62009621 | 26.59545708 | 26.73225021 |
| 23.39279556 | 23.44117355 | 23.35581779 |
| 29.9009285 | 29.93808174 | 29.86058998 |
| 28.5305481 | 28.46904945 | 28.53879356 |
| 29.02049065 | 29.01431274 | 29.1669178 |
| 26.62001228 | 26.62258148 | 26.75900841 |
| 29.38691139 | 29.31310272 | 29.43133926 |
| 29.99050903 | 29.9599781 | 29.93587875 |
| 24.20876884 | 24.09306717 | 24.14818764 |
| 24.93730354 | 24.86891556 | 24.88174057 |
| 28.6584034 | 28.70129967 | 28.69318771 |
| 29.97363281 | 29.90414047 | 29.92429733 |
| 27.42127419 | 27.4970932 | 27.4904995 |
| 24.66347694 | 24.58283043 | 24.68525505 |
| 28.22112274 | 28.25152588 | 28.2591362 |
| 23.21793556 | 23.33411407 | 23.28572464 |
| 28.35988808 | 28.31288719 | 28.37239647 |
| 27.44038582 | 27.34739876 | 27.38996124 |
| 25.42485428 | 25.33521652 | 25.45808029 |
| 27.06086922 | 27.06804657 | 27.15773582 |
| 25.22609711 | 25.24528122 | 25.14912987 |
| 24.28873634 | 24.26442909 | 24.21426964 |
| 24.12309074 | 24.10357285 | 24.05754089 |
| 31.0778141 | 31.07549095 | 31.10829926 |
| 27.93777466 | 28.02812004 | 27.97346306 |
| 29.16915703 | 29.20624924 | 29.07847404 |
| 26.63280678 | 26.55086517 | 26.58893585 |
| 23.30345345 | 23.2047863 | 23.22796631 |
| 28.33105278 | 28.32779694 | 28.37303162 |
| 27.08226776 | 26.97789574 | 27.01173782 |
| 26.61358643 | 26.52173233 | 26.6012249 |
| 24.33546829 | 24.40126801 | 24.34161377 |
| 32.21503448 | 32.16822052 | 32.18370438 |
| 32.64671707 | 32.64842987 | 32.57307816 |
| 26.86843681 | 26.88299179 | 26.80372238 |
| 25.16088295 | 25.09368134 | 25.12404633 |

| | | |
|---|---|---|
| 27.68153954 | 27.61583328 | 27.65037727 |
| 29.58695793 | 29.51377106 | 29.55937386 |
| 26.93140793 | 26.94366646 | 26.89355278 |
| 31.16094971 | 31.15891647 | 31.1147995 |

| Coverage [ | # Unique P | Mean | Standard d | Coefficient | Accession | Gene Name | Number |
|---|---|---|---|---|---|---|---|
| 14 | 1 | 24.73431 | 4.383935 | 0.177241 | Q15843 | NEDD8 | 1 |
| 14 | 5 | 22.36415 | 3.24238 | 0.144981 | Q9H8Y8 | GORASP2 | 2 |
| 2 | 1 | 18.57023 | 2.65506 | 0.142974 | Q6N069 | NAA16 | 3 |
| 28 | 4 | 21.4125 | 2.599552 | 0.121403 | P10451 | SPP1 | 4 |
| 19 | 5 | 19.77646 | 2.22442 | 0.112478 | Q99536 | VAT1 | 5 |
| 9 | 1 | 21.64093 | 2.313411 | 0.1069 | Q04941 | PLP2 | 6 |
| 4 | 16 | 23.69377 | 2.49609 | 0.105348 | Q7Z6Z7 | HUWE1 | 7 |
| 5 | 2 | 20.36175 | 2.049832 | 0.100671 | O14773 | TPP1 | 8 |
| 2 | 1 | 20.41062 | 2.017982 | 0.098869 | Q8N3Z6 | ZCCHC7 | 9 |
| 1 | 1 | 17.1568 | 1.676617 | 0.097723 | P42226 | STAT6 | 10 |
| 3 | 2 | 17.53418 | 1.687717 | 0.096253 | P02788 | LTF | 11 |
| 21 | 1 | 18.15516 | 1.735383 | 0.095586 | Q8IZP0 | ABI1 | 12 |
| 11 | 5 | 20.2403 | 1.862412 | 0.092015 | Q15172 | PPP2R5A | 13 |
| 3 | 4 | 20.13849 | 1.844466 | 0.091589 | P26640 | VARS1 | 14 |
| 3 | 3 | 14.75543 | 1.323258 | 0.089679 | O75122 | CLASP2 | 15 |
| 3 | 1 | 19.53788 | 1.705544 | 0.087294 | Q8NBI5 | SLC43A3 | 16 |
| 4 | 5 | 19.64674 | 1.690801 | 0.08606 | P01133 | EGF | 17 |
| 1 | 1 | 16.43767 | 1.360489 | 0.082767 | Q86V97 | KBTBD6 | 18 |
| 9 | 6 | 18.32665 | 1.512389 | 0.082524 | Q12933 | TRAF2 | 19 |
| 12 | 3 | 17.94313 | 1.478027 | 0.082373 | Q9Y673 | ALG5 | 20 |
| 2 | 5 | 23.44076 | 1.926446 | 0.082184 | Q6AHZ1 | ZNF518A | 21 |
| 1 | 1 | 19.01714 | 1.537282 | 0.080837 | P48736 | PIK3CG | 22 |
| 28 | 1 | 20.28367 | 1.635338 | 0.080623 | P14618 | PKM | 23 |
| 2 | 2 | 18.56511 | 1.482784 | 0.079869 | Q13619 | CUL4A | 24 |
| 7 | 3 | 19.54659 | 1.538433 | 0.078706 | P08559 | PDHA1 | 25 |
| 5 | 1 | 19.55217 | 1.535112 | 0.078514 | P28676 | GCA | 26 |
| 25 | 10 | 24.62135 | 1.909693 | 0.077563 | P07358 | C8B | 27 |
| 9 | 2 | 18.35245 | 1.412022 | 0.076939 | Q92572 | AP3S1 | 28 |
| 32 | 4 | 23.64487 | 1.818835 | 0.076923 | P13501 | CCL5 | 29 |
| 11 | 23 | 24.20383 | 1.860868 | 0.076883 | Q14643 | ITPR1 | 30 |
| 1 | 1 | 19.65034 | 1.498943 | 0.076281 | P28799 | GRN | 31 |
| 13 | 4 | 18.9169 | 1.431948 | 0.075697 | Q92890 | UFD1 | 32 |
| 8 | 2 | 19.97837 | 1.50266 | 0.075214 | Q16270 | IGFBP7 | 33 |
| 2 | 1 | 18.71011 | 1.405614 | 0.075126 | P15291 | B4GALT1 | 34 |
| 19 | 6 | 24.16989 | 1.787631 | 0.073961 | Q9NP79 | VTA1 | 35 |
| 11 | 8 | 20.79402 | 1.507374 | 0.072491 | Q96SB3 | PPP1R9B | 36 |
| 14 | 2 | 20.30697 | 1.46985 | 0.072382 | A0A075B6 | IGLV7-46 | 37 |
| 2 | 3 | 18.99032 | 1.369231 | 0.072102 | O15067 | PFAS | 38 |
| 2 | 1 | 18.87462 | 1.345091 | 0.071265 | Q96QS1 | TSPAN32 | 39 |
| 6 | 4 | 18.24628 | 1.289366 | 0.070665 | P47897 | QARS1 | 40 |
| 1 | 1 | 20.56975 | 1.453154 | 0.070645 | Q49MG5 | MAP9 | 41 |
| 4 | 3 | 23.04504 | 1.609879 | 0.069858 | Q9Y263 | PLAA | 42 |
| 1 | 1 | 17.28763 | 1.194239 | 0.069081 | Q8NBR6 | MINDY2 | 43 |
| 5 | 5 | 18.78351 | 1.294651 | 0.068925 | Q0VD83 | APOBR | 44 |

| | | | | | | | |
|---|---|---|---|---|---|---|---|
| 3 | 1 | 19.1979 | 1.323186 | 0.068923 | Q9UPZ9 | CILK1 | 45 |
| 14 | 3 | 23.00327 | 1.585073 | 0.068906 | P29972 | AQP1 | 46 |
| 11 | 2 | 22.37089 | 1.533115 | 0.068532 | P10153 | RNASE2 | 47 |
| 9 | 10 | 18.04818 | 1.234887 | 0.068422 | Q00722 | PLCB2 | 48 |
| 2 | 1 | 16.9442 | 1.158789 | 0.068388 | Q8NEZ2 | VPS37A | 49 |
| 1 | 1 | 18.78504 | 1.270944 | 0.067657 | Q9ULH0 | KIDINS220 | 50 |
| 7 | 4 | 21.80744 | 1.46443 | 0.067153 | Q9Y259 | CHKB | 51 |
| 2 | 1 | 20.85773 | 1.399581 | 0.067101 | P23352 | ANOS1 | 52 |
| 1 | 1 | 18.86779 | 1.262875 | 0.066933 | Q9UPV9 | TRAK1 | 53 |
| 4 | 3 | 19.73849 | 1.31544 | 0.066643 | Q9BVG8 | KIFC3 | 54 |
| 6 | 10 | 23.54622 | 1.551404 | 0.065888 | Q14571 | ITPR2 | 55 |
| 4 | 2 | 18.42017 | 1.212716 | 0.065836 | Q9Y2T2 | AP3M1 | 56 |
| 2 | 1 | 14.81431 | 0.972798 | 0.065666 | Q86TX2 | ACOT1 | 57 |
| 3 | 1 | 18.72427 | 1.216331 | 0.06496 | P48307 | TFPI2 | 58 |
| 7 | 4 | 19.34593 | 1.256228 | 0.064935 | P08473 | MME | 59 |
| 3 | 4 | 19.23827 | 1.246744 | 0.064805 | Q92835 | INPP5D | 60 |
| 5 | 2 | 19.60856 | 1.270463 | 0.064791 | P00742 | F10 | 61 |
| 0 | 1 | 19.14654 | 1.227924 | 0.064133 | Q9NZJ4 | SACS | 62 |
| 40 | 3 | 21.57897 | 1.376394 | 0.063784 | Q15836 | VAMP3 | 63 |
| 3 | 2 | 18.34785 | 1.163649 | 0.063422 | O00461 | GOLIM4 | 64 |
| 5 | 1 | 17.13296 | 1.082603 | 0.063188 | Q8TDH9 | BLOC1S5 | 65 |
| 8 | 4 | 18.6063 | 1.168267 | 0.062789 | Q16881 | TXNRD1 | 66 |
| 3 | 1 | 17.31143 | 1.085284 | 0.062692 | P45985 | MAP2K4 | 67 |
| 7 | 3 | 18.51128 | 1.158949 | 0.062608 | Q658P3 | STEAP3 | 68 |
| 7 | 1 | 15.36785 | 0.942925 | 0.061357 | Q9Y5J6 | TIMM10B | 69 |
| 14 | 16 | 21.79153 | 1.326683 | 0.060881 | O00203 | AP3B1 | 70 |
| 7 | 5 | 18.76012 | 1.14057 | 0.060798 | Q8TCJ2 | STT3B | 71 |
| 1 | 1 | 16.96563 | 1.028286 | 0.06061 | Q9BTW9 | TBCD | 72 |
| 15 | 2 | 20.4155 | 1.235647 | 0.060525 | Q15056 | EIF4H | 73 |
| 13 | 7 | 21.38909 | 1.285993 | 0.060124 | Q9H9C1 | VIPAS39 | 74 |
| 8 | 2 | 21.83512 | 1.293719 | 0.059249 | O96011 | PEX11B | 75 |
| 36 | 4 | 21.74474 | 1.287477 | 0.059209 | Q6GMV3 | PTRHD1 | 76 |
| 2 | 2 | 18.13776 | 1.061236 | 0.05851 | Q9UPN4 | CEP131 | 77 |
| 32 | 6 | 22.86707 | 1.307713 | 0.057188 | P07741 | APRT | 78 |
| 15 | 2 | 21.6031 | 1.230978 | 0.056982 | P24534 | EEF1B2 | 79 |
| 2 | 1 | 16.08394 | 0.910416 | 0.056604 | Q96FS4 | SIPA1 | 80 |
| 3 | 1 | 19.77723 | 1.118291 | 0.056544 | Q8IXJ6 | SIRT2 | 81 |
| 29 | 3 | 19.25753 | 1.082344 | 0.056204 | P60520 | GABARAPL2 | 82 |
| 9 | 3 | 20.26949 | 1.136805 | 0.056085 | P51665 | PSMD7 | 83 |
| 34 | 3 | 22.16095 | 1.231 | 0.055548 | P82909 | MRPS36 | 84 |
| 34 | 7 | 24.26297 | 1.346534 | 0.055497 | Q8N128 | FAM177A1 | 85 |
| 4 | 1 | 16.86371 | 0.927958 | 0.055027 | Q5TC84 | OGFRL1 | 86 |
| 4 | 1 | 17.19299 | 0.938459 | 0.054584 | P08311 | CTSG | 87 |
| 30 | 7 | 19.71446 | 1.068693 | 0.054209 | O95571 | ETHE1 | 88 |
| 3 | 4 | 17.48299 | 0.947527 | 0.054197 | O00159 | MYO1C | 89 |
| 19 | 7 | 20.61335 | 1.11528 | 0.054105 | Q8N1N4 | KRT78 | 90 |
| 5 | 5 | 15.98079 | 0.862754 | 0.053987 | P78344 | EIF4G2 | 91 |

| | | | | | | | |
|---|---|---|---|---|---|---|---|
| 22 | 4 | 25.47485 | 1.370931 | 0.053815 | P00492 | HPRT1 | 92 |
| 4 | 1 | 20.40811 | 1.097308 | 0.053768 | Q9Y315 | DERA | 93 |
| 16 | 6 | 23.6262 | 1.262243 | 0.053426 | P00740 | F9 | 94 |
| 11 | 7 | 18.64815 | 0.973565 | 0.052207 | P03951 | F11 | 95 |
| 5 | 6 | 19.88998 | 1.029661 | 0.051768 | Q99683 | MAP3K5 | 96 |
| 4 | 1 | 19.23174 | 0.994314 | 0.051702 | P49914 | MTHFS | 97 |
| 3 | 3 | 18.84502 | 0.964388 | 0.051175 | Q14764 | MVP | 98 |
| 1 | 1 | 17.06273 | 0.867307 | 0.05083 | P08183 | ABCB1 | 99 |
| 0 | 4 | 23.67896 | 1.200774 | 0.050711 | Q8NF91 | SYNE1 | 100 |
| 14 | 4 | 18.09314 | 0.915883 | 0.05062 | Q96A49 | SYAP1 | 101 |
| 12 | 5 | 18.01207 | 0.904027 | 0.05019 | Q9H0X4 | FAM234A | 102 |
| 7 | 5 | 18.94094 | 0.949471 | 0.050128 | O14639 | ABLIM1 | 103 |
| 8 | 9 | 19.83375 | 0.98099 | 0.049461 | Q6VY07 | PACS1 | 104 |
| 9 | 4 | 23.67298 | 1.168204 | 0.049348 | P53634 | CTSC | 105 |
| 16 | 3 | 21.18637 | 1.026382 | 0.048445 | P61086 | UBE2K | 106 |
| 19 | 2 | 20.95761 | 1.002318 | 0.047826 | Q8WUH6 | TMEM263 | 107 |
| 17 | 6 | 21.9222 | 1.043931 | 0.04762 | P61201 | COPS2 | 108 |
| 4 | 3 | 18.68304 | 0.887953 | 0.047527 | Q32P44 | EML3 | 109 |
| 14 | 3 | 19.85358 | 0.923081 | 0.046494 | O15126 | SCAMP1 | 110 |
| 4 | 3 | 22.39801 | 1.041165 | 0.046485 | Q6P1M0 | SLC27A4 | 111 |
| 0 | 1 | 19.84196 | 0.920836 | 0.046408 | Q99698 | LYST | 112 |
| 5 | 2 | 18.05086 | 0.833921 | 0.046198 | O95433 | AHSA1 | 113 |
| 5 | 3 | 17.95703 | 0.826403 | 0.046021 | O43776 | NARS1 | 114 |
| 15 | 2 | 21.17844 | 0.972274 | 0.045909 | A0A0C4DH | IGHV4-28 | 115 |
| 26 | 10 | 25.52662 | 1.171403 | 0.045889 | Q14254 | FLOT2 | 116 |
| 17 | 2 | 19.92973 | 0.905053 | 0.045412 | L0R6Q1 | SLC35A4 | 117 |
| 1 | 1 | 18.09494 | 0.821608 | 0.045405 | Q9H8L6 | MMRN2 | 118 |
| 3 | 4 | 18.9359 | 0.858837 | 0.045355 | Q9NZM3 | ITSN2 | 119 |
| 14 | 5 | 23.4171 | 1.06104 | 0.04531 | Q99952 | PTPN18 | 120 |
| 2 | 6 | 18.61388 | 0.841709 | 0.045219 | Q4LDE5 | SVEP1 | 121 |
| 1 | 1 | 21.57769 | 0.974407 | 0.045158 | P22735 | TGM1 | 122 |
| 3 | 2 | 19.97131 | 0.900247 | 0.045077 | Q6ZSS7 | MFSD6 | 123 |
| 33 | 3 | 19.23312 | 0.863157 | 0.044879 | O14548 | COX7A2L | 124 |
| 14 | 6 | 19.64576 | 0.880481 | 0.044818 | O94925 | GLS | 125 |
| 18 | 4 | 19.95428 | 0.892752 | 0.04474 | Q9Y385 | UBE2J1 | 126 |
| 2 | 1 | 16.09239 | 0.717795 | 0.044605 | Q9H0B8 | CRISPLD2 | 127 |
| 11 | 6 | 19.56121 | 0.872485 | 0.044603 | P12109 | COL6A1 | 128 |
| 19 | 4 | 21.9748 | 0.975867 | 0.044408 | P51970 | NDUFA8 | 129 |
| 2 | 1 | 19.06594 | 0.845224 | 0.044332 | P51570 | GALK1 | 130 |
| 26 | 3 | 24.54203 | 1.087486 | 0.044311 | Q562R1 | ACTBL2 | 131 |
| 46 | 5 | 20.71971 | 0.907409 | 0.043794 | P14927 | UQCRB | 132 |
| 30 | 7 | 20.11604 | 0.878115 | 0.043652 | O95721 | SNAP29 | 133 |
| 18 | 2 | 19.23807 | 0.833554 | 0.043328 | A0A0C4DH | IGKV1-8 | 134 |
| 7 | 17 | 21.86626 | 0.944537 | 0.043196 | Q6ZNJ1 | NBEAL2 | 135 |
| 20 | 3 | 20.96026 | 0.903051 | 0.043084 | Q9Y2B0 | CNPY2 | 136 |
| 15 | 3 | 20.80385 | 0.894721 | 0.043007 | P36543 | ATP6V1E1 | 137 |
| 10 | 4 | 18.55442 | 0.796487 | 0.042927 | P00352 | ALDH1A1 | 138 |

| | | | | | | | |
|---|---|---|---|---|---|---|---|
| 8 | 1 | 18.89221 | 0.810089 | 0.04288 | P10124 | SRGN | 139 |
| 10 | 4 | 20.67903 | 0.880292 | 0.042569 | O15533 | TAPBP | 140 |
| 15 | 9 | 22.68532 | 0.964572 | 0.04252 | Q13131 | PRKAA1 | 141 |
| 8 | 1 | 18.50417 | 0.783702 | 0.042353 | Q9BQB6 | VKORC1 | 142 |
| 30 | 11 | 23.00287 | 0.970782 | 0.042203 | Q12907 | LMAN2 | 143 |
| 14 | 7 | 19.6811 | 0.829449 | 0.042144 | Q9UHG3 | PCYOX1 | 144 |
| 16 | 4 | 19.73463 | 0.829228 | 0.042019 | Q9HCN8 | SDF2L1 | 145 |
| 3 | 2 | 18.38658 | 0.771734 | 0.041973 | Q9NW68 | BSDC1 | 146 |
| 5 | 2 | 23.2666 | 0.972022 | 0.041778 | Q9NWW5 | CLN6 | 147 |
| 10 | 4 | 20.57433 | 0.856823 | 0.041645 | P54922 | ADPRH | 148 |
| 10 | 4 | 19.8489 | 0.822284 | 0.041427 | P53582 | METAP1 | 149 |
| 1 | 1 | 16.26777 | 0.673189 | 0.041382 | Q8N5I2 | ARRDC1 | 150 |
| 1 | 1 | 18.87678 | 0.777094 | 0.041167 | Q969S3 | ZNF622 | 151 |
| 12 | 3 | 20.26668 | 0.832306 | 0.041068 | Q9NZ43 | USE1 | 152 |
| 15 | 9 | 19.77658 | 0.808375 | 0.040875 | P54136 | RARS1 | 153 |
| 13 | 2 | 17.9834 | 0.734495 | 0.040843 | P07498 | CSN3 | 154 |
| 17 | 12 | 23.91718 | 0.976146 | 0.040814 | P46459 | NSF | 155 |
| 4 | 6 | 19.67636 | 0.800442 | 0.04068 | O60449 | LY75 | 156 |
| 15 | 8 | 19.95365 | 0.809829 | 0.040585 | P21281 | ATP6V1B2 | 157 |
| 5 | 7 | 16.68727 | 0.676615 | 0.040547 | Q9HBL0 | TNS1 | 158 |
| 48 | 3 | 20.0855 | 0.814138 | 0.040534 | A0A0B4J1Y | IGHV3-72 | 159 |
| 1 | 1 | 17.3479 | 0.701627 | 0.040444 | O95155 | UBE4B | 160 |
| 3 | 3 | 19.67917 | 0.795323 | 0.040414 | Q9Y6Y8 | SEC23IP | 161 |
| 23 | 4 | 21.0316 | 0.848476 | 0.040343 | Q15762 | CD226 | 162 |
| 15 | 3 | 21.25396 | 0.856007 | 0.040275 | Q13445 | TMED1 | 163 |
| 27 | 3 | 21.08163 | 0.847293 | 0.040191 | Q9Y2R0 | COA3 | 164 |
| 6 | 5 | 19.08016 | 0.745398 | 0.039067 | P20810 | CAST | 165 |
| 23 | 2 | 22.43867 | 0.876178 | 0.039048 | Q8WUW1 | BRK1 | 166 |
| 22 | 2 | 22.27475 | 0.865834 | 0.038871 | Q3ZAQ7 | VMA21 | 167 |
| 26 | 3 | 21.41857 | 0.832319 | 0.03886 | O95182 | NDUFA7 | 168 |
| 15 | 2 | 18.81983 | 0.727942 | 0.03868 | Q8N5H3 | FAM89B | 169 |
| 3 | 1 | 19.05902 | 0.736155 | 0.038625 | Q15118 | PDK1 | 170 |
| 10 | 7 | 18.88099 | 0.728351 | 0.038576 | O95140 | MFN2 | 171 |
| 12 | 7 | 21.84514 | 0.840687 | 0.038484 | P23588 | EIF4B | 172 |
| 11 | 6 | 18.82422 | 0.721747 | 0.038341 | Q8TAT6 | NPLOC4 | 173 |
| 11 | 3 | 21.05453 | 0.805555 | 0.03826 | Q86X76 | NIT1 | 174 |
| 7 | 6 | 19.14496 | 0.728833 | 0.038069 | Q9Y4E8 | USP15 | 175 |
| 4 | 2 | 20.26972 | 0.770255 | 0.038 | Q9UHY1 | NRBP1 | 176 |
| 9 | 3 | 21.34359 | 0.809979 | 0.037949 | O75323 | NIPSNAP2 | 177 |
| 10 | 2 | 18.61626 | 0.704666 | 0.037852 | O15258 | RER1 | 178 |
| 2 | 1 | 18.6607 | 0.702891 | 0.037667 | P05067 | APP | 179 |
| 13 | 6 | 20.62775 | 0.77492 | 0.037567 | Q13510 | ASAH1 | 180 |
| 2 | 1 | 17.05848 | 0.639123 | 0.037467 | Q7Z7A4 | PXK | 181 |
| 27 | 4 | 23.05551 | 0.86057 | 0.037326 | P09972 | ALDOC | 182 |
| 7 | 3 | 19.75518 | 0.736938 | 0.037304 | Q9UEW8 | STK39 | 183 |
| 3 | 3 | 18.40222 | 0.682918 | 0.037111 | Q12959 | DLG1 | 184 |
| 17 | 5 | 22.03492 | 0.815653 | 0.037016 | P11177 | PDHB | 185 |

| | | | | | | | |
|---|---|---|---|---|---|---|---|
| 37 | 3 | 22.61302 | 0.831649 | 0.036777 | P61952 | GNG11 | 186 |
| 9 | 11 | 24.18951 | 0.888859 | 0.036746 | Q9P2E9 | RRBP1 | 187 |
| 11 | 6 | 22.07071 | 0.810693 | 0.036732 | P09622 | DLD | 188 |
| 27 | 2 | 20.06925 | 0.734671 | 0.036607 | Q9P1F3 | ABRACL | 189 |
| 3 | 1 | 18.71899 | 0.684595 | 0.036572 | Q8NCA5 | FAM98A | 190 |
| 11 | 2 | 19.57001 | 0.715356 | 0.036554 | P56377 | AP1S2 | 191 |
| 27 | 12 | 23.65891 | 0.864166 | 0.036526 | Q7L591 | DOK3 | 192 |
| 2 | 6 | 20.40156 | 0.744262 | 0.036481 | P42345 | MTOR | 193 |
| 1 | 1 | 17.80577 | 0.64711 | 0.036343 | Q12882 | DPYD | 194 |
| 8 | 4 | 19.50967 | 0.708588 | 0.03632 | O75298 | RTN2 | 195 |
| 43 | 4 | 22.58258 | 0.819934 | 0.036308 | P14209 | CD99 | 196 |
| 4 | 1 | 14.25122 | 0.517437 | 0.036308 | P48454 | PPP3CC | 197 |
| 3 | 1 | 21.03075 | 0.762455 | 0.036254 | P57764 | GSDMD | 198 |
| 18 | 3 | 21.8765 | 0.79219 | 0.036212 | Q969H8 | MYDGF | 199 |
| 34 | 10 | 23.26669 | 0.838335 | 0.036032 | Q9Y3F4 | STRAP | 200 |
| 20 | 2 | 20.73163 | 0.744943 | 0.035933 | P06310 | IGKV2-30 | 201 |
| 3 | 10 | 21.33451 | 0.766037 | 0.035906 | P78509 | RELN | 202 |
| 12 | 4 | 20.06978 | 0.720279 | 0.035889 | Q8IWB7 | WDFY1 | 203 |
| 53 | 12 | 23.95714 | 0.855739 | 0.03572 | O43399 | TPD52L2 | 204 |
| 12 | 5 | 19.48187 | 0.689943 | 0.035415 | P36955 | SERPINF1 | 205 |
| 39 | 3 | 22.27649 | 0.788356 | 0.03539 | P80748 | IGLV3-21 | 206 |
| 7 | 4 | 20.62513 | 0.72823 | 0.035308 | Q8NBM8 | PCYOX1L | 207 |
| 35 | 2 | 23.12426 | 0.814439 | 0.03522 | Q92930 | RAB8B | 208 |
| 4 | 3 | 19.33915 | 0.678686 | 0.035094 | P29122 | PCSK6 | 209 |
| 16 | 5 | 20.03365 | 0.701677 | 0.035025 | O94905 | ERLIN2 | 210 |
| 4 | 2 | 17.38612 | 0.605883 | 0.034849 | Q9HD26 | GOPC | 211 |
| 19 | 11 | 20.52703 | 0.712884 | 0.034729 | Q13492 | PICALM | 212 |
| 24 | 3 | 23.68776 | 0.822165 | 0.034708 | O60925 | PFDN1 | 213 |
| 6 | 4 | 22.81795 | 0.791719 | 0.034697 | P61221 | ABCE1 | 214 |
| 12 | 1 | 21.33297 | 0.73841 | 0.034614 | Q9NX55 | HYPK | 215 |
| 19 | 8 | 19.89267 | 0.685849 | 0.034477 | P62191 | PSMC1 | 216 |
| 2 | 1 | 18.70848 | 0.643439 | 0.034393 | Q9H1Z4 | WDR13 | 217 |
| 5 | 1 | 18.40675 | 0.631492 | 0.034308 | Q8TBP5 | FAM174A | 218 |
| 4 | 1 | 17.71968 | 0.606806 | 0.034245 | Q96MH2 | HEXIM2 | 219 |
| 29 | 11 | 22.79817 | 0.77868 | 0.034155 | O60664 | PLIN3 | 220 |
| 39 | 5 | 22.15271 | 0.75507 | 0.034085 | P09382 | LGALS1 | 221 |
| 17 | 5 | 22.84264 | 0.778327 | 0.034073 | P25787 | PSMA2 | 222 |
| 8 | 3 | 19.9592 | 0.675702 | 0.033854 | Q8NBQ5 | HSD17B11 | 223 |
| 10 | 4 | 18.58 | 0.628929 | 0.03385 | Q9Y6K9 | IKBKG | 224 |
| 7 | 2 | 20.70537 | 0.699428 | 0.03378 | P08865 | RPSA | 225 |
| 2 | 1 | 19.38124 | 0.651461 | 0.033613 | A1L0T0 | ILVBL | 226 |
| 5 | 2 | 19.79454 | 0.663591 | 0.033524 | Q13564 | NAE1 | 227 |
| 2 | 1 | 16.38218 | 0.543053 | 0.033149 | Q96B97 | SH3KBP1 | 228 |
| 5 | 4 | 21.83196 | 0.723326 | 0.033132 | O14920 | IKBKB | 229 |
| 20 | 5 | 21.14795 | 0.700106 | 0.033105 | P35625 | TIMP3 | 230 |
| 16 | 3 | 21.23368 | 0.701307 | 0.033028 | Q8N5K1 | CISD2 | 231 |
| 44 | 2 | 19.65968 | 0.646679 | 0.032894 | P02538 | KRT6A | 232 |

| | | | | | | | |
|---|---|---|---|---|---|---|---|
| 8 | 4 | 20.56314 | 0.675859 | 0.032867 | Q9BXR6 | CFHR5 | 233 |
| 18 | 2 | 21.27043 | 0.698356 | 0.032832 | P0DMM9 | SULT1A3 | 234 |
| 45 | 9 | 25.6623 | 0.841523 | 0.032792 | P30405 | PPIF | 235 |
| 16 | 3 | 21.88355 | 0.71714 | 0.032771 | Q9BVK6 | TMED9 | 236 |
| 3 | 3 | 19.03687 | 0.623074 | 0.03273 | Q8IWB9 | TEX2 | 237 |
| 2 | 1 | 18.28214 | 0.597512 | 0.032683 | Q9H9A5 | CNOT10 | 238 |
| 7 | 2 | 20.67917 | 0.673961 | 0.032591 | P29965 | CD40LG | 239 |
| 1 | 1 | 19.13598 | 0.623142 | 0.032564 | Q7Z7B0 | FILIP1 | 240 |
| 6 | 12 | 20.56068 | 0.669459 | 0.03256 | Q9HCM2 | PLXNA4 | 241 |
| 8 | 6 | 20.88709 | 0.679561 | 0.032535 | Q5VY43 | PEAR1 | 242 |
| 23 | 4 | 21.32722 | 0.691237 | 0.032411 | Q9Y3B3 | TMED7 | 243 |
| 23 | 4 | 20.3816 | 0.660307 | 0.032397 | P51571 | SSR4 | 244 |
| 5 | 1 | 18.20559 | 0.588782 | 0.032341 | Q9BY32 | ITPA | 245 |
| 16 | 4 | 19.63116 | 0.634394 | 0.032316 | Q9H8W4 | PLEKHF2 | 246 |
| 8 | 4 | 20.50547 | 0.66073 | 0.032222 | O95831 | AIFM1 | 247 |
| 28 | 5 | 19.91845 | 0.641555 | 0.032209 | P37235 | HPCAL1 | 248 |
| 23 | 6 | 23.48541 | 0.7553 | 0.03216 | Q15382 | RHEB | 249 |
| 12 | 10 | 19.66428 | 0.625953 | 0.031832 | Q7Z2W4 | ZC3HAV1 | 250 |
| 4 | 3 | 18.00883 | 0.57104 | 0.031709 | O94874 | UFL1 | 251 |
| 13 | 2 | 16.72882 | 0.530054 | 0.031685 | Q96C01 | FAM136A | 252 |
| 10 | 6 | 22.32385 | 0.706507 | 0.031648 | O60506 | SYNCRIP | 253 |
| 15 | 6 | 22.265 | 0.704462 | 0.03164 | Q9BWD1 | ACAT2 | 254 |
| 13 | 4 | 19.02332 | 0.600973 | 0.031591 | Q9BS40 | LXN | 255 |
| 12 | 2 | 18.79895 | 0.591941 | 0.031488 | Q969M7 | UBE2F | 256 |
| 26 | 4 | 22.70164 | 0.711722 | 0.031351 | P61626 | LYZ | 257 |
| 34 | 9 | 23.75849 | 0.743866 | 0.031309 | Q9Y696 | CLIC4 | 258 |
| 8 | 2 | 18.50859 | 0.579428 | 0.031306 | Q2TAA2 | IAH1 | 259 |
| 11 | 7 | 21.89252 | 0.685141 | 0.031296 | Q9NRL3 | STRN4 | 260 |
| 22 | 5 | 24.27592 | 0.754099 | 0.031064 | Q16762 | TST | 261 |
| 35 | 4 | 27.63431 | 0.857721 | 0.031038 | P68032 | ACTC1 | 262 |
| 28 | 10 | 23.99286 | 0.7432 | 0.030976 | Q14344 | GNA13 | 263 |
| 26 | 2 | 19.59646 | 0.606302 | 0.030939 | P60468 | SEC61B | 264 |
| 10 | 6 | 22.84755 | 0.706245 | 0.030911 | Q8NBF2 | NHLRC2 | 265 |
| 10 | 3 | 19.72332 | 0.608714 | 0.030863 | P80217 | IFI35 | 266 |
| 26 | 2 | 19.70143 | 0.605735 | 0.030746 | Q86YW5 | TREML1 | 267 |
| 3 | 1 | 21.75876 | 0.668271 | 0.030713 | Q9BXA7 | TSSK1B | 268 |
| 2 | 1 | 18.85752 | 0.573978 | 0.030438 | P49757 | NUMB | 269 |
| 17 | 3 | 20.39998 | 0.620648 | 0.030424 | P01127 | PDGFB | 270 |
| 32 | 3 | 20.19011 | 0.612106 | 0.030317 | P56385 | ATP5ME | 271 |
| 8 | 10 | 21.6107 | 0.653151 | 0.030224 | O14980 | XPO1 | 272 |
| 13 | 5 | 19.24647 | 0.581035 | 0.030189 | P55010 | EIF5 | 273 |
| 15 | 2 | 21.19186 | 0.639012 | 0.030154 | O15212 | PFDN6 | 274 |
| 7 | 4 | 20.49424 | 0.617365 | 0.030124 | Q5H9R7 | PPP6R3 | 275 |
| 48 | 4 | 25.12676 | 0.755607 | 0.030072 | P48668 | KRT6C | 276 |
| 10 | 6 | 23.86956 | 0.713197 | 0.029879 | Q8IVB4 | SLC9A9 | 277 |
| 24 | 1 | 19.26803 | 0.572669 | 0.029721 | P56134 | ATP5MF | 278 |
| 6 | 2 | 20.87257 | 0.620126 | 0.02971 | Q9BSK0 | MARVELD1 | 279 |

| | | | | | | | |
|---|---|---|---|---|---|---|---|
| 9 | 5 | 22.24101 | 0.660603 | 0.029702 | Q9Y6W5 | WASF2 | 280 |
| 11 | 3 | 18.21004 | 0.540529 | 0.029683 | O14828 | SCAMP3 | 281 |
| 3 | 1 | 17.56594 | 0.521135 | 0.029667 | Q9H0U6 | MRPL18 | 282 |
| 24 | 8 | 23.38857 | 0.692394 | 0.029604 | P09104 | ENO2 | 283 |
| 8 | 4 | 20.25055 | 0.59889 | 0.029574 | P08185 | SERPINA6 | 284 |
| 6 | 4 | 20.27623 | 0.597233 | 0.029455 | O15327 | INPP4B | 285 |
| 23 | 13 | 23.80611 | 0.696852 | 0.029272 | P13797 | PLS3 | 286 |
| 20 | 3 | 19.61003 | 0.57326 | 0.029233 | Q9UHV9 | PFDN2 | 287 |
| 15 | 3 | 21.93898 | 0.636557 | 0.029015 | O15551 | CLDN3 | 288 |
| 10 | 4 | 21.07616 | 0.610763 | 0.028979 | P25685 | DNAJB1 | 289 |
| 12 | 2 | 17.06674 | 0.493522 | 0.028917 | Q16774 | GUK1 | 290 |
| 4 | 6 | 20.57094 | 0.588057 | 0.028587 | O15439 | ABCC4 | 291 |
| 29 | 6 | 19.93311 | 0.567442 | 0.028467 | Q9Y3A3 | MOB4 | 292 |
| 7 | 8 | 21.31402 | 0.606213 | 0.028442 | Q7Z460 | CLASP1 | 293 |
| 9 | 7 | 21.95178 | 0.621125 | 0.028295 | Q08554 | DSC1 | 294 |
| 6 | 6 | 20.97354 | 0.592904 | 0.028269 | O14617 | AP3D1 | 295 |
| 1 | 1 | 19.27924 | 0.540103 | 0.028015 | Q68DL7 | C18orf63 | 296 |
| 6 | 5 | 19.83135 | 0.554891 | 0.027981 | Q86VS8 | HOOK3 | 297 |
| 8 | 15 | 20.20635 | 0.562791 | 0.027852 | Q96BY6 | DOCK10 | 298 |
| 3 | 1 | 19.28215 | 0.536683 | 0.027833 | Q15005 | SPCS2 | 299 |
| 2 | 1 | 18.00994 | 0.499444 | 0.027732 | Q9NZ09 | UBAP1 | 300 |
| 10 | 3 | 20.37825 | 0.564989 | 0.027725 | P54619 | PRKAG1 | 301 |
| 15 | 6 | 22.23518 | 0.613993 | 0.027614 | Q14012 | CAMK1 | 302 |
| 43 | 2 | 21.60462 | 0.596395 | 0.027605 | Q96IX5 | ATP5MK | 303 |
| 15 | 4 | 21.50474 | 0.592582 | 0.027556 | Q7L5N1 | COPS6 | 304 |
| 30 | 7 | 22.65944 | 0.619327 | 0.027332 | Q9BWM7 | SFXN3 | 305 |
| 15 | 3 | 22.42068 | 0.61167 | 0.027282 | P00403 | MT-CO2 | 306 |
| 10 | 3 | 20.18237 | 0.548775 | 0.027191 | Q9H0W9 | C11orf54 | 307 |
| 26 | 6 | 21.94555 | 0.595962 | 0.027156 | Q04760 | GLO1 | 308 |
| 19 | 6 | 21.72106 | 0.589779 | 0.027152 | Q96CN7 | ISOC1 | 309 |
| 3 | 1 | 19.03509 | 0.515717 | 0.027093 | Q9Y619 | SLC25A15 | 310 |
| 1 | 1 | 17.12983 | 0.46387 | 0.02708 | Q5VYS8 | TUT7 | 311 |
| 7 | 3 | 19.15485 | 0.5186 | 0.027074 | Q9NS86 | LANCL2 | 312 |
| 40 | 9 | 28.00244 | 0.755567 | 0.026982 | P05090 | APOD | 313 |
| 12 | 5 | 21.66149 | 0.583271 | 0.026927 | O75190 | DNAJB6 | 314 |
| 8 | 3 | 21.05418 | 0.565397 | 0.026854 | P08574 | CYC1 | 315 |
| 14 | 20 | 25.60457 | 0.687403 | 0.026847 | Q04637 | EIF4G1 | 316 |
| 8 | 6 | 20.93402 | 0.561977 | 0.026845 | P22059 | OSBP | 317 |
| 5 | 2 | 20.03645 | 0.536221 | 0.026762 | O15269 | SPTLC1 | 318 |
| 8 | 5 | 28.11685 | 0.750185 | 0.026681 | Q9NZQ3 | NCKIPSD | 319 |
| 5 | 2 | 19.33176 | 0.51449 | 0.026614 | P12268 | IMPDH2 | 320 |
| 9 | 2 | 19.12908 | 0.50878 | 0.026597 | Q9NRW3 | APOBEC3C | 321 |
| 32 | 16 | 27.04716 | 0.718322 | 0.026558 | P02774 | GC | 322 |
| 8 | 3 | 20.44227 | 0.542294 | 0.026528 | Q13347 | EIF3I | 323 |
| 10 | 1 | 19.4688 | 0.514512 | 0.026428 | P24311 | COX7B | 324 |
| 7 | 4 | 21.18779 | 0.559286 | 0.026397 | P11166 | SLC2A1 | 325 |
| 6 | 6 | 24.17525 | 0.634876 | 0.026261 | Q9Y4W6 | AFG3L2 | 326 |

| | | | | | | | |
|---|---|---|---|---|---|---|---|
| 14 | 4 | 21.29472 | 0.555619 | 0.026092 | Q8IZD0 | SAMD14 | 327 |
| 3 | 1 | 18.54905 | 0.481457 | 0.025956 | P19784 | CSNK2A2 | 328 |
| 19 | 1 | 19.17938 | 0.495752 | 0.025848 | Q8TAD7 | OCC1 | 329 |
| 2 | 1 | 17.73572 | 0.457545 | 0.025798 | P55285 | CDH6 | 330 |
| 19 | 7 | 20.4726 | 0.527909 | 0.025786 | O75351 | VPS4B | 331 |
| 5 | 1 | 18.88234 | 0.486093 | 0.025743 | P53680 | AP2S1 | 332 |
| 40 | 9 | 23.28683 | 0.596435 | 0.025613 | P16152 | CBR1 | 333 |
| 18 | 12 | 22.94144 | 0.586885 | 0.025582 | Q9NQW7 | XPNPEP1 | 334 |
| 18 | 7 | 21.99244 | 0.561308 | 0.025523 | P53990 | IST1 | 335 |
| 13 | 4 | 22.61978 | 0.575476 | 0.025441 | P98172 | EFNB1 | 336 |
| 4 | 3 | 20.92815 | 0.532274 | 0.025433 | Q5TCZ1 | SH3PXD2A | 337 |
| 3 | 1 | 17.59804 | 0.447462 | 0.025427 | Q92604 | LPGAT1 | 338 |
| 3 | 14 | 22.5925 | 0.573598 | 0.025389 | O75592 | MYCBP2 | 339 |
| 1 | 1 | 17.48243 | 0.443689 | 0.025379 | Q3LXA3 | TKFC | 340 |
| 7 | 5 | 18.93387 | 0.47827 | 0.02526 | Q8TD16 | BICD2 | 341 |
| 14 | 8 | 19.12224 | 0.482278 | 0.025221 | P05160 | F13B | 342 |
| 22 | 9 | 21.32366 | 0.532158 | 0.024956 | P35998 | PSMC2 | 343 |
| 17 | 3 | 19.45739 | 0.485145 | 0.024934 | P15374 | UCHL3 | 344 |
| 7 | 2 | 21.28946 | 0.529094 | 0.024852 | P08571 | CD14 | 345 |
| 26 | 5 | 20.68353 | 0.513513 | 0.024827 | O00154 | ACOT7 | 346 |
| 18 | 8 | 21.50512 | 0.53157 | 0.024718 | P49257 | LMAN1 | 347 |
| 8 | 5 | 21.24272 | 0.524041 | 0.024669 | Q14258 | TRIM25 | 348 |
| 18 | 8 | 21.30374 | 0.525456 | 0.024665 | P06132 | UROD | 349 |
| 2 | 2 | 19.0205 | 0.467464 | 0.024577 | Q9Y2H0 | DLGAP4 | 350 |
| 1 | 1 | 17.4302 | 0.427089 | 0.024503 | Q07889 | SOS1 | 351 |
| 24 | 6 | 20.97682 | 0.513897 | 0.024498 | Q9UJZ1 | STOML2 | 352 |
| 1 | 1 | 18.17798 | 0.44488 | 0.024474 | Q9P0K7 | RAI14 | 353 |
| 12 | 5 | 21.93499 | 0.53534 | 0.024406 | Q9Y5P6 | GMPPB | 354 |
| 12 | 8 | 21.28069 | 0.519013 | 0.024389 | Q8IZ83 | ALDH16A1 | 355 |
| 16 | 3 | 20.84882 | 0.507412 | 0.024338 | Q9GZP4 | PITHD1 | 356 |
| 2 | 9 | 22.7396 | 0.553122 | 0.024324 | Q8N3K9 | CMYA5 | 357 |
| 31 | 4 | 21.80885 | 0.529667 | 0.024287 | P61019 | RAB2A | 358 |
| 8 | 2 | 18.16976 | 0.441285 | 0.024287 | Q96C19 | EFHD2 | 359 |
| 7 | 13 | 22.37504 | 0.542396 | 0.024241 | Q08378 | GOLGA3 | 360 |
| 1 | 1 | 18.10591 | 0.438224 | 0.024203 | Q8NEN9 | PDZD8 | 361 |
| 3 | 1 | 17.90183 | 0.432282 | 0.024147 | Q9UBX7 | KLK11 | 362 |
| 1 | 1 | 19.09275 | 0.460688 | 0.024129 | Q8IY17 | PNPLA6 | 363 |
| 1 | 1 | 20.35462 | 0.490733 | 0.024109 | Q96M91 | CFAP53 | 364 |
| 6 | 5 | 21.66624 | 0.521772 | 0.024082 | P46977 | STT3A | 365 |
| 20 | 8 | 20.55153 | 0.49492 | 0.024082 | P23526 | AHCY | 366 |
| 14 | 5 | 20.73475 | 0.499241 | 0.024078 | Q05193 | DNM1 | 367 |
| 1 | 1 | 19.97168 | 0.47933 | 0.024 | O15259 | NPHP1 | 368 |
| 4 | 1 | 18.94022 | 0.454258 | 0.023984 | O43402 | EMC8 | 369 |
| 32 | 10 | 27.17477 | 0.647699 | 0.023835 | Q99623 | PHB2 | 370 |
| 16 | 2 | 20.56886 | 0.489204 | 0.023784 | P50225 | SULT1A1 | 371 |
| 19 | 3 | 21.64978 | 0.512773 | 0.023685 | O75832 | PSMD10 | 372 |
| 7 | 2 | 21.71542 | 0.513961 | 0.023668 | Q53GL0 | PLEKHO1 | 373 |

| | | | | | | | |
|---|---|---|---|---|---|---|---|
| 16 | 5 | 21.08082 | 0.49841 | 0.023643 | O76003 | GLRX3 | 374 |
| 4 | 6 | 23.37508 | 0.551637 | 0.023599 | Q8N2N9 | ANKRD36B | 375 |
| 5 | 3 | 20.94214 | 0.491782 | 0.023483 | P32189 | GK | 376 |
| 10 | 2 | 21.22111 | 0.497561 | 0.023447 | O43674 | NDUFB5 | 377 |
| 22 | 6 | 20.27007 | 0.474929 | 0.02343 | Q12846 | STX4 | 378 |
| 6 | 6 | 19.75856 | 0.462488 | 0.023407 | Q86UT6 | NLRX1 | 379 |
| 16 | 1 | 19.25662 | 0.44945 | 0.02334 | P36980 | CFHR2 | 380 |
| 6 | 3 | 18.35199 | 0.427168 | 0.023276 | P08195 | SLC3A2 | 381 |
| 4 | 1 | 19.40217 | 0.45061 | 0.023225 | Q9NVV5 | AIG1 | 382 |
| 17 | 9 | 21.33781 | 0.49467 | 0.023183 | Q9UNF1 | MAGED2 | 383 |
| 2 | 1 | 17.6036 | 0.407759 | 0.023163 | Q96Q11 | TRNT1 | 384 |
| 26 | 6 | 23.81994 | 0.551305 | 0.023145 | O14818 | PSMA7 | 385 |
| 15 | 12 | 21.48207 | 0.496642 | 0.023119 | P42566 | EPS15 | 386 |
| 2 | 1 | 17.75975 | 0.405912 | 0.022856 | Q93099 | HGD | 387 |
| 18 | 16 | 23.71051 | 0.541448 | 0.022836 | Q9UDT6 | CLIP2 | 388 |
| 6 | 2 | 21.27365 | 0.485665 | 0.022829 | O15427 | SLC16A3 | 389 |
| 6 | 6 | 21.07959 | 0.481137 | 0.022825 | Q9Y3P9 | RABGAP1 | 390 |
| 2 | 1 | 19.44501 | 0.44316 | 0.02279 | Q7Z7B1 | PIGW | 391 |
| 3 | 1 | 18.1616 | 0.413761 | 0.022782 | Q15233 | NONO | 392 |
| 3 | 1 | 19.38693 | 0.441357 | 0.022766 | Q8NFQ8 | TOR1AIP2 | 393 |
| 20 | 7 | 21.09277 | 0.479283 | 0.022723 | P29508 | SERPINB3 | 394 |
| 21 | 11 | 21.3672 | 0.485312 | 0.022713 | O00186 | STXBP3 | 395 |
| 12 | 3 | 18.03856 | 0.409476 | 0.0227 | P28289 | TMOD1 | 396 |
| 17 | 8 | 21.63441 | 0.490725 | 0.022683 | P55809 | OXCT1 | 397 |
| 6 | 2 | 19.22566 | 0.435938 | 0.022675 | P23258 | TUBG1 | 398 |
| 5 | 5 | 21.01034 | 0.476056 | 0.022658 | Q92973 | TNPO1 | 399 |
| 15 | 5 | 20.64111 | 0.466834 | 0.022617 | Q9H2M3 | BHMT2 | 400 |
| 27 | 24 | 25.66366 | 0.579966 | 0.022599 | Q8WUM4 | PDCD6IP | 401 |
| 4 | 2 | 19.30488 | 0.436002 | 0.022585 | Q53H12 | AGK | 402 |
| 8 | 3 | 19.51356 | 0.440274 | 0.022562 | Q9UPY6 | WASF3 | 403 |
| 8 | 3 | 19.00505 | 0.428043 | 0.022523 | Q08752 | PPID | 404 |
| 15 | 10 | 22.76833 | 0.510401 | 0.022417 | P26639 | TARS1 | 405 |
| 12 | 18 | 24.32027 | 0.544574 | 0.022392 | Q96P48 | ARAP1 | 406 |
| 12 | 6 | 21.47815 | 0.478943 | 0.022299 | Q13177 | PAK2 | 407 |
| 20 | 20 | 25.8087 | 0.57409 | 0.022244 | P53396 | ACLY | 408 |
| 2 | 5 | 20.98868 | 0.466025 | 0.022204 | Q5THR3 | EFCAB6 | 409 |
| 5 | 5 | 21.42608 | 0.474918 | 0.022165 | Q6WCQ1 | MPRIP | 410 |
| 12 | 4 | 20.51266 | 0.451681 | 0.02202 | P50213 | IDH3A | 411 |
| 2 | 1 | 17.90134 | 0.394147 | 0.022018 | P48163 | ME1 | 412 |
| 10 | 6 | 21.00339 | 0.458679 | 0.021838 | P33151 | CDH5 | 413 |
| 16 | 5 | 21.09764 | 0.459527 | 0.021781 | Q9HAB8 | PPCS | 414 |
| 6 | 4 | 22.07859 | 0.480726 | 0.021773 | Q3YEC7 | RABL6 | 415 |
| 4 | 1 | 19.07351 | 0.414524 | 0.021733 | A6NFX1 | MFSD2B | 416 |
| 3 | 1 | 18.74709 | 0.406636 | 0.021691 | Q9H813 | PACC1 | 417 |
| 2 | 2 | 22.326 | 0.483792 | 0.021669 | P0CW18 | PRSS56 | 418 |
| 9 | 1 | 19.9286 | 0.430644 | 0.021609 | Q9Y2Q5 | LAMTOR2 | 419 |
| 9 | 4 | 19.42439 | 0.419445 | 0.021594 | Q15435 | PPP1R7 | 420 |

| | | | | | | | |
|---|---|---|---|---|---|---|---|
| 2 | 5 | 20.3459 | 0.439148 | 0.021584 | P27708 | CAD | 421 |
| 19 | 4 | 21.0913 | 0.454307 | 0.02154 | P43487 | RANBP1 | 422 |
| 8 | 7 | 21.24886 | 0.457164 | 0.021515 | O95834 | EML2 | 423 |
| 1 | 1 | 18.09845 | 0.388382 | 0.021459 | Q6PJF5 | RHBDF2 | 424 |
| 15 | 4 | 19.95359 | 0.424118 | 0.021255 | Q8NFV4 | ABHD11 | 425 |
| 7 | 1 | 21.41213 | 0.455029 | 0.021251 | Q53FT3 | HIKESHI | 426 |
| 28 | 3 | 26.2175 | 0.5567 | 0.021234 | P26447 | S100A4 | 427 |
| 19 | 3 | 22.40985 | 0.475289 | 0.021209 | Q15185 | PTGES3 | 428 |
| 15 | 6 | 21.7981 | 0.46131 | 0.021163 | Q6NVY1 | HIBCH | 429 |
| 14 | 8 | 21.33466 | 0.451339 | 0.021155 | Q06124 | PTPN11 | 430 |
| 24 | 18 | 22.98981 | 0.485986 | 0.021139 | Q9UBC2 | EPS15L1 | 431 |
| 19 | 11 | 21.81052 | 0.460073 | 0.021094 | Q15642 | TRIP10 | 432 |
| 20 | 8 | 21.72854 | 0.457816 | 0.02107 | P29622 | SERPINA4 | 433 |
| 25 | 1 | 21.95168 | 0.4625 | 0.021069 | P62328 | TMSB4X | 434 |
| 36 | 6 | 21.81281 | 0.459363 | 0.021059 | P05452 | CLEC3B | 435 |
| 15 | 3 | 21.19431 | 0.445085 | 0.021 | P31944 | CASP14 | 436 |
| 17 | 13 | 23.10397 | 0.484492 | 0.02097 | Q02413 | DSG1 | 437 |
| 7 | 4 | 19.92738 | 0.416666 | 0.020909 | Q9UGP4 | LIMD1 | 438 |
| 59 | 4 | 20.45055 | 0.426419 | 0.020851 | P07108 | DBI | 439 |
| 47 | 2 | 21.42592 | 0.445293 | 0.020783 | P62834 | RAP1A | 440 |
| 25 | 9 | 21.66829 | 0.450207 | 0.020777 | P62195 | PSMC5 | 441 |
| 16 | 5 | 21.35803 | 0.442381 | 0.020713 | P55036 | PSMD4 | 442 |
| 17 | 5 | 22.27937 | 0.4614 | 0.02071 | P14324 | FDPS | 443 |
| 17 | 3 | 20.86179 | 0.432008 | 0.020708 | Q7L9L4 | MOB1B | 444 |
| 9 | 4 | 21.76618 | 0.45073 | 0.020708 | Q9UI12 | ATP6V1H | 445 |
| 15 | 11 | 22.81798 | 0.472427 | 0.020704 | Q96QK1 | VPS35 | 446 |
| 10 | 6 | 20.82131 | 0.43053 | 0.020677 | P07602 | PSAP | 447 |
| 29 | 2 | 20.06943 | 0.413665 | 0.020612 | Q9H3K6 | BOLA2 | 448 |
| 6 | 11 | 26.10461 | 0.538016 | 0.02061 | O95425 | SVIL | 449 |
| 25 | 4 | 23.9701 | 0.493355 | 0.020582 | P0C0L4 | C4A | 450 |
| 0 | 1 | 17.085 | 0.351229 | 0.020558 | Q9Y520 | PRRC2C | 451 |
| 0 | 1 | 17.95829 | 0.368692 | 0.02053 | Q5TAX3 | TUT4 | 452 |
| 13 | 2 | 20.92662 | 0.42916 | 0.020508 | O95298 | NDUFC2 | 453 |
| 23 | 4 | 20.04828 | 0.411043 | 0.020503 | O75223 | GGCT | 454 |
| 40 | 10 | 22.9078 | 0.469414 | 0.020491 | Q14696 | MESD | 455 |
| 4 | 1 | 18.46767 | 0.378335 | 0.020486 | Q96K17 | BTF3L4 | 456 |
| 10 | 9 | 21.74305 | 0.445284 | 0.020479 | Q02218 | OGDH | 457 |
| 38 | 1 | 20.50183 | 0.419376 | 0.020456 | P10720 | PF4V1 | 458 |
| 8 | 3 | 20.59204 | 0.420611 | 0.020426 | Q9NRY5 | FAM114A2 | 459 |
| 3 | 1 | 21.02809 | 0.429397 | 0.02042 | P05543 | SERPINA7 | 460 |
| 26 | 26 | 24.95768 | 0.509452 | 0.020413 | O14974 | PPP1R12A | 461 |
| 12 | 5 | 20.7836 | 0.423139 | 0.020359 | P26440 | IVD | 462 |
| 5 | 2 | 21.73458 | 0.439353 | 0.020214 | P52788 | SMS | 463 |
| 14 | 5 | 19.47622 | 0.393598 | 0.020209 | Q9Y285 | FARSA | 464 |
| 18 | 4 | 20.47849 | 0.413221 | 0.020178 | Q8N1Q1 | CA13 | 465 |
| 15 | 9 | 22.56324 | 0.454967 | 0.020164 | P14923 | JUP | 466 |
| 17 | 7 | 22.44068 | 0.451489 | 0.020119 | Q8NE86 | MCU | 467 |

| | | | | | | | |
|---|---|---|---|---|---|---|---|
| 15 | 6 | 20.51096 | 0.412221 | 0.020098 | P16930 | FAH | 468 |
| 17 | 6 | 21.22692 | 0.425629 | 0.020051 | Q9Y371 | SH3GLB1 | 469 |
| 14 | 10 | 24.06945 | 0.482031 | 0.020027 | P11940 | PABPC1 | 470 |
| 2 | 1 | 18.22232 | 0.361632 | 0.019846 | Q8N4C8 | MINK1 | 471 |
| 8 | 1 | 21.6975 | 0.430491 | 0.019841 | Q7Z5L0 | VMO1 | 472 |
| 2 | 1 | 18.98179 | 0.376208 | 0.019819 | Q01415 | GALK2 | 473 |
| 6 | 2 | 22.59794 | 0.447082 | 0.019784 | Q92905 | COPS5 | 474 |
| 14 | 4 | 22.66661 | 0.448208 | 0.019774 | Q96FW1 | OTUB1 | 475 |
| 14 | 3 | 24.14292 | 0.477048 | 0.019759 | P07477 | PRSS1 | 476 |
| 17 | 6 | 20.37769 | 0.398569 | 0.019559 | P46734 | MAP2K3 | 477 |
| 20 | 7 | 20.28785 | 0.396501 | 0.019544 | P21283 | ATP6V1C1 | 478 |
| 44 | 10 | 26.67496 | 0.520432 | 0.01951 | Q04695 | KRT17 | 479 |
| 25 | 10 | 22.11884 | 0.431334 | 0.019501 | Q7L1Q6 | BZW1 | 480 |
| 14 | 1 | 18.68104 | 0.363887 | 0.019479 | P12035 | KRT3 | 481 |
| 8 | 3 | 21.01104 | 0.408546 | 0.019444 | O60704 | TPST2 | 482 |
| 13 | 9 | 21.42091 | 0.416407 | 0.019439 | Q9C0I1 | MTMR12 | 483 |
| 14 | 1 | 19.09856 | 0.369634 | 0.019354 | Q9NYM9 | BET1L | 484 |
| 4 | 1 | 17.97039 | 0.347033 | 0.019311 | Q9H0F6 | SHARPIN | 485 |
| 3 | 1 | 16.1571 | 0.310626 | 0.019225 | Q5TEU4 | NDUFAF5 | 486 |
| 5 | 6 | 19.3195 | 0.371402 | 0.019224 | Q96K76 | USP47 | 487 |
| 25 | 2 | 18.99724 | 0.364309 | 0.019177 | P55774 | CCL18 | 488 |
| 18 | 1 | 22.89789 | 0.438753 | 0.019161 | Q9UDY2 | TJP2 | 489 |
| 13 | 5 | 22.15241 | 0.423022 | 0.019096 | P04070 | PROC | 490 |
| 20 | 20 | 25.53216 | 0.484342 | 0.01897 | P05023 | ATP1A1 | 491 |
| 14 | 4 | 21.08 | 0.39928 | 0.018941 | Q13126 | MTAP | 492 |
| 9 | 11 | 21.21569 | 0.400499 | 0.018878 | Q86UP2 | KTN1 | 493 |
| 1 | 1 | 16.39181 | 0.309255 | 0.018866 | Q9BX69 | CARD6 | 494 |
| 7 | 4 | 19.80155 | 0.373375 | 0.018856 | P28331 | NDUFS1 | 495 |
| 4 | 5 | 22.2019 | 0.418161 | 0.018834 | Q01970 | PLCB3 | 496 |
| 3 | 4 | 19.66599 | 0.3688 | 0.018753 | Q6YHK3 | CD109 | 497 |
| 31 | 3 | 21.34705 | 0.399541 | 0.018716 | P61956 | SUMO2 | 498 |
| 42 | 12 | 23.75655 | 0.444169 | 0.018697 | P31150 | GDI1 | 499 |
| 4 | 2 | 19.51387 | 0.364009 | 0.018654 | Q9UBS0 | RPS6KB2 | 500 |
| 5 | 2 | 19.53588 | 0.363776 | 0.018621 | Q8N6T3 | ARFGAP1 | 501 |
| 55 | 9 | 26.40312 | 0.491071 | 0.018599 | O00194 | RAB27B | 502 |
| 3 | 1 | 18.94649 | 0.351384 | 0.018546 | Q9H1E5 | TMX4 | 503 |
| 25 | 5 | 20.95949 | 0.388488 | 0.018535 | Q96FZ7 | CHMP6 | 504 |
| 19 | 5 | 22.36762 | 0.414051 | 0.018511 | Q9BWP8 | COLEC11 | 505 |
| 1 | 1 | 17.94437 | 0.332059 | 0.018505 | O15027 | SEC16A | 506 |
| 4 | 1 | 21.67593 | 0.400801 | 0.018491 | Q9UHK6 | AMACR | 507 |
| 14 | 4 | 20.54333 | 0.379272 | 0.018462 | P55212 | CASP6 | 508 |
| 2 | 2 | 16.27361 | 0.300406 | 0.01846 | Q9UP83 | COG5 | 509 |
| 22 | 10 | 23.60377 | 0.435469 | 0.018449 | Q9P270 | SLAIN2 | 510 |
| 5 | 1 | 20.04115 | 0.369462 | 0.018435 | P04156 | PRNP | 511 |
| 12 | 7 | 18.78602 | 0.346115 | 0.018424 | P48147 | PREP | 512 |
| 9 | 4 | 19.55625 | 0.360292 | 0.018423 | P49758 | RGS6 | 513 |
| 6 | 8 | 18.03613 | 0.33212 | 0.018414 | Q15075 | EEA1 | 514 |

| | | | | | | | |
|---|---|---|---|---|---|---|---|
| 15 | 5 | 21.32754 | 0.392413 | 0.018399 | P31689 | DNAJA1 | 515 |
| 1 | 4 | 17.99573 | 0.329589 | 0.018315 | Q8NEZ4 | KMT2C | 516 |
| 1 | 1 | 17.69003 | 0.323251 | 0.018273 | O60266 | ADCY3 | 517 |
| 2 | 1 | 18.75029 | 0.342574 | 0.01827 | Q15904 | ATP6AP1 | 518 |
| 2 | 2 | 24.33041 | 0.442432 | 0.018184 | Q8TDI8 | TMC1 | 519 |
| 6 | 3 | 21.90369 | 0.39622 | 0.018089 | Q96M27 | PRRC1 | 520 |
| 11 | 9 | 21.20453 | 0.381965 | 0.018013 | Q9Y2L6 | FRMD4B | 521 |
| 26 | 1 | 18.79485 | 0.338465 | 0.018008 | Q9NR12 | PDLIM7 | 522 |
| 30 | 3 | 17.99674 | 0.323696 | 0.017986 | P69891 | HBG1 | 523 |
| 10 | 1 | 20.37152 | 0.366288 | 0.01798 | Q9H098 | FAM107B | 524 |
| 9 | 2 | 20.68682 | 0.371715 | 0.017969 | P30536 | TSPO | 525 |
| 8 | 3 | 20.09749 | 0.361109 | 0.017968 | P00749 | PLAU | 526 |
| 4 | 1 | 19.52823 | 0.350737 | 0.017961 | O75663 | TIPRL | 527 |
| 36 | 13 | 22.56326 | 0.404958 | 0.017948 | O75874 | IDH1 | 528 |
| 17 | 4 | 18.16193 | 0.325873 | 0.017943 | P29966 | MARCKS | 529 |
| 5 | 1 | 20.69205 | 0.371059 | 0.017932 | O75489 | NDUFS3 | 530 |
| 10 | 1 | 17.92909 | 0.321501 | 0.017932 | Q9H1E1 | RNASE7 | 531 |
| 15 | 3 | 20.038 | 0.357511 | 0.017842 | Q7Z7H5 | TMED4 | 532 |
| 2 | 6 | 25.10997 | 0.447872 | 0.017836 | Q92794 | KAT6A | 533 |
| 10 | 3 | 21.52912 | 0.383975 | 0.017835 | Q9H8H3 | METTL7A | 534 |
| 7 | 2 | 23.2163 | 0.412275 | 0.017758 | Q6ICL3 | TANGO2 | 535 |
| 67 | 5 | 22.48206 | 0.398926 | 0.017744 | P02042 | HBD | 536 |
| 22 | 5 | 21.85671 | 0.387765 | 0.017741 | Q8TEA8 | DTD1 | 537 |
| 20 | 3 | 21.43329 | 0.378333 | 0.017652 | A0A075B6 | IGLV8-61 | 538 |
| 16 | 4 | 22.4391 | 0.394761 | 0.017593 | P20645 | M6PR | 539 |
| 24 | 4 | 23.29761 | 0.409165 | 0.017563 | Q14203 | DCTN1 | 540 |
| 31 | 5 | 19.79457 | 0.347375 | 0.017549 | P80723 | BASP1 | 541 |
| 8 | 7 | 23.66954 | 0.415225 | 0.017543 | Q9UHB6 | LIMA1 | 542 |
| 22 | 2 | 19.90214 | 0.349131 | 0.017542 | P01743 | IGHV1-46 | 543 |
| 25 | 5 | 21.36871 | 0.374436 | 0.017523 | P28072 | PSMB6 | 544 |
| 9 | 3 | 24.69729 | 0.432311 | 0.017504 | P48509 | CD151 | 545 |
| 12 | 5 | 22.33124 | 0.390707 | 0.017496 | P12955 | PEPD | 546 |
| 12 | 3 | 20.62663 | 0.360829 | 0.017493 | Q9GZY8 | MFF | 547 |
| 37 | 5 | 23.10249 | 0.403101 | 0.017448 | Q96A00 | PPP1R14A | 548 |
| 4 | 4 | 19.84343 | 0.346147 | 0.017444 | A1A4S6 | ARHGAP10 | 549 |
| 19 | 4 | 21.00625 | 0.365733 | 0.017411 | Q9HCN6 | GP6 | 550 |
| 22 | 4 | 22.18901 | 0.386314 | 0.01741 | P30085 | CMPK1 | 551 |
| 19 | 3 | 25.04795 | 0.435742 | 0.017396 | P06703 | S100A6 | 552 |
| 38 | 21 | 25.42121 | 0.441797 | 0.017379 | P11413 | G6PD | 553 |
| 77 | 7 | 24.8324 | 0.431518 | 0.017377 | P01040 | CSTA | 554 |
| 3 | 1 | 18.28726 | 0.317631 | 0.017369 | P50579 | METAP2 | 555 |
| 2 | 1 | 19.39853 | 0.336382 | 0.017341 | Q8TE85 | GRHL3 | 556 |
| 18 | 7 | 21.43035 | 0.371364 | 0.017329 | Q9Y6N5 | SQOR | 557 |
| 3 | 1 | 19.01462 | 0.329007 | 0.017303 | P21731 | TBXA2R | 558 |
| 3 | 1 | 17.89575 | 0.309215 | 0.017279 | O95497 | VNN1 | 559 |
| 20 | 1 | 20.89361 | 0.360916 | 0.017274 | Q99719 | SEPTIN5 | 560 |
| 8 | 7 | 21.75563 | 0.375637 | 0.017266 | Q4V328 | GRIPAP1 | 561 |

| | | | | | | | |
|---|---|---|---|---|---|---|---|
| 25 | 3 | 18.60699 | 0.320745 | 0.017238 | O75368 | SH3BGRL | 562 |
| 11 | 3 | 20.54016 | 0.353786 | 0.017224 | O14579 | COPE | 563 |
| 16 | 6 | 24.81163 | 0.426426 | 0.017187 | O00232 | PSMD12 | 564 |
| 1 | 2 | 20.77743 | 0.356685 | 0.017167 | O94966 | USP19 | 565 |
| 5 | 2 | 19.03347 | 0.326707 | 0.017165 | Q5VZF2 | MBNL2 | 566 |
| 26 | 5 | 24.07318 | 0.412996 | 0.017156 | Q9UFN0 | NIPSNAP3A | 567 |
| 30 | 5 | 22.29106 | 0.381875 | 0.017131 | P59998 | ARPC4 | 568 |
| 8 | 4 | 20.41741 | 0.34972 | 0.017129 | Q9NSD9 | FARSB | 569 |
| 13 | 7 | 19.72342 | 0.337273 | 0.0171 | Q8WWZ8 | OIT3 | 570 |
| 15 | 6 | 21.72903 | 0.370122 | 0.017034 | Q5VW32 | BROX | 571 |
| 27 | 3 | 22.60154 | 0.384845 | 0.017027 | P06331 | IGHV4-34 | 572 |
| 17 | 7 | 21.34372 | 0.362634 | 0.01699 | P52594 | AGFG1 | 573 |
| 15 | 7 | 21.60928 | 0.366439 | 0.016957 | P80303 | NUCB2 | 574 |
| 11 | 5 | 21.14532 | 0.357097 | 0.016888 | Q96S66 | CLCC1 | 575 |
| 23 | 1 | 22.59956 | 0.381401 | 0.016876 | P62879 | GNB2 | 576 |
| 8 | 3 | 19.39631 | 0.327322 | 0.016875 | O95319 | CELF2 | 577 |
| 26 | 4 | 20.39344 | 0.343995 | 0.016868 | Q96E17 | RAB3C | 578 |
| 22 | 3 | 22.80625 | 0.384216 | 0.016847 | P14174 | MIF | 579 |
| 14 | 2 | 22.38972 | 0.377083 | 0.016842 | P30049 | ATP5F1D | 580 |
| 40 | 5 | 21.11577 | 0.353884 | 0.016759 | P28161 | GSTM2 | 581 |
| 20 | 2 | 25.94121 | 0.4346 | 0.016753 | P09669 | COX6C | 582 |
| 18 | 7 | 21.8302 | 0.365474 | 0.016742 | Q8TD55 | PLEKHO2 | 583 |
| 4 | 1 | 19.22839 | 0.321705 | 0.016731 | O75439 | PMPCB | 584 |
| 38 | 11 | 23.00826 | 0.384151 | 0.016696 | P36959 | GMPR | 585 |
| 17 | 17 | 25.33605 | 0.422303 | 0.016668 | P17301 | ITGA2 | 586 |
| 40 | 5 | 23.56873 | 0.392418 | 0.01665 | Q8WWA1 | TMEM40 | 587 |
| 6 | 2 | 19.73162 | 0.328332 | 0.01664 | Q14232 | EIF2B1 | 588 |
| 29 | 11 | 22.84388 | 0.379876 | 0.016629 | P43034 | PAFAH1B1 | 589 |
| 8 | 6 | 20.82707 | 0.346045 | 0.016615 | Q15436 | SEC23A | 590 |
| 20 | 3 | 20.58209 | 0.341384 | 0.016586 | O15173 | PGRMC2 | 591 |
| 2 | 1 | 20.7526 | 0.344017 | 0.016577 | Q15006 | EMC2 | 592 |
| 4 | 1 | 19.12013 | 0.316535 | 0.016555 | Q9UNE7 | STUB1 | 593 |
| 19 | 5 | 21.65022 | 0.358016 | 0.016536 | P53801 | PTTG1IP | 594 |
| 3 | 1 | 18.64132 | 0.307972 | 0.016521 | Q07108 | CD69 | 595 |
| 4 | 3 | 20.22259 | 0.33373 | 0.016503 | O95373 | IPO7 | 596 |
| 22 | 5 | 22.58983 | 0.372793 | 0.016503 | P02753 | RBP4 | 597 |
| 10 | 4 | 22.5773 | 0.372445 | 0.016496 | P35580 | MYH10 | 598 |
| 18 | 22 | 23.82655 | 0.392646 | 0.016479 | P53621 | COPA | 599 |
| 4 | 3 | 21.93008 | 0.361107 | 0.016466 | Q96SU4 | OSBPL9 | 600 |
| 16 | 5 | 22.47571 | 0.369968 | 0.016461 | P20042 | EIF2S2 | 601 |
| 3 | 1 | 18.95513 | 0.311753 | 0.016447 | Q13501 | SQSTM1 | 602 |
| 35 | 3 | 24.9843 | 0.410331 | 0.016424 | A0A0B4J1V | IGHV3-15 | 603 |
| 15 | 8 | 22.3971 | 0.367599 | 0.016413 | Q9UK55 | SERPINA10 | 604 |
| 40 | 8 | 22.54588 | 0.37001 | 0.016411 | O15400 | STX7 | 605 |
| 28 | 7 | 21.65002 | 0.355171 | 0.016405 | P28062 | PSMB8 | 606 |
| 2 | 2 | 17.81775 | 0.291887 | 0.016382 | Q8N766 | EMC1 | 607 |
| 43 | 2 | 20.7393 | 0.33951 | 0.01637 | P35326 | SPRR2A | 608 |

| | | | | | | | |
|---|---|---|---|---|---|---|---|
| 1 | 2 | 19.08456 | 0.312245 | 0.016361 | Q6P996 | PDXDC1 | 609 |
| 14 | 8 | 21.94445 | 0.35844 | 0.016334 | Q86TP1 | PRUNE1 | 610 |
| 6 | 3 | 21.52679 | 0.350661 | 0.01629 | Q96JJ7 | TMX3 | 611 |
| 15 | 7 | 21.62303 | 0.351529 | 0.016257 | P04424 | ASL | 612 |
| 33 | 7 | 24.23301 | 0.392933 | 0.016215 | Q9NY65 | TUBA8 | 613 |
| 36 | 10 | 24.60082 | 0.39832 | 0.016191 | Q14141 | SEPTIN6 | 614 |
| 9 | 13 | 22.90671 | 0.370686 | 0.016182 | Q5D862 | FLG2 | 615 |
| 10 | 5 | 20.09291 | 0.325004 | 0.016175 | O95260 | ATE1 | 616 |
| 38 | 11 | 23.64437 | 0.380416 | 0.016089 | P50402 | EMD | 617 |
| 20 | 4 | 20.55978 | 0.330704 | 0.016085 | Q96BS2 | TESC | 618 |
| 13 | 2 | 19.86556 | 0.319494 | 0.016083 | O95989 | NUDT3 | 619 |
| 4 | 2 | 21.38745 | 0.343588 | 0.016065 | Q96A33 | CCDC47 | 620 |
| 6 | 2 | 20.07052 | 0.322325 | 0.01606 | Q16186 | ADRM1 | 621 |
| 34 | 3 | 21.29347 | 0.341554 | 0.01604 | P04080 | CSTB | 622 |
| 41 | 23 | 29.52023 | 0.47268 | 0.016012 | P13645 | KRT10 | 623 |
| 25 | 4 | 25.01914 | 0.400563 | 0.01601 | P23142 | FBLN1 | 624 |
| 47 | 19 | 25.85511 | 0.413782 | 0.016004 | Q08495 | DMTN | 625 |
| 4 | 1 | 17.64578 | 0.28229 | 0.015998 | Q68D91 | MBLAC2 | 626 |
| 22 | 5 | 18.24641 | 0.29103 | 0.01595 | P23919 | DTYMK | 627 |
| 7 | 3 | 19.60962 | 0.312576 | 0.01594 | Q9UJW0 | DCTN4 | 628 |
| 3 | 1 | 19.38233 | 0.308576 | 0.01592 | Q96HD1 | CRELD1 | 629 |
| 15 | 3 | 21.40022 | 0.340133 | 0.015894 | Q9NZT1 | CALML5 | 630 |
| 22 | 8 | 21.80391 | 0.346357 | 0.015885 | P05198 | EIF2S1 | 631 |
| 2 | 1 | 17.01128 | 0.26982 | 0.015861 | Q9HC07 | TMEM165 | 632 |
| 14 | 1 | 19.17716 | 0.30369 | 0.015836 | P04430 | IGKV1-16 | 633 |
| 29 | 5 | 21.71635 | 0.343464 | 0.015816 | P61081 | UBE2M | 634 |
| 4 | 2 | 18.93439 | 0.299036 | 0.015793 | Q9BYM8 | RBCK1 | 635 |
| 5 | 2 | 19.3016 | 0.303992 | 0.01575 | P05121 | SERPINE1 | 636 |
| 24 | 4 | 22.08869 | 0.346889 | 0.015704 | O96000 | NDUFB10 | 637 |
| 21 | 1 | 19.61244 | 0.307987 | 0.015704 | P04921 | GYPC | 638 |
| 17 | 9 | 23.15796 | 0.362156 | 0.015639 | P01008 | SERPINC1 | 639 |
| 21 | 4 | 21.83021 | 0.341051 | 0.015623 | P13747 | HLA-E | 640 |
| 2 | 1 | 17.75234 | 0.276479 | 0.015574 | Q9Y305 | ACOT9 | 641 |
| 20 | 4 | 23.28819 | 0.362493 | 0.015566 | P62136 | PPP1CA | 642 |
| 9 | 3 | 20.26814 | 0.315476 | 0.015565 | O75787 | ATP6AP2 | 643 |
| 14 | 9 | 21.49958 | 0.333901 | 0.015531 | Q5JSP0 | FGD3 | 644 |
| 3 | 1 | 17.99237 | 0.279393 | 0.015528 | P31641 | SLC6A6 | 645 |
| 12 | 8 | 22.96397 | 0.356577 | 0.015528 | Q02108 | GUCY1A1 | 646 |
| 62 | 2 | 23.27022 | 0.361306 | 0.015527 | P0CF74 | IGLC6 | 647 |
| 5 | 3 | 19.24135 | 0.297136 | 0.015443 | Q9Y5X1 | SNX9 | 648 |
| 10 | 5 | 18.58155 | 0.286222 | 0.015404 | P56181 | NDUFV3 | 649 |
| 17 | 7 | 22.28208 | 0.342687 | 0.015379 | O75695 | RP2 | 650 |
| 23 | 3 | 19.14972 | 0.29431 | 0.015369 | Q8WUD1 | RAB2B | 651 |
| 12 | 15 | 21.52583 | 0.330494 | 0.015353 | Q9BZF9 | UACA | 652 |
| 24 | 8 | 22.47314 | 0.344571 | 0.015333 | Q13043 | STK4 | 653 |
| 57 | 4 | 24.98762 | 0.38297 | 0.015326 | P63167 | DYNLL1 | 654 |
| 3 | 1 | 20.44366 | 0.311648 | 0.015244 | Q8IVE0 | CROCCP3 | 655 |

| | | | | | | | |
|---|---|---|---|---|---|---|---|
| 19 | 4 | 23.22304 | 0.353791 | 0.015234 | Q9HB71 | CACYBP | 656 |
| 12 | 11 | 20.52361 | 0.312544 | 0.015229 | Q99460 | PSMD1 | 657 |
| 66 | 32 | 29.20389 | 0.444248 | 0.015212 | P35908 | KRT2 | 658 |
| 54 | 7 | 22.15046 | 0.336663 | 0.015199 | Q01469 | FABP5 | 659 |
| 6 | 4 | 21.01149 | 0.31925 | 0.015194 | P20742 | PZP | 660 |
| 22 | 12 | 25.23261 | 0.383372 | 0.015194 | P05771 | PRKCB | 661 |
| 21 | 5 | 20.26419 | 0.307211 | 0.01516 | Q9NQR4 | NIT2 | 662 |
| 7 | 3 | 22.45074 | 0.339821 | 0.015136 | Q99442 | SEC62 | 663 |
| 2 | 1 | 19.02154 | 0.286029 | 0.015037 | Q8ND56 | LSM14A | 664 |
| 35 | 2 | 21.80042 | 0.327086 | 0.015004 | A0A0C4DH | IGHV5-51 | 665 |
| 14 | 14 | 23.90158 | 0.35823 | 0.014988 | P14543 | NID1 | 666 |
| 8 | 4 | 21.52058 | 0.321965 | 0.014961 | Q6XQN6 | NAPRT | 667 |
| 34 | 59 | 27.14954 | 0.405922 | 0.014951 | Q13576 | IQGAP2 | 668 |
| 3 | 1 | 18.81438 | 0.281116 | 0.014942 | P15907 | ST6GAL1 | 669 |
| 28 | 4 | 24.42434 | 0.364356 | 0.014918 | Q15771 | RAB30 | 670 |
| 9 | 2 | 18.53555 | 0.276462 | 0.014915 | P13861 | PRKAR2A | 671 |
| 9 | 2 | 21.39443 | 0.318812 | 0.014902 | Q86Y82 | STX12 | 672 |
| 10 | 6 | 20.05387 | 0.298728 | 0.014896 | Q9NRW7 | VPS45 | 673 |
| 7 | 11 | 23.22355 | 0.345772 | 0.014889 | Q5SW79 | CEP170 | 674 |
| 11 | 3 | 22.32256 | 0.331631 | 0.014856 | Q15631 | TSN | 675 |
| 15 | 3 | 21.17514 | 0.314096 | 0.014833 | P61077 | UBE2D3 | 676 |
| 5 | 2 | 18.88185 | 0.279645 | 0.01481 | P17174 | GOT1 | 677 |
| 4 | 1 | 19.60307 | 0.290184 | 0.014803 | P19256 | CD58 | 678 |
| 2 | 1 | 18.14123 | 0.267545 | 0.014748 | Q16534 | HLF | 679 |
| 19 | 3 | 22.543 | 0.332282 | 0.01474 | P13693 | TPT1 | 680 |
| 24 | 5 | 21.365 | 0.314879 | 0.014738 | P25942 | CD40 | 681 |
| 11 | 6 | 21.59436 | 0.318027 | 0.014727 | Q9UEY8 | ADD3 | 682 |
| 7 | 2 | 16.89352 | 0.248479 | 0.014709 | P78356 | PIP4K2B | 683 |
| 3 | 1 | 19.83897 | 0.29156 | 0.014696 | Q12765 | SCRN1 | 684 |
| 11 | 1 | 19.13003 | 0.280673 | 0.014672 | O43676 | NDUFB3 | 685 |
| 14 | 4 | 22.60454 | 0.331547 | 0.014667 | P53007 | SLC25A1 | 686 |
| 4 | 1 | 18.85029 | 0.276305 | 0.014658 | O75381 | PEX14 | 687 |
| 12 | 11 | 20.89552 | 0.305865 | 0.014638 | P27816 | MAP4 | 688 |
| 3 | 10 | 23.8247 | 0.34847 | 0.014626 | Q02224 | CENPE | 689 |
| 14 | 9 | 23.44816 | 0.342619 | 0.014612 | O00534 | VWA5A | 690 |
| 7 | 3 | 19.04597 | 0.278059 | 0.014599 | O43493 | TGOLN2 | 691 |
| 3 | 1 | 17.69773 | 0.258308 | 0.014596 | P35222 | CTNNB1 | 692 |
| 2 | 3 | 19.88928 | 0.289827 | 0.014572 | Q8TEW0 | PARD3 | 693 |
| 18 | 11 | 23.14294 | 0.336519 | 0.014541 | P04196 | HRG | 694 |
| 6 | 5 | 21.29448 | 0.309346 | 0.014527 | P55884 | EIF3B | 695 |
| 19 | 9 | 23.33407 | 0.338722 | 0.014516 | P42768 | WAS | 696 |
| 13 | 3 | 23.54708 | 0.341655 | 0.014509 | Q9NX76 | CMTM6 | 697 |
| 25 | 6 | 23.4944 | 0.339956 | 0.01447 | P48739 | PITPNB | 698 |
| 0 | 1 | 19.07771 | 0.276 | 0.014467 | Q13315 | ATM | 699 |
| 28 | 4 | 24.97792 | 0.360486 | 0.014432 | P55056 | APOC4 | 700 |
| 39 | 5 | 25.10761 | 0.362019 | 0.014419 | O43665 | RGS10 | 701 |
| 6 | 1 | 20.07016 | 0.289122 | 0.014406 | P61923 | COPZ1 | 702 |

| | | | | | | | |
|---|---|---|---|---|---|---|---|
| 18 | 7 | 24.59518 | 0.353794 | 0.014385 | P50570 | DNM2 | 703 |
| 11 | 3 | 23.42676 | 0.335877 | 0.014337 | Q04323 | UBXN1 | 704 |
| 8 | 3 | 20.87868 | 0.298169 | 0.014281 | Q99816 | TSG101 | 705 |
| 7 | 6 | 22.44872 | 0.320441 | 0.014274 | Q05397 | PTK2 | 706 |
| 20 | 5 | 22.58467 | 0.322267 | 0.014269 | Q7Z4W1 | DCXR | 707 |
| 9 | 7 | 20.49 | 0.291942 | 0.014248 | Q03518 | TAP1 | 708 |
| 8 | 3 | 17.7079 | 0.251371 | 0.014195 | P10645 | CHGA | 709 |
| 4 | 1 | 21.06116 | 0.298563 | 0.014176 | O15260 | SURF4 | 710 |
| 7 | 2 | 20.44846 | 0.289586 | 0.014162 | Q8NG11 | TSPAN14 | 711 |
| 26 | 6 | 22.07674 | 0.311799 | 0.014123 | O75935 | DCTN3 | 712 |
| 20 | 6 | 20.00295 | 0.282166 | 0.014106 | Q8N4L2 | PIP4P2 | 713 |
| 4 | 1 | 17.87507 | 0.252034 | 0.0141 | Q8WUY1 | THEM6 | 714 |
| 2 | 1 | 19.85041 | 0.279867 | 0.014099 | Q14587 | ZNF268 | 715 |
| 20 | 6 | 21.62332 | 0.304853 | 0.014098 | Q92820 | GGH | 716 |
| 5 | 1 | 18.5897 | 0.261904 | 0.014089 | Q9Y6M9 | NDUFB9 | 717 |
| 9 | 5 | 20.27947 | 0.285243 | 0.014066 | P31751 | AKT2 | 718 |
| 21 | 3 | 21.40596 | 0.301013 | 0.014062 | P62330 | ARF6 | 719 |
| 12 | 5 | 20.41087 | 0.286775 | 0.01405 | Q13409 | DYNC1I2 | 720 |
| 29 | 15 | 24.23112 | 0.338843 | 0.013984 | Q7Z794 | KRT77 | 721 |
| 7 | 2 | 22.30305 | 0.311553 | 0.013969 | P27701 | CD82 | 722 |
| 18 | 8 | 21.46743 | 0.299382 | 0.013946 | P35241 | RDX | 723 |
| 21 | 11 | 25.35108 | 0.353374 | 0.013939 | P05155 | SERPING1 | 724 |
| 33 | 6 | 22.95711 | 0.319808 | 0.013931 | Q9NP72 | RAB18 | 725 |
| 19 | 4 | 22.69604 | 0.315666 | 0.013908 | P51809 | VAMP7 | 726 |
| 47 | 4 | 21.94691 | 0.303378 | 0.013823 | P99999 | CYCS | 727 |
| 18 | 4 | 21.77418 | 0.300483 | 0.0138 | Q5VWZ2 | LYPLAL1 | 728 |
| 10 | 8 | 20.64316 | 0.284355 | 0.013775 | P21399 | ACO1 | 729 |
| 5 | 3 | 18.43129 | 0.253694 | 0.013764 | O95630 | STAMBP | 730 |
| 17 | 7 | 22.81173 | 0.313344 | 0.013736 | P15529 | CD46 | 731 |
| 25 | 9 | 23.09776 | 0.317072 | 0.013727 | Q9NV96 | TMEM30A | 732 |
| 18 | 2 | 22.67568 | 0.310209 | 0.01368 | A0A075B6 | IGLV4-69 | 733 |
| 18 | 5 | 20.23358 | 0.276427 | 0.013662 | Q9H7J1 | PPP1R3E | 734 |
| 37 | 3 | 21.29634 | 0.290156 | 0.013625 | Q9H4X1 | RGCC | 735 |
| 5 | 4 | 20.81573 | 0.283183 | 0.013604 | O75154 | RAB11FIP3 | 736 |
| 7 | 6 | 21.50004 | 0.29173 | 0.013569 | O14964 | HGS | 737 |
| 20 | 3 | 20.70805 | 0.279166 | 0.013481 | Q9BX68 | HINT2 | 738 |
| 2 | 1 | 20.0485 | 0.270163 | 0.013475 | P34913 | EPHX2 | 739 |
| 7 | 1 | 19.42539 | 0.261394 | 0.013456 | Q9BRA2 | TXNDC17 | 740 |
| 8 | 4 | 21.03573 | 0.282864 | 0.013447 | Q8N1G4 | LRRC47 | 741 |
| 20 | 9 | 22.61649 | 0.303486 | 0.013419 | P50995 | ANXA11 | 742 |
| 6 | 4 | 20.61239 | 0.27602 | 0.013391 | Q8WWI5 | SLC44A1 | 743 |
| 25 | 1 | 21.21559 | 0.283039 | 0.013341 | P01780 | IGHV3-7 | 744 |
| 4 | 3 | 22.94095 | 0.305809 | 0.01333 | Q9Y4K3 | TRAF6 | 745 |
| 22 | 4 | 23.15596 | 0.30809 | 0.013305 | P19652 | ORM2 | 746 |
| 10 | 10 | 24.01681 | 0.31952 | 0.013304 | Q4KMQ2 | ANO6 | 747 |
| 40 | 7 | 24.03429 | 0.319212 | 0.013282 | P18669 | PGAM1 | 748 |
| 12 | 2 | 22.08942 | 0.293097 | 0.013269 | P49720 | PSMB3 | 749 |

| | | | | | | | |
|---|---|---|---|---|---|---|---|
| 17 | 2 | 20.82711 | 0.276262 | 0.013265 | P03973 | SLPI | 750 |
| 19 | 14 | 24.62695 | 0.326533 | 0.013259 | P10643 | C7 | 751 |
| 3 | 1 | 19.44006 | 0.25755 | 0.013248 | Q12982 | BNIP2 | 752 |
| 25 | 8 | 23.98279 | 0.317505 | 0.013239 | P31323 | PRKAR2B | 753 |
| 8 | 3 | 18.63154 | 0.246153 | 0.013212 | O60684 | KPNA6 | 754 |
| 14 | 3 | 19.71226 | 0.25955 | 0.013167 | P57729 | RAB38 | 755 |
| 13 | 8 | 21.14871 | 0.278458 | 0.013167 | Q8WVM8 | SCFD1 | 756 |
| 0 | 1 | 20.65 | 0.271811 | 0.013163 | Q16787 | LAMA3 | 757 |
| 12 | 5 | 23.00721 | 0.302818 | 0.013162 | P49821 | NDUFV1 | 758 |
| 21 | 10 | 20.70678 | 0.271586 | 0.013116 | P31939 | ATIC | 759 |
| 4 | 7 | 27.67247 | 0.362682 | 0.013106 | O75533 | SF3B1 | 760 |
| 4 | 2 | 21.86192 | 0.286243 | 0.013093 | Q9C037 | TRIM4 | 761 |
| 17 | 2 | 21.1231 | 0.276473 | 0.013089 | P61970 | NUTF2 | 762 |
| 13 | 7 | 22.05571 | 0.288465 | 0.013079 | Q02252 | ALDH6A1 | 763 |
| 62 | 26 | 28.87839 | 0.377531 | 0.013073 | P35527 | KRT9 | 764 |
| 10 | 5 | 23.87287 | 0.311671 | 0.013055 | P23381 | WARS1 | 765 |
| 6 | 1 | 18.75013 | 0.244501 | 0.01304 | P52815 | MRPL12 | 766 |
| 4 | 2 | 18.74302 | 0.244373 | 0.013038 | Q2M3D2 | EXOC3L2 | 767 |
| 14 | 8 | 21.3805 | 0.27773 | 0.01299 | O75410 | TACC1 | 768 |
| 38 | 5 | 22.8325 | 0.2964 | 0.012981 | Q13404 | UBE2V1 | 769 |
| 6 | 1 | 20.8646 | 0.27034 | 0.012957 | O43561 | LAT | 770 |
| 35 | 17 | 24.24644 | 0.313808 | 0.012942 | Q99832 | CCT7 | 771 |
| 4 | 1 | 28.00473 | 0.361403 | 0.012905 | Q9H0W7 | THAP2 | 772 |
| 3 | 1 | 18.7794 | 0.242191 | 0.012897 | P0C7M8 | CLEC2L | 773 |
| 0 | 1 | 20.3462 | 0.262277 | 0.012891 | Q6ZTR5 | CFAP47 | 774 |
| 16 | 15 | 23.89974 | 0.307563 | 0.012869 | Q9NZ08 | ERAP1 | 775 |
| 17 | 16 | 25.41951 | 0.326525 | 0.012845 | Q86YZ3 | HRNR | 776 |
| 2 | 1 | 18.60523 | 0.238519 | 0.01282 | P11182 | DBT | 777 |
| 12 | 4 | 18.95761 | 0.242972 | 0.012817 | P11217 | PYGM | 778 |
| 25 | 2 | 21.72349 | 0.277774 | 0.012787 | Q9UJC5 | SH3BGRL2 | 779 |
| 25 | 5 | 21.8976 | 0.279762 | 0.012776 | Q9UL46 | PSME2 | 780 |
| 26 | 3 | 22.09863 | 0.281888 | 0.012756 | P25398 | RPS12 | 781 |
| 27 | 12 | 23.21103 | 0.295088 | 0.012713 | P19013 | KRT4 | 782 |
| 20 | 3 | 23.14015 | 0.294016 | 0.012706 | Q9NUQ9 | CYRIB | 783 |
| 11 | 2 | 18.35314 | 0.233122 | 0.012702 | P19404 | NDUFV2 | 784 |
| 8 | 7 | 20.19102 | 0.254278 | 0.012594 | P49619 | DGKG | 785 |
| 22 | 5 | 23.18366 | 0.291247 | 0.012563 | Q6UWP8 | SBSN | 786 |
| 3 | 1 | 16.79111 | 0.2108 | 0.012554 | Q8N9M5 | TMEM102 | 787 |
| 13 | 4 | 20.06123 | 0.251744 | 0.012549 | Q92734 | TFG | 788 |
| 6 | 5 | 20.78149 | 0.260557 | 0.012538 | P98082 | DAB2 | 789 |
| 9 | 4 | 21.50621 | 0.269501 | 0.012531 | Q9NVG8 | TBC1D13 | 790 |
| 20 | 7 | 21.65802 | 0.271143 | 0.012519 | P08758 | ANXA5 | 791 |
| 17 | 6 | 22.03133 | 0.274763 | 0.012471 | Q08257 | CRYZ | 792 |
| 27 | 9 | 22.1249 | 0.275218 | 0.012439 | Q16539 | MAPK14 | 793 |
| 3 | 5 | 19.61692 | 0.24285 | 0.01238 | Q14152 | EIF3A | 794 |
| 13 | 14 | 22.87359 | 0.282281 | 0.012341 | Q9H2G2 | SLK | 795 |
| 21 | 11 | 22.46838 | 0.277189 | 0.012337 | P49591 | SARS1 | 796 |

| | | | | | | | |
|---|---|---|---|---|---|---|---|
| 35 | 1 | 27.21286 | 0.33565 | 0.012334 | P02776 | PF4 | 797 |
| 21 | 8 | 24.328 | 0.299804 | 0.012323 | P22694 | PRKACB | 798 |
| 10 | 2 | 20.25334 | 0.248858 | 0.012287 | P61009 | SPCS3 | 799 |
| 13 | 7 | 21.81816 | 0.267717 | 0.01227 | O15523 | DDX3Y | 800 |
| 55 | 2 | 23.8106 | 0.291921 | 0.01226 | P07951 | TPM2 | 801 |
| 18 | 4 | 21.75661 | 0.26586 | 0.01222 | Q99685 | MGLL | 802 |
| 42 | 6 | 26.18645 | 0.319817 | 0.012213 | P35542 | SAA4 | 803 |
| 15 | 2 | 21.69271 | 0.264863 | 0.01221 | O75380 | NDUFS6 | 804 |
| 6 | 4 | 19.65062 | 0.239594 | 0.012193 | Q13496 | MTM1 | 805 |
| 66 | 6 | 21.80068 | 0.265276 | 0.012168 | Q15369 | ELOC | 806 |
| 21 | 8 | 21.94698 | 0.266233 | 0.012131 | P51575 | P2RX1 | 807 |
| 9 | 3 | 19.63473 | 0.238054 | 0.012124 | O95865 | DDAH2 | 808 |
| 13 | 3 | 20.9357 | 0.2537 | 0.012118 | Q12974 | PTP4A2 | 809 |
| 22 | 4 | 20.12726 | 0.243442 | 0.012095 | Q9BY43 | CHMP4A | 810 |
| 24 | 11 | 23.87188 | 0.288285 | 0.012076 | P29401 | TKT | 811 |
| 4 | 4 | 23.67274 | 0.284704 | 0.012027 | P12830 | CDH1 | 812 |
| 15 | 2 | 21.89598 | 0.263151 | 0.012018 | Q9H910 | JPT2 | 813 |
| 1 | 1 | 22.77907 | 0.273398 | 0.012002 | Q8NFP4 | MDGA1 | 814 |
| 6 | 1 | 18.78385 | 0.22487 | 0.011971 | P16298 | PPP3CB | 815 |
| 3 | 1 | 18.20422 | 0.217645 | 0.011956 | P07476 | IVL | 816 |
| 11 | 15 | 23.71447 | 0.283259 | 0.011945 | Q9Y5S2 | CDC42BPB | 817 |
| 12 | 15 | 22.27291 | 0.265823 | 0.011935 | Q9ULH1 | ASAP1 | 818 |
| 49 | 11 | 26.60213 | 0.317474 | 0.011934 | Q9Y277 | VDAC3 | 819 |
| 22 | 9 | 22.94212 | 0.273727 | 0.011931 | O95299 | NDUFA10 | 820 |
| 11 | 3 | 20.67016 | 0.246237 | 0.011913 | Q9H6Q3 | SLA2 | 821 |
| 4 | 2 | 18.40904 | 0.218285 | 0.011857 | P51116 | FXR2 | 822 |
| 25 | 6 | 22.49645 | 0.266649 | 0.011853 | P06241 | FYN | 823 |
| 21 | 7 | 21.52537 | 0.254992 | 0.011846 | Q16543 | CDC37 | 824 |
| 20 | 3 | 22.80014 | 0.26984 | 0.011835 | P59666 | DEFA3 | 825 |
| 1 | 1 | 19.31311 | 0.22812 | 0.011812 | O00471 | EXOC5 | 826 |
| 2 | 1 | 18.86209 | 0.222718 | 0.011808 | Q13557 | CAMK2D | 827 |
| 5 | 1 | 22.53026 | 0.265178 | 0.01177 | Q969X1 | TMBIM1 | 828 |
| 30 | 20 | 25.59849 | 0.30128 | 0.011769 | Q96C24 | SYTL4 | 829 |
| 14 | 3 | 20.50145 | 0.241114 | 0.011761 | P11908 | PRPS2 | 830 |
| 10 | 7 | 21.43757 | 0.251933 | 0.011752 | P18084 | ITGB5 | 831 |
| 12 | 2 | 19.77581 | 0.232214 | 0.011742 | Q9H9B4 | SFXN1 | 832 |
| 8 | 2 | 23.69641 | 0.277268 | 0.011701 | O15127 | SCAMP2 | 833 |
| 3 | 1 | 19.23839 | 0.223907 | 0.011639 | Q9Y3B8 | REXO2 | 834 |
| 21 | 2 | 19.14276 | 0.22277 | 0.011637 | O75531 | BANF1 | 835 |
| 30 | 24 | 27.3529 | 0.317368 | 0.011603 | P05556 | ITGB1 | 836 |
| 9 | 4 | 20.58265 | 0.238518 | 0.011588 | Q12912 | IRAG2 | 837 |
| 11 | 5 | 21.47007 | 0.248611 | 0.011579 | Q15208 | STK38 | 838 |
| 17 | 5 | 21.43193 | 0.248054 | 0.011574 | Q969X5 | ERGIC1 | 839 |
| 7 | 6 | 19.96895 | 0.230963 | 0.011566 | Q96MK2 | RIPOR3 | 840 |
| 23 | 6 | 23.51794 | 0.271759 | 0.011555 | P54709 | ATP1B3 | 841 |
| 24 | 4 | 25.64737 | 0.296331 | 0.011554 | O75947 | ATP5PD | 842 |
| 16 | 31 | 24.65504 | 0.282629 | 0.011463 | Q14008 | CKAP5 | 843 |

| | | | | | | | |
|---|---|---|---|---|---|---|---|
| 32 | 12 | 21.37965 | 0.244569 | 0.011439 | Q12805 | EFEMP1 | 844 |
| 35 | 11 | 23.66543 | 0.270018 | 0.01141 | Q9H4M9 | EHD1 | 845 |
| 11 | 2 | 20.5146 | 0.233329 | 0.011374 | Q96IU4 | ABHD14B | 846 |
| 3 | 2 | 20.52987 | 0.233222 | 0.01136 | Q13443 | ADAM9 | 847 |
| 12 | 1 | 20.54187 | 0.233317 | 0.011358 | P14406 | COX7A2 | 848 |
| 10 | 2 | 19.52158 | 0.221377 | 0.01134 | Q15166 | PON3 | 849 |
| 28 | 5 | 24.3482 | 0.275883 | 0.011331 | P00441 | SOD1 | 850 |
| 12 | 10 | 22.80982 | 0.258356 | 0.011327 | P41226 | UBA7 | 851 |
| 17 | 2 | 25.34987 | 0.286635 | 0.011307 | P06312 | IGKV4-1 | 852 |
| 11 | 9 | 22.99083 | 0.259658 | 0.011294 | Q5T447 | HECTD3 | 853 |
| 10 | 4 | 18.6646 | 0.210722 | 0.01129 | Q9Y3I0 | RTCB | 854 |
| 11 | 12 | 22.66207 | 0.255582 | 0.011278 | P55160 | NCKAP1L | 855 |
| 37 | 6 | 22.21705 | 0.250455 | 0.011273 | Q15366 | PCBP2 | 856 |
| 8 | 4 | 20.81309 | 0.234385 | 0.011261 | Q6NUK1 | SLC25A24 | 857 |
| 1 | 1 | 20.32811 | 0.228656 | 0.011248 | O95359 | TACC2 | 858 |
| 14 | 12 | 23.66474 | 0.265938 | 0.011238 | Q9Y2X7 | GIT1 | 859 |
| 19 | 6 | 23.21722 | 0.260072 | 0.011202 | P35914 | HMGCL | 860 |
| 4 | 2 | 21.87308 | 0.244894 | 0.011196 | P30038 | ALDH4A1 | 861 |
| 18 | 20 | 23.3605 | 0.261488 | 0.011194 | Q86VP6 | CAND1 | 862 |
| 12 | 35 | 24.40178 | 0.272861 | 0.011182 | P15924 | DSP | 863 |
| 9 | 2 | 18.9669 | 0.212076 | 0.011181 | P04085 | PDGFA | 864 |
| 14 | 12 | 23.09685 | 0.2581 | 0.011175 | Q13200 | PSMD2 | 865 |
| 18 | 4 | 21.2759 | 0.237551 | 0.011165 | Q00688 | FKBP3 | 866 |
| 11 | 1 | 18.7574 | 0.209182 | 0.011152 | O75688 | PPM1B | 867 |
| 10 | 6 | 23.03342 | 0.25512 | 0.011076 | P11279 | LAMP1 | 868 |
| 23 | 8 | 22.62937 | 0.250334 | 0.011062 | Q86WR7 | PROSER2 | 869 |
| 17 | 6 | 24.90326 | 0.275485 | 0.011062 | P54727 | RAD23B | 870 |
| 6 | 1 | 19.9521 | 0.220247 | 0.011039 | O00501 | CLDN5 | 871 |
| 4 | 1 | 18.70938 | 0.206499 | 0.011037 | Q8IZ81 | ELMOD2 | 872 |
| 16 | 2 | 19.29232 | 0.212692 | 0.011025 | Q14240 | EIF4A2 | 873 |
| 22 | 17 | 23.5527 | 0.259248 | 0.011007 | P53618 | COPB1 | 874 |
| 63 | 40 | 30.51523 | 0.33541 | 0.010992 | P04264 | KRT1 | 875 |
| 59 | 7 | 24.25751 | 0.26654 | 0.010988 | O75347 | TBCA | 876 |
| 10 | 4 | 21.58059 | 0.236341 | 0.010952 | Q6UX71 | PLXDC2 | 877 |
| 4 | 1 | 20.66529 | 0.226079 | 0.01094 | Q9UKB3 | DNAJC12 | 878 |
| 51 | 4 | 23.22258 | 0.253925 | 0.010934 | Q9BSF0 | C2orf88 | 879 |
| 4 | 3 | 20.53874 | 0.224117 | 0.010912 | Q13617 | CUL2 | 880 |
| 13 | 13 | 20.35131 | 0.22162 | 0.01089 | Q9Y210 | TRPC6 | 881 |
| 33 | 3 | 23.50097 | 0.255904 | 0.010889 | P61020 | RAB5B | 882 |
| 16 | 6 | 19.02318 | 0.206365 | 0.010848 | P17980 | PSMC3 | 883 |
| 8 | 5 | 18.41478 | 0.199721 | 0.010846 | P26599 | PTBP1 | 884 |
| 12 | 5 | 24.01235 | 0.260393 | 0.010844 | Q9BS26 | ERP44 | 885 |
| 16 | 9 | 22.1498 | 0.239753 | 0.010824 | Q9UHQ4 | BCAP29 | 886 |
| 19 | 1 | 19.43967 | 0.210311 | 0.010819 | P0DP04 | IGHV3-43D | 887 |
| 22 | 5 | 22.00706 | 0.237972 | 0.010813 | Q9NX40 | OCIAD1 | 888 |
| 29 | 5 | 21.30114 | 0.230202 | 0.010807 | P16949 | STMN1 | 889 |
| 19 | 5 | 22.74014 | 0.24564 | 0.010802 | P62140 | PPP1CB | 890 |

| | | | | | | | |
|---|---|---|---|---|---|---|---|
| 14 | 5 | 21.1588 | 0.228552 | 0.010802 | P61421 | ATP6V0D1 | 891 |
| 8 | 5 | 22.23426 | 0.240014 | 0.010795 | P63151 | PPP2R2A | 892 |
| 57 | 7 | 23.91654 | 0.257645 | 0.010773 | P62942 | FKBP1A | 893 |
| 20 | 2 | 22.88366 | 0.245916 | 0.010746 | P81605 | DCD | 894 |
| 27 | 10 | 23.80162 | 0.254928 | 0.010711 | P35237 | SERPINB6 | 895 |
| 9 | 6 | 21.06494 | 0.225555 | 0.010708 | P13798 | APEH | 896 |
| 12 | 3 | 21.3217 | 0.228138 | 0.0107 | O95249 | GOSR1 | 897 |
| 42 | 1 | 22.33144 | 0.238846 | 0.010695 | P62820 | RAB1A | 898 |
| 16 | 8 | 22.65158 | 0.241957 | 0.010682 | O60884 | DNAJA2 | 899 |
| 25 | 8 | 23.32066 | 0.247955 | 0.010632 | P61163 | ACTR1A | 900 |
| 49 | 24 | 27.45758 | 0.291791 | 0.010627 | P13647 | KRT5 | 901 |
| 21 | 3 | 21.0664 | 0.223816 | 0.010624 | P24666 | ACP1 | 902 |
| 6 | 4 | 24.26396 | 0.257761 | 0.010623 | Q5T749 | KPRP | 903 |
| 16 | 10 | 21.69656 | 0.229685 | 0.010586 | Q04759 | PRKCQ | 904 |
| 23 | 6 | 21.27348 | 0.224775 | 0.010566 | O00233 | PSMD9 | 905 |
| 34 | 12 | 22.2865 | 0.235452 | 0.010565 | O75955 | FLOT1 | 906 |
| 19 | 2 | 22.27548 | 0.23519 | 0.010558 | Q15286 | RAB35 | 907 |
| 7 | 2 | 19.89482 | 0.20974 | 0.010542 | O15194 | CTDSPL | 908 |
| 1 | 1 | 17.88046 | 0.188457 | 0.01054 | P40818 | USP8 | 909 |
| 11 | 2 | 20.41463 | 0.21499 | 0.010531 | Q99969 | RARRES2 | 910 |
| 28 | 6 | 22.21146 | 0.233853 | 0.010528 | Q9HBH0 | RHOF | 911 |
| 17 | 2 | 22.48574 | 0.235765 | 0.010485 | Q9H4G4 | GLIPR2 | 912 |
| 19 | 5 | 23.55612 | 0.246975 | 0.010485 | P02746 | C1QB | 913 |
| 43 | 5 | 21.41718 | 0.224162 | 0.010466 | Q15370 | ELOB | 914 |
| 6 | 8 | 22.96356 | 0.240252 | 0.010462 | Q8N3D4 | EHBP1L1 | 915 |
| 10 | 2 | 19.36345 | 0.202558 | 0.010461 | Q9UK41 | VPS28 | 916 |
| 40 | 10 | 25.34828 | 0.264938 | 0.010452 | Q15555 | MAPRE2 | 917 |
| 1 | 1 | 20.10876 | 0.210087 | 0.010448 | P17655 | CAPN2 | 918 |
| 9 | 4 | 22.29143 | 0.232884 | 0.010447 | O75844 | ZMPSTE24 | 919 |
| 15 | 2 | 19.83727 | 0.207133 | 0.010442 | P50851 | LRBA | 920 |
| 23 | 6 | 25.20906 | 0.263042 | 0.010434 | Q99733 | NAP1L4 | 921 |
| 7 | 1 | 20.19148 | 0.210386 | 0.01042 | P30046 | DDT | 922 |
| 4 | 1 | 18.77597 | 0.195384 | 0.010406 | P06734 | FCER2 | 923 |
| 19 | 5 | 23.31595 | 0.242576 | 0.010404 | Q9P289 | STK26 | 924 |
| 34 | 6 | 24.95324 | 0.259393 | 0.010395 | P61586 | RHOA | 925 |
| 26 | 10 | 20.42803 | 0.212236 | 0.010389 | P18031 | PTPN1 | 926 |
| 24 | 2 | 21.77644 | 0.225871 | 0.010372 | A0A0C4DH | IGHV1-18 | 927 |
| 26 | 13 | 24.85678 | 0.25763 | 0.010365 | Q9BR76 | CORO1B | 928 |
| 10 | 4 | 20.82058 | 0.21551 | 0.010351 | Q96CW1 | AP2M1 | 929 |
| 43 | 5 | 27.26107 | 0.281942 | 0.010342 | P69905 | HBA1 | 930 |
| 9 | 1 | 20.54773 | 0.212248 | 0.010329 | Q15170 | TCEAL1 | 931 |
| 3 | 2 | 19.63346 | 0.201815 | 0.010279 | O60716 | CTNND1 | 932 |
| 22 | 6 | 22.55411 | 0.231425 | 0.010261 | P21912 | SDHB | 933 |
| 22 | 10 | 23.2043 | 0.237789 | 0.010248 | Q9HDC9 | APMAP | 934 |
| 21 | 7 | 20.29358 | 0.207731 | 0.010236 | Q99747 | NAPG | 935 |
| 4 | 3 | 24.76804 | 0.251907 | 0.010171 | A6NDN3 | GOLGA6B | 936 |
| 21 | 7 | 21.80407 | 0.221655 | 0.010166 | O43765 | SGTA | 937 |

| | | | | | | | |
|---|---|---|---|---|---|---|---|
| 29 | 6 | 24.09589 | 0.244796 | 0.010159 | P49913 | CAMP | 938 |
| 4 | 2 | 19.59371 | 0.198873 | 0.01015 | P48637 | GSS | 939 |
| 8 | 6 | 22.05648 | 0.223754 | 0.010145 | Q03519 | TAP2 | 940 |
| 9 | 2 | 19.1838 | 0.193777 | 0.010101 | O14960 | LECT2 | 941 |
| 1 | 1 | 24.92979 | 0.251347 | 0.010082 | O43155 | FLRT2 | 942 |
| 2 | 2 | 19.20359 | 0.19355 | 0.010079 | O60841 | EIF5B | 943 |
| 9 | 5 | 22.90968 | 0.230603 | 0.010066 | Q04446 | GBE1 | 944 |
| 25 | 10 | 21.23312 | 0.213727 | 0.010066 | O00231 | PSMD11 | 945 |
| 15 | 2 | 21.67438 | 0.217773 | 0.010047 | Q99471 | PFDN5 | 946 |
| 31 | 22 | 25.0302 | 0.251151 | 0.010034 | Q16891 | IMMT | 947 |
| 26 | 10 | 22.17541 | 0.222406 | 0.010029 | P14317 | HCLS1 | 948 |
| 42 | 7 | 25.18471 | 0.252444 | 0.010024 | P60981 | DSTN | 949 |
| 11 | 11 | 23.61368 | 0.236457 | 0.010014 | O00410 | IPO5 | 950 |
| 5 | 1 | 19.53496 | 0.195263 | 0.009996 | Q9BRP8 | PYM1 | 951 |
| 2 | 1 | 18.131 | 0.181179 | 0.009993 | O43304 | SEC14L5 | 952 |
| 9 | 3 | 18.26646 | 0.182462 | 0.009989 | Q9UQ16 | DNM3 | 953 |
| 7 | 7 | 25.26607 | 0.2518 | 0.009966 | Q9UBF2 | COPG2 | 954 |
| 24 | 3 | 20.85566 | 0.206972 | 0.009924 | P84095 | RHOG | 955 |
| 22 | 5 | 22.7114 | 0.224993 | 0.009907 | P42574 | CASP3 | 956 |
| 36 | 11 | 25.56097 | 0.253117 | 0.009902 | P40926 | MDH2 | 957 |
| 16 | 8 | 23.82477 | 0.235734 | 0.009895 | Q9NXH8 | TOR4A | 958 |
| 14 | 5 | 22.85946 | 0.225991 | 0.009886 | Q08209 | PPP3CA | 959 |
| 4 | 3 | 18.69368 | 0.184664 | 0.009878 | O94929 | ABLIM3 | 960 |
| 17 | 5 | 23.68132 | 0.233829 | 0.009874 | P10768 | ESD | 961 |
| 28 | 4 | 22.17403 | 0.218898 | 0.009872 | Q5JQS6 | GCSAML | 962 |
| 8 | 3 | 18.48739 | 0.182074 | 0.009849 | Q9BXN1 | ASPN | 963 |
| 3 | 6 | 20.85813 | 0.205249 | 0.00984 | Q14C86 | GAPVD1 | 964 |
| 22 | 8 | 22.57756 | 0.22215 | 0.009839 | P45954 | ACADSB | 965 |
| 12 | 1 | 21.90051 | 0.215457 | 0.009838 | P27449 | ATP6V0C | 966 |
| 12 | 11 | 22.04112 | 0.216192 | 0.009809 | Q96A65 | EXOC4 | 967 |
| 20 | 2 | 18.95778 | 0.185621 | 0.009791 | Q16627 | CCL14 | 968 |
| 8 | 10 | 22.98159 | 0.224495 | 0.009768 | P32019 | INPP5B | 969 |
| 19 | 8 | 22.94852 | 0.224109 | 0.009766 | Q13867 | BLMH | 970 |
| 40 | 22 | 24.83089 | 0.242268 | 0.009757 | P13667 | PDIA4 | 971 |
| 24 | 30 | 24.46958 | 0.238061 | 0.009729 | Q7L576 | CYFIP1 | 972 |
| 6 | 2 | 20.61075 | 0.200301 | 0.009718 | Q99598 | TSNAX | 973 |
| 28 | 6 | 22.80765 | 0.22145 | 0.009709 | Q9NVK5 | FGFR1OP2 | 974 |
| 3 | 5 | 21.0918 | 0.204625 | 0.009702 | E9PAV3 | NACA | 975 |
| 8 | 3 | 20.25571 | 0.196481 | 0.0097 | P43686 | PSMC4 | 976 |
| 10 | 4 | 19.53548 | 0.189369 | 0.009694 | Q13451 | FKBP5 | 977 |
| 1 | 1 | 18.7213 | 0.181201 | 0.009679 | Q9C0E2 | XPO4 | 978 |
| 14 | 1 | 20.23921 | 0.195886 | 0.009679 | Q9Y2S6 | TMA7 | 979 |
| 2 | 6 | 23.5613 | 0.228035 | 0.009678 | Q8NDH2 | CCDC168 | 980 |
| 14 | 9 | 21.31583 | 0.206073 | 0.009668 | Q96T51 | RUFY1 | 981 |
| 27 | 7 | 24.80163 | 0.239743 | 0.009666 | P30040 | ERP29 | 982 |
| 7 | 2 | 21.88821 | 0.210763 | 0.009629 | Q99436 | PSMB7 | 983 |
| 5 | 7 | 18.8017 | 0.180934 | 0.009623 | P07814 | EPRS1 | 984 |

| | | | | | | | |
|---|---|---|---|---|---|---|---|
| 9 | 18 | 24.17873 | 0.232427 | 0.009613 | Q92614 | MYO18A | 985 |
| 48 | 4 | 24.77661 | 0.237635 | 0.009591 | P22392 | NME2 | 986 |
| 29 | 8 | 26.51742 | 0.254256 | 0.009588 | Q9H444 | CHMP4B | 987 |
| 22 | 5 | 22.65677 | 0.217132 | 0.009584 | Q96HS1 | PGAM5 | 988 |
| 43 | 5 | 23.27585 | 0.22286 | 0.009575 | Q9BV40 | VAMP8 | 989 |
| 9 | 13 | 21.41569 | 0.204822 | 0.009564 | Q9H2M9 | RAB3GAP2 | 990 |
| 23 | 1 | 18.89761 | 0.180613 | 0.009557 | O43852 | CALU | 991 |
| 22 | 7 | 22.5277 | 0.214887 | 0.009539 | P22695 | UQCRC2 | 992 |
| 18 | 8 | 22.31399 | 0.212608 | 0.009528 | P54578 | USP14 | 993 |
| 29 | 5 | 23.31202 | 0.222076 | 0.009526 | Q9P0L0 | VAPA | 994 |
| 25 | 10 | 23.24136 | 0.221047 | 0.009511 | Q93052 | LPP | 995 |
| 2 | 1 | 21.26776 | 0.20213 | 0.009504 | Q9BPX6 | MICU1 | 996 |
| 11 | 12 | 21.14745 | 0.200796 | 0.009495 | Q5M775 | SPECC1 | 997 |
| 9 | 8 | 21.73867 | 0.206074 | 0.00948 | Q9HC35 | EML4 | 998 |
| 11 | 4 | 18.54657 | 0.175793 | 0.009478 | O43813 | LANCL1 | 999 |
| 33 | 3 | 24.08119 | 0.228234 | 0.009478 | P30466 | HLA-B | 1000 |
| 5 | 8 | 19.43879 | 0.18394 | 0.009463 | Q66K14 | TBC1D9B | 1001 |
| 5 | 5 | 20.38291 | 0.192745 | 0.009456 | P14735 | IDE | 1002 |
| 15 | 3 | 20.90326 | 0.197403 | 0.009444 | P07360 | C8G | 1003 |
| 6 | 2 | 22.00337 | 0.207755 | 0.009442 | Q9NZJ7 | MTCH1 | 1004 |
| 9 | 11 | 23.38309 | 0.220656 | 0.009437 | Q14432 | PDE3A | 1005 |
| 32 | 9 | 23.56976 | 0.222279 | 0.009431 | P38117 | ETFB | 1006 |
| 17 | 6 | 22.64875 | 0.213168 | 0.009412 | O60496 | DOK2 | 1007 |
| 5 | 1 | 18.33263 | 0.172544 | 0.009412 | Q9Y237 | PIN4 | 1008 |
| 34 | 9 | 23.22191 | 0.218109 | 0.009392 | P25786 | PSMA1 | 1009 |
| 16 | 10 | 23.923 | 0.223523 | 0.009343 | P0C7P3 | SLFN14 | 1010 |
| 12 | 4 | 22.40113 | 0.209222 | 0.00934 | Q5SQ64 | LY6G6F | 1011 |
| 21 | 20 | 26.84494 | 0.2504 | 0.009328 | Q9Y4L1 | HYOU1 | 1012 |
| 2 | 1 | 19.65448 | 0.183172 | 0.00932 | Q9C035 | TRIM5 | 1013 |
| 17 | 8 | 23.37505 | 0.217756 | 0.009316 | Q8IUI8 | CRLF3 | 1014 |
| 16 | 11 | 21.8616 | 0.20341 | 0.009304 | Q684P5 | RAP1GAP2 | 1015 |
| 14 | 3 | 23.18425 | 0.215017 | 0.009274 | O75954 | TSPAN9 | 1016 |
| 17 | 1 | 22.18189 | 0.205718 | 0.009274 | P01614 | IGKV2D-40 | 1017 |
| 39 | 8 | 23.16226 | 0.214639 | 0.009267 | Q9NS28 | RGS18 | 1018 |
| 29 | 4 | 23.9078 | 0.221273 | 0.009255 | Q9NR31 | SAR1A | 1019 |
| 19 | 1 | 21.70962 | 0.200622 | 0.009241 | P01768 | IGHV3-30 | 1020 |
| 8 | 1 | 19.76208 | 0.18215 | 0.009217 | P61960 | UFM1 | 1021 |
| 29 | 17 | 24.55976 | 0.226339 | 0.009216 | P27338 | MAOB | 1022 |
| 6 | 4 | 22.43662 | 0.206757 | 0.009215 | P18433 | PTPRA | 1023 |
| 11 | 8 | 22.77569 | 0.209836 | 0.009213 | Q9NY33 | DPP3 | 1024 |
| 21 | 10 | 25.08474 | 0.230991 | 0.009208 | P11597 | CETP | 1025 |
| 25 | 1 | 21.54707 | 0.198173 | 0.009197 | A0A0B4J1X | IGHV3-74 | 1026 |
| 9 | 2 | 18.46362 | 0.169682 | 0.00919 | Q14088 | RAB33A | 1027 |
| 7 | 5 | 23.47431 | 0.215096 | 0.009163 | Q16204 | CCDC6 | 1028 |
| 3 | 1 | 19.2737 | 0.17629 | 0.009147 | O14545 | TRAFD1 | 1029 |
| 1 | 1 | 20.89339 | 0.19062 | 0.009123 | O43196 | MSH5 | 1030 |
| 25 | 8 | 22.55902 | 0.205404 | 0.009105 | Q9Y2Z0 | SUGT1 | 1031 |

| | | | | | | | |
|---|---|---|---|---|---|---|---|
| 9 | 8 | 22.4185 | 0.204028 | 0.009101 | P55060 | CSE1L | 1032 |
| 58 | 12 | 26.54475 | 0.24058 | 0.009063 | P52566 | ARHGDIB | 1033 |
| 17 | 3 | 24.6603 | 0.223286 | 0.009054 | P02766 | TTR | 1034 |
| 28 | 6 | 24.53847 | 0.222023 | 0.009048 | O15145 | ARPC3 | 1035 |
| 40 | 8 | 24.24783 | 0.219313 | 0.009045 | P52565 | ARHGDIA | 1036 |
| 3 | 1 | 19.05488 | 0.171974 | 0.009025 | O43681 | GET3 | 1037 |
| 13 | 5 | 24.16356 | 0.217783 | 0.009013 | Q99704 | DOK1 | 1038 |
| 17 | 3 | 20.27045 | 0.182276 | 0.008992 | O43583 | DENR | 1039 |
| 2 | 3 | 24.73497 | 0.222418 | 0.008992 | Q08050 | FOXM1 | 1040 |
| 6 | 7 | 22.13716 | 0.198852 | 0.008983 | P08648 | ITGA5 | 1041 |
| 7 | 3 | 22.06425 | 0.198187 | 0.008982 | P56202 | CTSW | 1042 |
| 5 | 2 | 21.03369 | 0.188781 | 0.008975 | Q9BTV4 | TMEM43 | 1043 |
| 3 | 1 | 20.75866 | 0.185886 | 0.008955 | Q92504 | SLC39A7 | 1044 |
| 54 | 8 | 26.88228 | 0.2404 | 0.008943 | P61981 | YWHAG | 1045 |
| 8 | 1 | 21.00712 | 0.187203 | 0.008911 | P01714 | IGLV3-19 | 1046 |
| 45 | 2 | 25.66869 | 0.227761 | 0.008873 | P68363 | TUBA1B | 1047 |
| 26 | 10 | 24.66496 | 0.218837 | 0.008872 | P48426 | PIP4K2A | 1048 |
| 11 | 6 | 21.57101 | 0.191119 | 0.00886 | Q15067 | ACOX1 | 1049 |
| 11 | 11 | 21.62556 | 0.191403 | 0.008851 | Q6ZU35 | CRACD | 1050 |
| 15 | 7 | 23.64138 | 0.209207 | 0.008849 | P17252 | PRKCA | 1051 |
| 23 | 3 | 22.4438 | 0.198578 | 0.008848 | P61018 | RAB4B | 1052 |
| 6 | 3 | 24.09039 | 0.212943 | 0.008839 | Q9HB21 | PLEKHA1 | 1053 |
| 6 | 4 | 21.69584 | 0.191601 | 0.008831 | Q4KMP7 | TBC1D10B | 1054 |
| 15 | 7 | 21.32749 | 0.188244 | 0.008826 | Q8WXF7 | ATL1 | 1055 |
| 21 | 2 | 21.36087 | 0.188518 | 0.008825 | P10599 | TXN | 1056 |
| 1 | 1 | 17.60353 | 0.15526 | 0.00882 | B1AK53 | ESPN | 1057 |
| 4 | 3 | 18.36159 | 0.161758 | 0.00881 | O43432 | EIF4G3 | 1058 |
| 14 | 5 | 22.4005 | 0.197326 | 0.008809 | Q15008 | PSMD6 | 1059 |
| 9 | 3 | 19.18474 | 0.168717 | 0.008794 | Q96IJ6 | GMPPA | 1060 |
| 20 | 11 | 22.70281 | 0.199001 | 0.008765 | Q9UHD8 | SEPTIN9 | 1061 |
| 8 | 5 | 23.99543 | 0.21021 | 0.00876 | O95197 | RTN3 | 1062 |
| 6 | 4 | 20.9418 | 0.183016 | 0.008739 | O00391 | QSOX1 | 1063 |
| 13 | 1 | 19.71856 | 0.172056 | 0.008726 | Q9P2X0 | DPM3 | 1064 |
| 12 | 2 | 25.93903 | 0.225681 | 0.0087 | P11169 | SLC2A3 | 1065 |
| 21 | 1 | 19.76014 | 0.171731 | 0.008691 | P20339 | RAB5A | 1066 |
| 24 | 7 | 22.99841 | 0.199735 | 0.008685 | P28482 | MAPK1 | 1067 |
| 40 | 8 | 25.60581 | 0.222124 | 0.008675 | P55145 | MANF | 1068 |
| 27 | 2 | 21.22172 | 0.183942 | 0.008668 | P51148 | RAB5C | 1069 |
| 9 | 3 | 25.05043 | 0.217061 | 0.008665 | P13598 | ICAM2 | 1070 |
| 42 | 14 | 22.3445 | 0.193412 | 0.008656 | P07355 | ANXA2 | 1071 |
| 9 | 2 | 19.36736 | 0.167591 | 0.008653 | P15121 | AKR1B1 | 1072 |
| 26 | 12 | 23.95692 | 0.20687 | 0.008635 | Q6DD88 | ATL3 | 1073 |
| 11 | 5 | 23.09858 | 0.199435 | 0.008634 | Q8NBX0 | SCCPDH | 1074 |
| 3 | 4 | 20.95476 | 0.180877 | 0.008632 | P20936 | RASA1 | 1075 |
| 13 | 4 | 22.66795 | 0.195599 | 0.008629 | Q92747 | ARPC1A | 1076 |
| 37 | 8 | 25.23371 | 0.217612 | 0.008624 | P08134 | RHOC | 1077 |
| 16 | 4 | 22.96915 | 0.197967 | 0.008619 | P55957 | BID | 1078 |

| | | | | | | | | |
|---|---|---|---|---|---|---|---|---|
| 13 | 1 | 21.50736 | 0.185158 | 0.008609 | P01704 | IGLV2-14 | | 1079 |
| 11 | 15 | 19.0727 | 0.164132 | 0.008606 | Q16531 | DDB1 | | 1080 |
| 27 | 6 | 23.18306 | 0.199129 | 0.008589 | P35813 | PPM1A | | 1081 |
| 28 | 7 | 22.82531 | 0.19596 | 0.008585 | P09497 | CLTB | | 1082 |
| 8 | 6 | 20.14247 | 0.172913 | 0.008584 | O94886 | TMEM63A | | 1083 |
| 6 | 3 | 20.98378 | 0.180013 | 0.008579 | Q92575 | UBXN4 | | 1084 |
| 12 | 4 | 23.5537 | 0.202035 | 0.008578 | P02686 | MBP | | 1085 |
| 5 | 3 | 28.29788 | 0.242433 | 0.008567 | Q8TEC5 | SH3RF2 | | 1086 |
| 33 | 23 | 25.87619 | 0.221167 | 0.008547 | Q9NQ75 | CASS4 | | 1087 |
| 6 | 1 | 19.0808 | 0.162957 | 0.00854 | Q13526 | PIN1 | | 1088 |
| 6 | 4 | 19.3091 | 0.164661 | 0.008528 | Q93050 | ATP6V0A1 | | 1089 |
| 24 | 3 | 22.57848 | 0.192162 | 0.008511 | P84085 | ARF5 | | 1090 |
| 23 | 8 | 22.34694 | 0.190039 | 0.008504 | Q9UNZ2 | NSFL1C | | 1091 |
| 20 | 9 | 21.05839 | 0.178933 | 0.008497 | P41091 | EIF2S3 | | 1092 |
| 51 | 3 | 26.56369 | 0.225521 | 0.00849 | Q9H0U4 | RAB1B | | 1093 |
| 7 | 5 | 19.11757 | 0.162224 | 0.008486 | Q7L1W4 | LRRC8D | | 1094 |
| 2 | 1 | 19.58908 | 0.16618 | 0.008483 | P02545 | LMNA | | 1095 |
| 17 | 2 | 20.22734 | 0.171551 | 0.008481 | P08493 | MGP | | 1096 |
| 13 | 5 | 20.1531 | 0.170598 | 0.008465 | P05154 | SERPINA5 | | 1097 |
| 23 | 23 | 29.24416 | 0.247541 | 0.008465 | P19823 | ITIH2 | | 1098 |
| 8 | 6 | 22.28374 | 0.188258 | 0.008448 | P42892 | ECE1 | | 1099 |
| 5 | 1 | 18.68171 | 0.157367 | 0.008424 | Q5W111 | SPRYD7 | | 1100 |
| 31 | 6 | 25.5125 | 0.214522 | 0.008409 | P14770 | GP9 | | 1101 |
| 27 | 17 | 24.46343 | 0.205635 | 0.008406 | P11216 | PYGB | | 1102 |
| 31 | 6 | 20.91991 | 0.175479 | 0.008388 | Q15126 | PMVK | | 1103 |
| 15 | 3 | 21.48369 | 0.180067 | 0.008382 | Q9UL25 | RAB21 | | 1104 |
| 10 | 1 | 21.42466 | 0.17875 | 0.008343 | Q96BM9 | ARL8A | | 1105 |
| 25 | 19 | 23.98541 | 0.199871 | 0.008333 | Q14BN4 | SLMAP | | 1106 |
| 5 | 2 | 19.85339 | 0.165193 | 0.008321 | Q9BZV1 | UBXN6 | | 1107 |
| 24 | 6 | 26.80665 | 0.222447 | 0.008298 | P25788 | PSMA3 | | 1108 |
| 15 | 5 | 22.19192 | 0.183991 | 0.008291 | P21964 | COMT | | 1109 |
| 8 | 6 | 17.60738 | 0.145847 | 0.008283 | Q9NUB1 | ACSS1 | | 1110 |
| 21 | 4 | 20.52868 | 0.169626 | 0.008263 | P28065 | PSMB9 | | 1111 |
| 27 | 28 | 25.8901 | 0.2139 | 0.008262 | Q27J81 | INF2 | | 1112 |
| 10 | 4 | 20.3978 | 0.168289 | 0.00825 | P62495 | ETF1 | | 1113 |
| 8 | 4 | 20.05542 | 0.165148 | 0.008235 | Q9Y265 | RUVBL1 | | 1114 |
| 22 | 10 | 22.33336 | 0.183407 | 0.008212 | Q86UE4 | MTDH | | 1115 |
| 24 | 20 | 21.84516 | 0.179312 | 0.008208 | P53814 | SMTN | | 1116 |
| 32 | 3 | 22.7648 | 0.186648 | 0.008199 | Q6IAA8 | LAMTOR1 | | 1117 |
| 1 | 1 | 18.57193 | 0.152192 | 0.008195 | Q6P3X3 | TTC27 | | 1118 |
| 40 | 6 | 23.2007 | 0.190104 | 0.008194 | P30086 | PEBP1 | | 1119 |
| 4 | 1 | 19.43486 | 0.159038 | 0.008183 | Q14093 | CYLC2 | | 1120 |
| 28 | 10 | 22.51184 | 0.184028 | 0.008175 | Q9HC38 | GLOD4 | | 1121 |
| 34 | 5 | 24.37263 | 0.199226 | 0.008174 | O60234 | GMFG | | 1122 |
| 25 | 10 | 23.74935 | 0.193735 | 0.008157 | O60268 | KIAA0513 | | 1123 |
| 38 | 8 | 24.68519 | 0.201217 | 0.008151 | P29692 | EEF1D | | 1124 |
| 9 | 10 | 23.60707 | 0.192134 | 0.008139 | P42338 | PIK3CB | | 1125 |

| | | | | | | | |
|---|---|---|---|---|---|---|---|
| 28 | 4 | 25.6576 | 0.208191 | 0.008114 | Q9HAV0 | GNB4 | 1126 |
| 20 | 6 | 21.34659 | 0.173054 | 0.008107 | P62714 | PPP2CB | 1127 |
| 22 | 1 | 21.28744 | 0.172443 | 0.008101 | A0A0C4DH | IGKV3D-20 | 1128 |
| 40 | 1 | 25.70838 | 0.208113 | 0.008095 | P07437 | TUBB | 1129 |
| 28 | 21 | 24.65479 | 0.199185 | 0.008079 | P43304 | GPD2 | 1130 |
| 1 | 1 | 21.0471 | 0.16987 | 0.008071 | Q9ULV0 | MYO5B | 1131 |
| 28 | 3 | 21.50793 | 0.17354 | 0.008069 | P61803 | DAD1 | 1132 |
| 25 | 10 | 24.17694 | 0.194818 | 0.008058 | P00390 | GSR | 1133 |
| 23 | 9 | 24.62245 | 0.198067 | 0.008044 | P62333 | PSMC6 | 1134 |
| 51 | 24 | 26.12626 | 0.210058 | 0.00804 | O43707 | ACTN4 | 1135 |
| 7 | 3 | 19.6108 | 0.157559 | 0.008034 | Q15517 | CDSN | 1136 |
| 32 | 14 | 24.1458 | 0.193967 | 0.008033 | Q9NTK5 | OLA1 | 1137 |
| 28 | 6 | 25.31657 | 0.203288 | 0.00803 | P20851 | C4BPB | 1138 |
| 31 | 2 | 23.01874 | 0.184826 | 0.008029 | Q8IZP0 | ABI1 | 1139 |
| 13 | 3 | 22.80576 | 0.182888 | 0.008019 | P68036 | UBE2L3 | 1140 |
| 14 | 4 | 19.09957 | 0.153077 | 0.008015 | Q9Y6C9 | MTCH2 | 1141 |
| 11 | 3 | 20.65899 | 0.165489 | 0.008011 | Q00059 | TFAM | 1142 |
| 8 | 5 | 21.65424 | 0.173416 | 0.008008 | Q8N573 | OXR1 | 1143 |
| 16 | 10 | 23.21285 | 0.185799 | 0.008004 | P05156 | CFI | 1144 |
| 32 | 25 | 28.86115 | 0.231008 | 0.008004 | P00736 | C1R | 1145 |
| 33 | 11 | 23.98334 | 0.191436 | 0.007982 | Q8TC12 | RDH11 | 1146 |
| 2 | 12 | 23.83071 | 0.19011 | 0.007978 | Q03164 | KMT2A | 1147 |
| 8 | 6 | 22.73785 | 0.180966 | 0.007959 | Q6EMK4 | VASN | 1148 |
| 18 | 11 | 23.78596 | 0.189133 | 0.007951 | Q32MZ4 | LRRFIP1 | 1149 |
| 24 | 13 | 23.88051 | 0.189664 | 0.007942 | Q96KN2 | CNDP1 | 1150 |
| 16 | 2 | 19.21697 | 0.152568 | 0.007939 | Q9Y3D6 | FIS1 | 1151 |
| 31 | 6 | 22.43759 | 0.177616 | 0.007916 | P36969 | GPX4 | 1152 |
| 51 | 10 | 26.5422 | 0.210051 | 0.007914 | P07203 | GPX1 | 1153 |
| 37 | 8 | 23.81185 | 0.188424 | 0.007913 | P00568 | AK1 | 1154 |
| 14 | 5 | 23.51751 | 0.185991 | 0.007909 | P61978 | HNRNPK | 1155 |
| 16 | 5 | 22.98451 | 0.181526 | 0.007898 | Q9UN19 | DAPP1 | 1156 |
| 32 | 4 | 24.78489 | 0.195638 | 0.007893 | P26885 | FKBP2 | 1157 |
| 3 | 2 | 20.41602 | 0.1605 | 0.007861 | P25098 | GRK2 | 1158 |
| 14 | 9 | 23.42012 | 0.183999 | 0.007856 | O60488 | ACSL4 | 1159 |
| 0 | 1 | 20.81557 | 0.163497 | 0.007855 | Q8TF72 | SHROOM3 | 1160 |
| 1 | 1 | 19.89939 | 0.156249 | 0.007852 | Q2KHT3 | CLEC16A | 1161 |
| 24 | 10 | 22.68807 | 0.17776 | 0.007835 | O43516 | WIPF1 | 1162 |
| 11 | 29 | 24.54002 | 0.192169 | 0.007831 | P49327 | FASN | 1163 |
| 29 | 6 | 21.70032 | 0.169879 | 0.007828 | O75396 | SEC22B | 1164 |
| 19 | 10 | 23.78574 | 0.186083 | 0.007823 | P51606 | RENBP | 1165 |
| 12 | 13 | 23.23987 | 0.181763 | 0.007821 | Q9Y2Q0 | ATP8A1 | 1166 |
| 49 | 19 | 28.06892 | 0.219481 | 0.007819 | Q9UBW5 | BIN2 | 1167 |
| 4 | 1 | 22.86163 | 0.178714 | 0.007817 | P08962 | CD63 | 1168 |
| 18 | 6 | 23.98923 | 0.187462 | 0.007814 | P40925 | MDH1 | 1169 |
| 11 | 1 | 20.79557 | 0.162267 | 0.007803 | P01700 | IGLV1-47 | 1170 |
| 23 | 5 | 23.26425 | 0.181232 | 0.00779 | O95336 | PGLS | 1171 |
| 14 | 11 | 22.34874 | 0.173487 | 0.007763 | Q9H2K8 | TAOK3 | 1172 |

| | | | | | | | |
|---|---|---|---|---|---|---|---|
| 44 | 13 | 29.71555 | 0.230126 | 0.007744 | O75636 | FCN3 | 1173 |
| 13 | 5 | 21.68019 | 0.16755 | 0.007728 | Q53GQ0 | HSD17B12 | 1174 |
| 16 | 9 | 23.29885 | 0.17997 | 0.007724 | P35611 | ADD1 | 1175 |
| 7 | 1 | 18.66909 | 0.143822 | 0.007704 | Q99627 | COPS8 | 1176 |
| 11 | 3 | 21.31413 | 0.163954 | 0.007692 | O94903 | PLPBP | 1177 |
| 4 | 18 | 23.89171 | 0.183746 | 0.007691 | Q86WI1 | PKHD1L1 | 1178 |
| 33 | 8 | 23.40699 | 0.179519 | 0.007669 | P60842 | EIF4A1 | 1179 |
| 26 | 13 | 24.10047 | 0.184259 | 0.007645 | Q14554 | PDIA5 | 1180 |
| 13 | 3 | 20.18625 | 0.154283 | 0.007643 | Q9H479 | FN3K | 1181 |
| 24 | 2 | 24.0639 | 0.18388 | 0.007641 | P06396 | GSN | 1182 |
| 23 | 10 | 23.66247 | 0.180727 | 0.007638 | Q99961 | SH3GL1 | 1183 |
| 18 | 7 | 23.78058 | 0.181001 | 0.007611 | Q9Y274 | ST3GAL6 | 1184 |
| 2 | 1 | 19.83712 | 0.15088 | 0.007606 | Q92609 | TBC1D5 | 1185 |
| 22 | 14 | 23.88853 | 0.181594 | 0.007602 | P34932 | HSPA4 | 1186 |
| 13 | 7 | 19.63681 | 0.148795 | 0.007577 | Q9Y6G9 | DYNC1LI1 | 1187 |
| 23 | 11 | 25.46893 | 0.192909 | 0.007574 | P12081 | HARS1 | 1188 |
| 4 | 2 | 21.46049 | 0.162467 | 0.007571 | Q14157 | UBAP2L | 1189 |
| 36 | 8 | 23.36184 | 0.176682 | 0.007563 | P54819 | AK2 | 1190 |
| 18 | 3 | 24.8616 | 0.187888 | 0.007557 | P01701 | IGLV1-51 | 1191 |
| 2 | 1 | 20.87669 | 0.157575 | 0.007548 | Q13107 | USP4 | 1192 |
| 22 | 9 | 24.78362 | 0.186827 | 0.007538 | Q15019 | SEPTIN2 | 1193 |
| 24 | 8 | 23.17423 | 0.174426 | 0.007527 | Q13561 | DCTN2 | 1194 |
| 26 | 16 | 27.05158 | 0.203531 | 0.007524 | Q10567 | AP1B1 | 1195 |
| 19 | 5 | 23.58058 | 0.177351 | 0.007521 | P49755 | TMED10 | 1196 |
| 9 | 14 | 22.85229 | 0.17169 | 0.007513 | P20930 | FLG | 1197 |
| 17 | 4 | 22.14349 | 0.166348 | 0.007512 | Q9Y3Q8 | TSC22D4 | 1198 |
| 49 | 9 | 24.86739 | 0.186744 | 0.00751 | P32119 | PRDX2 | 1199 |
| 55 | 15 | 25.41221 | 0.190695 | 0.007504 | P23284 | PPIB | 1200 |
| 22 | 11 | 23.86538 | 0.179052 | 0.007503 | O95870 | ABHD16A | 1201 |
| 30 | 13 | 25.33165 | 0.189762 | 0.007491 | P01137 | TGFB1 | 1202 |
| 3 | 2 | 20.25822 | 0.151445 | 0.007476 | Q96M96 | FGD4 | 1203 |
| 26 | 8 | 22.14231 | 0.165481 | 0.007474 | Q02978 | SLC25A11 | 1204 |
| 18 | 6 | 22.84639 | 0.170468 | 0.007461 | Q9H0P0 | NT5C3A | 1205 |
| 34 | 5 | 25.35548 | 0.188889 | 0.00745 | P10606 | COX5B | 1206 |
| 15 | 12 | 21.53004 | 0.159624 | 0.007414 | P98194 | ATP2C1 | 1207 |
| 40 | 4 | 24.34683 | 0.18036 | 0.007408 | P00739 | HPR | 1208 |
| 19 | 24 | 23.82179 | 0.1759 | 0.007384 | Q13464 | ROCK1 | 1209 |
| 0 | 1 | 21.11439 | 0.155905 | 0.007384 | Q9Y6V0 | PCLO | 1210 |
| 11 | 5 | 22.65871 | 0.167285 | 0.007383 | P36507 | MAP2K2 | 1211 |
| 19 | 19 | 28.1519 | 0.207819 | 0.007382 | O60313 | OPA1 | 1212 |
| 16 | 6 | 22.05989 | 0.1627 | 0.007375 | P49137 | MAPKAPK2 | 1213 |
| 8 | 8 | 21.99872 | 0.161776 | 0.007354 | Q15042 | RAB3GAP1 | 1214 |
| 19 | 2 | 22.12427 | 0.162697 | 0.007354 | P60985 | KRTDAP | 1215 |
| 4 | 1 | 19.9944 | 0.147014 | 0.007353 | P49366 | DHPS | 1216 |
| 49 | 12 | 25.63631 | 0.188435 | 0.00735 | O00161 | SNAP23 | 1217 |
| 7 | 1 | 23.43064 | 0.172142 | 0.007347 | Q9UII2 | ATP5IF1 | 1218 |
| 27 | 5 | 24.47606 | 0.179594 | 0.007338 | Q9UMY4 | SNX12 | 1219 |

| | | | | | | | |
|---|---|---|---|---|---|---|---|
| 7 | 8 | 26.03849 | 0.190911 | 0.007332 | Q9Y4G6 | TLN2 | 1220 |
| 14 | 6 | 29.00253 | 0.21126 | 0.007284 | Q6ZVM7 | TOM1L2 | 1221 |
| 24 | 14 | 23.18616 | 0.168817 | 0.007281 | P48740 | MASP1 | 1222 |
| 18 | 5 | 21.05114 | 0.153236 | 0.007279 | P29218 | IMPA1 | 1223 |
| 41 | 30 | 28.80633 | 0.209198 | 0.007262 | P00488 | F13A1 | 1224 |
| 31 | 3 | 20.92574 | 0.151624 | 0.007246 | P14854 | COX6B1 | 1225 |
| 42 | 22 | 25.28551 | 0.182936 | 0.007235 | P54577 | YARS1 | 1226 |
| 42 | 9 | 24.15372 | 0.17471 | 0.007233 | P21796 | VDAC1 | 1227 |
| 11 | 2 | 24.64707 | 0.178151 | 0.007228 | Q7RTS7 | KRT74 | 1228 |
| 48 | 31 | 29.30907 | 0.211808 | 0.007227 | Q86UX7 | FERMT3 | 1229 |
| 2 | 5 | 23.66824 | 0.170954 | 0.007223 | O75037 | KIF21B | 1230 |
| 15 | 3 | 21.79016 | 0.157241 | 0.007216 | O43633 | CHMP2A | 1231 |
| 16 | 5 | 21.64281 | 0.156097 | 0.007212 | Q9UHD1 | CHORDC1 | 1232 |
| 28 | 6 | 25.93408 | 0.186967 | 0.007209 | P02743 | APCS | 1233 |
| 18 | 12 | 23.74148 | 0.170698 | 0.00719 | O95782 | AP2A1 | 1234 |
| 1 | 2 | 20.72517 | 0.148841 | 0.007182 | Q9BX66 | SORBS1 | 1235 |
| 16 | 14 | 23.95697 | 0.171861 | 0.007174 | Q7L7X3 | TAOK1 | 1236 |
| 2 | 1 | 20.41516 | 0.145938 | 0.007148 | Q8NH72 | OR4C6 | 1237 |
| 13 | 6 | 22.9841 | 0.164282 | 0.007148 | Q16610 | ECM1 | 1238 |
| 7 | 10 | 22.38165 | 0.159957 | 0.007147 | A2RUS2 | DENND3 | 1239 |
| 6 | 5 | 24.81845 | 0.177312 | 0.007144 | P13639 | EEF2 | 1240 |
| 32 | 22 | 25.09419 | 0.179178 | 0.00714 | Q05209 | PTPN12 | 1241 |
| 43 | 6 | 28.80561 | 0.204847 | 0.007111 | P0DOX5 | #N/A | 1242 |
| 30 | 16 | 24.57362 | 0.174456 | 0.007099 | P17987 | TCP1 | 1243 |
| 23 | 13 | 22.96316 | 0.162962 | 0.007097 | O15231 | ZNF185 | 1244 |
| 0 | 1 | 20.81595 | 0.147406 | 0.007081 | Q9NSI6 | BRWD1 | 1245 |
| 4 | 1 | 18.61071 | 0.131668 | 0.007075 | Q9UKK9 | NUDT5 | 1246 |
| 16 | 4 | 22.17735 | 0.156866 | 0.007073 | P00746 | CFD | 1247 |
| 10 | 27 | 24.13003 | 0.17053 | 0.007067 | Q13813 | SPTAN1 | 1248 |
| 5 | 3 | 21.55984 | 0.152338 | 0.007066 | Q9H845 | ACAD9 | 1249 |
| 29 | 15 | 24.636 | 0.173894 | 0.007059 | Q8N9U0 | TC2N | 1250 |
| 28 | 16 | 24.78649 | 0.174876 | 0.007055 | P49368 | CCT3 | 1251 |
| 25 | 17 | 24.67496 | 0.174069 | 0.007054 | Q8TF42 | UBASH3B | 1252 |
| 10 | 3 | 24.16077 | 0.17044 | 0.007054 | Q9Y3A6 | TMED5 | 1253 |
| 34 | 21 | 24.59918 | 0.173468 | 0.007052 | Q06187 | BTK | 1254 |
| 49 | 7 | 25.44915 | 0.179291 | 0.007045 | P09493 | TPM1 | 1255 |
| 4 | 9 | 25.3747 | 0.17864 | 0.00704 | O00507 | USP9Y | 1256 |
| 15 | 10 | 25.42673 | 0.178525 | 0.007021 | O00187 | MASP2 | 1257 |
| 5 | 1 | 20.97096 | 0.147236 | 0.007021 | P0C025 | NUDT17 | 1258 |
| 22 | 11 | 24.0881 | 0.168889 | 0.007011 | P43405 | SYK | 1259 |
| 3 | 1 | 23.55388 | 0.16495 | 0.007003 | A6NCD4 | CC2D2B | 1260 |
| 11 | 6 | 23.49121 | 0.164388 | 0.006998 | Q9Y251 | HPSE | 1261 |
| 35 | 28 | 26.18276 | 0.183223 | 0.006998 | Q0ZGT2 | NEXN | 1262 |
| 30 | 7 | 21.16907 | 0.148077 | 0.006995 | Q9NX63 | CHCHD3 | 1263 |
| 45 | 12 | 26.43399 | 0.184059 | 0.006963 | Q13642 | FHL1 | 1264 |
| 22 | 7 | 22.89679 | 0.15941 | 0.006962 | Q4G0F5 | VPS26B | 1265 |
| 29 | 24 | 24.58411 | 0.171128 | 0.006961 | P11586 | MTHFD1 | 1266 |

| | | | | | | | |
|---|---|---|---|---|---|---|---|
| 16 | 5 | 20.29689 | 0.14075 | 0.006935 | Q9Y6I9 | TEX264 | 1267 |
| 16 | 1 | 20.50196 | 0.142159 | 0.006934 | Q5VTU8 | ATP5F1EP2 | 1268 |
| 2 | 20 | 24.78358 | 0.171705 | 0.006928 | Q8WXH0 | SYNE2 | 1269 |
| 20 | 4 | 22.22077 | 0.153589 | 0.006912 | P10301 | RRAS | 1270 |
| 2 | 1 | 21.53265 | 0.148831 | 0.006912 | Q9BUH8 | BEGAIN | 1271 |
| 54 | 4 | 28.45995 | 0.19671 | 0.006912 | P24844 | MYL9 | 1272 |
| 28 | 4 | 25.99513 | 0.179605 | 0.006909 | P61769 | B2M | 1273 |
| 33 | 6 | 25.60904 | 0.176763 | 0.006902 | P12236 | SLC25A6 | 1274 |
| 18 | 10 | 23.19408 | 0.160019 | 0.006899 | P34897 | SHMT2 | 1275 |
| 36 | 6 | 23.6848 | 0.163221 | 0.006891 | Q13637 | RAB32 | 1276 |
| 9 | 6 | 21.67577 | 0.148544 | 0.006853 | P08174 | CD55 | 1277 |
| 10 | 6 | 20.92174 | 0.142829 | 0.006827 | Q5T0N5 | FNBP1L | 1278 |
| 7 | 1 | 25.48651 | 0.173723 | 0.006816 | P60673 | PFN3 | 1279 |
| 3 | 11 | 22.95618 | 0.156379 | 0.006812 | Q709C8 | VPS13C | 1280 |
| 4 | 12 | 21.43507 | 0.145877 | 0.006806 | P78559 | MAP1A | 1281 |
| 28 | 19 | 27.43773 | 0.186669 | 0.006803 | Q9ULV4 | CORO1C | 1282 |
| 17 | 2 | 22.98953 | 0.156374 | 0.006802 | P01111 | NRAS | 1283 |
| 34 | 10 | 24.41817 | 0.165888 | 0.006794 | Q9P126 | CLEC1B | 1284 |
| 26 | 7 | 23.70361 | 0.161025 | 0.006793 | Q96AP7 | ESAM | 1285 |
| 41 | 5 | 22.97544 | 0.156073 | 0.006793 | P61225 | RAP2B | 1286 |
| 3 | 1 | 20.0837 | 0.136242 | 0.006784 | Q9Y2S2 | CRYL1 | 1287 |
| 7 | 7 | 22.07098 | 0.149574 | 0.006777 | O94804 | STK10 | 1288 |
| 2 | 19 | 23.68841 | 0.160169 | 0.006761 | Q5VST9 | OBSCN | 1289 |
| 11 | 11 | 23.24784 | 0.157187 | 0.006761 | O43747 | AP1G1 | 1290 |
| 20 | 15 | 23.62348 | 0.15919 | 0.006739 | Q9BX10 | GTPBP2 | 1291 |
| 28 | 5 | 22.54709 | 0.151648 | 0.006726 | P20618 | PSMB1 | 1292 |
| 19 | 11 | 24.59342 | 0.164913 | 0.006706 | O94826 | TOMM70 | 1293 |
| 49 | 1 | 23.08818 | 0.154628 | 0.006697 | P09493 | TPM1 | 1294 |
| 0 | 1 | 18.95363 | 0.126934 | 0.006697 | Q8TCU4 | ALMS1 | 1295 |
| 51 | 9 | 23.68263 | 0.157875 | 0.006666 | P28066 | PSMA5 | 1296 |
| 7 | 1 | 20.21973 | 0.134536 | 0.006654 | Q9NX14 | NDUFB11 | 1297 |
| 29 | 5 | 21.60151 | 0.143631 | 0.006649 | Q13442 | PDAP1 | 1298 |
| 10 | 7 | 23.13715 | 0.153603 | 0.006639 | Q8IY22 | CMIP | 1299 |
| 41 | 4 | 25.96613 | 0.172101 | 0.006628 | P37840 | SNCA | 1300 |
| 48 | 3 | 23.04523 | 0.152532 | 0.006619 | P67936 | TPM4 | 1301 |
| 39 | 9 | 24.84595 | 0.164082 | 0.006604 | O00139 | KIF2A | 1302 |
| 27 | 9 | 23.98943 | 0.158195 | 0.006594 | P04217 | A1BG | 1303 |
| 46 | 10 | 25.90616 | 0.170268 | 0.006572 | Q9Y624 | F11R | 1304 |
| 4 | 3 | 22.26955 | 0.14622 | 0.006566 | Q9NQ79 | CRTAC1 | 1305 |
| 29 | 15 | 25.89944 | 0.169945 | 0.006562 | P40227 | CCT6A | 1306 |
| 1 | 1 | 19.3056 | 0.126141 | 0.006534 | P47712 | PLA2G4A | 1307 |
| 6 | 2 | 21.48123 | 0.140233 | 0.006528 | Q96QR8 | PURB | 1308 |
| 23 | 19 | 24.04189 | 0.156691 | 0.006517 | P35606 | COPB2 | 1309 |
| 39 | 8 | 25.97398 | 0.169174 | 0.006513 | O00299 | CLIC1 | 1310 |
| 32 | 2 | 26.07483 | 0.169404 | 0.006497 | P0DOX8 | #N/A | 1311 |
| 57 | 8 | 21.26021 | 0.138078 | 0.006495 | Q9UKY7 | CDV3 | 1312 |
| 35 | 13 | 28.23762 | 0.183228 | 0.006489 | Q15404 | RSU1 | 1313 |

| | | | | | | | |
|---|---|---|---|---|---|---|---|
| 45 | 3 | 23.81607 | 0.154527 | 0.006488 | Q9BQE3 | TUBA1C | 1314 |
| 29 | 6 | 23.07434 | 0.149649 | 0.006486 | P30626 | SRI | 1315 |
| 10 | 6 | 21.21106 | 0.137253 | 0.006471 | O43312 | MTSS1 | 1316 |
| 33 | 18 | 27.04196 | 0.174913 | 0.006468 | Q16851 | UGP2 | 1317 |
| 44 | 20 | 28.33459 | 0.182831 | 0.006453 | P01009 | SERPINA1 | 1318 |
| 2 | 10 | 24.48077 | 0.157854 | 0.006448 | Q63HN8 | RNF213 | 1319 |
| 13 | 4 | 20.86783 | 0.134218 | 0.006432 | Q9NUJ1 | ABHD10 | 1320 |
| 12 | 4 | 21.75416 | 0.139724 | 0.006423 | O43772 | SLC25A20 | 1321 |
| 8 | 6 | 22.13856 | 0.14206 | 0.006417 | P15498 | VAV1 | 1322 |
| 21 | 5 | 23.45942 | 0.150204 | 0.006403 | Q9H7M9 | VSIR | 1323 |
| 19 | 5 | 24.23085 | 0.155101 | 0.006401 | Q969T9 | WBP2 | 1324 |
| 5 | 6 | 22.95565 | 0.146774 | 0.006394 | P16885 | PLCG2 | 1325 |
| 10 | 4 | 22.21503 | 0.141706 | 0.006379 | Q96G23 | CERS2 | 1326 |
| 1 | 1 | 20.01688 | 0.127446 | 0.006367 | Q96NL6 | SCLT1 | 1327 |
| 19 | 12 | 26.07572 | 0.165942 | 0.006364 | Q9ULC5 | ACSL5 | 1328 |
| 1 | 1 | 21.46399 | 0.136445 | 0.006357 | Q5T5U3 | ARHGAP21 | 1329 |
| 42 | 20 | 30.2551 | 0.192153 | 0.006351 | P06396 | GSN | 1330 |
| 15 | 4 | 21.65092 | 0.137493 | 0.00635 | P09486 | SPARC | 1331 |
| 19 | 13 | 23.75091 | 0.150609 | 0.006341 | P38606 | ATP6V1A | 1332 |
| 34 | 14 | 25.18899 | 0.15963 | 0.006337 | P37837 | TALDO1 | 1333 |
| 24 | 4 | 23.14276 | 0.1466 | 0.006335 | P61088 | UBE2N | 1334 |
| 59 | 14 | 28.53616 | 0.180618 | 0.006329 | P02533 | KRT14 | 1335 |
| 21 | 14 | 23.6677 | 0.149736 | 0.006327 | P40763 | STAT3 | 1336 |
| 32 | 3 | 24.79973 | 0.156805 | 0.006323 | Q9H299 | SH3BGRL3 | 1337 |
| 12 | 9 | 22.34124 | 0.141141 | 0.006318 | P35612 | ADD2 | 1338 |
| 30 | 5 | 22.22502 | 0.140381 | 0.006316 | Q04837 | SSBP1 | 1339 |
| 20 | 13 | 25.90333 | 0.163574 | 0.006315 | Q13976 | PRKG1 | 1340 |
| 16 | 19 | 23.49619 | 0.148309 | 0.006312 | Q9Y2A7 | NCKAP1 | 1341 |
| 27 | 12 | 24.95305 | 0.15716 | 0.006298 | Q96PD5 | PGLYRP2 | 1342 |
| 15 | 9 | 24.83845 | 0.156403 | 0.006297 | Q9NUY8 | TBC1D23 | 1343 |
| 10 | 12 | 23.9331 | 0.150427 | 0.006285 | O94979 | SEC31A | 1344 |
| 20 | 9 | 22.80327 | 0.143122 | 0.006276 | Q96KP4 | CNDP2 | 1345 |
| 32 | 5 | 25.4462 | 0.159704 | 0.006276 | P04792 | HSPB1 | 1346 |
| 37 | 11 | 24.13755 | 0.151398 | 0.006272 | P54920 | NAPA | 1347 |
| 21 | 4 | 21.65983 | 0.135806 | 0.00627 | P20674 | COX5A | 1348 |
| 41 | 9 | 23.57518 | 0.147515 | 0.006257 | Q9BUL8 | PDCD10 | 1349 |
| 1 | 1 | 23.54594 | 0.1473 | 0.006256 | Q9P2P1 | NYNRIN | 1350 |
| 24 | 7 | 23.1275 | 0.144121 | 0.006232 | Q9NVA2 | SEPTIN11 | 1351 |
| 29 | 3 | 27.85149 | 0.172946 | 0.00621 | Q86YW5 | TREML1 | 1352 |
| 42 | 16 | 25.44178 | 0.157902 | 0.006206 | Q9Y608 | LRRFIP2 | 1353 |
| 51 | 4 | 23.79665 | 0.147197 | 0.006186 | P15531 | NME1 | 1354 |
| 16 | 5 | 20.36018 | 0.125612 | 0.006169 | O00743 | PPP6C | 1355 |
| 26 | 25 | 25.36092 | 0.156411 | 0.006167 | Q14697 | GANAB | 1356 |
| 20 | 14 | 23.4401 | 0.144529 | 0.006166 | A0FGR8 | ESYT2 | 1357 |
| 7 | 5 | 19.72002 | 0.121402 | 0.006156 | Q17RC7 | EXOC3L4 | 1358 |
| 21 | 2 | 21.96801 | 0.134975 | 0.006144 | Q14203 | DCTN1 | 1359 |
| 1 | 1 | 20.10222 | 0.12337 | 0.006137 | O14917 | PCDH17 | 1360 |

| | | | | | | | |
|---|---|---|---|---|---|---|---|
| 18 | 14 | 23.66355 | 0.144785 | 0.006118 | Q06033 | ITIH3 | 1361 |
| 13 | 3 | 22.24669 | 0.1358 | 0.006104 | P28070 | PSMB4 | 1362 |
| 32 | 3 | 25.95724 | 0.158275 | 0.006098 | O00483 | NDUFA4 | 1363 |
| 16 | 6 | 24.15322 | 0.147146 | 0.006092 | Q9UNS2 | COPS3 | 1364 |
| 8 | 1 | 21.05531 | 0.128122 | 0.006085 | P17096 | HMGA1 | 1365 |
| 29 | 15 | 23.47596 | 0.142439 | 0.006067 | P00367 | GLUD1 | 1366 |
| 27 | 14 | 23.74784 | 0.144024 | 0.006065 | P36957 | DLST | 1367 |
| 30 | 6 | 26.14367 | 0.158529 | 0.006064 | P04179 | SOD2 | 1368 |
| 50 | 3 | 28.22498 | 0.171073 | 0.006061 | P0DOX7 | #N/A | 1369 |
| 5 | 3 | 18.25757 | 0.110248 | 0.006038 | Q9NVE7 | PANK4 | 1370 |
| 10 | 2 | 24.6096 | 0.148595 | 0.006038 | P00414 | MT-CO3 | 1371 |
| 17 | 7 | 23.2198 | 0.140202 | 0.006038 | Q96AG4 | LRRC59 | 1372 |
| 22 | 1 | 24.00903 | 0.14427 | 0.006009 | O43852 | CALU | 1373 |
| 14 | 8 | 21.80491 | 0.130868 | 0.006002 | P43155 | CRAT | 1374 |
| 19 | 12 | 24.00774 | 0.143703 | 0.005986 | P23368 | ME2 | 1375 |
| 13 | 2 | 22.35755 | 0.133627 | 0.005977 | P55209 | NAP1L1 | 1376 |
| 38 | 5 | 26.1555 | 0.15625 | 0.005974 | P02775 | PPBP | 1377 |
| 14 | 11 | 22.51159 | 0.134027 | 0.005954 | P41250 | GARS1 | 1378 |
| 15 | 2 | 25.14101 | 0.14966 | 0.005953 | Q96DZ9 | CMTM5 | 1379 |
| 28 | 8 | 23.90566 | 0.142153 | 0.005946 | P30042 | GATD3A | 1380 |
| 3 | 2 | 27.08043 | 0.160591 | 0.00593 | P27987 | ITPKB | 1381 |
| 4 | 6 | 24.17333 | 0.143159 | 0.005922 | Q9NQC3 | RTN4 | 1382 |
| 30 | 8 | 24.87839 | 0.147297 | 0.005921 | P51572 | BCAP31 | 1383 |
| 34 | 6 | 24.11125 | 0.142445 | 0.005908 | P05141 | SLC25A5 | 1384 |
| 27 | 11 | 22.02156 | 0.130075 | 0.005907 | Q8NG06 | TRIM58 | 1385 |
| 30 | 10 | 25.63656 | 0.15137 | 0.005904 | P02760 | AMBP | 1386 |
| 15 | 30 | 25.0918 | 0.148001 | 0.005898 | Q15746 | MYLK | 1387 |
| 22 | 3 | 22.01281 | 0.129794 | 0.005896 | P20338 | RAB4A | 1388 |
| 25 | 9 | 22.26433 | 0.131204 | 0.005893 | P22234 | PAICS | 1389 |
| 22 | 16 | 23.77115 | 0.139148 | 0.005854 | P33176 | KIF5B | 1390 |
| 19 | 18 | 23.48926 | 0.137308 | 0.005846 | O60229 | KALRN | 1391 |
| 4 | 2 | 19.10744 | 0.111638 | 0.005843 | Q9C0E8 | LNPK | 1392 |
| 32 | 10 | 24.48974 | 0.142904 | 0.005835 | Q99426 | TBCB | 1393 |
| 35 | 3 | 25.46236 | 0.148214 | 0.005821 | A0A075B6 | IGLV3-10 | 1394 |
| 8 | 1 | 20.71762 | 0.120585 | 0.00582 | P07919 | UQCRH | 1395 |
| 10 | 3 | 21.16356 | 0.123179 | 0.00582 | Q86UF1 | TSPAN33 | 1396 |
| 12 | 10 | 27.10236 | 0.157389 | 0.005807 | Q8NDB2 | BANK1 | 1397 |
| 13 | 6 | 25.28786 | 0.14684 | 0.005807 | P01019 | AGT | 1398 |
| 25 | 3 | 21.4802 | 0.124543 | 0.005798 | O43169 | CYB5B | 1399 |
| 44 | 8 | 25.36062 | 0.146914 | 0.005793 | P09211 | GSTP1 | 1400 |
| 18 | 7 | 25.6208 | 0.148268 | 0.005787 | Q9UHQ9 | CYB5R1 | 1401 |
| 3 | 1 | 21.02208 | 0.121591 | 0.005784 | Q9Y3E7 | CHMP3 | 1402 |
| 15 | 6 | 27.34948 | 0.157989 | 0.005777 | P21926 | CD9 | 1403 |
| 27 | 9 | 23.40511 | 0.135001 | 0.005768 | Q9BXS5 | AP1M1 | 1404 |
| 32 | 14 | 24.85014 | 0.143329 | 0.005768 | P14868 | DARS1 | 1405 |
| 8 | 3 | 20.72272 | 0.119463 | 0.005765 | Q15738 | NSDHL | 1406 |
| 2 | 4 | 28.47893 | 0.163943 | 0.005757 | Q8N3C0 | ASCC3 | 1407 |

| | | | | | | | |
|---|---|---|---|---|---|---|---|
| 17 | 7 | 23.25134 | 0.133562 | 0.005744 | Q9NR46 | SH3GLB2 | 1408 |
| 3 | 1 | 22.27504 | 0.127672 | 0.005732 | Q9NZN8 | CNOT2 | 1409 |
| 14 | 7 | 26.95467 | 0.154494 | 0.005732 | P16671 | CD36 | 1410 |
| 51 | 6 | 24.85349 | 0.14244 | 0.005731 | P05109 | S100A8 | 1411 |
| 6 | 9 | 24.44351 | 0.14006 | 0.00573 | Q5T5C0 | STXBP5 | 1412 |
| 25 | 3 | 23.09069 | 0.131971 | 0.005715 | P13987 | CD59 | 1413 |
| 8 | 1 | 19.49425 | 0.110791 | 0.005683 | O95167 | NDUFA3 | 1414 |
| 27 | 6 | 26.62848 | 0.151258 | 0.00568 | P30084 | ECHS1 | 1415 |
| 22 | 4 | 21.40501 | 0.121163 | 0.00566 | Q86YL5 | TDRP | 1416 |
| 31 | 15 | 26.02845 | 0.14731 | 0.00566 | P02790 | HPX | 1417 |
| 26 | 1 | 19.26531 | 0.109008 | 0.005658 | P07948 | LYN | 1418 |
| 62 | 11 | 27.54338 | 0.155836 | 0.005658 | P62258 | YWHAE | 1419 |
| 11 | 5 | 21.22226 | 0.119619 | 0.005637 | Q8N490 | PNKD | 1420 |
| 34 | 9 | 25.73099 | 0.144891 | 0.005631 | P08238 | HSP90AB1 | 1421 |
| 24 | 14 | 23.93078 | 0.134485 | 0.00562 | Q16799 | RTN1 | 1422 |
| 20 | 10 | 22.59986 | 0.12685 | 0.005613 | Q9BW04 | SARG | 1423 |
| 35 | 20 | 24.23431 | 0.135767 | 0.005602 | P48444 | ARCN1 | 1424 |
| 24 | 8 | 22.99531 | 0.128652 | 0.005595 | O75791 | GRAP2 | 1425 |
| 27 | 17 | 25.14354 | 0.140447 | 0.005586 | O00429 | DNM1L | 1426 |
| 30 | 5 | 22.9449 | 0.127906 | 0.005574 | O95292 | VAPB | 1427 |
| 29 | 18 | 26.59827 | 0.148248 | 0.005574 | P31146 | CORO1A | 1428 |
| 14 | 5 | 21.69813 | 0.1209 | 0.005572 | Q9UBS4 | DNAJB11 | 1429 |
| 20 | 17 | 24.53893 | 0.136622 | 0.005568 | P13671 | C6 | 1430 |
| 15 | 4 | 20.281 | 0.112887 | 0.005566 | P53004 | BLVRA | 1431 |
| 9 | 4 | 21.23088 | 0.118032 | 0.005559 | Q92598 | HSPH1 | 1432 |
| 13 | 4 | 23.78451 | 0.132211 | 0.005559 | Q9Y376 | CAB39 | 1433 |
| 17 | 8 | 25.25047 | 0.140353 | 0.005558 | O75390 | CS | 1434 |
| 21 | 9 | 22.85223 | 0.126637 | 0.005542 | P08670 | VIM | 1435 |
| 31 | 6 | 24.30928 | 0.134316 | 0.005525 | P48047 | ATP5PO | 1436 |
| 14 | 5 | 21.07177 | 0.116029 | 0.005506 | Q9HB40 | SCPEP1 | 1437 |
| 3 | 13 | 25.63438 | 0.141123 | 0.005505 | O15050 | TRANK1 | 1438 |
| 26 | 18 | 27.19125 | 0.149353 | 0.005493 | P09871 | C1S | 1439 |
| 28 | 22 | 25.91757 | 0.142264 | 0.005489 | P16615 | ATP2A2 | 1440 |
| 20 | 11 | 22.94954 | 0.125958 | 0.005488 | P07954 | FH | 1441 |
| 10 | 7 | 21.72929 | 0.119065 | 0.005479 | Q6UWL2 | SUSD1 | 1442 |
| 27 | 10 | 21.49942 | 0.117541 | 0.005467 | Q9UNM6 | PSMD13 | 1443 |
| 6 | 3 | 19.7753 | 0.108037 | 0.005463 | P10515 | DLAT | 1444 |
| 28 | 12 | 26.46135 | 0.144505 | 0.005461 | O43294 | TGFB1I1 | 1445 |
| 52 | 13 | 24.49626 | 0.133761 | 0.00546 | P08779 | KRT16 | 1446 |
| 29 | 19 | 25.5709 | 0.139525 | 0.005456 | P49748 | ACADVL | 1447 |
| 17 | 15 | 24.80073 | 0.135165 | 0.00545 | Q9H3U1 | UNC45A | 1448 |
| 2 | 2 | 19.8079 | 0.107835 | 0.005444 | P22102 | GART | 1449 |
| 22 | 35 | 28.68609 | 0.156029 | 0.005439 | P16157 | ANK1 | 1450 |
| 8 | 14 | 24.92903 | 0.135552 | 0.005438 | Q92954 | PRG4 | 1451 |
| 39 | 24 | 28.52784 | 0.154892 | 0.005429 | Q01518 | CAP1 | 1452 |
| 6 | 7 | 22.75481 | 0.123418 | 0.005424 | Q9UBN7 | HDAC6 | 1453 |
| 36 | 16 | 24.42007 | 0.132095 | 0.005409 | P41240 | CSK | 1454 |

| | | | | | | | |
|---|---|---|---|---|---|---|---|
| 38 | 4 | 22.19506 | 0.120025 | 0.005408 | O43768 | ENSA | 1455 |
| 26 | 2 | 24.70195 | 0.133458 | 0.005403 | A2NJV5 | IGKV2-29 | 1456 |
| 5 | 3 | 24.65157 | 0.133088 | 0.005399 | Q8TC07 | TBC1D15 | 1457 |
| 22 | 25 | 23.56899 | 0.127123 | 0.005394 | Q9Y4D1 | DAAM1 | 1458 |
| 35 | 10 | 23.9582 | 0.128861 | 0.005379 | Q9UIB8 | CD84 | 1459 |
| 29 | 18 | 25.23018 | 0.135563 | 0.005373 | Q7LDG7 | RASGRP2 | 1460 |
| 26 | 25 | 26.23782 | 0.140765 | 0.005365 | P49588 | AARS1 | 1461 |
| 43 | 11 | 28.27079 | 0.151594 | 0.005362 | P00738 | HP | 1462 |
| 35 | 7 | 24.61401 | 0.131912 | 0.005359 | P51149 | RAB7A | 1463 |
| 34 | 5 | 23.6457 | 0.126449 | 0.005348 | P13073 | COX4I1 | 1464 |
| 9 | 29 | 25.31824 | 0.135196 | 0.00534 | Q96RL7 | VPS13A | 1465 |
| 18 | 11 | 22.76437 | 0.121282 | 0.005328 | P17858 | PFKL | 1466 |
| 21 | 4 | 26.12577 | 0.13916 | 0.005327 | P0DOX2 | #N/A | 1467 |
| 25 | 12 | 23.4661 | 0.124975 | 0.005326 | P63010 | AP2B1 | 1468 |
| 12 | 7 | 22.78154 | 0.120685 | 0.005297 | P51812 | RPS6KA3 | 1469 |
| 2 | 7 | 22.30488 | 0.118146 | 0.005297 | Q12923 | PTPN13 | 1470 |
| 34 | 15 | 26.44991 | 0.139185 | 0.005262 | P27797 | CALR | 1471 |
| 15 | 3 | 21.10617 | 0.110934 | 0.005256 | Q9H2U2 | PPA2 | 1472 |
| 32 | 7 | 26.13599 | 0.137094 | 0.005245 | P24539 | ATP5PB | 1473 |
| 25 | 7 | 24.23312 | 0.126986 | 0.00524 | Q15257 | PTPA | 1474 |
| 5 | 2 | 23.37525 | 0.122489 | 0.00524 | Q12840 | KIF5A | 1475 |
| 24 | 4 | 23.10231 | 0.120088 | 0.005198 | Q3ZCW2 | LGALSL | 1476 |
| 37 | 3 | 28.34925 | 0.147105 | 0.005189 | P01717 | IGLV3-25 | 1477 |
| 39 | 11 | 27.34116 | 0.141717 | 0.005183 | Q14847 | LASP1 | 1478 |
| 40 | 15 | 25.16826 | 0.130428 | 0.005182 | O75563 | SKAP2 | 1479 |
| 25 | 14 | 24.51372 | 0.126916 | 0.005177 | Q13094 | LCP2 | 1480 |
| 5 | 3 | 22.88144 | 0.118404 | 0.005175 | Q96AC1 | FERMT2 | 1481 |
| 1 | 1 | 21.59723 | 0.111545 | 0.005165 | O43861 | ATP9B | 1482 |
| 23 | 19 | 25.84198 | 0.133455 | 0.005164 | P07384 | CAPN1 | 1483 |
| 21 | 2 | 24.07145 | 0.124112 | 0.005156 | P01624 | IGKV3-15 | 1484 |
| 22 | 5 | 22.81809 | 0.117467 | 0.005148 | O75915 | ARL6IP5 | 1485 |
| 28 | 17 | 22.80998 | 0.1172 | 0.005138 | P31040 | SDHA | 1486 |
| 16 | 3 | 23.04244 | 0.11795 | 0.005119 | Q9UBQ0 | VPS29 | 1487 |
| 10 | 7 | 23.00231 | 0.117733 | 0.005118 | O95202 | LETM1 | 1488 |
| 15 | 13 | 24.67199 | 0.126077 | 0.00511 | Q9UKE5 | TNIK | 1489 |
| 3 | 2 | 20.22808 | 0.103268 | 0.005105 | Q96K49 | TMEM87B | 1490 |
| 43 | 24 | 28.71872 | 0.146449 | 0.005099 | Q13418 | ILK | 1491 |
| 20 | 10 | 23.9398 | 0.12205 | 0.005098 | O43865 | AHCYL1 | 1492 |
| 31 | 18 | 25.18788 | 0.128274 | 0.005093 | P48643 | CCT5 | 1493 |
| 18 | 7 | 23.23158 | 0.118299 | 0.005092 | P04180 | LCAT | 1494 |
| 51 | 3 | 24.94135 | 0.126929 | 0.005089 | O14950 | MYL12B | 1495 |
| 38 | 17 | 24.55361 | 0.124942 | 0.005089 | P49411 | TUFM | 1496 |
| 23 | 8 | 23.66611 | 0.120415 | 0.005088 | P09496 | CLTA | 1497 |
| 18 | 9 | 25.33236 | 0.128687 | 0.00508 | P49908 | SELENOP | 1498 |
| 8 | 3 | 22.65327 | 0.115048 | 0.005079 | Q6PL24 | TMED8 | 1499 |
| 16 | 3 | 22.36951 | 0.113527 | 0.005075 | Q9BRG1 | VPS25 | 1500 |
| 24 | 4 | 25.41191 | 0.128673 | 0.005064 | P62873 | GNB1 | 1501 |

| | | | | | | | |
|---|---|---|---|---|---|---|---|
| 68 | 27 | 30.34694 | 0.153467 | 0.005057 | P02649 | APOE | 1502 |
| 4 | 1 | 18.28962 | 0.092411 | 0.005053 | O60762 | DPM1 | 1503 |
| 32 | 15 | 27.45047 | 0.138676 | 0.005052 | P08567 | PLEK | 1504 |
| 21 | 20 | 25.88657 | 0.130722 | 0.00505 | Q70J99 | UNC13D | 1505 |
| 72 | 11 | 27.44516 | 0.138263 | 0.005038 | P62937 | PPIA | 1506 |
| 13 | 3 | 21.81315 | 0.109874 | 0.005037 | Q9NR28 | DIABLO | 1507 |
| 62 | 7 | 23.25248 | 0.117087 | 0.005035 | P30044 | PRDX5 | 1508 |
| 12 | 3 | 20.13728 | 0.101285 | 0.00503 | P00915 | CA1 | 1509 |
| 15 | 3 | 23.78258 | 0.119617 | 0.00503 | Q00765 | REEP5 | 1510 |
| 41 | 4 | 23.75762 | 0.119364 | 0.005024 | P61604 | HSPE1 | 1511 |
| 32 | 17 | 24.43719 | 0.122705 | 0.005021 | Q96HC4 | PDLIM5 | 1512 |
| 1 | 1 | 25.10862 | 0.125974 | 0.005017 | Q5SNT2 | TMEM201 | 1513 |
| 21 | 8 | 26.3441 | 0.132156 | 0.005017 | P26641 | EEF1G | 1514 |
| 5 | 1 | 20.65073 | 0.103393 | 0.005007 | O60636 | TSPAN2 | 1515 |
| 26 | 3 | 20.99355 | 0.104986 | 0.005001 | Q9Y6B6 | SAR1B | 1516 |
| 22 | 16 | 24.69064 | 0.123314 | 0.004994 | P80108 | GPLD1 | 1517 |
| 40 | 17 | 26.32411 | 0.131426 | 0.004993 | P78371 | CCT2 | 1518 |
| 29 | 8 | 20.96205 | 0.104487 | 0.004985 | Q9Y266 | NUDC | 1519 |
| 67 | 10 | 29.02121 | 0.14447 | 0.004978 | P60660 | MYL6 | 1520 |
| 1 | 1 | 20.9319 | 0.104089 | 0.004973 | Q96G01 | BICD1 | 1521 |
| 14 | 8 | 24.86512 | 0.123532 | 0.004968 | P18428 | LBP | 1522 |
| 23 | 7 | 25.97706 | 0.128803 | 0.004958 | P51884 | LUM | 1523 |
| 41 | 11 | 23.96591 | 0.118799 | 0.004957 | Q14165 | MLEC | 1524 |
| 19 | 10 | 24.70536 | 0.122138 | 0.004944 | P40197 | GP5 | 1525 |
| 9 | 11 | 23.51157 | 0.116013 | 0.004934 | O43306 | ADCY6 | 1526 |
| 37 | 6 | 23.57772 | 0.116243 | 0.00493 | P51452 | DUSP3 | 1527 |
| 52 | 7 | 26.71274 | 0.131481 | 0.004922 | P21291 | CSRP1 | 1528 |
| 39 | 12 | 25.08986 | 0.123031 | 0.004904 | O75558 | STX11 | 1529 |
| 54 | 14 | 27.6437 | 0.135449 | 0.0049 | O00151 | PDLIM1 | 1530 |
| 42 | 3 | 28.5861 | 0.139996 | 0.004897 | P61006 | RAB8A | 1531 |
| 21 | 8 | 23.70723 | 0.116082 | 0.004896 | P11766 | ADH5 | 1532 |
| 22 | 9 | 25.09367 | 0.122816 | 0.004894 | P55058 | PLTP | 1533 |
| 36 | 10 | 28.41269 | 0.138888 | 0.004888 | P78417 | GSTO1 | 1534 |
| 51 | 11 | 25.5166 | 0.124567 | 0.004882 | P00918 | CA2 | 1535 |
| 39 | 13 | 24.85561 | 0.121154 | 0.004874 | P13646 | KRT13 | 1536 |
| 21 | 33 | 24.71903 | 0.120222 | 0.004864 | Q9NYU2 | UGGT1 | 1537 |
| 18 | 10 | 22.02787 | 0.107005 | 0.004858 | Q05655 | PRKCD | 1538 |
| 25 | 11 | 24.31184 | 0.118089 | 0.004857 | P00505 | GOT2 | 1539 |
| 39 | 16 | 24.91548 | 0.12097 | 0.004855 | P10644 | PRKAR1A | 1540 |
| 30 | 3 | 24.60925 | 0.119326 | 0.004849 | P15153 | RAC2 | 1541 |
| 29 | 2 | 24.12134 | 0.11676 | 0.004841 | P07948 | LYN | 1542 |
| 25 | 7 | 23.71059 | 0.114745 | 0.004839 | Q9UGM5 | FETUB | 1543 |
| 29 | 8 | 24.36352 | 0.117633 | 0.004828 | Q16698 | DECR1 | 1544 |
| 17 | 9 | 21.99372 | 0.105948 | 0.004817 | O00116 | AGPS | 1545 |
| 19 | 10 | 23.31863 | 0.111987 | 0.004802 | P52306 | RAP1GDS1 | 1546 |
| 48 | 16 | 26.16775 | 0.125641 | 0.004801 | P00338 | LDHA | 1547 |
| 13 | 7 | 24.05349 | 0.115412 | 0.004798 | P10619 | CTSA | 1548 |

| | | | | | | | |
|---|---|---|---|---|---|---|---|
| 12 | 3 | 21.68064 | 0.103864 | 0.004791 | O75822 | EIF3J | 1549 |
| 25 | 16 | 26.69695 | 0.127716 | 0.004784 | P02748 | C9 | 1550 |
| 10 | 4 | 18.87629 | 0.090067 | 0.004771 | Q13217 | DNAJC3 | 1551 |
| 29 | 10 | 23.47896 | 0.112003 | 0.00477 | Q7Z434 | MAVS | 1552 |
| 26 | 46 | 26.44061 | 0.125588 | 0.00475 | P01031 | C5 | 1553 |
| 3 | 1 | 21.85951 | 0.103786 | 0.004748 | P31995 | FCGR2C | 1554 |
| 33 | 18 | 25.85024 | 0.122641 | 0.004744 | P04040 | CAT | 1555 |
| 41 | 10 | 27.09439 | 0.127367 | 0.004701 | P01876 | IGHA1 | 1556 |
| 20 | 11 | 24.8103 | 0.116556 | 0.004698 | P16452 | EPB42 | 1557 |
| 3 | 1 | 19.50813 | 0.091491 | 0.00469 | P68402 | PAFAH1B2 | 1558 |
| 37 | 22 | 29.16837 | 0.136788 | 0.00469 | P10909 | CLU | 1559 |
| 28 | 4 | 26.37063 | 0.123667 | 0.00469 | P62158 | CALM1 | 1560 |
| 34 | 16 | 26.69727 | 0.125016 | 0.004683 | P48735 | IDH2 | 1561 |
| 26 | 11 | 24.61595 | 0.11522 | 0.004681 | P49189 | ALDH9A1 | 1562 |
| 35 | 23 | 25.70773 | 0.120319 | 0.00468 | Q9NTJ5 | SACM1L | 1563 |
| 35 | 26 | 26.6929 | 0.124552 | 0.004666 | P55072 | VCP | 1564 |
| 19 | 3 | 22.00781 | 0.102551 | 0.00466 | P07947 | YES1 | 1565 |
| 17 | 18 | 25.87801 | 0.12052 | 0.004657 | P00450 | CP | 1566 |
| 19 | 7 | 25.65967 | 0.119093 | 0.004641 | Q9H223 | EHD4 | 1567 |
| 17 | 8 | 24.67035 | 0.114459 | 0.00464 | P08697 | SERPINF2 | 1568 |
| 19 | 91 | 25.90997 | 0.120185 | 0.004639 | Q15149 | PLEC | 1569 |
| 37 | 8 | 23.77794 | 0.110246 | 0.004636 | Q13162 | PRDX4 | 1570 |
| 17 | 16 | 24.28725 | 0.112538 | 0.004634 | P55786 | NPEPPS | 1571 |
| 15 | 5 | 22.84647 | 0.105646 | 0.004624 | Q13011 | ECH1 | 1572 |
| 21 | 4 | 24.97549 | 0.115469 | 0.004623 | Q15363 | TMED2 | 1573 |
| 25 | 27 | 25.50344 | 0.117524 | 0.004608 | Q92619 | ARHGAP45 | 1574 |
| 12 | 7 | 23.84128 | 0.109794 | 0.004605 | P00748 | F12 | 1575 |
| 44 | 10 | 28.43472 | 0.130834 | 0.004601 | Q15907 | RAB11B | 1576 |
| 2 | 2 | 22.82233 | 0.104976 | 0.0046 | Q9UIA9 | XPO7 | 1577 |
| 27 | 18 | 25.63616 | 0.117706 | 0.004591 | P00751 | CFB | 1578 |
| 13 | 4 | 22.39274 | 0.102579 | 0.004581 | Q16775 | HAGH | 1579 |
| 24 | 17 | 25.21174 | 0.115107 | 0.004566 | O14672 | ADAM10 | 1580 |
| 49 | 25 | 27.78426 | 0.126679 | 0.004559 | P26038 | MSN | 1581 |
| 18 | 10 | 23.41741 | 0.106742 | 0.004558 | P39656 | DDOST | 1582 |
| 20 | 7 | 24.57319 | 0.11177 | 0.004548 | Q15084 | PDIA6 | 1583 |
| 23 | 11 | 24.76708 | 0.111821 | 0.004515 | P20073 | ANXA7 | 1584 |
| 27 | 14 | 27.50771 | 0.124084 | 0.004511 | P14314 | PRKCSH | 1585 |
| 8 | 3 | 21.97346 | 0.099058 | 0.004508 | Q9NZP8 | C1RL | 1586 |
| 35 | 11 | 27.50604 | 0.123899 | 0.004504 | P35232 | PHB | 1587 |
| 41 | 26 | 26.11128 | 0.117593 | 0.004504 | Q13586 | STIM1 | 1588 |
| 9 | 2 | 27.44725 | 0.123378 | 0.004495 | Q9H0E2 | TOLLIP | 1589 |
| 36 | 4 | 21.89273 | 0.098349 | 0.004492 | P09488 | GSTM1 | 1590 |
| 6 | 5 | 25.30607 | 0.11326 | 0.004476 | P09327 | VIL1 | 1591 |
| 10 | 5 | 20.39993 | 0.090893 | 0.004456 | Q9HD45 | TM9SF3 | 1592 |
| 23 | 15 | 23.905 | 0.10639 | 0.004451 | P06737 | PYGL | 1593 |
| 11 | 7 | 21.80202 | 0.096707 | 0.004436 | Q9Y5K6 | CD2AP | 1594 |
| 19 | 5 | 22.26278 | 0.098613 | 0.004429 | O14618 | CCS | 1595 |

| | | | | | | | |
|---|---|---|---|---|---|---|---|
| 16 | 5 | 24.68843 | 0.109001 | 0.004415 | O95445 | APOM | 1596 |
| 25 | 5 | 24.47899 | 0.107862 | 0.004406 | P02747 | C1QC | 1597 |
| 41 | 11 | 27.55969 | 0.121417 | 0.004406 | Q99439 | CNN2 | 1598 |
| 62 | 7 | 23.66852 | 0.104224 | 0.004403 | O75964 | ATP5MG | 1599 |
| 0 | 1 | 22.01867 | 0.096568 | 0.004386 | Q86YA3 | ZGRF1 | 1600 |
| 30 | 15 | 25.46519 | 0.111279 | 0.00437 | Q14520 | HABP2 | 1601 |
| 34 | 71 | 28.61594 | 0.124832 | 0.004362 | P11277 | SPTB | 1602 |
| 6 | 12 | 21.18957 | 0.092213 | 0.004352 | Q8NF50 | DOCK8 | 1603 |
| 51 | 8 | 26.10023 | 0.113488 | 0.004348 | P61026 | RAB10 | 1604 |
| 37 | 11 | 23.30354 | 0.100959 | 0.004332 | P12429 | ANXA3 | 1605 |
| 43 | 18 | 25.51054 | 0.110482 | 0.004331 | Q9UJU6 | DBNL | 1606 |
| 4 | 1 | 19.7562 | 0.085544 | 0.00433 | Q92882 | OSTF1 | 1607 |
| 46 | 11 | 26.64486 | 0.115222 | 0.004324 | P04899 | GNAI2 | 1608 |
| 39 | 17 | 28.31463 | 0.122325 | 0.00432 | P11142 | HSPA8 | 1609 |
| 20 | 13 | 24.01399 | 0.103696 | 0.004318 | Q07866 | KLC1 | 1610 |
| 28 | 5 | 27.50519 | 0.118397 | 0.004305 | P60953 | CDC42 | 1611 |
| 50 | 7 | 28.47689 | 0.122451 | 0.0043 | P68366 | TUBA4A | 1612 |
| 18 | 1 | 25.56227 | 0.109592 | 0.004287 | Q92686 | NRGN | 1613 |
| 32 | 7 | 24.93685 | 0.106911 | 0.004287 | P62826 | RAN | 1614 |
| 48 | 11 | 25.91301 | 0.110972 | 0.004282 | Q15691 | MAPRE1 | 1615 |
| 17 | 14 | 23.50092 | 0.100613 | 0.004281 | Q14974 | KPNB1 | 1616 |
| 60 | 9 | 24.29643 | 0.103797 | 0.004272 | P47755 | CAPZA2 | 1617 |
| 56 | 42 | 28.8966 | 0.123196 | 0.004263 | P02787 | TF | 1618 |
| 39 | 13 | 26.27255 | 0.112001 | 0.004263 | O43866 | CD5L | 1619 |
| 13 | 10 | 23.48254 | 0.099932 | 0.004256 | P11171 | EPB41 | 1620 |
| 28 | 17 | 25.88368 | 0.109918 | 0.004247 | Q01813 | PFKP | 1621 |
| 30 | 8 | 24.33863 | 0.103223 | 0.004241 | P25311 | AZGP1 | 1622 |
| 39 | 42 | 26.97266 | 0.114164 | 0.004233 | P08603 | CFH | 1623 |
| 12 | 4 | 25.01305 | 0.105774 | 0.004229 | P51692 | STAT5B | 1624 |
| 26 | 21 | 24.12066 | 0.101879 | 0.004224 | Q7KZF4 | SND1 | 1625 |
| 30 | 15 | 25.34471 | 0.106939 | 0.004219 | Q9UDY2 | TJP2 | 1626 |
| 28 | 2 | 26.56187 | 0.111972 | 0.004216 | Q03591 | CFHR1 | 1627 |
| 35 | 9 | 24.20401 | 0.101783 | 0.004205 | P13804 | ETFA | 1628 |
| 5 | 4 | 24.1005 | 0.101266 | 0.004202 | Q99805 | TM9SF2 | 1629 |
| 7 | 25 | 29.34119 | 0.123262 | 0.004201 | P46939 | UTRN | 1630 |
| 18 | 14 | 24.1013 | 0.101224 | 0.0042 | P43652 | AFM | 1631 |
| 15 | 2 | 21.06175 | 0.088246 | 0.00419 | Q99576 | TSC22D3 | 1632 |
| 21 | 12 | 25.87466 | 0.108303 | 0.004186 | P06744 | GPI | 1633 |
| 22 | 4 | 25.78739 | 0.107815 | 0.004181 | P02763 | ORM1 | 1634 |
| 7 | 2 | 21.99621 | 0.091874 | 0.004177 | P61073 | CXCR4 | 1635 |
| 22 | 24 | 26.10759 | 0.108665 | 0.004162 | Q9Y613 | FHOD1 | 1636 |
| 29 | 13 | 24.69678 | 0.102529 | 0.004152 | Q00013 | MPP1 | 1637 |
| 59 | 11 | 27.37036 | 0.113627 | 0.004151 | P61106 | RAB14 | 1638 |
| 37 | 6 | 22.84904 | 0.094824 | 0.00415 | Q9NVJ2 | ARL8B | 1639 |
| 11 | 5 | 24.10489 | 0.099991 | 0.004148 | P22792 | CPN2 | 1640 |
| 49 | 7 | 29.61388 | 0.12281 | 0.004147 | P01860 | IGHG3 | 1641 |
| 19 | 23 | 24.5517 | 0.101778 | 0.004145 | P29144 | TPP2 | 1642 |

| | | | | | | | |
|---|---|---|---|---|---|---|---|
| 42 | 8 | 24.36298 | 0.100977 | 0.004145 | O95866 | MPIG6B | 1643 |
| 33 | 23 | 25.50175 | 0.104453 | 0.004096 | P51659 | HSD17B4 | 1644 |
| 41 | 3 | 24.73066 | 0.101228 | 0.004093 | P01619 | IGKV3-20 | 1645 |
| 27 | 4 | 24.8692 | 0.10126 | 0.004072 | P63241 | EIF5A | 1646 |
| 2 | 2 | 21.07512 | 0.085736 | 0.004068 | Q9NTJ4 | MAN2C1 | 1647 |
| 39 | 8 | 23.79761 | 0.096708 | 0.004064 | Q9Y2Q3 | GSTK1 | 1648 |
| 25 | 10 | 23.37536 | 0.094936 | 0.004061 | Q9BT78 | COPS4 | 1649 |
| 75 | 14 | 32.94168 | 0.133768 | 0.004061 | P60709 | ACTB | 1650 |
| 15 | 7 | 21.56351 | 0.08756 | 0.004061 | P07093 | SERPINE2 | 1651 |
| 61 | 12 | 28.84296 | 0.11704 | 0.004058 | P23528 | CFL1 | 1652 |
| 25 | 18 | 25.82763 | 0.104615 | 0.00405 | P04843 | RPN1 | 1653 |
| 27 | 16 | 24.22567 | 0.098106 | 0.00405 | P55084 | HADHB | 1654 |
| 9 | 13 | 23.78603 | 0.095853 | 0.00403 | Q96RT1 | ERBIN | 1655 |
| 44 | 48 | 28.82317 | 0.115993 | 0.004024 | P07996 | THBS1 | 1656 |
| 14 | 66 | 24.46039 | 0.098219 | 0.004015 | Q14204 | DYNC1H1 | 1657 |
| 25 | 18 | 26.61413 | 0.106661 | 0.004008 | O43182 | ARHGAP6 | 1658 |
| 37 | 19 | 26.36552 | 0.105651 | 0.004007 | P10809 | HSPD1 | 1659 |
| 36 | 10 | 22.80594 | 0.091266 | 0.004002 | Q9H425 | C1orf198 | 1660 |
| 50 | 8 | 27.03702 | 0.107867 | 0.00399 | P01861 | IGHG4 | 1661 |
| 18 | 16 | 24.79523 | 0.098892 | 0.003988 | P22314 | UBA1 | 1662 |
| 20 | 5 | 22.5877 | 0.089864 | 0.003978 | P49721 | PSMB2 | 1663 |
| 33 | 10 | 27.17091 | 0.107475 | 0.003956 | P00387 | CYB5R3 | 1664 |
| 33 | 5 | 21.99935 | 0.086893 | 0.00395 | Q9UMX5 | NENF | 1665 |
| 70 | 8 | 27.64505 | 0.108821 | 0.003936 | P06702 | S100A9 | 1666 |
| 29 | 16 | 25.68862 | 0.101049 | 0.003934 | P68104 | EEF1A1 | 1667 |
| 22 | 6 | 22.21047 | 0.087103 | 0.003922 | P25789 | PSMA4 | 1668 |
| 27 | 11 | 24.05026 | 0.094311 | 0.003921 | P09543 | CNP | 1669 |
| 27 | 11 | 25.3953 | 0.09938 | 0.003913 | P19086 | GNAZ | 1670 |
| 39 | 4 | 28.53788 | 0.111643 | 0.003912 | P02652 | APOA2 | 1671 |
| 58 | 4 | 27.55361 | 0.10744 | 0.003899 | P61224 | RAP1B | 1672 |
| 49 | 83 | 29.25557 | 0.113975 | 0.003896 | P01024 | C3 | 1673 |
| 43 | 3 | 28.6259 | 0.111089 | 0.003881 | P68371 | TUBB4B | 1674 |
| 3 | 10 | 24.32995 | 0.094389 | 0.00388 | Q6ZQQ6 | WDR87 | 1675 |
| 37 | 8 | 27.28489 | 0.105805 | 0.003878 | P14618 | PKM | 1676 |
| 1 | 4 | 22.65582 | 0.087838 | 0.003877 | O94915 | FRYL | 1677 |
| 14 | 17 | 25.91664 | 0.100398 | 0.003874 | O60610 | DIAPH1 | 1678 |
| 10 | 8 | 23.51372 | 0.090964 | 0.003869 | Q6PJW8 | CNST | 1679 |
| 26 | 28 | 25.31977 | 0.097945 | 0.003868 | Q9Y6C2 | EMILIN1 | 1680 |
| 76 | 26 | 32.05339 | 0.123973 | 0.003868 | P02647 | APOA1 | 1681 |
| 17 | 4 | 21.98002 | 0.084997 | 0.003867 | O75608 | LYPLA1 | 1682 |
| 51 | 12 | 24.37835 | 0.094119 | 0.003861 | Q06323 | PSME1 | 1683 |
| 31 | 9 | 29.1117 | 0.112163 | 0.003853 | P02765 | AHSG | 1684 |
| 8 | 7 | 23.41368 | 0.090077 | 0.003847 | Q8TDZ2 | MICAL1 | 1685 |
| 17 | 4 | 25.44241 | 0.097536 | 0.003834 | O00264 | PGRMC1 | 1686 |
| 27 | 14 | 27.0051 | 0.103215 | 0.003822 | O14791 | APOL1 | 1687 |
| 35 | 23 | 25.80799 | 0.098392 | 0.003812 | Q8N392 | ARHGAP18 | 1688 |
| 32 | 16 | 24.82536 | 0.094613 | 0.003811 | P50991 | CCT4 | 1689 |

| | | | | | | | |
|---|---|---|---|---|---|---|---|
| 17 | 11 | 23.55465 | 0.089686 | 0.003808 | Q96G03 | PGM2 | 1690 |
| 20 | 14 | 27.54423 | 0.104718 | 0.003802 | Q14155 | ARHGEF7 | 1691 |
| 30 | 14 | 27.4529 | 0.104104 | 0.003792 | P07900 | HSP90AA1 | 1692 |
| 26 | 11 | 23.72333 | 0.089933 | 0.003791 | Q9UQ80 | PA2G4 | 1693 |
| 14 | 26 | 24.21405 | 0.091515 | 0.003779 | Q01082 | SPTBN1 | 1694 |
| 26 | 10 | 24.55798 | 0.092237 | 0.003756 | P31930 | UQCRC1 | 1695 |
| 13 | 2 | 19.64037 | 0.0736 | 0.003747 | P03950 | ANG | 1696 |
| 21 | 51 | 27.48272 | 0.102764 | 0.003739 | P04275 | VWF | 1697 |
| 72 | 5 | 30.78677 | 0.115044 | 0.003737 | P02656 | APOC3 | 1698 |
| 16 | 8 | 26.83755 | 0.100039 | 0.003728 | P13224 | GP1BB | 1699 |
| 40 | 19 | 25.82917 | 0.095949 | 0.003715 | Q9H939 | PSTPIP2 | 1700 |
| 28 | 8 | 24.87323 | 0.091986 | 0.003698 | P04632 | CAPNS1 | 1701 |
| 9 | 17 | 27.88942 | 0.103091 | 0.003696 | Q9Y4I1 | MYO5A | 1702 |
| 5 | 1 | 19.1639 | 0.070612 | 0.003685 | Q07021 | C1QBP | 1703 |
| 1 | 1 | 19.85755 | 0.073105 | 0.003681 | O75891 | ALDH1L1 | 1704 |
| 36 | 17 | 26.05155 | 0.095449 | 0.003664 | P00734 | F2 | 1705 |
| 31 | 4 | 26.46595 | 0.09669 | 0.003653 | P63000 | RAC1 | 1706 |
| 37 | 8 | 23.60249 | 0.086138 | 0.00365 | P60900 | PSMA6 | 1707 |
| 34 | 20 | 25.96728 | 0.094468 | 0.003638 | P31948 | STIP1 | 1708 |
| 24 | 3 | 23.74847 | 0.086302 | 0.003634 | P18085 | ARF4 | 1709 |
| 27 | 19 | 24.41267 | 0.08827 | 0.003616 | Q99798 | ACO2 | 1710 |
| 29 | 16 | 24.82168 | 0.089743 | 0.003615 | O60749 | SNX2 | 1711 |
| 35 | 21 | 27.22951 | 0.098223 | 0.003607 | P03952 | KLKB1 | 1712 |
| 7 | 5 | 22.35768 | 0.080544 | 0.003603 | Q99501 | GAS2L1 | 1713 |
| 17 | 8 | 24.25663 | 0.08708 | 0.00359 | Q9Y394 | DHRS7 | 1714 |
| 18 | 9 | 23.77546 | 0.085255 | 0.003586 | Q9P2R7 | SUCLA2 | 1715 |
| 9 | 6 | 19.93343 | 0.071426 | 0.003583 | O43252 | PAPSS1 | 1716 |
| 33 | 26 | 24.28423 | 0.086936 | 0.00358 | Q9Y6F6 | IRAG1 | 1717 |
| 23 | 5 | 26.55965 | 0.094623 | 0.003563 | P30048 | PRDX3 | 1718 |
| 28 | 5 | 23.29753 | 0.082929 | 0.00356 | O60493 | SNX3 | 1719 |
| 38 | 22 | 26.5947 | 0.094509 | 0.003554 | P24557 | TBXAS1 | 1720 |
| 27 | 16 | 26.0736 | 0.092312 | 0.00354 | P18054 | ALOX12 | 1721 |
| 27 | 11 | 25.03475 | 0.088614 | 0.00354 | O94919 | ENDOD1 | 1722 |
| 16 | 5 | 23.59602 | 0.083162 | 0.003524 | Q8N699 | MYCT1 | 1723 |
| 38 | 12 | 25.20603 | 0.088608 | 0.003515 | Q9BX67 | JAM3 | 1724 |
| 12 | 16 | 25.22292 | 0.088208 | 0.003497 | Q9C0C9 | UBE2O | 1725 |
| 26 | 4 | 24.19528 | 0.084461 | 0.003491 | Q99719 | SEPTIN5 | 1726 |
| 45 | 15 | 27.05863 | 0.094442 | 0.00349 | P50552 | VASP | 1727 |
| 33 | 3 | 25.4699 | 0.088636 | 0.00348 | P30273 | FCER1G | 1728 |
| 14 | 4 | 25.0517 | 0.087179 | 0.00348 | P35613 | BSG | 1729 |
| 43 | 4 | 25.99233 | 0.090404 | 0.003478 | P31946 | YWHAB | 1730 |
| 30 | 10 | 24.46982 | 0.085106 | 0.003478 | Q6Q788 | APOA5 | 1731 |
| 30 | 22 | 25.21175 | 0.087445 | 0.003468 | P40939 | HADHA | 1732 |
| 49 | 13 | 26.35171 | 0.091385 | 0.003468 | P47756 | CAPZB | 1733 |
| 11 | 10 | 24.8271 | 0.085783 | 0.003455 | P45974 | USP5 | 1734 |
| 28 | 8 | 27.12978 | 0.093711 | 0.003454 | P27169 | PON1 | 1735 |
| 2 | 1 | 19.70658 | 0.067732 | 0.003437 | O75683 | SURF6 | 1736 |

| | | | | | | | |
|---|---|---|---|---|---|---|---|
| 19 | 16 | 24.04091 | 0.081737 | 0.0034 | O60763 | USO1 | 1737 |
| 5 | 2 | 24.19107 | 0.082171 | 0.003397 | Q08722 | CD47 | 1738 |
| 23 | 16 | 24.80888 | 0.084236 | 0.003395 | P50416 | CPT1A | 1739 |
| 4 | 26 | 25.71308 | 0.087256 | 0.003393 | Q5T4S7 | UBR4 | 1740 |
| 37 | 13 | 22.6732 | 0.076929 | 0.003393 | P50453 | SERPINB9 | 1741 |
| 39 | 26 | 26.73915 | 0.089987 | 0.003365 | P16284 | PECAM1 | 1742 |
| 28 | 6 | 25.83578 | 0.086896 | 0.003363 | Q13103 | SPP2 | 1743 |
| 29 | 17 | 29.32057 | 0.098233 | 0.00335 | P01042 | KNG1 | 1744 |
| 14 | 10 | 23.69433 | 0.078786 | 0.003325 | Q9Y678 | COPG1 | 1745 |
| 20 | 9 | 27.12025 | 0.089982 | 0.003318 | P0DMV8 | HSPA1A | 1746 |
| 35 | 15 | 26.43602 | 0.087706 | 0.003318 | P01011 | SERPINA3 | 1747 |
| 49 | 8 | 25.27662 | 0.083779 | 0.003315 | Q15365 | PCBP1 | 1748 |
| 38 | 8 | 24.06873 | 0.079753 | 0.003314 | P01892 | HLA-A | 1749 |
| 31 | 6 | 23.73167 | 0.07841 | 0.003304 | Q99714 | HSD17B10 | 1750 |
| 18 | 6 | 24.43989 | 0.080743 | 0.003304 | P0C7P4 | UQCRFS1P1 | 1751 |
| 29 | 40 | 29.59496 | 0.097447 | 0.003293 | Q14766 | LTBP1 | 1752 |
| 5 | 15 | 23.59343 | 0.077636 | 0.003291 | P12111 | COL6A3 | 1753 |
| 29 | 26 | 26.519 | 0.087202 | 0.003288 | P19367 | HK1 | 1754 |
| 28 | 26 | 27.00527 | 0.088523 | 0.003278 | O76074 | PDE5A | 1755 |
| 7 | 3 | 19.7558 | 0.064752 | 0.003278 | Q9NWZ3 | IRAK4 | 1756 |
| 33 | 6 | 25.06531 | 0.081961 | 0.00327 | P84077 | ARF1 | 1757 |
| 23 | 18 | 26.41888 | 0.085818 | 0.003248 | P02730 | SLC4A1 | 1758 |
| 8 | 6 | 30.46275 | 0.098188 | 0.003223 | O00712 | NFIB | 1759 |
| 39 | 13 | 25.86256 | 0.082805 | 0.003202 | O15144 | ARPC2 | 1760 |
| 31 | 17 | 26.61237 | 0.084626 | 0.00318 | Q16643 | DBN1 | 1761 |
| 26 | 12 | 25.49238 | 0.080569 | 0.003161 | P23219 | PTGS1 | 1762 |
| 14 | 8 | 24.15358 | 0.076184 | 0.003154 | Q13045 | FLII | 1763 |
| 29 | 10 | 23.68098 | 0.074543 | 0.003148 | O43639 | NCK2 | 1764 |
| 44 | 15 | 27.48926 | 0.086143 | 0.003134 | P07195 | LDHB | 1765 |
| 24 | 7 | 24.95098 | 0.077974 | 0.003125 | P25325 | MPST | 1766 |
| 14 | 6 | 23.28928 | 0.072601 | 0.003117 | P07339 | CTSD | 1767 |
| 18 | 10 | 24.94814 | 0.077541 | 0.003108 | P15170 | GSPT1 | 1768 |
| 56 | 5 | 29.82735 | 0.09233 | 0.003095 | P02655 | APOC2 | 1769 |
| 24 | 15 | 28.11593 | 0.086422 | 0.003074 | P07359 | GP1BA | 1770 |
| 27 | 10 | 29.71003 | 0.091321 | 0.003074 | P0C0L5 | C4B | 1771 |
| 51 | 19 | 26.89218 | 0.082511 | 0.003068 | P00558 | PGK1 | 1772 |
| 14 | 8 | 23.99036 | 0.073192 | 0.003051 | P07357 | C8A | 1773 |
| 37 | 22 | 26.65605 | 0.080819 | 0.003032 | Q15833 | STXBP2 | 1774 |
| 30 | 7 | 25.9802 | 0.078706 | 0.003029 | P00491 | PNP | 1775 |
| 33 | 15 | 25.39824 | 0.076769 | 0.003023 | P52209 | PGD | 1776 |
| 50 | 5 | 25.09127 | 0.075519 | 0.00301 | P20340 | RAB6A | 1777 |
| 40 | 11 | 24.35494 | 0.07327 | 0.003008 | P45880 | VDAC2 | 1778 |
| 54 | 3 | 28.14269 | 0.084401 | 0.002999 | P09493 | TPM1 | 1779 |
| 59 | 11 | 29.94752 | 0.089494 | 0.002988 | P37802 | TAGLN2 | 1780 |
| 22 | 6 | 27.19889 | 0.081186 | 0.002985 | P46109 | CRKL | 1781 |
| 19 | 4 | 20.62647 | 0.06144 | 0.002979 | P22352 | GPX3 | 1782 |
| 76 | 12 | 29.32997 | 0.087342 | 0.002978 | P07737 | PFN1 | 1783 |

| | | | | | | | |
|---|---|---|---|---|---|---|---|
| 42 | 21 | 26.77877 | 0.079717 | 0.002977 | P50990 | CCT8 | 1784 |
| 47 | 14 | 25.69001 | 0.07647 | 0.002977 | O15143 | ARPC1B | 1785 |
| 27 | 19 | 27.22793 | 0.080778 | 0.002967 | P16109 | SELP | 1786 |
| 51 | 25 | 27.60558 | 0.081789 | 0.002963 | Q14247 | CTTN | 1787 |
| 21 | 9 | 25.35007 | 0.075032 | 0.00296 | Q00325 | SLC25A3 | 1788 |
| 84 | 15 | 25.45765 | 0.075057 | 0.002948 | P52907 | CAPZA1 | 1789 |
| 26 | 8 | 25.93545 | 0.076253 | 0.00294 | Q9Y6E0 | STK24 | 1790 |
| 26 | 6 | 25.3976 | 0.074355 | 0.002928 | P11234 | RALB | 1791 |
| 15 | 8 | 24.32892 | 0.070578 | 0.002901 | Q14162 | SCARF1 | 1792 |
| 82 | 6 | 29.2778 | 0.084878 | 0.002899 | P68871 | HBB | 1793 |
| 18 | 4 | 24.34466 | 0.07051 | 0.002896 | P22061 | PCMT1 | 1794 |
| 13 | 12 | 25.51733 | 0.073899 | 0.002896 | Q6P3W7 | SCYL2 | 1795 |
| 41 | 16 | 25.95082 | 0.075043 | 0.002892 | P12931 | SRC | 1796 |
| 45 | 21 | 29.62739 | 0.085372 | 0.002882 | P01871 | IGHM | 1797 |
| 34 | 11 | 25.97417 | 0.074739 | 0.002877 | Q9NYL9 | TMOD3 | 1798 |
| 50 | 21 | 27.57258 | 0.079254 | 0.002874 | P06733 | ENO1 | 1799 |
| 5 | 3 | 24.58018 | 0.070647 | 0.002874 | O95573 | ACSL3 | 1800 |
| 7 | 2 | 24.43719 | 0.07012 | 0.002869 | Q13790 | APOF | 1801 |
| 44 | 28 | 27.94878 | 0.080144 | 0.002868 | P11021 | HSPA5 | 1802 |
| 22 | 22 | 26.56996 | 0.076136 | 0.002866 | Q01433 | AMPD2 | 1803 |
| 60 | 119 | 32.3813 | 0.092647 | 0.002861 | Q9Y490 | TLN1 | 1804 |
| 67 | 7 | 26.17047 | 0.074874 | 0.002861 | O15511 | ARPC5 | 1805 |
| 22 | 8 | 25.40994 | 0.071918 | 0.00283 | P11310 | ACADM | 1806 |
| 63 | 15 | 30.72021 | 0.086804 | 0.002826 | P06753 | TPM3 | 1807 |
| 38 | 16 | 27.30382 | 0.076845 | 0.002814 | P61160 | ACTR2 | 1808 |
| 34 | 17 | 25.37126 | 0.071146 | 0.002804 | P29350 | PTPN6 | 1809 |
| 32 | 18 | 25.04325 | 0.070193 | 0.002803 | Q14677 | CLINT1 | 1810 |
| 18 | 14 | 27.43641 | 0.076526 | 0.002789 | P50851 | LRBA | 1811 |
| 40 | 5 | 24.97639 | 0.069569 | 0.002785 | Q14019 | COTL1 | 1812 |
| 4 | 5 | 21.68378 | 0.060385 | 0.002785 | B0I1T2 | MYO1G | 1813 |
| 33 | 5 | 22.21122 | 0.06162 | 0.002774 | Q9HBL7 | PLGRKT | 1814 |
| 38 | 29 | 27.77416 | 0.076891 | 0.002768 | Q5JSH3 | WDR44 | 1815 |
| 6 | 4 | 21.67612 | 0.059894 | 0.002763 | Q8N5J2 | MINDY1 | 1816 |
| 34 | 36 | 27.26072 | 0.074895 | 0.002747 | P23229 | ITGA6 | 1817 |
| 25 | 55 | 27.58179 | 0.075766 | 0.002747 | P12259 | F5 | 1818 |
| 37 | 13 | 27.46618 | 0.075397 | 0.002745 | P48059 | LIMS1 | 1819 |
| 27 | 12 | 25.93538 | 0.071112 | 0.002742 | Q16181 | SEPTIN7 | 1820 |
| 3 | 2 | 23.11475 | 0.063091 | 0.002729 | O95674 | CDS2 | 1821 |
| 41 | 13 | 24.28425 | 0.065995 | 0.002718 | O14745 | SLC9A3R1 | 1822 |
| 77 | 47 | 31.64323 | 0.085949 | 0.002716 | P02675 | FGB | 1823 |
| 20 | 5 | 21.87585 | 0.059199 | 0.002706 | Q9UEU0 | VTI1B | 1824 |
| 22 | 9 | 25.6641 | 0.069361 | 0.002703 | P50502 | ST13 | 1825 |
| 24 | 5 | 22.16332 | 0.059542 | 0.002687 | P06730 | EIF4E | 1826 |
| 9 | 8 | 24.37536 | 0.065463 | 0.002686 | Q8TD19 | NEK9 | 1827 |
| 17 | 5 | 24.05862 | 0.064579 | 0.002684 | Q9H3N1 | TMX1 | 1828 |
| 53 | 33 | 28.11402 | 0.075378 | 0.002681 | P00747 | PLG | 1829 |
| 48 | 7 | 26.1997 | 0.070142 | 0.002677 | P62987 | UBA52 | 1830 |

| | | | | | | | |
|---|---|---|---|---|---|---|---|
| 39 | 35 | 28.81307 | 0.077097 | 0.002676 | P14625 | HSP90B1 | 1831 |
| 55 | 33 | 28.53007 | 0.076241 | 0.002672 | Q05682 | CALD1 | 1832 |
| 57 | 9 | 26.16415 | 0.069683 | 0.002663 | Q06830 | PRDX1 | 1833 |
| 26 | 17 | 25.54669 | 0.067762 | 0.002652 | Q8NEU8 | APPL2 | 1834 |
| 41 | 10 | 23.76023 | 0.062742 | 0.002641 | Q9UIJ7 | AK3 | 1835 |
| 34 | 68 | 30.02347 | 0.078725 | 0.002622 | P02751 | FN1 | 1836 |
| 40 | 18 | 24.97504 | 0.065437 | 0.00262 | Q07960 | ARHGAP1 | 1837 |
| 44 | 27 | 28.72232 | 0.074917 | 0.002608 | P04003 | C4BPA | 1838 |
| 51 | 15 | 26.04096 | 0.067684 | 0.002599 | P50395 | GDI2 | 1839 |
| 9 | 3 | 24.87349 | 0.064419 | 0.00259 | P02745 | C1QA | 1840 |
| 36 | 16 | 26.9521 | 0.069616 | 0.002583 | P06576 | ATP5F1B | 1841 |
| 13 | 13 | 25.04741 | 0.064641 | 0.002581 | Q9BSJ8 | ESYT1 | 1842 |
| 59 | 55 | 31.80719 | 0.081775 | 0.002571 | P02671 | FGA | 1843 |
| 70 | 37 | 29.93653 | 0.076936 | 0.00257 | P06727 | APOA4 | 1844 |
| 45 | 19 | 28.39398 | 0.071824 | 0.00253 | O95810 | CAVIN2 | 1845 |
| 6 | 5 | 23.67145 | 0.059507 | 0.002514 | Q8IWA5 | SLC44A2 | 1846 |
| 32 | 2 | 24.49105 | 0.061451 | 0.002509 | A0A0B4J1V | IGHV3-73 | 1847 |
| 24 | 28 | 25.79061 | 0.064501 | 0.002501 | Q12913 | PTPRJ | 1848 |
| 35 | 7 | 24.49677 | 0.061167 | 0.002497 | P62993 | GRB2 | 1849 |
| 37 | 16 | 26.65573 | 0.066497 | 0.002495 | P61158 | ACTR3 | 1850 |
| 28 | 9 | 24.20217 | 0.060092 | 0.002483 | P49407 | ARRB1 | 1851 |
| 47 | 13 | 27.09639 | 0.06708 | 0.002476 | P05534 | HLA-A | 1852 |
| 28 | 3 | 23.14751 | 0.057249 | 0.002473 | P51153 | RAB13 | 1853 |
| 23 | 21 | 27.19562 | 0.066863 | 0.002459 | Q14624 | ITIH4 | 1854 |
| 36 | 32 | 29.32843 | 0.071471 | 0.002437 | P08514 | ITGA2B | 1855 |
| 44 | 7 | 24.83181 | 0.060433 | 0.002434 | P01591 | JCHAIN | 1856 |
| 21 | 23 | 30.30137 | 0.073618 | 0.00243 | Q13423 | NNT | 1857 |
| 29 | 68 | 27.07517 | 0.065678 | 0.002426 | P02549 | SPTA1 | 1858 |
| 39 | 9 | 22.92152 | 0.055434 | 0.002418 | O43396 | TXNL1 | 1859 |
| 24 | 14 | 26.19357 | 0.063103 | 0.002409 | P05546 | SERPIND1 | 1860 |
| 41 | 15 | 27.73869 | 0.066806 | 0.002408 | P04406 | GAPDH | 1861 |
| 20 | 8 | 24.8423 | 0.058017 | 0.002335 | Q6IBS0 | TWF2 | 1862 |
| 24 | 10 | 27.18567 | 0.06307 | 0.00232 | Q9HBI1 | PARVB | 1863 |
| 35 | 5 | 25.62532 | 0.059428 | 0.002319 | Q9NR12 | PDLIM7 | 1864 |
| 36 | 16 | 26.32235 | 0.060316 | 0.002291 | P30740 | SERPINB1 | 1865 |
| 35 | 18 | 26.00278 | 0.059509 | 0.002289 | P36871 | PGM1 | 1866 |
| 31 | 13 | 23.85819 | 0.054127 | 0.002269 | P13489 | RNH1 | 1867 |
| 36 | 21 | 27.29878 | 0.061636 | 0.002258 | P07237 | P4HB | 1868 |
| 45 | 13 | 28.19927 | 0.063103 | 0.002238 | P02749 | APOH | 1869 |
| 19 | 7 | 25.00781 | 0.055381 | 0.002215 | O43488 | AKR7A2 | 1870 |
| 45 | 15 | 27.3373 | 0.060438 | 0.002211 | Q9NZN3 | EHD3 | 1871 |
| 1 | 1 | 17.45488 | 0.038588 | 0.002211 | P15144 | ANPEP | 1872 |
| 4 | 126 | 27.452 | 0.060617 | 0.002208 | Q8WZ42 | TTN | 1873 |
| 20 | 15 | 23.81096 | 0.052482 | 0.002204 | O43150 | ASAP2 | 1874 |
| 71 | 51 | 33.42104 | 0.073392 | 0.002196 | P02768 | ALB | 1875 |
| 29 | 18 | 25.00487 | 0.054518 | 0.00218 | P38646 | HSPA9 | 1876 |
| 18 | 2 | 21.86808 | 0.047558 | 0.002175 | P49773 | HINT1 | 1877 |

| | | | | | | | |
|---|---|---|---|---|---|---|---|
| 48 | 5 | 26.75676 | 0.058112 | 0.002172 | P01859 | IGHG2 | 1878 |
| 37 | 22 | 27.78698 | 0.059767 | 0.002151 | O75083 | WDR1 | 1879 |
| 36 | 13 | 26.0463 | 0.05592 | 0.002147 | P50148 | GNAQ | 1880 |
| 49 | 236 | 31.99771 | 0.068446 | 0.002139 | P04114 | APOB | 1881 |
| 34 | 26 | 25.47534 | 0.054323 | 0.002132 | Q14644 | RASA3 | 1882 |
| 66 | 19 | 28.19371 | 0.060045 | 0.00213 | P04075 | ALDOA | 1883 |
| 69 | 80 | 31.07481 | 0.06554 | 0.002109 | P18206 | VCL | 1884 |
| 28 | 14 | 25.81653 | 0.054176 | 0.002098 | P19971 | TYMP | 1885 |
| 66 | 31 | 31.64428 | 0.065834 | 0.00208 | P02679 | FGG | 1886 |
| 4 | 31 | 24.91441 | 0.051336 | 0.002061 | Q9UPN3 | MACF1 | 1887 |
| 44 | 5 | 24.98956 | 0.051307 | 0.002053 | P27348 | YWHAQ | 1888 |
| 43 | 22 | 26.62426 | 0.054399 | 0.002043 | P25705 | ATP5F1A | 1889 |
| 26 | 12 | 23.38152 | 0.047745 | 0.002042 | Q13884 | SNTB1 | 1890 |
| 67 | 9 | 29.92553 | 0.060952 | 0.002037 | P67936 | TPM4 | 1891 |
| 63 | 12 | 28.47527 | 0.054795 | 0.001924 | P63104 | YWHAZ | 1892 |
| 48 | 15 | 29.06479 | 0.055753 | 0.001918 | Q9H4B7 | TUBB1 | 1893 |
| 47 | 9 | 26.66813 | 0.051123 | 0.001917 | Q04917 | YWHAH | 1894 |
| 49 | 41 | 29.40625 | 0.055987 | 0.001904 | P08519 | LPA | 1895 |
| 53 | 38 | 30.01079 | 0.056738 | 0.001891 | P05106 | ITGB3 | 1896 |
| 5 | 7 | 24.16092 | 0.045563 | 0.001886 | P30622 | CLIP1 | 1897 |
| 36 | 4 | 24.89558 | 0.046088 | 0.001851 | P58546 | MTPN | 1898 |
| 51 | 22 | 28.71829 | 0.051986 | 0.00181 | Q15942 | ZYX | 1899 |
| 35 | 39 | 29.89155 | 0.053077 | 0.001776 | P01023 | A2M | 1900 |
| 51 | 13 | 27.49959 | 0.04875 | 0.001773 | P60174 | TPI1 | 1901 |
| 49 | 5 | 24.62241 | 0.04324 | 0.001756 | P18859 | ATP5PF | 1902 |
| 33 | 27 | 28.22506 | 0.047816 | 0.001694 | Q93084 | ATP2A3 | 1903 |
| 26 | 11 | 23.28365 | 0.03887 | 0.001669 | Q8NBS9 | TXNDC5 | 1904 |
| 44 | 12 | 28.3169 | 0.047239 | 0.001668 | P27105 | STOM | 1905 |
| 40 | 31 | 27.40188 | 0.045078 | 0.001645 | O15117 | FYB1 | 1906 |
| 14 | 8 | 25.40533 | 0.041378 | 0.001629 | P30153 | PPP2R1A | 1907 |
| 31 | 14 | 27.10934 | 0.043689 | 0.001612 | Q9UNF0 | PACSIN2 | 1908 |
| 37 | 16 | 25.20183 | 0.039954 | 0.001585 | P28838 | LAP3 | 1909 |
| 36 | 16 | 24.26652 | 0.036621 | 0.001509 | Q16555 | DPYSL2 | 1910 |
| 23 | 7 | 24.10011 | 0.036019 | 0.001495 | P13716 | ALAD | 1911 |
| 41 | 6 | 31.09876 | 0.046141 | 0.001484 | P02654 | APOC1 | 1912 |
| 42 | 24 | 27.95559 | 0.04103 | 0.001468 | P30101 | PDIA3 | 1913 |
| 34 | 23 | 29.15039 | 0.042685 | 0.001464 | P27824 | CANX | 1914 |
| 50 | 12 | 26.60223 | 0.038766 | 0.001457 | P30041 | PRDX6 | 1915 |
| 28 | 10 | 23.24255 | 0.032808 | 0.001412 | P49593 | PPM1F | 1916 |
| 36 | 42 | 28.34972 | 0.039259 | 0.001385 | Q13201 | MMRN1 | 1917 |
| 66 | 8 | 27.02669 | 0.036965 | 0.001368 | Q9NRW1 | RAB6B | 1918 |
| 34 | 42 | 26.58033 | 0.035975 | 0.001353 | O75116 | ROCK2 | 1919 |
| 6 | 5 | 24.3762 | 0.032383 | 0.001328 | Q5T9A4 | ATAD3B | 1920 |
| 63 | 117 | 32.19624 | 0.042659 | 0.001325 | P35579 | MYH9 | 1921 |
| 62 | 148 | 32.64041 | 0.042808 | 0.001312 | P21333 | FLNA | 1922 |
| 23 | 37 | 26.83933 | 0.030623 | 0.001141 | Q00610 | CLTC | 1923 |
| 46 | 10 | 25.11677 | 0.027403 | 0.001091 | Q99497 | PARK7 | 1924 |

| | | | | | | | |
|---|---|---|---|---|---|---|---|
| 24 | 23 | 27.66098 | 0.03008 | 0.001087 | P19827 | ITIH1 | 1925 |
| 53 | 28 | 29.564 | 0.032143 | 0.001087 | P04004 | VTN | 1926 |
| 32 | 24 | 26.90336 | 0.027705 | 0.00103 | P07225 | PROS1 | 1927 |
| 62 | 35 | 31.15241 | 0.025753 | 0.000827 | P12814 | ACTN1 | 1928 |

| Description |
|---|
| NEDD8 [OS=Homo sapiens] |
| Golgi reassembly-stacking protein 2 [OS=Homo sapiens] |
| N-alpha-acetyltransferase 16, NatA auxiliary subunit [OS=Homo sapiens] |
| Isoform 5 of Osteopontin [OS=Homo sapiens] |
| Synaptic vesicle membrane protein VAT-1 homolog [OS=Homo sapiens] |
| proteolipid protein 2 [OS=Homo sapiens] |
| Isoform 3 of E3 ubiquitin-protein ligase HUWE1 [OS=Homo sapiens] |
| Tripeptidyl-peptidase 1 [OS=Homo sapiens] |
| Zinc finger CCHC domain-containing protein 7 [OS=Homo sapiens] |
| Signal transducer and activator of transcription 6 [OS=Homo sapiens] |
| Lactotransferrin [OS=Homo sapiens] |
| abl interactor 1 [OS=Homo sapiens] |
| Serine/threonine-protein phosphatase 2A 56 kDa regulatory subunit alpha isoform [OS=Homo sapiens] |
| Valine--tRNA ligase [OS=Homo sapiens] |
| CLIP-associating protein 2 [OS=Homo sapiens] |
| Solute carrier family 43 member 3 [OS=Homo sapiens] |
| Pro-epidermal growth factor [OS=Homo sapiens] |
| kelch repeat and BTB domain-containing protein 6 [OS=Homo sapiens] |
| Isoform 2 of TNF receptor-associated factor 2 [OS=Homo sapiens] |
| dolichyl-phosphate beta-glucosyltransferase [OS=Homo sapiens] |
| Zinc finger protein 518A [OS=Homo sapiens] |
| Phosphatidylinositol 4,5-bisphosphate 3-kinase catalytic subunit gamma isoform [OS=Homo sapiens] |
| Isoform M1 of Pyruvate kinase PKM [OS=Homo sapiens] |
| cullin-4A [OS=Homo sapiens] |
| Isoform 4 of Pyruvate dehydrogenase E1 component subunit alpha, somatic form, mitochondrial [OS=Homo |
| Grancalcin [OS=Homo sapiens] |
| Complement component C8 beta chain [OS=Homo sapiens] |
| AP-3 complex subunit sigma-1 [OS=Homo sapiens] |
| C-C motif chemokine 5 [OS=Homo sapiens] |
| Isoform 3 of Inositol 1,4,5-trisphosphate receptor type 1 [OS=Homo sapiens] |
| Granulins [OS=Homo sapiens] |
| Isoform Long of Ubiquitin recognition factor in ER-associated degradation protein 1 [OS=Homo sapiens] |
| Insulin-like growth factor-binding protein 7 [OS=Homo sapiens] |
| Beta-1,4-galactosyltransferase 1 [OS=Homo sapiens] |
| Vacuolar protein sorting-associated protein VTA1 homolog [OS=Homo sapiens] |
| Neurabin-2 [OS=Homo sapiens] |
| Immunoglobulin lambda variable 7-46 [OS=Homo sapiens] |
| Phosphoribosylformylglycinamidine synthase [OS=Homo sapiens] |
| Tetraspanin-32 [OS=Homo sapiens] |
| glutamine--tRNA ligase [OS=Homo sapiens] |
| Microtubule-associated protein 9 [OS=Homo sapiens] |
| Phospholipase A-2-activating protein [OS=Homo sapiens] |
| Ubiquitin carboxyl-terminal hydrolase MINDY-2 [OS=Homo sapiens] |
| Apolipoprotein B receptor [OS=Homo sapiens] |

| |
|---|
| Serine/threonine-protein kinase ICK [OS=Homo sapiens] |
| aquaporin-1 [OS=Homo sapiens] |
| Non-secretory ribonuclease [OS=Homo sapiens] |
| 1-phosphatidylinositol 4,5-bisphosphate phosphodiesterase beta-2 [OS=Homo sapiens] |
| Vacuolar protein sorting-associated protein 37A [OS=Homo sapiens] |
| Kinase D-interacting substrate of 220 kDa [OS=Homo sapiens] |
| Choline/ethanolamine kinase [OS=Homo sapiens] |
| Anosmin-1 [OS=Homo sapiens] |
| Trafficking kinesin-binding protein 1 [OS=Homo sapiens] |
| Kinesin-like protein KIFC3 [OS=Homo sapiens] |
| Inositol 1,4,5-trisphosphate receptor type 2 [OS=Homo sapiens] |
| ap-3 complex subunit mu-1 [OS=Homo sapiens] |
| acyl-coenzyme A thioesterase 1 [OS=Homo sapiens] |
| Tissue factor pathway inhibitor 2 [OS=Homo sapiens] |
| Neprilysin [OS=Homo sapiens] |
| Phosphatidylinositol 3,4,5-trisphosphate 5-phosphatase 1 [OS=Homo sapiens] |
| Coagulation factor X [OS=Homo sapiens] |
| Sacsin [OS=Homo sapiens] |
| Vesicle-associated membrane protein 3 [OS=Homo sapiens] |
| Golgi integral membrane protein 4 [OS=Homo sapiens] |
| Biogenesis of lysosome-related organelles complex 1 subunit 5 [OS=Homo sapiens] |
| Thioredoxin reductase 1, cytoplasmic [OS=Homo sapiens] |
| Dual specificity mitogen-activated protein kinase kinase 4 [OS=Homo sapiens] |
| Metalloreductase STEAP3 [OS=Homo sapiens] |
| Mitochondrial import inner membrane translocase subunit Tim10 B [OS=Homo sapiens] |
| AP-3 complex subunit beta-1 [OS=Homo sapiens] |
| Dolichyl-diphosphooligosaccharide--protein glycosyltransferase subunit STT3B [OS=Homo sapiens] |
| Tubulin-specific chaperone D [OS=Homo sapiens] |
| Eukaryotic translation initiation factor 4H [OS=Homo sapiens] |
| Spermatogenesis-defective protein 39 homolog [OS=Homo sapiens] |
| Peroxisomal membrane protein 11B [OS=Homo sapiens] |
| Putative peptidyl-tRNA hydrolase PTRHD1 [OS=Homo sapiens] |
| centrosomal protein of 131 kDa [OS=Homo sapiens] |
| Adenine phosphoribosyltransferase [OS=Homo sapiens] |
| Elongation factor 1-beta [OS=Homo sapiens] |
| signal-induced proliferation-associated protein 1 [OS=Homo sapiens] |
| NAD-dependent protein deacetylase sirtuin-2 [OS=Homo sapiens] |
| Gamma-aminobutyric acid receptor-associated protein-like 2 [OS=Homo sapiens] |
| 26S proteasome non-ATPase regulatory subunit 7 [OS=Homo sapiens] |
| 28S ribosomal protein S36, mitochondrial [OS=Homo sapiens] |
| Protein FAM177A1 [OS=Homo sapiens] |
| Opioid growth factor receptor-like protein 1 [OS=Homo sapiens] |
| Cathepsin G [OS=Homo sapiens] |
| Persulfide dioxygenase ETHE1, mitochondrial [OS=Homo sapiens] |
| Unconventional myosin-Ic [OS=Homo sapiens] |
| Keratin, type II cytoskeletal 78 [OS=Homo sapiens] |
| Eukaryotic translation initiation factor 4 gamma 2 [OS=Homo sapiens] |

| |
|---|
| Hypoxanthine-guanine phosphoribosyltransferase [OS=Homo sapiens] |
| deoxyribose-phosphate aldolase [OS=Homo sapiens] |
| Coagulation factor IX [OS=Homo sapiens] |
| Coagulation factor XI [OS=Homo sapiens] |
| Mitogen-activated protein kinase kinase kinase 5 [OS=Homo sapiens] |
| 5-formyltetrahydrofolate cyclo-ligase [OS=Homo sapiens] |
| major vault protein [OS=Homo sapiens] |
| multidrug resistance protein 1 [OS=Homo sapiens] |
| Isoform 7 of Nesprin-1 [OS=Homo sapiens] |
| Synapse-associated protein 1 [OS=Homo sapiens] |
| Protein FAM234A [OS=Homo sapiens] |
| Isoform 6 of Actin-binding LIM protein 1 [OS=Homo sapiens] |
| Phosphofurin acidic cluster sorting protein 1 [OS=Homo sapiens] |
| Dipeptidyl peptidase 1 [OS=Homo sapiens] |
| Ubiquitin-conjugating enzyme E2 K [OS=Homo sapiens] |
| Transmembrane protein 263 [OS=Homo sapiens] |
| COP9 signalosome complex subunit 2 [OS=Homo sapiens] |
| Echinoderm microtubule-associated protein-like 3 [OS=Homo sapiens] |
| Secretory carrier-associated membrane protein 1 [OS=Homo sapiens] |
| Long-chain fatty acid transport protein 4 [OS=Homo sapiens] |
| Lysosomal-trafficking regulator [OS=Homo sapiens] |
| activator of 90 kDa heat shock protein ATPase homolog 1 [OS=Homo sapiens] |
| Asparagine--tRNA ligase, cytoplasmic [OS=Homo sapiens] |
| immunoglobulin heavy variable 4-28 [OS=Homo sapiens] |
| Flotillin-2 [OS=Homo sapiens] |
| SLC35A4 upstream open reading frame protein [OS=Homo sapiens] |
| Multimerin-2 [OS=Homo sapiens] |
| Isoform 4 of Intersectin-2 [OS=Homo sapiens] |
| Tyrosine-protein phosphatase non-receptor type 18 [OS=Homo sapiens] |
| Sushi, von Willebrand factor type A, EGF and pentraxin domain-containing protein 1 [OS=Homo sapiens] |
| Protein-glutamine gamma-glutamyltransferase K [OS=Homo sapiens] |
| Major facilitator superfamily domain-containing protein 6 [OS=Homo sapiens] |
| Cytochrome c oxidase subunit 7A-related protein, mitochondrial [OS=Homo sapiens] |
| Isoform 3 of Glutaminase kidney isoform, mitochondrial [OS=Homo sapiens] |
| ubiquitin-conjugating enzyme e2 j1 [OS=Homo sapiens] |
| Cysteine-rich secretory protein LCCL domain-containing 2 [OS=Homo sapiens] |
| Collagen alpha-1(VI) chain [OS=Homo sapiens] |
| NADH dehydrogenase [ubiquinone] 1 alpha subcomplex subunit 8 [OS=Homo sapiens] |
| galactokinase [OS=Homo sapiens] |
| Beta-actin-like protein 2 [OS=Homo sapiens] |
| Cytochrome b-c1 complex subunit 7 [OS=Homo sapiens] |
| Synaptosomal-associated protein 29 [OS=Homo sapiens] |
| immunoglobulin kappa variable 1-8 [OS=Homo sapiens] |
| Neurobeachin-like protein 2 [OS=Homo sapiens] |
| Protein canopy homolog 2 [OS=Homo sapiens] |
| V-type proton ATPase subunit E 1 [OS=Homo sapiens] |
| Retinal dehydrogenase 1 [OS=Homo sapiens] |

| |
|---|
| Serglycin [OS=Homo sapiens] |
| Tapasin [OS=Homo sapiens] |
| 5'-AMP-activated protein kinase catalytic subunit alpha-1 [OS=Homo sapiens] |
| vitamin K epoxide reductase complex subunit 1 [OS=Homo sapiens] |
| Vesicular integral-membrane protein VIP36 [OS=Homo sapiens] |
| prenylcysteine oxidase 1 [OS=Homo sapiens] |
| Stromal cell-derived factor 2-like protein 1 [OS=Homo sapiens] |
| Isoform 2 of BSD domain-containing protein 1 [OS=Homo sapiens] |
| ceroid-lipofuscinosis neuronal protein 6 [OS=Homo sapiens] |
| [Protein ADP-ribosylarginine] hydrolase [OS=Homo sapiens] |
| Methionine aminopeptidase 1 [OS=Homo sapiens] |
| arrestin domain-containing protein 1 [OS=Homo sapiens] |
| Zinc finger protein 622 [OS=Homo sapiens] |
| Vesicle transport protein USE1 [OS=Homo sapiens] |
| arginine--tRNA ligase, cytoplasmic [OS=Homo sapiens] |
| kappa-casein [OS=Homo sapiens] |
| Vesicle-fusing ATPase [OS=Homo sapiens] |
| Lymphocyte antigen 75 [OS=Homo sapiens] |
| V-type proton ATPase subunit B, brain isoform [OS=Homo sapiens] |
| Tensin-1 [OS=Homo sapiens] |
| immunoglobulin heavy variable 3-72 [OS=Homo sapiens] |
| Isoform 3 of Ubiquitin conjugation factor E4 B [OS=Homo sapiens] |
| SEC23-interacting protein [OS=Homo sapiens] |
| CD226 antigen [OS=Homo sapiens] |
| Transmembrane emp24 domain-containing protein 1 [OS=Homo sapiens] |
| Cytochrome c oxidase assembly factor 3 homolog, mitochondrial [OS=Homo sapiens] |
| Calpastatin [OS=Homo sapiens] |
| Protein BRICK1 [OS=Homo sapiens] |
| Vacuolar ATPase assembly integral membrane protein vma21 [OS=Homo sapiens] |
| NADH dehydrogenase [ubiquinone] 1 alpha subcomplex subunit 7 [OS=Homo sapiens] |
| Isoform 1 of Leucine repeat adapter protein 25 [OS=Homo sapiens] |
| [Pyruvate dehydrogenase (Acetyl-transferring)] kinase isozyme 1, mitochondrial [OS=Homo sapiens] |
| Mitofusin-2 [OS=Homo sapiens] |
| eukaryotic translation initiation factor 4B [OS=Homo sapiens] |
| Nuclear protein localization protein 4 homolog [OS=Homo sapiens] |
| Deaminated glutathione amidase [OS=Homo sapiens] |
| ubiquitin carboxyl-terminal hydrolase 15 [OS=Homo sapiens] |
| Nuclear receptor-binding protein [OS=Homo sapiens] |
| Protein NipSnap homolog 2 [OS=Homo sapiens] |
| Protein RER1 [OS=Homo sapiens] |
| Amyloid-beta A4 protein [OS=Homo sapiens] |
| Acid ceramidase [OS=Homo sapiens] |
| PX domain-containing protein kinase-like protein [OS=Homo sapiens] |
| Fructose-bisphosphate aldolase C [OS=Homo sapiens] |
| STE20/SPS1-related proline-alanine-rich protein kinase [OS=Homo sapiens] |
| Disks large homolog 1 [OS=Homo sapiens] |
| Pyruvate dehydrogenase E1 component subunit beta, mitochondrial [OS=Homo sapiens] |

| |
|---|
| Guanine nucleotide-binding protein G(I)/G(S)/G(O) subunit gamma-11 [OS=Homo sapiens] |
| Ribosome-binding protein 1 [OS=Homo sapiens] |
| Dihydrolipoyl dehydrogenase, mitochondrial [OS=Homo sapiens] |
| Costars family protein ABRACL [OS=Homo sapiens] |
| protein FAM98A [OS=Homo sapiens] |
| AP-1 complex subunit sigma-2 [OS=Homo sapiens] |
| Docking protein 3 [OS=Homo sapiens] |
| Serine/threonine-protein kinase mTOR [OS=Homo sapiens] |
| Dihydropyrimidine dehydrogenase [NADP(+)] [OS=Homo sapiens] |
| Reticulon-2 [OS=Homo sapiens] |
| CD99 antigen [OS=Homo sapiens] |
| Serine/threonine-protein phosphatase 2B catalytic subunit gamma isoform [OS=Homo sapiens] |
| Gasdermin-D [OS=Homo sapiens] |
| Myeloid-derived growth factor [OS=Homo sapiens] |
| Serine-threonine kinase receptor-associated protein [OS=Homo sapiens] |
| Immunoglobulin kappa variable 2-30 [OS=Homo sapiens] |
| Reelin [OS=Homo sapiens] |
| WD repeat and FYVE domain-containing protein 1 [OS=Homo sapiens] |
| Tumor protein D54 [OS=Homo sapiens] |
| Pigment epithelium-derived factor [OS=Homo sapiens] |
| Immunoglobulin lambda variable 3-21 [OS=Homo sapiens] |
| Prenylcysteine oxidase-like [OS=Homo sapiens] |
| Ras-related protein Rab-8B [OS=Homo sapiens] |
| Isoform PACE4C of Proprotein convertase subtilisin/kexin type 6 [OS=Homo sapiens] |
| Erlin-2 [OS=Homo sapiens] |
| Isoform 2 of Golgi-associated PDZ and coiled-coil motif-containing protein [OS=Homo sapiens] |
| Phosphatidylinositol-binding clathrin assembly protein [OS=Homo sapiens] |
| prefoldin subunit 1 [OS=Homo sapiens] |
| ATP-binding cassette sub-family E member 1 [OS=Homo sapiens] |
| Isoform 3 of Huntingtin-interacting protein K [OS=Homo sapiens] |
| 26S proteasome regulatory subunit 4 [OS=Homo sapiens] |
| WD repeat-containing protein 13 [OS=Homo sapiens] |
| Membrane protein FAM174A [OS=Homo sapiens] |
| Protein HEXIM2 [OS=Homo sapiens] |
| Perilipin-3 [OS=Homo sapiens] |
| Galectin-1 [OS=Homo sapiens] |
| Proteasome subunit alpha type-2 [OS=Homo sapiens] |
| Estradiol 17-beta-dehydrogenase 11 [OS=Homo sapiens] |
| Isoform 2 of NF-kappa-B essential modulator [OS=Homo sapiens] |
| 40S ribosomal protein SA [OS=Homo sapiens] |
| Acetolactate synthase-like protein [OS=Homo sapiens] |
| Isoform 2 of NEDD8-activating enzyme E1 regulatory subunit [OS=Homo sapiens] |
| SH3 domain-containing kinase-binding protein 1 [OS=Homo sapiens] |
| Inhibitor of nuclear factor kappa-B kinase subunit beta [OS=Homo sapiens] |
| Metalloproteinase inhibitor 3 [OS=Homo sapiens] |
| CDGSH iron-sulfur domain-containing protein 2 [OS=Homo sapiens] |
| Keratin, type II cytoskeletal 6A [OS=Homo sapiens] |

| |
|---|
| Complement factor H-related protein 5 [OS=Homo sapiens] |
| Sulfotransferase 1A3 [OS=Homo sapiens] |
| Peptidyl-prolyl cis-trans isomerase F, mitochondrial [OS=Homo sapiens] |
| Transmembrane emp24 domain-containing protein 9 [OS=Homo sapiens] |
| Testis-expressed protein 2 [OS=Homo sapiens] |
| CCR4-NOT transcription complex subunit 10 [OS=Homo sapiens] |
| CD40 ligand [OS=Homo sapiens] |
| Filamin-A-interacting protein 1 [OS=Homo sapiens] |
| Plexin-A4 [OS=Homo sapiens] |
| Platelet endothelial aggregation receptor 1 [OS=Homo sapiens] |
| Transmembrane emp24 domain-containing protein 7 [OS=Homo sapiens] |
| translocon-associated protein subunit delta [OS=Homo sapiens] |
| Inosine triphosphate pyrophosphatase [OS=Homo sapiens] |
| Pleckstrin homology domain-containing family F member 2 [OS=Homo sapiens] |
| Isoform 3 of Apoptosis-inducing factor 1, mitochondrial [OS=Homo sapiens] |
| Hippocalcin-like protein 1 [OS=Homo sapiens] |
| GTP-binding protein rheb [OS=Homo sapiens] |
| zinc finger CCCH-type antiviral protein 1 [OS=Homo sapiens] |
| E3 UFM1-protein ligase 1 [OS=Homo sapiens] |
| Protein FAM136A [OS=Homo sapiens] |
| Heterogeneous nuclear ribonucleoprotein Q [OS=Homo sapiens] |
| Acetyl-CoA acetyltransferase, cytosolic [OS=Homo sapiens] |
| Latexin [OS=Homo sapiens] |
| NEDD8-conjugating enzyme UBE2F [OS=Homo sapiens] |
| lysozyme c [OS=Homo sapiens] |
| Chloride intracellular channel protein 4 [OS=Homo sapiens] |
| Isoamyl acetate-hydrolyzing esterase 1 homolog [OS=Homo sapiens] |
| Striatin-4 [OS=Homo sapiens] |
| thiosulfate sulfurtransferase [OS=Homo sapiens] |
| Actin, alpha cardiac muscle 1 [OS=Homo sapiens] |
| Guanine nucleotide-binding protein subunit alpha-13 [OS=Homo sapiens] |
| protein transport protein Sec61 subunit beta [OS=Homo sapiens] |
| NHL repeat-containing protein 2 [OS=Homo sapiens] |
| interferon-induced 35 kDa protein [OS=Homo sapiens] |
| Isoform 2 of Trem-like transcript 1 protein [OS=Homo sapiens] |
| Testis-specific serine/threonine-protein kinase 1 [OS=Homo sapiens] |
| Protein numb homolog [OS=Homo sapiens] |
| Platelet-derived growth factor subunit B [OS=Homo sapiens] |
| ATP synthase subunit e, mitochondrial [OS=Homo sapiens] |
| Exportin-1 [OS=Homo sapiens] |
| Eukaryotic translation initiation factor 5 [OS=Homo sapiens] |
| Prefoldin subunit 6 [OS=Homo sapiens] |
| Serine/threonine-protein phosphatase 6 regulatory subunit 3 [OS=Homo sapiens] |
| Keratin, type II cytoskeletal 6C [OS=Homo sapiens] |
| Sodium/hydrogen exchanger 9 [OS=Homo sapiens] |
| Isoform 3 of ATP synthase subunit f, mitochondrial [OS=Homo sapiens] |
| MARVEL domain-containing protein 1 [OS=Homo sapiens] |

| |
|---|
| Wiskott-Aldrich syndrome protein family member 2 [OS=Homo sapiens] |
| Secretory carrier-associated membrane protein 3 [OS=Homo sapiens] |
| 39S ribosomal protein L18, mitochondrial [OS=Homo sapiens] |
| Gamma-enolase [OS=Homo sapiens] |
| corticosteroid-binding globulin [OS=Homo sapiens] |
| Type II inositol 3,4-bisphosphate 4-phosphatase [OS=Homo sapiens] |
| Plastin-3 [OS=Homo sapiens] |
| Prefoldin subunit 2 [OS=Homo sapiens] |
| Claudin-3 [OS=Homo sapiens] |
| dnaJ homolog subfamily B member 1 [OS=Homo sapiens] |
| guanylate kinase [OS=Homo sapiens] |
| multidrug resistance-associated protein 4 [OS=Homo sapiens] |
| Isoform 3 of MOB-like protein phocein [OS=Homo sapiens] |
| Isoform 2 of CLIP-associating protein 1 [OS=Homo sapiens] |
| Desmocollin-1 [OS=Homo sapiens] |
| Isoform 4 of AP-3 complex subunit delta-1 [OS=Homo sapiens] |
| Uncharacterized protein C18orf63 [OS=Homo sapiens] |
| Protein Hook homolog 3 [OS=Homo sapiens] |
| Isoform 3 of Dedicator of cytokinesis protein 10 [OS=Homo sapiens] |
| Signal peptidase complex subunit 2 [OS=Homo sapiens] |
| Ubiquitin-associated protein 1 [OS=Homo sapiens] |
| 5'-AMP-activated protein kinase subunit gamma-1 [OS=Homo sapiens] |
| calcium/calmodulin-dependent protein kinase type 1 [OS=Homo sapiens] |
| Up-regulated during skeletal muscle growth protein 5 [OS=Homo sapiens] |
| COP9 signalosome complex subunit 6 [OS=Homo sapiens] |
| Sideroflexin-3 [OS=Homo sapiens] |
| Cytochrome c oxidase subunit 2 [OS=Homo sapiens] |
| Ester hydrolase C11orf54 [OS=Homo sapiens] |
| lactoylglutathione lyase [OS=Homo sapiens] |
| Isochorismatase domain-containing protein 1 [OS=Homo sapiens] |
| mitochondrial ornithine transporter 1 [OS=Homo sapiens] |
| terminal uridylyltransferase 7 [OS=Homo sapiens] |
| lanC-like protein 2 [OS=Homo sapiens] |
| apolipoprotein D [OS=Homo sapiens] |
| DnaJ homolog subfamily B member 6 [OS=Homo sapiens] |
| Cytochrome c1, heme protein, mitochondrial [OS=Homo sapiens] |
| Isoform D of Eukaryotic translation initiation factor 4 gamma 1 [OS=Homo sapiens] |
| Oxysterol-binding protein 1 [OS=Homo sapiens] |
| serine palmitoyltransferase 1 [OS=Homo sapiens] |
| NCK-interacting protein with SH3 domain [OS=Homo sapiens] |
| inosine-5'-monophosphate dehydrogenase 2 [OS=Homo sapiens] |
| DNA dC->dU-editing enzyme APOBEC-3C [OS=Homo sapiens] |
| vitamin D-binding protein [OS=Homo sapiens] |
| Eukaryotic translation initiation factor 3 subunit I [OS=Homo sapiens] |
| Cytochrome c oxidase subunit 7B, mitochondrial [OS=Homo sapiens] |
| Solute carrier family 2, facilitated glucose transporter member 1 [OS=Homo sapiens] |
| AFG3-like protein 2 [OS=Homo sapiens] |

| |
|---|
| Sterile alpha motif domain-containing protein 14 [OS=Homo sapiens] |
| casein kinase II subunit alpha' [OS=Homo sapiens] |
| Overexpressed in colon carcinoma 1 protein [OS=Homo sapiens] |
| Cadherin-6 [OS=Homo sapiens] |
| Vacuolar protein sorting-associated protein 4B [OS=Homo sapiens] |
| AP-2 complex subunit sigma [OS=Homo sapiens] |
| Carbonyl reductase [NADPH] 1 [OS=Homo sapiens] |
| Isoform 3 of Xaa-Pro aminopeptidase 1 [OS=Homo sapiens] |
| Isoform 5 of IST1 homolog [OS=Homo sapiens] |
| ephrin-B1 [OS=Homo sapiens] |
| Isoform 2 of SH3 and PX domain-containing protein 2A [OS=Homo sapiens] |
| Acyl-CoA:lysophosphatidylglycerol acyltransferase 1 [OS=Homo sapiens] |
| E3 ubiquitin-protein ligase MYCBP2 [OS=Homo sapiens] |
| Triokinase/FMN cyclase [OS=Homo sapiens] |
| Protein bicaudal D homolog 2 [OS=Homo sapiens] |
| Coagulation factor XIII B chain [OS=Homo sapiens] |
| 26S proteasome regulatory subunit 7 [OS=Homo sapiens] |
| Ubiquitin carboxyl-terminal hydrolase isozyme L3 [OS=Homo sapiens] |
| Monocyte differentiation antigen CD14 [OS=Homo sapiens] |
| Isoform 2 of Cytosolic acyl coenzyme A thioester hydrolase [OS=Homo sapiens] |
| Protein ERGIC-53 [OS=Homo sapiens] |
| E3 ubiquitin/ISG15 ligase TRIM25 [OS=Homo sapiens] |
| Uroporphyrinogen decarboxylase [OS=Homo sapiens] |
| Isoform 2 of Disks large-associated protein 4 [OS=Homo sapiens] |
| Son of sevenless homolog 1 [OS=Homo sapiens] |
| Stomatin-like protein 2, mitochondrial [OS=Homo sapiens] |
| Ankycorbin [OS=Homo sapiens] |
| Mannose-1-phosphate guanyltransferase beta [OS=Homo sapiens] |
| Aldehyde dehydrogenase family 16 member A1 [OS=Homo sapiens] |
| PITH domain-containing protein 1 [OS=Homo sapiens] |
| Cardiomyopathy-associated protein 5 [OS=Homo sapiens] |
| Ras-related protein Rab-2A [OS=Homo sapiens] |
| EF-hand domain-containing protein D2 [OS=Homo sapiens] |
| Golgin subfamily A member 3 [OS=Homo sapiens] |
| PDZ domain-containing protein 8 [OS=Homo sapiens] |
| Isoform 1 of Kallikrein-11 [OS=Homo sapiens] |
| Neuropathy target esterase [OS=Homo sapiens] |
| cilia- and flagella-associated protein 53 [OS=Homo sapiens] |
| Dolichyl-diphosphooligosaccharide--protein glycosyltransferase subunit STT3A [OS=Homo sapiens] |
| Adenosylhomocysteinase [OS=Homo sapiens] |
| Dynamin-1 [OS=Homo sapiens] |
| Nephrocystin-1 [OS=Homo sapiens] |
| ER membrane protein complex subunit 8 [OS=Homo sapiens] |
| Prohibitin-2 [OS=Homo sapiens] |
| Sulfotransferase 1A1 [OS=Homo sapiens] |
| 26S proteasome non-ATPase regulatory subunit 10 [OS=Homo sapiens] |
| pleckstrin homology domain-containing family O member 1 [OS=Homo sapiens] |

| |
|---|
| glutaredoxin-3 [OS=Homo sapiens] |
| Isoform 2 of Ankyrin repeat domain-containing protein 36B [OS=Homo sapiens] |
| glycerol kinase [OS=Homo sapiens] |
| NADH dehydrogenase [ubiquinone] 1 beta subcomplex subunit 5, mitochondrial [OS=Homo sapiens] |
| Syntaxin-4 [OS=Homo sapiens] |
| NLR family member X1 [OS=Homo sapiens] |
| Complement factor H-related protein 2 [OS=Homo sapiens] |
| 4F2 cell-surface antigen heavy chain [OS=Homo sapiens] |
| Androgen-induced gene 1 protein [OS=Homo sapiens] |
| Melanoma-associated antigen D2 [OS=Homo sapiens] |
| CCA tRNA nucleotidyltransferase 1, mitochondrial [OS=Homo sapiens] |
| Proteasome subunit alpha type-7 [OS=Homo sapiens] |
| epidermal growth factor receptor substrate 15 [OS=Homo sapiens] |
| Homogentisate 1,2-dioxygenase [OS=Homo sapiens] |
| Isoform 2 of CAP-Gly domain-containing linker protein 2 [OS=Homo sapiens] |
| Monocarboxylate transporter 4 [OS=Homo sapiens] |
| Rab GTPase-activating protein 1 [OS=Homo sapiens] |
| Phosphatidylinositol-glycan biosynthesis class W protein [OS=Homo sapiens] |
| Non-POU domain-containing octamer-binding protein [OS=Homo sapiens] |
| Torsin-1A-interacting protein 2 [OS=Homo sapiens] |
| Serpin B3 [OS=Homo sapiens] |
| syntaxin-binding protein 3 [OS=Homo sapiens] |
| Tropomodulin-1 [OS=Homo sapiens] |
| Succinyl-CoA:3-ketoacid coenzyme A transferase 1, mitochondrial [OS=Homo sapiens] |
| tubulin gamma-1 chain [OS=Homo sapiens] |
| transportin-1 [OS=Homo sapiens] |
| S-methylmethionine--homocysteine S-methyltransferase BHMT2 [OS=Homo sapiens] |
| Programmed cell death 6-interacting protein [OS=Homo sapiens] |
| Acylglycerol kinase, mitochondrial [OS=Homo sapiens] |
| Wiskott-Aldrich syndrome protein family member 3 [OS=Homo sapiens] |
| peptidyl-prolyl cis-trans isomerase D [OS=Homo sapiens] |
| Isoform 2 of Threonine--tRNA ligase, cytoplasmic [OS=Homo sapiens] |
| Arf-GAP with Rho-GAP domain, ANK repeat and PH domain-containing protein 1 [OS=Homo sapiens] |
| Serine/threonine-protein kinase PAK 2 [OS=Homo sapiens] |
| ATP-citrate synthase [OS=Homo sapiens] |
| EF-hand calcium-binding domain-containing protein 6 [OS=Homo sapiens] |
| Myosin phosphatase Rho-interacting protein [OS=Homo sapiens] |
| Isocitrate dehydrogenase [NAD] subunit alpha, mitochondrial [OS=Homo sapiens] |
| NADP-dependent malic enzyme [OS=Homo sapiens] |
| Cadherin-5 [OS=Homo sapiens] |
| phosphopantothenate--cysteine ligase [OS=Homo sapiens] |
| Isoform 2 of Rab-like protein 6 [OS=Homo sapiens] |
| Major facilitator superfamily domain-containing protein 2B [OS=Homo sapiens] |
| Transmembrane protein 206 [OS=Homo sapiens] |
| Serine protease 56 [OS=Homo sapiens] |
| Ragulator complex protein LAMTOR2 [OS=Homo sapiens] |
| Protein phosphatase 1 regulatory subunit 7 [OS=Homo sapiens] |

| |
|---|
| CAD protein [OS=Homo sapiens] |
| Ran-specific GTPase-activating protein [OS=Homo sapiens] |
| Isoform 3 of Echinoderm microtubule-associated protein-like 2 [OS=Homo sapiens] |
| Inactive rhomboid protein 2 [OS=Homo sapiens] |
| Protein Abhd11 [OS=Homo sapiens] |
| Protein Hikeshi [OS=Homo sapiens] |
| Protein S100-A4 [OS=Homo sapiens] |
| Prostaglandin E synthase 3 [OS=Homo sapiens] |
| 3-hydroxyisobutyryl-CoA hydrolase, mitochondrial [OS=Homo sapiens] |
| tyrosine-protein phosphatase non-receptor type 11 [OS=Homo sapiens] |
| Epidermal growth factor receptor substrate 15-like 1 [OS=Homo sapiens] |
| Isoform 2 of Cdc42-interacting protein 4 [OS=Homo sapiens] |
| Kallistatin [OS=Homo sapiens] |
| Thymosin beta-4 [OS=Homo sapiens] |
| Tetranectin [OS=Homo sapiens] |
| Caspase-14 [OS=Homo sapiens] |
| Desmoglein-1 [OS=Homo sapiens] |
| LIM domain-containing protein 1 [OS=Homo sapiens] |
| acyl-CoA-binding protein [OS=Homo sapiens] |
| ras-related protein Rap-1A [OS=Homo sapiens] |
| Isoform 2 of 26S proteasome regulatory subunit 8 [OS=Homo sapiens] |
| 26S proteasome non-ATPase regulatory subunit 4 [OS=Homo sapiens] |
| Isoform 2 of Farnesyl pyrophosphate synthase [OS=Homo sapiens] |
| mob kinase activator 1b [OS=Homo sapiens] |
| V-type proton ATPase subunit H [OS=Homo sapiens] |
| Vacuolar protein sorting-associated protein 35 [OS=Homo sapiens] |
| Prosaposin [OS=Homo sapiens] |
| bola-like protein 2 [OS=Homo sapiens] |
| Supervillin [OS=Homo sapiens] |
| Complement C4-A [OS=Homo sapiens] |
| Protein Prrc2c [OS=Homo sapiens] |
| Terminal uridylyltransferase 4 [OS=Homo sapiens] |
| NADH dehydrogenase [ubiquinone] 1 subunit C2 [OS=Homo sapiens] |
| gamma-glutamylcyclotransferase [OS=Homo sapiens] |
| LRP chaperone MESD [OS=Homo sapiens] |
| transcription factor BTF3 homolog 4 [OS=Homo sapiens] |
| 2-oxoglutarate dehydrogenase, mitochondrial [OS=Homo sapiens] |
| Platelet factor 4 variant [OS=Homo sapiens] |
| Protein FAM114A2 [OS=Homo sapiens] |
| thyroxine-binding globulin [OS=Homo sapiens] |
| Isoform 5 of Protein phosphatase 1 regulatory subunit 12A [OS=Homo sapiens] |
| Isovaleryl-CoA dehydrogenase, mitochondrial [OS=Homo sapiens] |
| Spermine synthase [OS=Homo sapiens] |
| Phenylalanine--tRNA ligase alpha subunit [OS=Homo sapiens] |
| Carbonic anhydrase 13 [OS=Homo sapiens] |
| Junction plakoglobin [OS=Homo sapiens] |
| Calcium uniporter protein, mitochondrial [OS=Homo sapiens] |

| |
|---|
| Fumarylacetoacetase [OS=Homo sapiens] |
| Isoform 2 of Endophilin-B1 [OS=Homo sapiens] |
| Polyadenylate-binding protein 1 [OS=Homo sapiens] |
| Misshapen-like kinase 1 [OS=Homo sapiens] |
| Vitelline membrane outer layer protein 1 homolog [OS=Homo sapiens] |
| N-acetylgalactosamine kinase [OS=Homo sapiens] |
| COP9 signalosome complex subunit 5 [OS=Homo sapiens] |
| Ubiquitin thioesterase otub1 [OS=Homo sapiens] |
| Trypsin-1 [OS=Homo sapiens] |
| dual specificity mitogen-activated protein kinase kinase 3 [OS=Homo sapiens] |
| V-type proton ATPase subunit C 1 [OS=Homo sapiens] |
| Keratin, type I cytoskeletal 17 [OS=Homo sapiens] |
| Isoform 2 of Basic leucine zipper and W2 domain-containing protein 1 [OS=Homo sapiens] |
| Keratin, type II cytoskeletal 3 [OS=Homo sapiens] |
| Protein-tyrosine sulfotransferase 2 [OS=Homo sapiens] |
| Myotubularin-related protein 12 [OS=Homo sapiens] |
| BET1-like protein [OS=Homo sapiens] |
| Sharpin [OS=Homo sapiens] |
| Arginine-hydroxylase NDUFAF5, mitochondrial [OS=Homo sapiens] |
| Isoform 4 of Ubiquitin carboxyl-terminal hydrolase 47 [OS=Homo sapiens] |
| C-C motif chemokine 18 [OS=Homo sapiens] |
| Isoform 6 of Tight junction protein ZO-2 [OS=Homo sapiens] |
| Vitamin K-dependent protein C [OS=Homo sapiens] |
| Sodium/potassium-transporting ATPase subunit alpha-1 [OS=Homo sapiens] |
| S-methyl-5'-thioadenosine phosphorylase [OS=Homo sapiens] |
| Isoform 2 of Kinectin [OS=Homo sapiens] |
| Caspase recruitment domain-containing protein 6 [OS=Homo sapiens] |
| Isoform 4 of NADH-ubiquinone oxidoreductase 75 kDa subunit, mitochondrial [OS=Homo sapiens] |
| 1-phosphatidylinositol 4,5-bisphosphate phosphodiesterase beta-3 [OS=Homo sapiens] |
| CD109 antigen [OS=Homo sapiens] |
| small ubiquitin-related modifier 2 [OS=Homo sapiens] |
| Rab GDP dissociation inhibitor alpha [OS=Homo sapiens] |
| Ribosomal protein S6 kinase beta-2 [OS=Homo sapiens] |
| ADP-ribosylation factor GTPase-activating protein 1 [OS=Homo sapiens] |
| ras-related protein rab-27b [OS=Homo sapiens] |
| Thioredoxin-related transmembrane protein 4 [OS=Homo sapiens] |
| charged multivesicular body protein 6 [OS=Homo sapiens] |
| Collectin-11 [OS=Homo sapiens] |
| Protein transport protein Sec16A [OS=Homo sapiens] |
| alpha-methylacyl-CoA racemase [OS=Homo sapiens] |
| Caspase-6 [OS=Homo sapiens] |
| conserved oligomeric Golgi complex subunit 5 [OS=Homo sapiens] |
| SLAIN motif-containing protein 2 [OS=Homo sapiens] |
| Major prion protein [OS=Homo sapiens] |
| prolyl endopeptidase [OS=Homo sapiens] |
| Isoform 3 of Regulator of G-protein signaling 6 [OS=Homo sapiens] |
| Early endosome antigen 1 [OS=Homo sapiens] |

| |
|---|
| Isoform 2 of DnaJ homolog subfamily A member 1 [OS=Homo sapiens] |
| Isoform 3 of Histone-lysine N-methyltransferase 2C [OS=Homo sapiens] |
| Adenylate cyclase type 3 [OS=Homo sapiens] |
| V-type proton ATPase subunit S1 [OS=Homo sapiens] |
| Transmembrane channel-like protein 1 [OS=Homo sapiens] |
| protein PRRC1 [OS=Homo sapiens] |
| FERM domain-containing protein 4B [OS=Homo sapiens] |
| Isoform 2 of PDZ and LIM domain protein 7 [OS=Homo sapiens] |
| Hemoglobin subunit gamma-1 [OS=Homo sapiens] |
| Protein FAM107B [OS=Homo sapiens] |
| translocator protein [OS=Homo sapiens] |
| Urokinase-type plasminogen activator [OS=Homo sapiens] |
| TIP41-like protein [OS=Homo sapiens] |
| Isocitrate dehydrogenase [NADP] cytoplasmic [OS=Homo sapiens] |
| Myristoylated alanine-rich C-kinase substrate [OS=Homo sapiens] |
| NADH dehydrogenase [ubiquinone] iron-sulfur protein 3, mitochondrial [OS=Homo sapiens] |
| Ribonuclease 7 [OS=Homo sapiens] |
| Transmembrane emp24 domain-containing protein 4 [OS=Homo sapiens] |
| histone acetyltransferase KAT6A [OS=Homo sapiens] |
| methyltransferase-like protein 7A [OS=Homo sapiens] |
| transport and Golgi organization protein 2 homolog [OS=Homo sapiens] |
| Hemoglobin subunit delta [OS=Homo sapiens] |
| D-aminoacyl-tRNA deacylase 1 [OS=Homo sapiens] |
| immunoglobulin lambda variable 8-61 [OS=Homo sapiens] |
| Cation-dependent mannose-6-phosphate receptor [OS=Homo sapiens] |
| Isoform p135 of Dynactin subunit 1 [OS=Homo sapiens] |
| Brain acid soluble protein 1 [OS=Homo sapiens] |
| LIM domain and actin-binding protein 1 [OS=Homo sapiens] |
| immunoglobulin heavy variable 1-46 [OS=Homo sapiens] |
| Proteasome subunit beta type-6 [OS=Homo sapiens] |
| CD151 antigen [OS=Homo sapiens] |
| xaa-Pro dipeptidase [OS=Homo sapiens] |
| Isoform 5 of Mitochondrial fission factor [OS=Homo sapiens] |
| Protein phosphatase 1 regulatory subunit 14A [OS=Homo sapiens] |
| Rho GTPase-activating protein 10 [OS=Homo sapiens] |
| Platelet glycoprotein VI [OS=Homo sapiens] |
| UMP-CMP kinase [OS=Homo sapiens] |
| protein S100-A6 [OS=Homo sapiens] |
| Glucose-6-phosphate 1-dehydrogenase [OS=Homo sapiens] |
| Cystatin-A [OS=Homo sapiens] |
| methionine aminopeptidase 2 [OS=Homo sapiens] |
| grainyhead-like protein 3 homolog [OS=Homo sapiens] |
| Sulfide:quinone oxidoreductase, mitochondrial [OS=Homo sapiens] |
| Thromboxane A2 receptor [OS=Homo sapiens] |
| Pantetheinase [OS=Homo sapiens] |
| Isoform 2 of Septin-5 [OS=Homo sapiens] |
| GRIP1-associated protein 1 [OS=Homo sapiens] |

| |
|---|
| SH3 domain-binding glutamic acid-rich-like protein [OS=Homo sapiens] |
| coatomer subunit epsilon [OS=Homo sapiens] |
| 26s proteasome non-atpase regulatory subunit 12 [OS=Homo sapiens] |
| Isoform 7 of Ubiquitin carboxyl-terminal hydrolase 19 [OS=Homo sapiens] |
| muscleblind-like protein 2 [OS=Homo sapiens] |
| protein NipSnap homolog 3A [OS=Homo sapiens] |
| Actin-related protein 2/3 complex subunit 4 [OS=Homo sapiens] |
| Phenylalanine--tRNA ligase beta subunit [OS=Homo sapiens] |
| Oncoprotein-induced transcript 3 protein [OS=Homo sapiens] |
| BRO1 domain-containing protein BROX [OS=Homo sapiens] |
| immunoglobulin heavy variable 4-34 [OS=Homo sapiens] |
| Isoform 2 of Arf-GAP domain and FG repeat-containing protein 1 [OS=Homo sapiens] |
| Nucleobindin-2 [OS=Homo sapiens] |
| chloride channel CLIC-like protein 1 [OS=Homo sapiens] |
| Guanine nucleotide-binding protein G(I)/G(S)/G(T) subunit beta-2 [OS=Homo sapiens] |
| CUGBP Elav-like family member 2 [OS=Homo sapiens] |
| ras-related protein Rab-3C [OS=Homo sapiens] |
| Macrophage Migration inhibitory factor [OS=Homo sapiens] |
| ATP synthase subunit delta, mitochondrial [OS=Homo sapiens] |
| Glutathione S-transferase Mu 2 [OS=Homo sapiens] |
| Cytochrome c oxidase subunit 6C [OS=Homo sapiens] |
| pleckstrin homology domain-containing family O member 2 [OS=Homo sapiens] |
| mitochondrial-processing peptidase subunit beta [OS=Homo sapiens] |
| GMP reductase 1 [OS=Homo sapiens] |
| Integrin alpha-2 [OS=Homo sapiens] |
| Isoform 2 of Transmembrane protein 40 [OS=Homo sapiens] |
| Translation initiation factor eIF-2B subunit alpha [OS=Homo sapiens] |
| Platelet-activating factor acetylhydrolase IB subunit alpha [OS=Homo sapiens] |
| protein transport protein Sec23A [OS=Homo sapiens] |
| Membrane-associated progesterone receptor component 2 [OS=Homo sapiens] |
| ER membrane protein complex subunit 2 [OS=Homo sapiens] |
| E3 ubiquitin-protein ligase CHIP [OS=Homo sapiens] |
| pituitary tumor-transforming gene 1 protein-interacting protein [OS=Homo sapiens] |
| Early activation antigen CD69 [OS=Homo sapiens] |
| Importin-7 [OS=Homo sapiens] |
| Retinol-binding protein 4 [OS=Homo sapiens] |
| Isoform 2 of Myosin-10 [OS=Homo sapiens] |
| coatomer subunit alpha [OS=Homo sapiens] |
| Oxysterol-binding protein-related protein 9 [OS=Homo sapiens] |
| Eukaryotic translation initiation factor 2 subunit 2 [OS=Homo sapiens] |
| sequestosome-1 [OS=Homo sapiens] |
| Immunoglobulin heavy variable 3-15 [OS=Homo sapiens] |
| Protein Z-dependent protease inhibitor [OS=Homo sapiens] |
| Isoform 2 of Syntaxin-7 [OS=Homo sapiens] |
| Proteasome subunit beta type-8 [OS=Homo sapiens] |
| ER membrane protein complex subunit 1 [OS=Homo sapiens] |
| Small proline-rich protein 2A [OS=Homo sapiens] |

| |
|---|
| Pyridoxal-dependent decarboxylase domain-containing protein 1 [OS=Homo sapiens] |
| Exopolyphosphatase PRUNE1 [OS=Homo sapiens] |
| Protein disulfide-isomerase TMX3 [OS=Homo sapiens] |
| argininosuccinate lyase [OS=Homo sapiens] |
| Tubulin alpha-8 chain [OS=Homo sapiens] |
| Isoform V of Septin-6 [OS=Homo sapiens] |
| Filaggrin-2 [OS=Homo sapiens] |
| Arginyl-tRNA--protein transferase 1 [OS=Homo sapiens] |
| Emerin [OS=Homo sapiens] |
| calcineurin B homologous protein 3 [OS=Homo sapiens] |
| Diphosphoinositol polyphosphate phosphohydrolase 1 [OS=Homo sapiens] |
| Coiled-coil domain-containing protein 47 [OS=Homo sapiens] |
| Proteasomal ubiquitin receptor ADRM1 [OS=Homo sapiens] |
| Cystatin-B [OS=Homo sapiens] |
| Keratin, type I cytoskeletal 10 [OS=Homo sapiens] |
| Fibulin-1 [OS=Homo sapiens] |
| Dematin [OS=Homo sapiens] |
| Metallo-beta-lactamase domain-containing protein 2 [OS=Homo sapiens] |
| thymidylate kinase [OS=Homo sapiens] |
| Dynactin subunit 4 [OS=Homo sapiens] |
| Cysteine-rich with EGF-like domain protein 1 [OS=Homo sapiens] |
| Calmodulin-like protein 5 [OS=Homo sapiens] |
| Eukaryotic translation initiation factor 2 subunit 1 [OS=Homo sapiens] |
| Transmembrane protein 165 [OS=Homo sapiens] |
| Immunoglobulin kappa variable 1-16 [OS=Homo sapiens] |
| NEDD8-conjugating enzyme Ubc12 [OS=Homo sapiens] |
| ranBP-type and C3HC4-type zinc finger-containing protein 1 [OS=Homo sapiens] |
| Plasminogen activator inhibitor 1 [OS=Homo sapiens] |
| NADH dehydrogenase [ubiquinone] 1 beta subcomplex subunit 10 [OS=Homo sapiens] |
| Glycophorin-C [OS=Homo sapiens] |
| Antithrombin-III [OS=Homo sapiens] |
| HLA class I histocompatibility antigen, alpha chain E [OS=Homo sapiens] |
| Acyl-coenzyme A thioesterase 9, mitochondrial [OS=Homo sapiens] |
| serine/threonine-protein phosphatase PP1-alpha catalytic subunit [OS=Homo sapiens] |
| Renin receptor [OS=Homo sapiens] |
| FYVE, RhoGEF and PH domain-containing protein 3 [OS=Homo sapiens] |
| Sodium- and chloride-dependent taurine transporter [OS=Homo sapiens] |
| Guanylate cyclase soluble subunit alpha-3 [OS=Homo sapiens] |
| Immunoglobulin lambda constant 6 [OS=Homo sapiens] |
| Sorting nexin-9 [OS=Homo sapiens] |
| Isoform 2 of NADH dehydrogenase [ubiquinone] flavoprotein 3, mitochondrial [OS=Homo sapiens] |
| Protein XRP2 [OS=Homo sapiens] |
| Ras-related protein Rab-2B [OS=Homo sapiens] |
| Uveal autoantigen with coiled-coil domains and ankyrin repeats [OS=Homo sapiens] |
| serine/threonine-protein kinase 4 [OS=Homo sapiens] |
| Dynein light chain 1, cytoplasmic [OS=Homo sapiens] |
| Putative ciliary rootlet coiled-coil protein-like 2 protein [OS=Homo sapiens] |

| |
|---|
| Calcyclin-binding protein [OS=Homo sapiens] |
| 26S proteasome non-ATPase regulatory subunit 1 [OS=Homo sapiens] |
| Keratin, type II cytoskeletal 2 epidermal [OS=Homo sapiens] |
| Fatty acid-binding protein, epidermal [OS=Homo sapiens] |
| Pregnancy zone protein [OS=Homo sapiens] |
| Isoform Beta-II of Protein kinase C beta type [OS=Homo sapiens] |
| Omega-amidase NIT2 [OS=Homo sapiens] |
| translocation protein sec62 [OS=Homo sapiens] |
| protein LSM14 homolog A [OS=Homo sapiens] |
| immunoglobulin heavy variable 5-51 [OS=Homo sapiens] |
| Nidogen-1 [OS=Homo sapiens] |
| nicotinate phosphoribosyltransferase [OS=Homo sapiens] |
| Ras GTPase-activating-like protein IQGAP2 [OS=Homo sapiens] |
| Beta-galactoside alpha-2,6-sialyltransferase 1 [OS=Homo sapiens] |
| Ras-related protein Rab-30 [OS=Homo sapiens] |
| cAMP-dependent protein kinase type II-alpha regulatory subunit [OS=Homo sapiens] |
| Syntaxin-12 [OS=Homo sapiens] |
| Isoform 2 of Vacuolar protein sorting-associated protein 45 [OS=Homo sapiens] |
| Centrosomal protein of 170 kDa [OS=Homo sapiens] |
| Translin [OS=Homo sapiens] |
| Isoform 3 of Ubiquitin-conjugating enzyme E2 D3 [OS=Homo sapiens] |
| Aspartate aminotransferase, cytoplasmic [OS=Homo sapiens] |
| Lymphocyte function-associated antigen 3 [OS=Homo sapiens] |
| Hepatic leukemia factor [OS=Homo sapiens] |
| Translationally-controlled tumor protein [OS=Homo sapiens] |
| Tumor necrosis factor receptor superfamily member 5 [OS=Homo sapiens] |
| Gamma-adducin [OS=Homo sapiens] |
| Phosphatidylinositol 5-phosphate 4-kinase type-2 beta [OS=Homo sapiens] |
| Secernin-1 [OS=Homo sapiens] |
| NADH dehydrogenase [ubiquinone] 1 beta subcomplex subunit 3 [OS=Homo sapiens] |
| Tricarboxylate transport protein, mitochondrial [OS=Homo sapiens] |
| Peroxisomal membrane protein PEX14 [OS=Homo sapiens] |
| Isoform 6 of Microtubule-associated protein 4 [OS=Homo sapiens] |
| Centromere-associated protein E [OS=Homo sapiens] |
| von Willebrand factor A domain-containing protein 5A [OS=Homo sapiens] |
| Trans-Golgi network integral membrane protein 2 [OS=Homo sapiens] |
| Catenin beta-1 [OS=Homo sapiens] |
| Partitioning defective 3 homolog [OS=Homo sapiens] |
| Histidine-rich glycoprotein [OS=Homo sapiens] |
| Isoform 2 of Eukaryotic translation initiation factor 3 subunit B [OS=Homo sapiens] |
| Wiskott-Aldrich syndrome protein [OS=Homo sapiens] |
| CKLF-like MARVEL transmembrane domain-containing protein 6 [OS=Homo sapiens] |
| Phosphatidylinositol transfer protein beta isoform [OS=Homo sapiens] |
| Serine-protein kinase ATM [OS=Homo sapiens] |
| Apolipoprotein C-IV [OS=Homo sapiens] |
| Isoform 3 of Regulator of G-protein signaling 10 [OS=Homo sapiens] |
| Coatomer subunit zeta-1 [OS=Homo sapiens] |

| |
|---|
| Isoform 4 of Dynamin-2 [OS=Homo sapiens] |
| Isoform 2 of UBX domain-containing protein 1 [OS=Homo sapiens] |
| tumor susceptibility gene 101 protein [OS=Homo sapiens] |
| Focal adhesion kinase 1 [OS=Homo sapiens] |
| L-xylulose reductase [OS=Homo sapiens] |
| Antigen peptide transporter 1 [OS=Homo sapiens] |
| Chromogranin-A [OS=Homo sapiens] |
| surfeit locus protein 4 [OS=Homo sapiens] |
| Tetraspanin-14 [OS=Homo sapiens] |
| Dynactin subunit 3 [OS=Homo sapiens] |
| Type 2 phosphatidylinositol 4,5-bisphosphate 4-phosphatase [OS=Homo sapiens] |
| Protein THEM6 [OS=Homo sapiens] |
| Zinc finger protein 268 [OS=Homo sapiens] |
| Gamma-glutamyl hydrolase [OS=Homo sapiens] |
| NADH dehydrogenase [ubiquinone] 1 beta subcomplex subunit 9 [OS=Homo sapiens] |
| RAC-beta serine/threonine-protein kinase [OS=Homo sapiens] |
| ADP-ribosylation factor 6 [OS=Homo sapiens] |
| cytoplasmic dynein 1 intermediate chain 2 [OS=Homo sapiens] |
| Keratin, type II cytoskeletal 1b [OS=Homo sapiens] |
| CD82 antigen [OS=Homo sapiens] |
| radixin [OS=Homo sapiens] |
| Plasma protease C1 inhibitor [OS=Homo sapiens] |
| Ras-related protein Rab-18 [OS=Homo sapiens] |
| vesicle-associated membrane protein 7 [OS=Homo sapiens] |
| cytochrome c [OS=Homo sapiens] |
| Lysophospholipase-like protein 1 [OS=Homo sapiens] |
| Cytoplasmic aconitate hydratase [OS=Homo sapiens] |
| STAM-binding protein [OS=Homo sapiens] |
| Isoform L of Membrane cofactor protein [OS=Homo sapiens] |
| Cell cycle control protein 50A [OS=Homo sapiens] |
| immunoglobulin lambda variable 4-69 [OS=Homo sapiens] |
| Protein phosphatase 1 regulatory subunit 3E [OS=Homo sapiens] |
| regulator of cell cycle RGCC [OS=Homo sapiens] |
| Isoform 3 of Rab11 family-interacting protein 3 [OS=Homo sapiens] |
| Hepatocyte growth factor-regulated tyrosine kinase substrate [OS=Homo sapiens] |
| Histidine triad nucleotide-binding protein 2, mitochondrial [OS=Homo sapiens] |
| Bifunctional epoxide hydrolase 2 [OS=Homo sapiens] |
| Thioredoxin domain-containing protein 17 [OS=Homo sapiens] |
| Leucine-rich repeat-containing protein 47 [OS=Homo sapiens] |
| annexin A11 [OS=Homo sapiens] |
| Isoform 3 of Choline transporter-like protein 1 [OS=Homo sapiens] |
| Immunoglobulin heavy variable 3-7 [OS=Homo sapiens] |
| TNF receptor-associated factor 6 [OS=Homo sapiens] |
| Alpha-1-acid glycoprotein 2 [OS=Homo sapiens] |
| Isoform 3 of Anoctamin-6 [OS=Homo sapiens] |
| Phosphoglycerate mutase 1 [OS=Homo sapiens] |
| proteasome subunit beta type-3 [OS=Homo sapiens] |

| |
|---|
| Antileukoproteinase [OS=Homo sapiens] |
| Complement component C7 [OS=Homo sapiens] |
| Isoform 2 of BCL2/adenovirus E1B 19 kDa protein-interacting protein 2 [OS=Homo sapiens] |
| cAMP-dependent protein kinase type II-beta regulatory subunit [OS=Homo sapiens] |
| Importin subunit alpha-7 [OS=Homo sapiens] |
| ras-related protein Rab-38 [OS=Homo sapiens] |
| Sec1 family domain-containing protein 1 [OS=Homo sapiens] |
| Isoform 1 of Laminin subunit alpha-3 [OS=Homo sapiens] |
| Isoform 2 of NADH dehydrogenase [ubiquinone] flavoprotein 1, mitochondrial [OS=Homo sapiens] |
| bifunctional purine biosynthesis protein purH [OS=Homo sapiens] |
| splicing factor 3B subunit 1 [OS=Homo sapiens] |
| E3 ubiquitin-protein ligase TRIM4 [OS=Homo sapiens] |
| nuclear transport factor 2 [OS=Homo sapiens] |
| Methylmalonate-semialdehyde dehydrogenase [acylating], mitochondrial [OS=Homo sapiens] |
| Keratin, type I cytoskeletal 9 [OS=Homo sapiens] |
| Tryptophan--tRNA ligase, cytoplasmic [OS=Homo sapiens] |
| 39S ribosomal protein L12, mitochondrial [OS=Homo sapiens] |
| Exocyst complex component 3-like protein 2 [OS=Homo sapiens] |
| Transforming acidic coiled-coil-containing protein 1 [OS=Homo sapiens] |
| Isoform 1 of Ubiquitin-conjugating enzyme E2 variant 1 [OS=Homo sapiens] |
| Linker for activation of T-cells family member 1 [OS=Homo sapiens] |
| T-complex protein 1 subunit eta [OS=Homo sapiens] |
| THAP domain-containing protein 2 [OS=Homo sapiens] |
| C-type lectin domain family 2 member L [OS=Homo sapiens] |
| Cilia- and flagella-associated protein 47 [OS=Homo sapiens] |
| endoplasmic reticulum aminopeptidase 1 [OS=Homo sapiens] |
| Hornerin [OS=Homo sapiens] |
| Lipoamide acyltransferase component of branched-chain alpha-keto acid dehydrogenase complex, mitochor |
| Isoform 2 of Glycogen phosphorylase, muscle form [OS=Homo sapiens] |
| SH3 domain-binding glutamic acid-rich-like protein 2 [OS=Homo sapiens] |
| proteasome activator complex subunit 2 [OS=Homo sapiens] |
| 40S ribosomal protein S12 [OS=Homo sapiens] |
| Keratin, type II cytoskeletal 4 [OS=Homo sapiens] |
| Isoform 2 of Protein FAM49B [OS=Homo sapiens] |
| NADH dehydrogenase [ubiquinone] flavoprotein 2, mitochondrial [OS=Homo sapiens] |
| Isoform 2 of Diacylglycerol kinase gamma [OS=Homo sapiens] |
| Suprabasin [OS=Homo sapiens] |
| transmembrane protein 102 [OS=Homo sapiens] |
| Protein TFG [OS=Homo sapiens] |
| Disabled homolog 2 [OS=Homo sapiens] |
| TBC1 domain family member 13 [OS=Homo sapiens] |
| annexin A5 [OS=Homo sapiens] |
| Quinone oxidoreductase [OS=Homo sapiens] |
| Mitogen-activated protein kinase 14 [OS=Homo sapiens] |
| Eukaryotic translation initiation factor 3 subunit A [OS=Homo sapiens] |
| Isoform 2 of STE20-like serine/threonine-protein kinase [OS=Homo sapiens] |
| Serine--tRNA ligase, cytoplasmic [OS=Homo sapiens] |

| |
|---|
| Platelet factor 4 [OS=Homo sapiens] |
| Isoform 2 of cAMP-dependent protein kinase catalytic subunit beta [OS=Homo sapiens] |
| Signal peptidase complex subunit 3 [OS=Homo sapiens] |
| atp-dependent rna helicase ddx3y [OS=Homo sapiens] |
| Isoform 2 of Tropomyosin beta chain [OS=Homo sapiens] |
| monoglyceride lipase [OS=Homo sapiens] |
| Serum amyloid A-4 protein [OS=Homo sapiens] |
| NADH dehydrogenase [ubiquinone] iron-sulfur protein 6, mitochondrial [OS=Homo sapiens] |
| Myotubularin [OS=Homo sapiens] |
| elongin-C [OS=Homo sapiens] |
| P2X purinoceptor 1 [OS=Homo sapiens] |
| N(G),N(G)-dimethylarginine dimethylaminohydrolase 2 [OS=Homo sapiens] |
| Protein tyrosine phosphatase type IVA 2 [OS=Homo sapiens] |
| Charged multivesicular body protein 4a [OS=Homo sapiens] |
| Transketolase [OS=Homo sapiens] |
| Cadherin-1 [OS=Homo sapiens] |
| Jupiter microtubule associated homolog 2 [OS=Homo sapiens] |
| MAM domain-containing glycosylphosphatidylinositol anchor protein 1 [OS=Homo sapiens] |
| serine/threonine-protein phosphatase 2B catalytic subunit beta isoform [OS=Homo sapiens] |
| Involucrin [OS=Homo sapiens] |
| Serine/threonine-protein kinase MRCK beta [OS=Homo sapiens] |
| Isoform 1 of Arf-GAP with SH3 domain, ANK repeat and PH domain-containing protein 1 [OS=Homo sapiens] |
| Voltage-dependent anion-selective channel protein 3 [OS=Homo sapiens] |
| NADH dehydrogenase [ubiquinone] 1 alpha subcomplex subunit 10, mitochondrial [OS=Homo sapiens] |
| Src-like-adapter 2 [OS=Homo sapiens] |
| Fragile X mental retardation syndrome-related protein 2 [OS=Homo sapiens] |
| Isoform 3 of Tyrosine-protein kinase Fyn [OS=Homo sapiens] |
| Hsp90 co-chaperone Cdc37 [OS=Homo sapiens] |
| Neutrophil defensin 3 [OS=Homo sapiens] |
| Exocyst complex component 5 [OS=Homo sapiens] |
| Calcium/calmodulin-dependent protein kinase type II subunit delta [OS=Homo sapiens] |
| Protein lifeguard 3 [OS=Homo sapiens] |
| Synaptotagmin-like protein 4 [OS=Homo sapiens] |
| ribose-phosphate pyrophosphokinase 2 [OS=Homo sapiens] |
| Integrin beta-5 [OS=Homo sapiens] |
| Sideroflexin-1 [OS=Homo sapiens] |
| Secretory carrier-associated membrane protein 2 [OS=Homo sapiens] |
| Oligoribonuclease, mitochondrial [OS=Homo sapiens] |
| Barrier-to-autointegration factor [OS=Homo sapiens] |
| Integrin beta-1 [OS=Homo sapiens] |
| Lymphoid-restricted membrane protein [OS=Homo sapiens] |
| serine/threonine-protein kinase 38 [OS=Homo sapiens] |
| Endoplasmic reticulum-Golgi intermediate compartment protein 1 [OS=Homo sapiens] |
| RIPOR family member 3 [OS=Homo sapiens] |
| sodium/potassium-transporting ATPase subunit beta-3 [OS=Homo sapiens] |
| ATP synthase subunit d, mitochondrial [OS=Homo sapiens] |
| Cytoskeleton-associated protein 5 [OS=Homo sapiens] |

| |
|---|
| EGF-containing fibulin-like extracellular matrix protein 1 [OS=Homo sapiens] |
| EH domain-containing protein 1 [OS=Homo sapiens] |
| Protein ABHD14B [OS=Homo sapiens] |
| disintegrin and metalloproteinase domain-containing protein 9 [OS=Homo sapiens] |
| Cytochrome c oxidase subunit 7A2, mitochondrial [OS=Homo sapiens] |
| Serum paraoxonase/lactonase 3 [OS=Homo sapiens] |
| Superoxide dismutase [Cu-Zn] [OS=Homo sapiens] |
| ubiquitin-like modifier-activating enzyme 7 [OS=Homo sapiens] |
| immunoglobulin kappa variable 4-1 [OS=Homo sapiens] |
| E3 ubiquitin-protein ligase HECTD3 [OS=Homo sapiens] |
| tRNA-splicing ligase RtcB homolog [OS=Homo sapiens] |
| Nck-associated protein 1-like [OS=Homo sapiens] |
| Isoform 3 of Poly(rC)-binding protein 2 [OS=Homo sapiens] |
| Calcium-binding mitochondrial carrier protein SCaMC-1 [OS=Homo sapiens] |
| Transforming acidic coiled-coil-containing protein 2 [OS=Homo sapiens] |
| ARF GTPase-activating protein GIT1 [OS=Homo sapiens] |
| Hydroxymethylglutaryl-CoA lyase, mitochondrial [OS=Homo sapiens] |
| Delta-1-pyrroline-5-carboxylate dehydrogenase, mitochondrial [OS=Homo sapiens] |
| cullin-associated nedd8-dissociated protein 1 [OS=Homo sapiens] |
| Desmoplakin [OS=Homo sapiens] |
| Platelet-derived growth factor subunit A [OS=Homo sapiens] |
| 26S proteasome non-ATPase regulatory subunit 2 [OS=Homo sapiens] |
| peptidyl-prolyl cis-trans isomerase FKBP3 [OS=Homo sapiens] |
| Protein phosphatase 1B [OS=Homo sapiens] |
| Lysosome-associated membrane glycoprotein 1 [OS=Homo sapiens] |
| Proline and serine-rich protein 2 [OS=Homo sapiens] |
| UV excision repair protein RAD23 homolog B [OS=Homo sapiens] |
| Claudin-5 [OS=Homo sapiens] |
| ELMO domain-containing protein 2 [OS=Homo sapiens] |
| Isoform 2 of Eukaryotic initiation factor 4A-II [OS=Homo sapiens] |
| Coatomer subunit beta [OS=Homo sapiens] |
| Keratin, type II cytoskeletal 1 [OS=Homo sapiens] |
| Tubulin-specific chaperone A [OS=Homo sapiens] |
| Plexin domain-containing protein 2 [OS=Homo sapiens] |
| DnaJ homolog subfamily C member 12 [OS=Homo sapiens] |
| Small membrane A-kinase anchor protein [OS=Homo sapiens] |
| Cullin-2 [OS=Homo sapiens] |
| Short transient receptor potential channel 6 [OS=Homo sapiens] |
| Ras-related protein Rab-5B [OS=Homo sapiens] |
| 26S proteasome regulatory subunit 6A [OS=Homo sapiens] |
| Isoform 3 of Polypyrimidine tract-binding protein 1 [OS=Homo sapiens] |
| Endoplasmic reticulum resident protein 44 [OS=Homo sapiens] |
| Isoform 2 of B-cell receptor-associated protein 29 [OS=Homo sapiens] |
| Immunoglobulin heavy variable 3-43D [OS=Homo sapiens] |
| OCIA domain-containing protein 1 [OS=Homo sapiens] |
| Stathmin [OS=Homo sapiens] |
| Serine/threonine-protein phosphatase PP1-beta catalytic subunit [OS=Homo sapiens] |

| |
|---|
| V-type proton ATPase subunit d 1 [OS=Homo sapiens] |
| Serine/threonine-protein phosphatase 2A 55 kDa regulatory subunit B alpha isoform [OS=Homo sapiens] |
| Peptidyl-prolyl cis-trans isomerase FKBP1A [OS=Homo sapiens] |
| Dermcidin [OS=Homo sapiens] |
| serpin B6 [OS=Homo sapiens] |
| Acylamino-acid-releasing enzyme [OS=Homo sapiens] |
| Golgi SNAP receptor complex member 1 [OS=Homo sapiens] |
| Ras-related protein Rab-1A [OS=Homo sapiens] |
| DnaJ homolog subfamily A member 2 [OS=Homo sapiens] |
| Alpha-centractin [OS=Homo sapiens] |
| keratin, type II cytoskeletal 5 [OS=Homo sapiens] |
| Low molecular weight phosphotyrosine protein phosphatase [OS=Homo sapiens] |
| Keratinocyte proline-rich protein [OS=Homo sapiens] |
| Protein kinase C theta type [OS=Homo sapiens] |
| 26S proteasome non-ATPase regulatory subunit 9 [OS=Homo sapiens] |
| Flotillin-1 [OS=Homo sapiens] |
| Ras-related protein Rab-35 [OS=Homo sapiens] |
| CTD small phosphatase-like protein [OS=Homo sapiens] |
| ubiquitin carboxyl-terminal hydrolase 8 [OS=Homo sapiens] |
| Retinoic acid receptor responder protein 2 [OS=Homo sapiens] |
| Rho-related GTP-binding protein RhoF [OS=Homo sapiens] |
| Golgi-associated plant pathogenesis-related protein 1 [OS=Homo sapiens] |
| Complement C1q subcomponent subunit B [OS=Homo sapiens] |
| Elongin-B [OS=Homo sapiens] |
| EH domain-binding protein 1-like protein 1 [OS=Homo sapiens] |
| vacuolar protein sorting-associated protein 28 homolog [OS=Homo sapiens] |
| Microtubule-associated protein RP/EB family member 2 [OS=Homo sapiens] |
| Calpain-2 catalytic subunit [OS=Homo sapiens] |
| caax prenyl protease 1 homolog [OS=Homo sapiens] |
| Lipopolysaccharide-responsive and beige-like anchor protein [OS=Homo sapiens] |
| Nucleosome assembly protein 1-like 4 [OS=Homo sapiens] |
| D-dopachrome decarboxylase [OS=Homo sapiens] |
| Low affinity immunoglobulin epsilon Fc receptor [OS=Homo sapiens] |
| serine/threonine-protein kinase 26 [OS=Homo sapiens] |
| Transforming protein RhoA [OS=Homo sapiens] |
| Tyrosine-protein phosphatase non-receptor type 1 [OS=Homo sapiens] |
| Immunoglobulin heavy variable 1-18 [OS=Homo sapiens] |
| Coronin-1B [OS=Homo sapiens] |
| Isoform 2 of AP-2 complex subunit mu [OS=Homo sapiens] |
| Hemoglobin subunit alpha [OS=Homo sapiens] |
| transcription elongation factor A protein-like 1 [OS=Homo sapiens] |
| Catenin delta-1 [OS=Homo sapiens] |
| Succinate dehydrogenase [ubiquinone] iron-sulfur subunit, mitochondrial [OS=Homo sapiens] |
| Adipocyte plasma membrane-associated protein [OS=Homo sapiens] |
| Gamma-soluble NSF attachment protein [OS=Homo sapiens] |
| Golgin subfamily A member 6B [OS=Homo sapiens] |
| Small glutamine-rich tetratricopeptide repeat-containing protein alpha [OS=Homo sapiens] |

| |
|---|
| cathelicidin antimicrobial peptide [OS=Homo sapiens] |
| glutathione synthetase [OS=Homo sapiens] |
| antigen peptide transporter 2 [OS=Homo sapiens] |
| Leukocyte cell-derived chemotaxin-2 [OS=Homo sapiens] |
| Leucine-rich repeat transmembrane protein FLRT2 [OS=Homo sapiens] |
| Eukaryotic translation initiation factor 5B [OS=Homo sapiens] |
| 1,4-alpha-glucan-branching enzyme [OS=Homo sapiens] |
| 26S proteasome non-ATPase regulatory subunit 11 [OS=Homo sapiens] |
| prefoldin subunit 5 [OS=Homo sapiens] |
| Isoform 2 of MICOS complex subunit MIC60 [OS=Homo sapiens] |
| Hematopoietic lineage cell-specific protein [OS=Homo sapiens] |
| Destrin [OS=Homo sapiens] |
| Importin-5 [OS=Homo sapiens] |
| Partner of Y14 and mago [OS=Homo sapiens] |
| SEC14-like protein 5 [OS=Homo sapiens] |
| Dynamin-3 [OS=Homo sapiens] |
| Coatomer subunit gamma-2 [OS=Homo sapiens] |
| Rho-related GTP-binding protein RhoG [OS=Homo sapiens] |
| Caspase-3 [OS=Homo sapiens] |
| Malate dehydrogenase, mitochondrial [OS=Homo sapiens] |
| Torsin-4A [OS=Homo sapiens] |
| Serine/threonine-protein phosphatase 2B catalytic subunit alpha isoform [OS=Homo sapiens] |
| Actin-binding LIM protein 3 [OS=Homo sapiens] |
| S-formylglutathione hydrolase [OS=Homo sapiens] |
| Germinal center-associated signaling and motility-like protein [OS=Homo sapiens] |
| Asporin [OS=Homo sapiens] |
| Isoform 3 of GTPase-activating protein and VPS9 domain-containing protein 1 [OS=Homo sapiens] |
| Short/branched chain specific acyl-CoA dehydrogenase, mitochondrial [OS=Homo sapiens] |
| V-type proton ATPase 16 kDa proteolipid subunit [OS=Homo sapiens] |
| Exocyst complex component 4 [OS=Homo sapiens] |
| Isoform HCC-3 of C-C motif chemokine 14 [OS=Homo sapiens] |
| Type II inositol 1,4,5-trisphosphate 5-phosphatase [OS=Homo sapiens] |
| bleomycin hydrolase [OS=Homo sapiens] |
| Protein disulfide-isomerase A4 [OS=Homo sapiens] |
| cytoplasmic FMR1-interacting protein 1 [OS=Homo sapiens] |
| Translin-associated protein X [OS=Homo sapiens] |
| FGFR1 oncogene partner 2 [OS=Homo sapiens] |
| Nascent polypeptide-associated complex subunit alpha, muscle-specific form [OS=Homo sapiens] |
| Isoform 2 of 26S proteasome regulatory subunit 6B [OS=Homo sapiens] |
| Peptidyl-prolyl cis-trans isomerase FKBP5 [OS=Homo sapiens] |
| Exportin-4 [OS=Homo sapiens] |
| Translation machinery-associated protein 7 [OS=Homo sapiens] |
| Coiled-coil domain-containing protein 168 [OS=Homo sapiens] |
| RUN and FYVE domain-containing protein 1 [OS=Homo sapiens] |
| Endoplasmic reticulum resident protein 29 [OS=Homo sapiens] |
| Proteasome subunit beta type-7 [OS=Homo sapiens] |
| Bifunctional glutamate/proline--tRNA ligase [OS=Homo sapiens] |

| |
|---|
| Unconventional myosin-XVIIIa [OS=Homo sapiens] |
| Isoform 3 of Nucleoside diphosphate kinase B [OS=Homo sapiens] |
| Charged multivesicular body protein 4b [OS=Homo sapiens] |
| Serine/threonine-protein phosphatase Pgam5, mitochondrial [OS=Homo sapiens] |
| vesicle-associated membrane protein 8 [OS=Homo sapiens] |
| Rab3 GTPase-activating protein non-catalytic subunit [OS=Homo sapiens] |
| Calumenin [OS=Homo sapiens] |
| Cytochrome b-c1 complex subunit 2, mitochondrial [OS=Homo sapiens] |
| Ubiquitin carboxyl-terminal hydrolase 14 [OS=Homo sapiens] |
| vesicle-associated membrane protein-associated protein A [OS=Homo sapiens] |
| Lipoma-preferred partner [OS=Homo sapiens] |
| Calcium uptake protein 1, mitochondrial [OS=Homo sapiens] |
| Cytospin-B [OS=Homo sapiens] |
| Echinoderm microtubule-associated protein-like 4 [OS=Homo sapiens] |
| LanC-like protein 1 [OS=Homo sapiens] |
| HLA class I histocompatibility antigen, B-18 alpha chain [OS=Homo sapiens] |
| TBC1 domain family member 9B [OS=Homo sapiens] |
| insulin-degrading enzyme [OS=Homo sapiens] |
| Complement component C8 gamma chain [OS=Homo sapiens] |
| Mitochondrial carrier homolog 1 [OS=Homo sapiens] |
| cGMP-inhibited 3',5'-cyclic phosphodiesterase A [OS=Homo sapiens] |
| electron transfer flavoprotein subunit beta [OS=Homo sapiens] |
| Docking protein 2 [OS=Homo sapiens] |
| Peptidyl-prolyl cis-trans isomerase NIMA-interacting 4 [OS=Homo sapiens] |
| Proteasome subunit alpha type-1 [OS=Homo sapiens] |
| Isoform 2 of Protein SLFN14 [OS=Homo sapiens] |
| Lymphocyte antigen 6 complex locus protein G6f [OS=Homo sapiens] |
| Hypoxia up-regulated protein 1 [OS=Homo sapiens] |
| Tripartite motif-containing protein 5 [OS=Homo sapiens] |
| Cytokine receptor-like factor 3 [OS=Homo sapiens] |
| Isoform 2 of Rap1 GTPase-activating protein 2 [OS=Homo sapiens] |
| tetraspanin-9 [OS=Homo sapiens] |
| Immunoglobulin kappa variable 2D-40 [OS=Homo sapiens] |
| Regulator of G-protein signaling 18 [OS=Homo sapiens] |
| GTP-binding protein SAR1a [OS=Homo sapiens] |
| Immunoglobulin heavy variable 3-30 [OS=Homo sapiens] |
| Ubiquitin-fold modifier 1 [OS=Homo sapiens] |
| Amine oxidase [flavin-containing] B [OS=Homo sapiens] |
| Receptor-type tyrosine-protein phosphatase alpha [OS=Homo sapiens] |
| Isoform 4 of Dipeptidyl peptidase 3 [OS=Homo sapiens] |
| cholesteryl ester transfer protein [OS=Homo sapiens] |
| immunoglobulin heavy variable 3-74 [OS=Homo sapiens] |
| Ras-related protein Rab-33A [OS=Homo sapiens] |
| coiled-coil domain-containing protein 6 [OS=Homo sapiens] |
| TRAF-type zinc finger domain-containing protein 1 [OS=Homo sapiens] |
| mutS protein homolog 5 [OS=Homo sapiens] |
| Protein SGT1 homolog [OS=Homo sapiens] |

| |
|---|
| Isoform 3 of Exportin-2 [OS=Homo sapiens] |
| Rho GDP-dissociation inhibitor 2 [OS=Homo sapiens] |
| Transthyretin [OS=Homo sapiens] |
| Actin-related protein 2/3 complex subunit 3 [OS=Homo sapiens] |
| rho GDP-dissociation inhibitor 1 [OS=Homo sapiens] |
| ATPase ASNA1 [OS=Homo sapiens] |
| docking protein 1 [OS=Homo sapiens] |
| density-regulated protein [OS=Homo sapiens] |
| Isoform 4 of Forkhead box protein M1 [OS=Homo sapiens] |
| Integrin alpha-5 [OS=Homo sapiens] |
| cathepsin W [OS=Homo sapiens] |
| Transmembrane protein 43 [OS=Homo sapiens] |
| Zinc transporter SLC39A7 [OS=Homo sapiens] |
| 14-3-3 protein gamma [OS=Homo sapiens] |
| Immunoglobulin lambda variable 3-19 [OS=Homo sapiens] |
| Tubulin alpha-1B chain [OS=Homo sapiens] |
| phosphatidylinositol 5-phosphate 4-kinase type-2 alpha [OS=Homo sapiens] |
| Peroxisomal acyl-coenzyme A oxidase 1 [OS=Homo sapiens] |
| Uncharacterized protein KIAA1211 [OS=Homo sapiens] |
| Protein kinase C alpha type [OS=Homo sapiens] |
| Ras-related protein Rab-4B [OS=Homo sapiens] |
| pleckstrin homology domain-containing family A member 1 [OS=Homo sapiens] |
| TBC1 domain family member 10B [OS=Homo sapiens] |
| Atlastin-1 [OS=Homo sapiens] |
| thioredoxin [OS=Homo sapiens] |
| espin [OS=Homo sapiens] |
| Eukaryotic translation initiation factor 4 gamma 3 [OS=Homo sapiens] |
| 26S proteasome non-ATPase regulatory subunit 6 [OS=Homo sapiens] |
| Mannose-1-phosphate guanyltransferase alpha [OS=Homo sapiens] |
| Isoform 2 of Septin-9 [OS=Homo sapiens] |
| Isoform 2 of Reticulon-3 [OS=Homo sapiens] |
| Sulfhydryl oxidase 1 [OS=Homo sapiens] |
| Dolichol-phosphate mannosyltransferase subunit 3 [OS=Homo sapiens] |
| Solute carrier family 2, facilitated glucose transporter member 3 [OS=Homo sapiens] |
| Ras-related protein Rab-5A [OS=Homo sapiens] |
| mitogen-activated protein kinase 1 [OS=Homo sapiens] |
| Mesencephalic astrocyte-derived neurotrophic factor [OS=Homo sapiens] |
| Ras-related protein Rab-5C [OS=Homo sapiens] |
| Intercellular adhesion molecule 2 [OS=Homo sapiens] |
| Annexin A2 [OS=Homo sapiens] |
| aldose reductase [OS=Homo sapiens] |
| Atlastin-3 [OS=Homo sapiens] |
| saccharopine dehydrogenase-like oxidoreductase [OS=Homo sapiens] |
| ras GTPase-activating protein 1 [OS=Homo sapiens] |
| Actin-related protein 2/3 complex subunit 1A [OS=Homo sapiens] |
| Rho-related GTP-binding protein RhoC [OS=Homo sapiens] |
| BH3-interacting domain death agonist [OS=Homo sapiens] |

| |
|---|
| Immunoglobulin lambda variable 2-14 [OS=Homo sapiens] |
| DNA damage-binding protein 1 [OS=Homo sapiens] |
| Protein phosphatase 1A [OS=Homo sapiens] |
| clathrin light chain B [OS=Homo sapiens] |
| CSC1-like protein 1 [OS=Homo sapiens] |
| UBX domain-containing protein 4 [OS=Homo sapiens] |
| myelin basic protein [OS=Homo sapiens] |
| Putative E3 ubiquitin-protein ligase SH3RF2 [OS=Homo sapiens] |
| Cas scaffolding protein family member 4 [OS=Homo sapiens] |
| peptidyl-prolyl cis-trans isomerase NIMA-interacting 1 [OS=Homo sapiens] |
| Isoform 2 of V-type proton ATPase 116 kDa subunit a isoform 1 [OS=Homo sapiens] |
| ADP-ribosylation factor 5 [OS=Homo sapiens] |
| NSFL1 cofactor p47 [OS=Homo sapiens] |
| eukaryotic translation initiation factor 2 subunit 3 [OS=Homo sapiens] |
| ras-related protein Rab-1B [OS=Homo sapiens] |
| volume-regulated anion channel subunit LRRC8D [OS=Homo sapiens] |
| Prelamin-A/C [OS=Homo sapiens] |
| matrix Gla protein [OS=Homo sapiens] |
| Plasma serine protease inhibitor [OS=Homo sapiens] |
| Inter-alpha-trypsin inhibitor heavy chain H2 [OS=Homo sapiens] |
| Endothelin-converting enzyme 1 [OS=Homo sapiens] |
| SPRY domain-containing protein 7 [OS=Homo sapiens] |
| Platelet glycoprotein IX [OS=Homo sapiens] |
| Glycogen phosphorylase, brain form [OS=Homo sapiens] |
| phosphomevalonate kinase [OS=Homo sapiens] |
| Ras-related protein Rab-21 [OS=Homo sapiens] |
| ADP-ribosylation factor-like protein 8A [OS=Homo sapiens] |
| Isoform 3 of Sarcolemmal membrane-associated protein [OS=Homo sapiens] |
| UBX domain-containing protein 6 [OS=Homo sapiens] |
| Isoform 2 of Proteasome subunit alpha type-3 [OS=Homo sapiens] |
| Catechol O-methyltransferase [OS=Homo sapiens] |
| Acetyl-coenzyme A synthetase 2-like, mitochondrial [OS=Homo sapiens] |
| Proteasome subunit beta type-9 [OS=Homo sapiens] |
| Isoform 2 of Inverted formin-2 [OS=Homo sapiens] |
| Eukaryotic peptide chain release factor subunit 1 [OS=Homo sapiens] |
| RuvB-like 1 [OS=Homo sapiens] |
| protein LYRIC [OS=Homo sapiens] |
| Smoothelin [OS=Homo sapiens] |
| ragulator complex protein LAMTOR1 [OS=Homo sapiens] |
| tetratricopeptide repeat protein 27 [OS=Homo sapiens] |
| phosphatidylethanolamine-binding protein 1 [OS=Homo sapiens] |
| Cylicin-2 [OS=Homo sapiens] |
| Glyoxalase domain-containing protein 4 [OS=Homo sapiens] |
| glia maturation factor gamma [OS=Homo sapiens] |
| Uncharacterized protein KIAA0513 [OS=Homo sapiens] |
| Elongation factor 1-delta [OS=Homo sapiens] |
| Phosphatidylinositol 4,5-bisphosphate 3-kinase catalytic subunit beta isoform [OS=Homo sapiens] |

| |
|---|
| Guanine nucleotide-binding protein subunit beta-4 [OS=Homo sapiens] |
| serine/threonine-protein phosphatase 2A catalytic subunit beta isoform [OS=Homo sapiens] |
| immunoglobulin kappa variable 3D-20 [OS=Homo sapiens] |
| tubulin beta chain [OS=Homo sapiens] |
| Glycerol-3-phosphate dehydrogenase, mitochondrial [OS=Homo sapiens] |
| unconventional myosin-Vb [OS=Homo sapiens] |
| Dolichyl-diphosphooligosaccharide--protein glycosyltransferase subunit DAD1 [OS=Homo sapiens] |
| Isoform Cytoplasmic of Glutathione reductase, mitochondrial [OS=Homo sapiens] |
| 26S proteasome regulatory subunit 10B [OS=Homo sapiens] |
| Alpha-actinin-4 [OS=Homo sapiens] |
| corneodesmosin [OS=Homo sapiens] |
| obg-like ATPase 1 [OS=Homo sapiens] |
| C4b-binding protein beta chain [OS=Homo sapiens] |
| Isoform 8 of Abl interactor 1 [OS=Homo sapiens] |
| Ubiquitin-conjugating enzyme E2 L3 [OS=Homo sapiens] |
| Mitochondrial carrier homolog 2 [OS=Homo sapiens] |
| Transcription factor A, mitochondrial [OS=Homo sapiens] |
| Isoform 5 of Oxidation resistance protein 1 [OS=Homo sapiens] |
| Complement factor I [OS=Homo sapiens] |
| Complement C1r subcomponent [OS=Homo sapiens] |
| Retinol dehydrogenase 11 [OS=Homo sapiens] |
| Histone-lysine N-methyltransferase 2A [OS=Homo sapiens] |
| vasorin [OS=Homo sapiens] |
| Isoform 2 of Leucine-rich repeat flightless-interacting protein 1 [OS=Homo sapiens] |
| Beta-Ala-His dipeptidase [OS=Homo sapiens] |
| mitochondrial fission 1 protein [OS=Homo sapiens] |
| Phospholipid hydroperoxide glutathione peroxidase, mitochondrial [OS=Homo sapiens] |
| Glutathione peroxidase 1 [OS=Homo sapiens] |
| Adenylate kinase isoenzyme 1 [OS=Homo sapiens] |
| Heterogeneous nuclear ribonucleoprotein K [OS=Homo sapiens] |
| Dual adapter for phosphotyrosine and 3-phosphotyrosine and 3-phosphoinositide [OS=Homo sapiens] |
| Peptidyl-prolyl cis-trans isomerase FKBP2 [OS=Homo sapiens] |
| Beta-adrenergic receptor kinase 1 [OS=Homo sapiens] |
| Long-chain-fatty-acid--CoA ligase 4 [OS=Homo sapiens] |
| protein Shroom3 [OS=Homo sapiens] |
| protein CLEC16A [OS=Homo sapiens] |
| WAS/WASL-interacting protein family member 1 [OS=Homo sapiens] |
| Fatty acid synthase [OS=Homo sapiens] |
| Vesicle-trafficking protein SEC22b [OS=Homo sapiens] |
| N-acylglucosamine 2-epimerase [OS=Homo sapiens] |
| Phospholipid-transporting ATPase IA [OS=Homo sapiens] |
| Bridging integrator 2 [OS=Homo sapiens] |
| CD63 antigen [OS=Homo sapiens] |
| Malate dehydrogenase, cytoplasmic [OS=Homo sapiens] |
| Immunoglobulin lambda variable 1-47 [OS=Homo sapiens] |
| 6-phosphogluconolactonase [OS=Homo sapiens] |
| Serine/threonine-protein kinase TAO3 [OS=Homo sapiens] |

| |
|---|
| Ficolin-3 [OS=Homo sapiens] |
| Very-long-chain 3-oxoacyl-CoA reductase [OS=Homo sapiens] |
| Alpha-adducin [OS=Homo sapiens] |
| COP9 signalosome complex subunit 8 [OS=Homo sapiens] |
| Pyridoxal phosphate homeostasis protein [OS=Homo sapiens] |
| Fibrocystin-L [OS=Homo sapiens] |
| Eukaryotic initiation factor 4A-I [OS=Homo sapiens] |
| Protein disulfide-isomerase A5 [OS=Homo sapiens] |
| fructosamine-3-kinase [OS=Homo sapiens] |
| Gelsolin [OS=Homo sapiens] |
| Endophilin-A2 [OS=Homo sapiens] |
| Type 2 lactosamine alpha-2,3-sialyltransferase [OS=Homo sapiens] |
| TBC1 domain family member 5 [OS=Homo sapiens] |
| Heat shock 70 kDa protein 4 [OS=Homo sapiens] |
| Cytoplasmic dynein 1 light intermediate chain 1 [OS=Homo sapiens] |
| Isoform 2 of Histidine--tRNA ligase, cytoplasmic [OS=Homo sapiens] |
| Isoform 2 of Ubiquitin-associated protein 2-like [OS=Homo sapiens] |
| Adenylate kinase 2, mitochondrial [OS=Homo sapiens] |
| immunoglobulin lambda variable 1-51 [OS=Homo sapiens] |
| Ubiquitin carboxyl-terminal hydrolase 4 [OS=Homo sapiens] |
| septin-2 [OS=Homo sapiens] |
| dynactin subunit 2 [OS=Homo sapiens] |
| AP-1 complex subunit beta-1 [OS=Homo sapiens] |
| Transmembrane emp24 domain-containing protein 10 [OS=Homo sapiens] |
| Filaggrin [OS=Homo sapiens] |
| TSC22 domain family protein 4 [OS=Homo sapiens] |
| Peroxiredoxin-2 [OS=Homo sapiens] |
| peptidyl-prolyl cis-trans isomerase B [OS=Homo sapiens] |
| Protein ABHD16A [OS=Homo sapiens] |
| Transforming growth factor beta-1 [OS=Homo sapiens] |
| FYVE, RhoGEF and PH domain-containing protein 4 [OS=Homo sapiens] |
| Isoform 2 of Mitochondrial 2-oxoglutarate/malate carrier protein [OS=Homo sapiens] |
| Cytosolic 5'-nucleotidase 3A [OS=Homo sapiens] |
| Cytochrome c oxidase subunit 5B, mitochondrial [OS=Homo sapiens] |
| Calcium-transporting ATPase type 2C member 1 [OS=Homo sapiens] |
| Haptoglobin-related protein [OS=Homo sapiens] |
| Rho-associated protein kinase 1 [OS=Homo sapiens] |
| Protein piccolo [OS=Homo sapiens] |
| Dual specificity mitogen-activated protein kinase kinase 2 [OS=Homo sapiens] |
| Isoform 2 of Dynamin-like 120 kDa protein, mitochondrial [OS=Homo sapiens] |
| MAP kinase-activated protein kinase 2 [OS=Homo sapiens] |
| Rab3 GTPase-activating protein catalytic subunit [OS=Homo sapiens] |
| Isoform 2 of Keratinocyte differentiation-associated protein [OS=Homo sapiens] |
| Isoform 3 of Deoxyhypusine synthase [OS=Homo sapiens] |
| Synaptosomal-associated protein 23 [OS=Homo sapiens] |
| ATPase inhibitor, mitochondrial [OS=Homo sapiens] |
| Sorting nexin-12 [OS=Homo sapiens] |

| |
|---|
| Talin-2 [OS=Homo sapiens] |
| TOM1-like protein 2 [OS=Homo sapiens] |
| Mannan-binding lectin serine protease 1 [OS=Homo sapiens] |
| Isoform 3 of Inositol monophosphatase 1 [OS=Homo sapiens] |
| Coagulation factor XIII A chain [OS=Homo sapiens] |
| Cytochrome c oxidase subunit 6B1 [OS=Homo sapiens] |
| Tyrosine--tRNA ligase, cytoplasmic [OS=Homo sapiens] |
| voltage-dependent anion-selective channel protein 1 [OS=Homo sapiens] |
| Keratin, type II cytoskeletal 74 [OS=Homo sapiens] |
| Fermitin family homolog 3 [OS=Homo sapiens] |
| Kinesin-like protein KIF21B [OS=Homo sapiens] |
| charged multivesicular body protein 2a [OS=Homo sapiens] |
| cysteine and histidine-rich domain-containing protein 1 [OS=Homo sapiens] |
| Serum amyloid P-component [OS=Homo sapiens] |
| AP-2 complex subunit alpha-1 [OS=Homo sapiens] |
| Isoform 12 of Sorbin and SH3 domain-containing protein 1 [OS=Homo sapiens] |
| Serine/threonine-protein kinase TAO1 [OS=Homo sapiens] |
| Olfactory receptor 4C6 [OS=Homo sapiens] |
| Isoform 4 of Extracellular matrix protein 1 [OS=Homo sapiens] |
| DENN domain-containing protein 3 [OS=Homo sapiens] |
| Elongation factor 2 [OS=Homo sapiens] |
| Tyrosine-protein phosphatase non-receptor type 12 [OS=Homo sapiens] |
| immunoglobulin gamma-1 heavy chain [OS=Homo sapiens] |
| T-complex protein 1 subunit alpha [OS=Homo sapiens] |
| Isoform 3 of Zinc finger protein 185 [OS=Homo sapiens] |
| bromodomain and WD repeat-containing protein 1 [OS=Homo sapiens] |
| ADP-sugar pyrophosphatase [OS=Homo sapiens] |
| Complement factor D [OS=Homo sapiens] |
| Spectrin alpha chain, non-erythrocytic 1 [OS=Homo sapiens] |
| Acyl-CoA dehydrogenase family member 9, mitochondrial [OS=Homo sapiens] |
| Tandem C2 domains nuclear protein [OS=Homo sapiens] |
| T-complex protein 1 subunit gamma [OS=Homo sapiens] |
| ubiquitin-associated and SH3 domain-containing protein B [OS=Homo sapiens] |
| Transmembrane emp24 domain-containing protein 5 [OS=Homo sapiens] |
| Isoform BTK-C of Tyrosine-protein kinase BTK [OS=Homo sapiens] |
| Isoform 5 of Tropomyosin alpha-1 chain [OS=Homo sapiens] |
| Probable ubiquitin carboxyl-terminal hydrolase FAF-Y [OS=Homo sapiens] |
| Mannan-binding lectin serine protease 2 [OS=Homo sapiens] |
| Nucleoside diphosphate-linked moiety X motif 17 [OS=Homo sapiens] |
| Tyrosine-protein kinase SYK [OS=Homo sapiens] |
| Uncharacterized protein C10orf131 [OS=Homo sapiens] |
| Heparanase [OS=Homo sapiens] |
| Nexilin [OS=Homo sapiens] |
| MICOS complex subunit MIC19 [OS=Homo sapiens] |
| Isoform 5 of Four and a half LIM domains protein 1 [OS=Homo sapiens] |
| Vacuolar protein sorting-associated protein 26B [OS=Homo sapiens] |
| C-1-tetrahydrofolate synthase, cytoplasmic [OS=Homo sapiens] |

| |
|---|
| Testis-expressed protein 264 [OS=Homo sapiens] |
| ATP synthase subunit epsilon-like protein, mitochondrial [OS=Homo sapiens] |
| Nesprin-2 [OS=Homo sapiens] |
| Ras-related protein R-Ras [OS=Homo sapiens] |
| Brain-enriched guanylate kinase-associated protein [OS=Homo sapiens] |
| Myosin regulatory light polypeptide 9 [OS=Homo sapiens] |
| Beta-2-microglobulin [OS=Homo sapiens] |
| ADP/ATP translocase 3 [OS=Homo sapiens] |
| Serine hydroxymethyltransferase, mitochondrial [OS=Homo sapiens] |
| Ras-related protein Rab-32 [OS=Homo sapiens] |
| Isoform 7 of Complement decay-accelerating factor [OS=Homo sapiens] |
| Isoform 4 of Formin-binding protein 1-like [OS=Homo sapiens] |
| Profilin-3 [OS=Homo sapiens] |
| Isoform 4 of Vacuolar protein sorting-associated protein 13C [OS=Homo sapiens] |
| Microtubule-associated protein 1A [OS=Homo sapiens] |
| Isoform 3 of Coronin-1C [OS=Homo sapiens] |
| GTPase NRas [OS=Homo sapiens] |
| C-type lectin domain family 1 member B [OS=Homo sapiens] |
| Endothelial cell-selective adhesion molecule [OS=Homo sapiens] |
| ras-related protein Rap-2b [OS=Homo sapiens] |
| Lambda-crystallin homolog [OS=Homo sapiens] |
| Serine/threonine-protein kinase 10 [OS=Homo sapiens] |
| Isoform 5 of Obscurin [OS=Homo sapiens] |
| AP-1 complex subunit gamma-1 [OS=Homo sapiens] |
| GTP-binding protein 2 [OS=Homo sapiens] |
| proteasome subunit beta type-1 [OS=Homo sapiens] |
| Mitochondrial import receptor subunit TOM70 [OS=Homo sapiens] |
| Isoform 3 of Tropomyosin alpha-1 chain [OS=Homo sapiens] |
| Alstrom syndrome protein 1 [OS=Homo sapiens] |
| Proteasome subunit alpha type-5 [OS=Homo sapiens] |
| NADH dehydrogenase [ubiquinone] 1 beta subcomplex subunit 11, mitochondrial [OS=Homo sapiens] |
| 28 kDa heat- and acid-stable phosphoprotein [OS=Homo sapiens] |
| C-Maf-inducing protein [OS=Homo sapiens] |
| Alpha-synuclein [OS=Homo sapiens] |
| Isoform 2 of Tropomyosin alpha-4 chain [OS=Homo sapiens] |
| Isoform 2 of Kinesin-like protein KIF2A [OS=Homo sapiens] |
| Alpha-1B-glycoprotein [OS=Homo sapiens] |
| Junctional adhesion molecule A [OS=Homo sapiens] |
| cartilage acidic protein 1 [OS=Homo sapiens] |
| T-complex protein 1 subunit zeta [OS=Homo sapiens] |
| Cytosolic phospholipase A2 [OS=Homo sapiens] |
| Transcriptional activator protein Pur-beta [OS=Homo sapiens] |
| Coatomer subunit beta' [OS=Homo sapiens] |
| chloride intracellular channel protein 1 [OS=Homo sapiens] |
| Immunoglobulin lambda-1 light chain [OS=Homo sapiens] |
| Protein CDV3 homolog [OS=Homo sapiens] |
| Ras suppressor protein 1 [OS=Homo sapiens] |

| |
|---|
| Tubulin alpha-1C chain [OS=Homo sapiens] |
| Sorcin [OS=Homo sapiens] |
| Metastasis suppressor protein 1 [OS=Homo sapiens] |
| UTP--glucose-1-phosphate uridylyltransferase [OS=Homo sapiens] |
| alpha-1-antitrypsin [OS=Homo sapiens] |
| E3 ubiquitin-protein ligase RNF213 [OS=Homo sapiens] |
| Mycophenolic acid acyl-glucuronide esterase, mitochondrial [OS=Homo sapiens] |
| mitochondrial carnitine/acylcarnitine carrier protein [OS=Homo sapiens] |
| Proto-oncogene vav [OS=Homo sapiens] |
| V-type immunoglobulin domain-containing suppressor of T-cell activation [OS=Homo sapiens] |
| WW domain-binding protein 2 [OS=Homo sapiens] |
| 1-phosphatidylinositol 4,5-bisphosphate phosphodiesterase gamma-2 [OS=Homo sapiens] |
| Ceramide synthase 2 [OS=Homo sapiens] |
| Sodium channel and clathrin linker 1 [OS=Homo sapiens] |
| Long-chain-fatty-acid--CoA ligase 5 [OS=Homo sapiens] |
| rho GTPase-activating protein 21 [OS=Homo sapiens] |
| Isoform 2 of Gelsolin [OS=Homo sapiens] |
| Sparc [OS=Homo sapiens] |
| V-type proton ATPase catalytic subunit A [OS=Homo sapiens] |
| Transaldolase [OS=Homo sapiens] |
| ubiquitin-conjugating enzyme E2 N [OS=Homo sapiens] |
| Keratin, type I cytoskeletal 14 [OS=Homo sapiens] |
| Signal transducer and activator of transcription 3 [OS=Homo sapiens] |
| SH3 domain-binding glutamic acid-rich-like protein 3 [OS=Homo sapiens] |
| Beta-adducin [OS=Homo sapiens] |
| Single-stranded DNA-binding protein, mitochondrial [OS=Homo sapiens] |
| Isoform Beta of cGMP-dependent protein kinase 1 [OS=Homo sapiens] |
| Nck-associated protein 1 [OS=Homo sapiens] |
| N-acetylmuramoyl-L-alanine amidase [OS=Homo sapiens] |
| TBC1 domain family member 23 [OS=Homo sapiens] |
| Protein transport protein Sec31A [OS=Homo sapiens] |
| cytosolic non-specific dipeptidase [OS=Homo sapiens] |
| Heat shock protein beta-1 [OS=Homo sapiens] |
| alpha-soluble nsf attachment protein [OS=Homo sapiens] |
| Cytochrome c oxidase subunit 5A, mitochondrial [OS=Homo sapiens] |
| programmed cell death protein 10 [OS=Homo sapiens] |
| Protein NYNRIN [OS=Homo sapiens] |
| Septin-11 [OS=Homo sapiens] |
| Trem-like transcript 1 protein [OS=Homo sapiens] |
| Isoform 4 of Leucine-rich repeat flightless-interacting protein 2 [OS=Homo sapiens] |
| Nucleoside diphosphate kinase A [OS=Homo sapiens] |
| Serine/threonine-protein phosphatase 6 catalytic subunit [OS=Homo sapiens] |
| Neutral alpha-glucosidase AB [OS=Homo sapiens] |
| Extended synaptotagmin-2 [OS=Homo sapiens] |
| Exocyst complex component 3-like protein 4 [OS=Homo sapiens] |
| Dynactin subunit 1 [OS=Homo sapiens] |
| Protocadherin-17 [OS=Homo sapiens] |

| |
|---|
| Inter-alpha-trypsin inhibitor heavy chain H3 [OS=Homo sapiens] |
| Proteasome subunit beta type-4 [OS=Homo sapiens] |
| Cytochrome c oxidase subunit NDUFA4 [OS=Homo sapiens] |
| COP9 signalosome complex subunit 3 [OS=Homo sapiens] |
| High mobility group protein HMG-I/HMG-Y [OS=Homo sapiens] |
| Glutamate dehydrogenase 1, mitochondrial [OS=Homo sapiens] |
| Dihydrolipoyllysine-residue succinyltransferase component of 2-oxoglutarate dehydrogenase complex, mito |
| Superoxide dismutase [Mn], mitochondrial [OS=Homo sapiens] |
| immunoglobulin kappa light chain [OS=Homo sapiens] |
| pantothenate kinase 4 [OS=Homo sapiens] |
| Cytochrome c oxidase subunit 3 [OS=Homo sapiens] |
| Leucine-rich repeat-containing protein 59 [OS=Homo sapiens] |
| Isoform 4 of Calumenin [OS=Homo sapiens] |
| Carnitine O-acetyltransferase [OS=Homo sapiens] |
| NAD-dependent malic enzyme, mitochondrial [OS=Homo sapiens] |
| Nucleosome assembly protein 1-like 1 [OS=Homo sapiens] |
| Platelet basic protein [OS=Homo sapiens] |
| Glycine--tRNA ligase [OS=Homo sapiens] |
| Isoform 2 of CKLF-like MARVEL transmembrane domain-containing protein 5 [OS=Homo sapiens] |
| ES1 protein homolog, mitochondrial [OS=Homo sapiens] |
| inositol-trisphosphate 3-kinase B [OS=Homo sapiens] |
| Reticulon-4 [OS=Homo sapiens] |
| B-cell receptor-associated protein 31 [OS=Homo sapiens] |
| ADP/ATP translocase 2 [OS=Homo sapiens] |
| E3 ubiquitin-protein ligase TRIM58 [OS=Homo sapiens] |
| Protein AMBP [OS=Homo sapiens] |
| Isoform 3B of Myosin light chain kinase, smooth muscle [OS=Homo sapiens] |
| Ras-related protein Rab-4A [OS=Homo sapiens] |
| multifunctional protein ADE2 [OS=Homo sapiens] |
| Kinesin-1 heavy chain [OS=Homo sapiens] |
| Isoform 6 of Kalirin [OS=Homo sapiens] |
| Protein lunapark [OS=Homo sapiens] |
| tubulin-folding cofactor B [OS=Homo sapiens] |
| immunoglobulin lambda variable 3-10 [OS=Homo sapiens] |
| Cytochrome b-c1 complex subunit 6, mitochondrial [OS=Homo sapiens] |
| Tetraspanin-33 [OS=Homo sapiens] |
| Isoform 2 of B-cell scaffold protein with ankyrin repeats [OS=Homo sapiens] |
| Angiotensinogen [OS=Homo sapiens] |
| Cytochrome b5 type B [OS=Homo sapiens] |
| Glutathione S-transferase P [OS=Homo sapiens] |
| NADH-cytochrome b5 reductase 1 [OS=Homo sapiens] |
| Charged multivesicular body protein 3 [OS=Homo sapiens] |
| CD9 antigen [OS=Homo sapiens] |
| AP-1 complex subunit mu-1 [OS=Homo sapiens] |
| Aspartate--tRNA ligase, cytoplasmic [OS=Homo sapiens] |
| sterol-4-alpha-carboxylate 3-dehydrogenase, decarboxylating [OS=Homo sapiens] |
| Activating signal cointegrator 1 complex subunit 3 [OS=Homo sapiens] |

| |
|---|
| Endophilin-B2 [OS=Homo sapiens] |
| CCR4-NOT transcription complex subunit 2 [OS=Homo sapiens] |
| Platelet glycoprotein 4 [OS=Homo sapiens] |
| Protein S100-A8 [OS=Homo sapiens] |
| Syntaxin-binding protein 5 [OS=Homo sapiens] |
| CD59 glycoprotein [OS=Homo sapiens] |
| NADH dehydrogenase [ubiquinone] 1 alpha subcomplex subunit 3 [OS=Homo sapiens] |
| Enoyl-CoA hydratase, mitochondrial [OS=Homo sapiens] |
| Testis development-related protein [OS=Homo sapiens] |
| Hemopexin [OS=Homo sapiens] |
| Tyrosine-protein kinase Lyn [OS=Homo sapiens] |
| 14-3-3 protein epsilon [OS=Homo sapiens] |
| Isoform 3 of Probable hydrolase PNKD [OS=Homo sapiens] |
| Heat shock protein HSP 90-beta [OS=Homo sapiens] |
| Reticulon-1 [OS=Homo sapiens] |
| Specifically androgen-regulated gene protein [OS=Homo sapiens] |
| Coatomer subunit delta [OS=Homo sapiens] |
| GRB2-related adapter protein 2 [OS=Homo sapiens] |
| Dynamin-1-like protein [OS=Homo sapiens] |
| Vesicle-associated membrane protein-associated protein B/C [OS=Homo sapiens] |
| Coronin-1A [OS=Homo sapiens] |
| DnaJ homolog subfamily B member 11 [OS=Homo sapiens] |
| Complement component c6 [OS=Homo sapiens] |
| Biliverdin reductase A [OS=Homo sapiens] |
| Isoform 4 of Heat shock protein 105 kDa [OS=Homo sapiens] |
| Calcium-binding protein 39 [OS=Homo sapiens] |
| citrate synthase, mitochondrial [OS=Homo sapiens] |
| Vimentin [OS=Homo sapiens] |
| ATP synthase subunit O, mitochondrial [OS=Homo sapiens] |
| Retinoid-inducible serine carboxypeptidase [OS=Homo sapiens] |
| TPR and ankyrin repeat-containing protein 1 [OS=Homo sapiens] |
| Complement C1s subcomponent [OS=Homo sapiens] |
| Sarcoplasmic/endoplasmic reticulum calcium ATPase 2 [OS=Homo sapiens] |
| fumarate hydratase, mitochondrial [OS=Homo sapiens] |
| Isoform 2 of Sushi domain-containing protein 1 [OS=Homo sapiens] |
| 26S proteasome non-ATPase regulatory subunit 13 [OS=Homo sapiens] |
| Dihydrolipoyllysine-residue acetyltransferase component of pyruvate dehydrogenase complex, mitochondri[illegible] |
| Transforming growth factor beta-1-induced transcript 1 protein [OS=Homo sapiens] |
| Keratin, type I cytoskeletal 16 [OS=Homo sapiens] |
| Very long-chain specific acyl-CoA dehydrogenase, mitochondrial [OS=Homo sapiens] |
| Isoform 2 of Protein unc-45 homolog A [OS=Homo sapiens] |
| trifunctional purine biosynthetic protein adenosine-3 [OS=Homo sapiens] |
| ankyrin-1 [OS=Homo sapiens] |
| Proteoglycan 4 [OS=Homo sapiens] |
| adenylyl cyclase-associated protein 1 [OS=Homo sapiens] |
| histone deacetylase 6 [OS=Homo sapiens] |
| tyrosine-protein kinase CSK [OS=Homo sapiens] |

| |
|---|
| alpha-endosulfine [OS=Homo sapiens] |
| Immunoglobulin kappa variable 2-29 [OS=Homo sapiens] |
| Isoform 2 of TBC1 domain family member 15 [OS=Homo sapiens] |
| Isoform 2 of Disheveled-associated activator of morphogenesis 1 [OS=Homo sapiens] |
| SLAM family member 5 [OS=Homo sapiens] |
| RAS guanyl-releasing protein 2 [OS=Homo sapiens] |
| Alanine--tRNA ligase, cytoplasmic [OS=Homo sapiens] |
| Haptoglobin [OS=Homo sapiens] |
| ras-related protein Rab-7a [OS=Homo sapiens] |
| Cytochrome c oxidase subunit 4 isoform 1, mitochondrial [OS=Homo sapiens] |
| Isoform 4 of Vacuolar protein sorting-associated protein 13A [OS=Homo sapiens] |
| ATP-dependent 6-phosphofructokinase, liver type [OS=Homo sapiens] |
| Immunoglobulin alpha-2 heavy chain [OS=Homo sapiens] |
| AP-2 complex subunit beta [OS=Homo sapiens] |
| Ribosomal protein S6 kinase alpha-3 [OS=Homo sapiens] |
| Isoform 4 of Tyrosine-protein phosphatase non-receptor type 13 [OS=Homo sapiens] |
| Calreticulin [OS=Homo sapiens] |
| Isoform 2 of Inorganic pyrophosphatase 2, mitochondrial [OS=Homo sapiens] |
| ATP synthase F(0) complex subunit B1, mitochondrial [OS=Homo sapiens] |
| Isoform 4 of Serine/threonine-protein phosphatase 2A activator [OS=Homo sapiens] |
| Kinesin heavy chain isoform 5A [OS=Homo sapiens] |
| Galectin-related protein [OS=Homo sapiens] |
| Immunoglobulin lambda variable 3-25 [OS=Homo sapiens] |
| Isoform 2 of LIM and SH3 domain protein 1 [OS=Homo sapiens] |
| src kinase-associated phosphoprotein 2 [OS=Homo sapiens] |
| lymphocyte cytosolic protein 2 [OS=Homo sapiens] |
| Fermitin family homolog 2 [OS=Homo sapiens] |
| Probable phospholipid-transporting ATPase IIB [OS=Homo sapiens] |
| Calpain-1 catalytic subunit [OS=Homo sapiens] |
| Immunoglobulin kappa variable 3-15 [OS=Homo sapiens] |
| PRA1 family protein 3 [OS=Homo sapiens] |
| Succinate dehydrogenase [ubiquinone] flavoprotein subunit, mitochondrial [OS=Homo sapiens] |
| Vacuolar protein sorting-associated protein 29 [OS=Homo sapiens] |
| mitochondrial proton/calcium exchanger protein [OS=Homo sapiens] |
| Isoform 4 of TRAF2 and NCK-interacting protein kinase [OS=Homo sapiens] |
| Isoform 2 of Transmembrane protein 87B [OS=Homo sapiens] |
| Integrin-linked protein kinase [OS=Homo sapiens] |
| S-adenosylhomocysteine hydrolase-like protein 1 [OS=Homo sapiens] |
| T-complex protein 1 subunit epsilon [OS=Homo sapiens] |
| phosphatidylcholine-sterol acyltransferase [OS=Homo sapiens] |
| Myosin regulatory light chain 12B [OS=Homo sapiens] |
| elongation factor Tu, mitochondrial [OS=Homo sapiens] |
| clathrin light chain A [OS=Homo sapiens] |
| Selenoprotein P [OS=Homo sapiens] |
| Protein TMED8 [OS=Homo sapiens] |
| Vacuolar protein-sorting-associated protein 25 [OS=Homo sapiens] |
| Guanine nucleotide-binding protein G(I)/G(S)/G(T) subunit beta-1 [OS=Homo sapiens] |

| |
|---|
| Apolipoprotein E [OS=Homo sapiens] |
| Dolichol-phosphate mannosyltransferase subunit 1 [OS=Homo sapiens] |
| pleckstrin [OS=Homo sapiens] |
| Protein unc-13 homolog D [OS=Homo sapiens] |
| peptidyl-prolyl cis-trans isomerase A [OS=Homo sapiens] |
| Diablo homolog, mitochondrial [OS=Homo sapiens] |
| Isoform Cytoplasmic+peroxisomal of Peroxiredoxin-5, mitochondrial [OS=Homo sapiens] |
| carbonic anhydrase 1 [OS=Homo sapiens] |
| Receptor expression-enhancing protein 5 [OS=Homo sapiens] |
| 10 kDa heat shock protein, mitochondrial [OS=Homo sapiens] |
| PDZ and LIM domain protein 5 [OS=Homo sapiens] |
| Transmembrane protein 201 [OS=Homo sapiens] |
| elongation factor 1-gamma [OS=Homo sapiens] |
| tetraspanin-2 [OS=Homo sapiens] |
| GTP-binding protein SAR1b [OS=Homo sapiens] |
| Phosphatidylinositol-glycan-specific phospholipase D [OS=Homo sapiens] |
| T-complex protein 1 subunit beta [OS=Homo sapiens] |
| nuclear migration protein nudC [OS=Homo sapiens] |
| Myosin light polypeptide 6 [OS=Homo sapiens] |
| Protein bicaudal D homolog 1 [OS=Homo sapiens] |
| lipopolysaccharide-binding protein [OS=Homo sapiens] |
| Lumican [OS=Homo sapiens] |
| Malectin [OS=Homo sapiens] |
| Platelet glycoprotein V [OS=Homo sapiens] |
| Adenylate cyclase type 6 [OS=Homo sapiens] |
| dual specificity protein phosphatase 3 [OS=Homo sapiens] |
| Cysteine and glycine-rich protein 1 [OS=Homo sapiens] |
| syntaxin-11 [OS=Homo sapiens] |
| PDZ and LIM domain protein 1 [OS=Homo sapiens] |
| Ras-related protein Rab-8A [OS=Homo sapiens] |
| alcohol dehydrogenase class-3 [OS=Homo sapiens] |
| phospholipid transfer protein [OS=Homo sapiens] |
| Glutathione S-transferase omega-1 [OS=Homo sapiens] |
| Carbonic anhydrase 2 [OS=Homo sapiens] |
| Keratin, type I cytoskeletal 13 [OS=Homo sapiens] |
| UDP-glucose:glycoprotein glucosyltransferase 1 [OS=Homo sapiens] |
| protein kinase C delta type [OS=Homo sapiens] |
| Aspartate aminotransferase, mitochondrial [OS=Homo sapiens] |
| cAMP-dependent protein kinase type I-alpha regulatory subunit [OS=Homo sapiens] |
| Ras-related C3 botulinum toxin substrate 2 [OS=Homo sapiens] |
| Isoform 2 of Tyrosine-protein kinase Lyn [OS=Homo sapiens] |
| Fetuin-B [OS=Homo sapiens] |
| Isoform 2 of 2,4-dienoyl-CoA reductase, mitochondrial [OS=Homo sapiens] |
| Alkyldihydroxyacetonephosphate synthase, peroxisomal [OS=Homo sapiens] |
| Rap1 GTPase-GDP dissociation stimulator 1 [OS=Homo sapiens] |
| L-lactate dehydrogenase A chain [OS=Homo sapiens] |
| lysosomal protective protein [OS=Homo sapiens] |

| |
|---|
| Eukaryotic translation initiation factor 3 subunit J [OS=Homo sapiens] |
| complement component C9 [OS=Homo sapiens] |
| DnaJ homolog subfamily C member 3 [OS=Homo sapiens] |
| Mitochondrial antiviral-signaling protein [OS=Homo sapiens] |
| Complement C5 [OS=Homo sapiens] |
| Low affinity immunoglobulin gamma Fc region receptor II-c [OS=Homo sapiens] |
| catalase [OS=Homo sapiens] |
| immunoglobulin heavy constant alpha 1 [OS=Homo sapiens] |
| Isoform Long of Erythrocyte membrane protein band 4.2 [OS=Homo sapiens] |
| platelet-activating factor acetylhydrolase IB subunit beta [OS=Homo sapiens] |
| Isoform 2 of Clusterin [OS=Homo sapiens] |
| Calmodulin [OS=Homo sapiens] |
| Isocitrate dehydrogenase [NADP], mitochondrial [OS=Homo sapiens] |
| 4-trimethylaminobutyraldehyde dehydrogenase [OS=Homo sapiens] |
| Phosphatidylinositide phosphatase SAC1 [OS=Homo sapiens] |
| Transitional endoplasmic reticulum ATPase [OS=Homo sapiens] |
| tyrosine-protein kinase Yes [OS=Homo sapiens] |
| Ceruloplasmin [OS=Homo sapiens] |
| EH domain-containing protein 4 [OS=Homo sapiens] |
| Alpha-2-antiplasmin [OS=Homo sapiens] |
| Isoform 7 of Plectin [OS=Homo sapiens] |
| Peroxiredoxin-4 [OS=Homo sapiens] |
| puromycin-sensitive aminopeptidase [OS=Homo sapiens] |
| Delta(3,5)-Delta(2,4)-dienoyl-CoA isomerase, mitochondrial [OS=Homo sapiens] |
| Transmembrane emp24 domain-containing protein 2 [OS=Homo sapiens] |
| Rho GTPase-activating protein 45 [OS=Homo sapiens] |
| Coagulation factor XII [OS=Homo sapiens] |
| Ras-related protein Rab-11B [OS=Homo sapiens] |
| exportin-7 [OS=Homo sapiens] |
| Complement factor B [OS=Homo sapiens] |
| Hydroxyacylglutathione hydrolase, mitochondrial [OS=Homo sapiens] |
| Disintegrin and metalloproteinase domain-containing protein 10 [OS=Homo sapiens] |
| Moesin [OS=Homo sapiens] |
| Dolichyl-diphosphooligosaccharide--protein glycosyltransferase 48 kDa subunit [OS=Homo sapiens] |
| Protein disulfide-isomerase A6 [OS=Homo sapiens] |
| Annexin A7 [OS=Homo sapiens] |
| Isoform 2 of Glucosidase 2 subunit beta [OS=Homo sapiens] |
| Complement C1r subcomponent-like protein [OS=Homo sapiens] |
| Prohibitin [OS=Homo sapiens] |
| stromal interaction molecule 1 [OS=Homo sapiens] |
| Toll-interacting protein [OS=Homo sapiens] |
| Isoform 2 of Glutathione S-transferase Mu 1 [OS=Homo sapiens] |
| Villin-1 [OS=Homo sapiens] |
| Transmembrane 9 superfamily member 3 [OS=Homo sapiens] |
| Glycogen phosphorylase, liver form [OS=Homo sapiens] |
| CD2-associated protein [OS=Homo sapiens] |
| copper chaperone for superoxide dismutase [OS=Homo sapiens] |

| |
|---|
| apolipoprotein M [OS=Homo sapiens] |
| Complement C1q subcomponent subunit C [OS=Homo sapiens] |
| Calponin-2 [OS=Homo sapiens] |
| ATP synthase subunit g, mitochondrial [OS=Homo sapiens] |
| Protein ZGRF1 [OS=Homo sapiens] |
| Hyaluronan-binding protein 2 [OS=Homo sapiens] |
| Isoform 2 of Spectrin beta chain, erythrocytic [OS=Homo sapiens] |
| Dedicator of cytokinesis protein 8 [OS=Homo sapiens] |
| ras-related protein rab-10 [OS=Homo sapiens] |
| annexin A3 [OS=Homo sapiens] |
| Drebrin-like protein [OS=Homo sapiens] |
| osteoclast-stimulating factor 1 [OS=Homo sapiens] |
| guanine nucleotide-binding protein G(i) subunit alpha-2 [OS=Homo sapiens] |
| Heat shock cognate 71 kDa protein [OS=Homo sapiens] |
| Isoform G of Kinesin light chain 1 [OS=Homo sapiens] |
| Cell division control protein 42 homolog [OS=Homo sapiens] |
| Tubulin alpha-4A chain [OS=Homo sapiens] |
| Neurogranin [OS=Homo sapiens] |
| GTP-binding nuclear protein RAN [OS=Homo sapiens] |
| Microtubule-associated protein RP/EB family member 1 [OS=Homo sapiens] |
| Importin subunit beta-1 [OS=Homo sapiens] |
| F-actin-capping protein subunit alpha-2 [OS=Homo sapiens] |
| Serotransferrin [OS=Homo sapiens] |
| CD5 antigen-like [OS=Homo sapiens] |
| protein 4.1 [OS=Homo sapiens] |
| ATP-dependent 6-phosphofructokinase, platelet type [OS=Homo sapiens] |
| Zinc-alpha-2-glycoprotein [OS=Homo sapiens] |
| complement factor H [OS=Homo sapiens] |
| Signal transducer and activator of transcription 5B [OS=Homo sapiens] |
| staphylococcal nuclease domain-containing protein 1 [OS=Homo sapiens] |
| Tight junction protein ZO-2 [OS=Homo sapiens] |
| Complement factor H-related protein 1 [OS=Homo sapiens] |
| Electron transfer flavoprotein subunit alpha, mitochondrial [OS=Homo sapiens] |
| Transmembrane 9 superfamily member 2 [OS=Homo sapiens] |
| Utrophin [OS=Homo sapiens] |
| Afamin [OS=Homo sapiens] |
| Isoform 2 of TSC22 domain family protein 3 [OS=Homo sapiens] |
| glucose-6-phosphate isomerase [OS=Homo sapiens] |
| Alpha-1-acid glycoprotein 1 [OS=Homo sapiens] |
| Isoform 2 of C-X-C chemokine receptor type 4 [OS=Homo sapiens] |
| FH1/FH2 domain-containing protein 1 [OS=Homo sapiens] |
| 55 kDa erythrocyte membrane protein [OS=Homo sapiens] |
| Ras-related protein Rab-14 [OS=Homo sapiens] |
| ADP-ribosylation factor-like protein 8B [OS=Homo sapiens] |
| Carboxypeptidase n subunit 2 [OS=Homo sapiens] |
| Immunoglobulin heavy constant gamma 3 [OS=Homo sapiens] |
| Tripeptidyl-peptidase 2 [OS=Homo sapiens] |

| |
|---|
| Isoform A of Megakaryocyte and platelet inhibitory receptor G6b [OS=Homo sapiens] |
| peroxisomal multifunctional enzyme type 2 [OS=Homo sapiens] |
| Immunoglobulin kappa variable 3-20 [OS=Homo sapiens] |
| Eukaryotic translation initiation factor 5A-1 [OS=Homo sapiens] |
| alpha-mannosidase 2C1 [OS=Homo sapiens] |
| Isoform 2 of Glutathione S-transferase kappa 1 [OS=Homo sapiens] |
| COP9 signalosome complex subunit 4 [OS=Homo sapiens] |
| Actin, cytoplasmic 1 [OS=Homo sapiens] |
| Glia-derived nexin [OS=Homo sapiens] |
| Cofilin-1 [OS=Homo sapiens] |
| Dolichyl-diphosphooligosaccharide--protein glycosyltransferase subunit 1 [OS=Homo sapiens] |
| Trifunctional enzyme subunit beta, mitochondrial [OS=Homo sapiens] |
| Erbin [OS=Homo sapiens] |
| thrombospondin-1 [OS=Homo sapiens] |
| Cytoplasmic dynein 1 heavy chain 1 [OS=Homo sapiens] |
| Rho GTPase-activating protein 6 [OS=Homo sapiens] |
| 60 kDa heat shock protein, mitochondrial [OS=Homo sapiens] |
| Uncharacterized protein C1orf198 [OS=Homo sapiens] |
| Immunoglobulin heavy constant gamma 4 [OS=Homo sapiens] |
| Ubiquitin-like modifier-activating enzyme 1 [OS=Homo sapiens] |
| proteasome subunit beta type-2 [OS=Homo sapiens] |
| Isoform 2 of NADH-cytochrome b5 reductase 3 [OS=Homo sapiens] |
| Neudesin [OS=Homo sapiens] |
| Protein S100-A9 [OS=Homo sapiens] |
| Elongation factor 1-alpha 1 [OS=Homo sapiens] |
| Proteasome subunit alpha type-4 [OS=Homo sapiens] |
| 2',3'-cyclic-nucleotide 3'-phosphodiesterase [OS=Homo sapiens] |
| guanine nucleotide-binding protein G(z) subunit alpha [OS=Homo sapiens] |
| Apolipoprotein A-II [OS=Homo sapiens] |
| Isoform 3 of Ras-related protein Rap-1b [OS=Homo sapiens] |
| Complement C3 [OS=Homo sapiens] |
| Tubulin beta-4B chain [OS=Homo sapiens] |
| WD repeat-containing protein 87 [OS=Homo sapiens] |
| Pyruvate kinase PKM [OS=Homo sapiens] |
| protein furry homolog-like [OS=Homo sapiens] |
| Protein diaphanous homolog 1 [OS=Homo sapiens] |
| Consortin [OS=Homo sapiens] |
| EMILIN-1 [OS=Homo sapiens] |
| Apolipoprotein A-I [OS=Homo sapiens] |
| Acyl-protein thioesterase 1 [OS=Homo sapiens] |
| Isoform 2 of Proteasome activator complex subunit 1 [OS=Homo sapiens] |
| Alpha-2-HS-glycoprotein [OS=Homo sapiens] |
| [F-actin]-monooxygenase MICAL1 [OS=Homo sapiens] |
| Membrane-associated progesterone receptor component 1 [OS=Homo sapiens] |
| Isoform 2 of Apolipoprotein L1 [OS=Homo sapiens] |
| Rho GTPase-activating protein 18 [OS=Homo sapiens] |
| T-complex protein 1 subunit delta [OS=Homo sapiens] |

| |
|---|
| Phosphoglucomutase-2 [OS=Homo sapiens] |
| Isoform 1 of Rho guanine nucleotide exchange factor 7 [OS=Homo sapiens] |
| Isoform 2 of Heat shock protein HSP 90-alpha [OS=Homo sapiens] |
| proliferation-associated protein 2G4 [OS=Homo sapiens] |
| Spectrin beta chain, non-erythrocytic 1 [OS=Homo sapiens] |
| Cytochrome b-c1 complex subunit 1, mitochondrial [OS=Homo sapiens] |
| angiogenin [OS=Homo sapiens] |
| Von Willebrand factor [OS=Homo sapiens] |
| Apolipoprotein C-III [OS=Homo sapiens] |
| Isoform 2 of Platelet glycoprotein Ib beta chain [OS=Homo sapiens] |
| Proline-serine-threonine phosphatase-interacting protein 2 [OS=Homo sapiens] |
| Calpain small subunit 1 [OS=Homo sapiens] |
| Unconventional myosin-Va [OS=Homo sapiens] |
| Complement component 1 Q subcomponent-binding protein, mitochondrial [OS=Homo sapiens] |
| Cytosolic 10-formyltetrahydrofolate dehydrogenase [OS=Homo sapiens] |
| Prothrombin [OS=Homo sapiens] |
| Ras-related C3 botulinum toxin substrate 1 [OS=Homo sapiens] |
| Proteasome subunit alpha type-6 [OS=Homo sapiens] |
| Isoform 2 of Stress-induced-phosphoprotein 1 [OS=Homo sapiens] |
| ADP-ribosylation factor 4 [OS=Homo sapiens] |
| Aconitate hydratase, mitochondrial [OS=Homo sapiens] |
| Sorting nexin-2 [OS=Homo sapiens] |
| Plasma kallikrein [OS=Homo sapiens] |
| GAS2-like protein 1 [OS=Homo sapiens] |
| Dehydrogenase/reductase SDR family member 7 [OS=Homo sapiens] |
| Succinate--CoA ligase [ADP-forming] subunit beta, mitochondrial [OS=Homo sapiens] |
| bifunctional 3'-phosphoadenosine 5'-phosphosulfate synthase 1 [OS=Homo sapiens] |
| Isoform 7 of Protein MRVI1 [OS=Homo sapiens] |
| Thioredoxin-dependent peroxide reductase, mitochondrial [OS=Homo sapiens] |
| sorting nexin-3 [OS=Homo sapiens] |
| Thromboxane-A synthase [OS=Homo sapiens] |
| Arachidonate 12-lipoxygenase, 12S-type [OS=Homo sapiens] |
| endonuclease domain-containing 1 protein [OS=Homo sapiens] |
| Myc target protein 1 [OS=Homo sapiens] |
| Junctional adhesion molecule C [OS=Homo sapiens] |
| (E3-independent) E2 ubiquitin-conjugating enzyme [OS=Homo sapiens] |
| Septin-5 [OS=Homo sapiens] |
| Vasodilator-stimulated phosphoprotein [OS=Homo sapiens] |
| High affinity immunoglobulin epsilon receptor subunit gamma [OS=Homo sapiens] |
| Basigin [OS=Homo sapiens] |
| 14-3-3 protein beta/alpha [OS=Homo sapiens] |
| Apolipoprotein A-V [OS=Homo sapiens] |
| Trifunctional enzyme subunit alpha, mitochondrial [OS=Homo sapiens] |
| Isoform 2 of F-actin-capping protein subunit beta [OS=Homo sapiens] |
| Ubiquitin carboxyl-terminal hydrolase 5 [OS=Homo sapiens] |
| Serum paraoxonase/arylesterase 1 [OS=Homo sapiens] |
| Surfeit locus protein 6 [OS=Homo sapiens] |

| |
|---|
| Isoform 2 of General vesicular transport factor p115 [OS=Homo sapiens] |
| Leukocyte surface antigen CD47 [OS=Homo sapiens] |
| Carnitine O-palmitoyltransferase 1, liver isoform [OS=Homo sapiens] |
| e3 ubiquitin-protein ligase UBR4 [OS=Homo sapiens] |
| Serpin B9 [OS=Homo sapiens] |
| Platelet endothelial cell adhesion molecule [OS=Homo sapiens] |
| Secreted phosphoprotein 24 [OS=Homo sapiens] |
| kininogen-1 [OS=Homo sapiens] |
| Coatomer subunit gamma-1 [OS=Homo sapiens] |
| heat shock 70 kDa protein 1A [OS=Homo sapiens] |
| Alpha-1-antichymotrypsin [OS=Homo sapiens] |
| Poly(RC)-binding protein 1 [OS=Homo sapiens] |
| HLA class I histocompatibility antigen, A-2 alpha chain [OS=Homo sapiens] |
| 3-hydroxyacyl-CoA dehydrogenase type-2 [OS=Homo sapiens] |
| Putative cytochrome b-c1 complex subunit Rieske-like protein 1 [OS=Homo sapiens] |
| Isoform 4 of Latent-transforming growth factor beta-binding protein 1 [OS=Homo sapiens] |
| Isoform 2 of Collagen alpha-3(VI) chain [OS=Homo sapiens] |
| Hexokinase-1 [OS=Homo sapiens] |
| cGMP-specific 3',5'-cyclic phosphodiesterase [OS=Homo sapiens] |
| Interleukin-1 receptor-associated kinase 4 [OS=Homo sapiens] |
| ADP-ribosylation factor 1 [OS=Homo sapiens] |
| Band 3 anion transport protein [OS=Homo sapiens] |
| Isoform 4 of Nuclear factor 1 B-type [OS=Homo sapiens] |
| Actin-related protein 2/3 complex subunit 2 [OS=Homo sapiens] |
| Isoform 3 of Drebrin [OS=Homo sapiens] |
| Prostaglandin G/H synthase 1 [OS=Homo sapiens] |
| protein flightless-1 homolog [OS=Homo sapiens] |
| Cytoplasmic protein NCK2 [OS=Homo sapiens] |
| L-lactate dehydrogenase B chain [OS=Homo sapiens] |
| 3-mercaptopyruvate sulfurtransferase [OS=Homo sapiens] |
| Cathepsin D [OS=Homo sapiens] |
| Eukaryotic peptide chain release factor GTP-binding subunit ERF3A [OS=Homo sapiens] |
| Apolipoprotein C-II [OS=Homo sapiens] |
| Platelet glycoprotein Ib alpha chain [OS=Homo sapiens] |
| Complement C4-B [OS=Homo sapiens] |
| phosphoglycerate kinase 1 [OS=Homo sapiens] |
| Complement component C8 alpha chain [OS=Homo sapiens] |
| Syntaxin-binding protein 2 [OS=Homo sapiens] |
| purine nucleoside phosphorylase [OS=Homo sapiens] |
| 6-phosphogluconate dehydrogenase, decarboxylating [OS=Homo sapiens] |
| Ras-related protein Rab-6A [OS=Homo sapiens] |
| Isoform 1 of Voltage-dependent anion-selective channel protein 2 [OS=Homo sapiens] |
| Isoform 8 of Tropomyosin alpha-1 chain [OS=Homo sapiens] |
| Transgelin-2 [OS=Homo sapiens] |
| Crk-like protein [OS=Homo sapiens] |
| Glutathione peroxidase 3 [OS=Homo sapiens] |
| profilin-1 [OS=Homo sapiens] |

| |
|---|
| T-complex protein 1 subunit theta [OS=Homo sapiens] |
| Actin-related protein 2/3 complex subunit 1B [OS=Homo sapiens] |
| P-selectin [OS=Homo sapiens] |
| Src substrate cortactin [OS=Homo sapiens] |
| Phosphate carrier protein, mitochondrial [OS=Homo sapiens] |
| F-actin-capping protein subunit alpha-1 [OS=Homo sapiens] |
| Serine/threonine-protein kinase 24 [OS=Homo sapiens] |
| Isoform 2 of Ras-related protein Ral-B [OS=Homo sapiens] |
| Scavenger receptor class F member 1 [OS=Homo sapiens] |
| Hemoglobin subunit beta [OS=Homo sapiens] |
| protein-L-isoaspartate(D-aspartate) O-methyltransferase [OS=Homo sapiens] |
| SCY1-like protein 2 [OS=Homo sapiens] |
| Proto-oncogene tyrosine-protein kinase Src [OS=Homo sapiens] |
| immunoglobulin heavy constant mu [OS=Homo sapiens] |
| tropomodulin-3 [OS=Homo sapiens] |
| alpha-enolase [OS=Homo sapiens] |
| long-chain-fatty-acid--CoA ligase 3 [OS=Homo sapiens] |
| Apolipoprotein F [OS=Homo sapiens] |
| 78 kDa glucose-regulated protein [OS=Homo sapiens] |
| AMP deaminase 2 [OS=Homo sapiens] |
| Talin-1 [OS=Homo sapiens] |
| Actin-related protein 2/3 complex subunit 5 [OS=Homo sapiens] |
| Isoform 2 of Medium-chain specific acyl-CoA dehydrogenase, mitochondrial [OS=Homo sapiens] |
| Isoform 2 of Tropomyosin alpha-3 chain [OS=Homo sapiens] |
| Actin-related protein 2 [OS=Homo sapiens] |
| Isoform 2 of Tyrosine-protein phosphatase non-receptor type 6 [OS=Homo sapiens] |
| Isoform 3 of Clathrin interactor 1 [OS=Homo sapiens] |
| Isoform 2 of Lipopolysaccharide-responsive and beige-like anchor protein [OS=Homo sapiens] |
| coactosin-like protein [OS=Homo sapiens] |
| unconventional myosin-Ig [OS=Homo sapiens] |
| Plasminogen receptor (KT) [OS=Homo sapiens] |
| WD repeat-containing protein 44 [OS=Homo sapiens] |
| Isoform 3 of Ubiquitin carboxyl-terminal hydrolase MINDY-1 [OS=Homo sapiens] |
| Isoform Alpha-6X1X2A of Integrin alpha-6 [OS=Homo sapiens] |
| Coagulation factor V [OS=Homo sapiens] |
| LIM and senescent cell antigen-like-containing domain protein 1 [OS=Homo sapiens] |
| Septin-7 [OS=Homo sapiens] |
| phosphatidate cytidylyltransferase 2 [OS=Homo sapiens] |
| Na(+)/H(+) exchange regulatory cofactor NHE-RF1 [OS=Homo sapiens] |
| Fibrinogen beta chain [OS=Homo sapiens] |
| vesicle transport through interaction with t-SNAREs homolog 1B [OS=Homo sapiens] |
| Hsc70-interacting protein [OS=Homo sapiens] |
| Eukaryotic translation initiation factor 4E [OS=Homo sapiens] |
| Serine/threonine-protein kinase Nek9 [OS=Homo sapiens] |
| Thioredoxin-related transmembrane protein 1 [OS=Homo sapiens] |
| Plasminogen [OS=Homo sapiens] |
| Ubiquitin-60S ribosomal protein L40 [OS=Homo sapiens] |

| |
|---|
| Endoplasmin [OS=Homo sapiens] |
| Isoform 4 of Caldesmon [OS=Homo sapiens] |
| peroxiredoxin-1 [OS=Homo sapiens] |
| DCC-interacting protein 13-beta [OS=Homo sapiens] |
| GTP:AMP phosphotransferase AK3, mitochondrial [OS=Homo sapiens] |
| Isoform 8 of Fibronectin [OS=Homo sapiens] |
| rho GTPase-activating protein 1 [OS=Homo sapiens] |
| C4b-binding protein alpha chain [OS=Homo sapiens] |
| Rab GDP dissociation inhibitor beta [OS=Homo sapiens] |
| Complement C1q subcomponent subunit A [OS=Homo sapiens] |
| ATP synthase subunit beta, mitochondrial [OS=Homo sapiens] |
| Extended synaptotagmin-1 [OS=Homo sapiens] |
| Fibrinogen alpha chain [OS=Homo sapiens] |
| Apolipoprotein A-IV [OS=Homo sapiens] |
| caveolae-associated protein 2 [OS=Homo sapiens] |
| Choline transporter-like protein 2 [OS=Homo sapiens] |
| immunoglobulin heavy variable 3-73 [OS=Homo sapiens] |
| Receptor-type tyrosine-protein phosphatase eta [OS=Homo sapiens] |
| Growth factor receptor-bound protein 2 [OS=Homo sapiens] |
| actin-related protein 3 [OS=Homo sapiens] |
| Beta-arrestin-1 [OS=Homo sapiens] |
| HLA class I histocompatibility antigen, A-24 alpha chain [OS=Homo sapiens] |
| Ras-related protein Rab-13 [OS=Homo sapiens] |
| Inter-alpha-trypsin inhibitor heavy chain H4 [OS=Homo sapiens] |
| Isoform 2 of Integrin alpha-IIb [OS=Homo sapiens] |
| Immunoglobulin J chain [OS=Homo sapiens] |
| NAD(P) transhydrogenase, mitochondrial [OS=Homo sapiens] |
| spectrin alpha chain, erythrocytic 1 [OS=Homo sapiens] |
| Thioredoxin-like protein 1 [OS=Homo sapiens] |
| Heparin cofactor 2 [OS=Homo sapiens] |
| glyceraldehyde-3-phosphate dehydrogenase [OS=Homo sapiens] |
| Twinfilin-2 [OS=Homo sapiens] |
| Beta-parvin [OS=Homo sapiens] |
| PDZ and LIM domain protein 7 [OS=Homo sapiens] |
| Leukocyte elastase inhibitor [OS=Homo sapiens] |
| Phosphoglucomutase-1 [OS=Homo sapiens] |
| Ribonuclease inhibitor [OS=Homo sapiens] |
| Protein disulfide-isomerase [OS=Homo sapiens] |
| Beta-2-glycoprotein 1 [OS=Homo sapiens] |
| aflatoxin B1 aldehyde reductase member 2 [OS=Homo sapiens] |
| EH domain-containing protein 3 [OS=Homo sapiens] |
| aminopeptidase N [OS=Homo sapiens] |
| Isoform 12 of Titin [OS=Homo sapiens] |
| Arf-GAP with SH3 domain, ANK repeat and PH domain-containing protein 2 [OS=Homo sapiens] |
| Serum albumin [OS=Homo sapiens] |
| Stress-70 protein, mitochondrial [OS=Homo sapiens] |
| Histidine triad nucleotide-binding protein 1 [OS=Homo sapiens] |

| |
|---|
| Immunoglobulin heavy constant gamma 2 [OS=Homo sapiens] |
| WD repeat-containing protein 1 [OS=Homo sapiens] |
| Guanine nucleotide-binding protein G(Q) subunit alpha [OS=Homo sapiens] |
| apolipoprotein B-100 [OS=Homo sapiens] |
| Ras GTPase-activating protein 3 [OS=Homo sapiens] |
| fructose-bisphosphate aldolase A [OS=Homo sapiens] |
| Isoform 1 of Vinculin [OS=Homo sapiens] |
| thymidine phosphorylase [OS=Homo sapiens] |
| Fibrinogen gamma chain [OS=Homo sapiens] |
| Microtubule-actin cross-linking factor 1, isoforms 1/2/3/5 [OS=Homo sapiens] |
| 14-3-3 protein theta [OS=Homo sapiens] |
| ATP synthase subunit alpha, mitochondrial [OS=Homo sapiens] |
| Beta-1-syntrophin [OS=Homo sapiens] |
| Tropomyosin alpha-4 chain [OS=Homo sapiens] |
| 14-3-3 protein zeta/delta [OS=Homo sapiens] |
| tubulin beta-1 chain [OS=Homo sapiens] |
| 14-3-3 protein eta [OS=Homo sapiens] |
| apolipoprotein(a) [OS=Homo sapiens] |
| Integrin beta-3 [OS=Homo sapiens] |
| Isoform 3 of CAP-Gly domain-containing linker protein 1 [OS=Homo sapiens] |
| Myotrophin [OS=Homo sapiens] |
| Zyxin [OS=Homo sapiens] |
| alpha-2-macroglobulin [OS=Homo sapiens] |
| Isoform 2 of Triosephosphate isomerase [OS=Homo sapiens] |
| ATP synthase-coupling factor 6, mitochondrial [OS=Homo sapiens] |
| Sarcoplasmic/endoplasmic reticulum calcium atpase 3 [OS=Homo sapiens] |
| Thioredoxin domain-containing protein 5 [OS=Homo sapiens] |
| erythrocyte band 7 integral membrane protein [OS=Homo sapiens] |
| Isoform 3 of FYN-binding protein 1 [OS=Homo sapiens] |
| serine/threonine-protein phosphatase 2A 65 kDa regulatory subunit A alpha isoform [OS=Homo sapiens] |
| Protein kinase C and casein kinase substrate in neurons protein 2 [OS=Homo sapiens] |
| cytosol aminopeptidase [OS=Homo sapiens] |
| Dihydropyrimidinase-related protein 2 [OS=Homo sapiens] |
| Delta-aminolevulinic acid dehydratase [OS=Homo sapiens] |
| Apolipoprotein C-I [OS=Homo sapiens] |
| Protein disulfide-isomerase A3 [OS=Homo sapiens] |
| Calnexin [OS=Homo sapiens] |
| Peroxiredoxin-6 [OS=Homo sapiens] |
| Protein phosphatase 1F [OS=Homo sapiens] |
| Multimerin-1 [OS=Homo sapiens] |
| Ras-related protein Rab-6B [OS=Homo sapiens] |
| Rho-associated protein kinase 2 [OS=Homo sapiens] |
| ATPase family AAA domain-containing protein 3B [OS=Homo sapiens] |
| Myosin-9 [OS=Homo sapiens] |
| Isoform 2 of Filamin-A [OS=Homo sapiens] |
| Isoform 2 of Clathrin heavy chain 1 [OS=Homo sapiens] |
| protein/nucleic acid deglycase DJ-1 [OS=Homo sapiens] |

| |
|---|
| Inter-alpha-trypsin inhibitor heavy chain H1 [OS=Homo sapiens] |
| Vitronectin [OS=Homo sapiens] |
| Vitamin K-dependent protein S [OS=Homo sapiens] |
| Alpha-actinin-1 [OS=Homo sapiens] |

ndrial [OS=Homo sapiens]

chondrial [OS=Homo sapiens]
