## Supplementary Table 4 for "A Large-Scale Proteomics Resource of Circulating Extracellular Vesicles for Biomarker Discovery in Pancreatic Cancer"

| **Supplementary Table 4: List of EV proteins that met the eligibility criteria for principal component analysis.** | | | | | | | | |
| --- | --- | --- | --- | --- | --- | --- | --- | --- |
| **ProteinID** | **GeneName** | **EntrezID** | **HCvsPDAC log2Fold Change** | **HCvsPDAC pValue** | **HCvsCP log2Fold Change** | **HCvsCP pValue** | **HCvsIPMN log2Fold Change** | **HCvsIPMN pValue** |
| P07148 | FABP1 | 2168 | 20.374 | 2.25E-06 | 21.833 | 6.75E-05 | 23.836 | 6.38E-05 |
| Q8WUM4 | PDCD6IP | 10015 | 20.027 | 5.19E-07 | 4.731 | 1.20E-01 | 10.449 | 1.87E-02 |
| Q8IW75 | SERPINA12 | 145264 | 14.806 | 1.27E-04 | -0.072 | 3.38E-01 | -0.072 | 4.56E-01 |
| Q9P0V3-1 | SH3BP4 | 23677 | 13.933 | 1.60E-04 | 0.000 | NA | 0.000 | NA |
| P61764 | STXBP1 | 6812 | 13.720 | 2.46E-05 | 9.304 | 1.07E-02 | 7.163 | 8.81E-02 |
| P05165-1 | PCCA | 5095 | 13.074 | 1.06E-04 | 12.018 | 1.64E-03 | 2.479 | 2.86E-01 |
| Q9BSJ8 | ESYT1 | 23344 | 12.627 | 5.16E-04 | 10.171 | 9.62E-03 | 7.601 | 3.75E-02 |
| Q01628 | IFITM3 | 10410 | 12.505 | 2.04E-04 | 8.382 | 2.26E-02 | 4.964 | 2.24E-01 |
| O15162 | PLSCR1 | 5359 | 12.432 | 2.40E-04 | 4.864 | 9.26E-02 | 0.000 | NA |
| Q5JXB2 | UBE2NL | 389898 | 11.964 | 3.21E-04 | -0.061 | 1.00E+00 | 4.026 | 2.24E-01 |
| P48509 | CD151 | 977 | 11.863 | 4.28E-04 | 13.420 | 1.64E-03 | 14.600 | 1.14E-03 |
| Q99808 | SLC29A1 | 2030 | 11.809 | 2.12E-04 | 10.574 | 5.60E-03 | 10.213 | 3.59E-02 |
| P63208 | SKP1 | 6500 | 11.788 | 7.43E-04 | 0.000 | NA | 5.499 | 1.04E-01 |
| P20810-1 | CAST | 831 | 11.283 | 1.52E-05 | 5.983 | 2.67E-02 | 5.066 | 1.33E-01 |
| Q5TDH0-3 | DDI2 | 84301 | 11.258 | 5.16E-04 | 3.166 | 1.86E-01 | 0.000 | NA |
| P24539 | ATP5F1 | NA | 11.154 | 9.30E-04 | 8.053 | 3.02E-02 | -3.506 | 2.45E-01 |
| P08473 | MME | 4311 | 11.118 | 6.99E-04 | 9.355 | 1.07E-02 | 11.289 | 1.07E-02 |
| Q92820 | GGH | 8836 | 11.114 | 1.76E-03 | 0.000 | NA | 0.000 | NA |
| P54920 | NAPA | 8775 | 10.734 | 5.29E-04 | 4.520 | 2.73E-01 | 7.443 | 1.66E-01 |
| O15400-2 | STX7 | 8417 | 10.636 | 1.00E-03 | 1.484 | 6.36E-01 | 3.185 | 3.98E-01 |
| Q8NBP7-1 | PCSK9 | 255738 | 10.338 | 2.38E-04 | 4.890 | 1.42E-01 | 4.510 | 2.33E-01 |
| P61163 | ACTR1A | 10121 | 10.101 | 4.09E-03 | 3.199 | 5.63E-01 | 9.540 | 3.78E-02 |
| Q7Z5J8-1 | ANKAR | 150709 | 9.819 | 5.16E-04 | 0.722 | 3.84E-01 | 0.000 | NA |
| Q13576-1 | IQGAP2 | 10788 | 9.612 | 1.63E-03 | 8.893 | 5.76E-03 | 2.426 | 3.98E-01 |
| O95466-3 | FMNL1 | 752 | 9.603 | 2.43E-03 | 6.798 | 2.14E-02 | 6.029 | 3.75E-02 |
| Q9C0C9 | UBE2O | 63893 | 9.601 | 4.72E-03 | 3.518 | 2.21E-01 | 2.719 | 5.15E-01 |
| Q15555 | MAPRE2 | 10982 | 9.556 | 2.43E-03 | 4.753 | 9.26E-02 | 2.467 | 2.86E-01 |
| A0A0C4DH24 | IGKV6-21 | 28906 | 9.447 | 5.94E-03 | 8.933 | 4.68E-02 | 4.597 | 2.73E-01 |
| P23276 | KEL | 3792 | 9.413 | 2.28E-03 | 4.666 | 5.41E-02 | 12.156 | 6.55E-03 |
| P48426 | PIP4K2A | 5305 | 9.334 | 2.07E-03 | 4.990 | 4.53E-02 | 0.000 | NA |
| P05186-1 | ALPL | 249 | 9.281 | 2.43E-03 | 11.631 | 4.10E-03 | 12.746 | 4.03E-03 |
| P51149 | RAB7A | 7879 | 9.214 | 7.05E-04 | 6.844 | 6.12E-02 | 8.442 | 3.99E-02 |
| A0A075B6K0 | IGLV3-16 | 28799 | 9.173 | 4.52E-03 | 12.801 | 1.64E-03 | 0.000 | NA |
| P10619 | CTSA | 5476 | 9.053 | 1.05E-02 | -3.170 | 4.75E-01 | 6.127 | 2.84E-01 |
| P17858-2 | PFKL | 5211 | 8.844 | 3.19E-03 | -1.637 | 3.38E-01 | 0.225 | 4.75E-01 |
| P16615-4 | ATP2A2 | 488 | 8.347 | 3.55E-03 | 2.664 | 5.50E-01 | 6.556 | 1.27E-01 |
| P63218 | GNG5 | 2787 | 8.301 | 1.82E-04 | 8.739 | 7.72E-04 | 8.400 | 1.14E-02 |
| Q9ULV4-3 | CORO1C | 23603 | 7.824 | 1.09E-02 | 14.685 | 1.60E-04 | 7.394 | 7.99E-02 |
| O75955 | FLOT1 | 10211 | 7.779 | 3.49E-07 | 8.129 | 5.92E-06 | 4.161 | 9.08E-02 |
| P09622 | DLD | 1738 | 7.431 | 6.21E-03 | -4.926 | 6.59E-02 | -4.926 | 1.35E-01 |
| P49862 | KLK7 | 5650 | 6.732 | 2.33E-02 | 13.521 | 6.14E-04 | 5.119 | 1.04E-01 |
| Q14974 | KPNB1 | 3837 | 6.625 | 3.25E-03 | 4.850 | 1.93E-01 | 2.505 | 1.89E-01 |
| P31025 | LCN1 | 3933 | 6.550 | 2.85E-03 | 3.821 | 4.53E-02 | 2.741 | 1.04E-01 |
| P27824 | CANX | 821 | 6.159 | 4.60E-06 | 6.039 | 7.91E-04 | -0.144 | 5.35E-01 |
| P29350-1 | PTPN6 | 5777 | 6.079 | 4.46E-02 | 12.509 | 2.60E-03 | -9.183 | 3.78E-02 |
| P62879 | GNB2 | 2783 | 6.052 | 1.38E-01 | 11.169 | 2.12E-03 | 2.969 | 8.96E-01 |
| P27487 | DPP4 | 1803 | 5.898 | 9.28E-03 | 13.016 | 6.72E-05 | 3.221 | 3.75E-02 |
| P11234-2 | RALB | 5899 | 5.785 | 1.19E-06 | 4.007 | 9.05E-02 | 3.700 | 3.86E-01 |
| O60832-1 | DKC1 | 1736 | 5.777 | 7.69E-04 | -3.416 | 6.19E-01 | -2.030 | 3.82E-01 |
| Q08830 | FGL1 | 2267 | 5.492 | 5.22E-03 | 0.680 | 4.17E-01 | 6.800 | 5.19E-02 |
| Q9NZD2 | GLTP | 51228 | 5.117 | 4.89E-02 | 13.063 | 6.20E-04 | -2.924 | 2.45E-01 |
| Q15833-1 | STXBP2 | 6813 | 4.636 | 9.44E-02 | 12.232 | 1.99E-03 | 3.035 | 3.98E-01 |
| P25786-1 | PSMA1 | 5682 | 4.431 | 4.58E-04 | -0.923 | 9.75E-01 | 3.366 | 7.10E-01 |
| P13716-1 | ALAD | 210 | 4.163 | 6.59E-07 | 2.405 | 6.98E-03 | 1.829 | 6.92E-02 |
| O94919 | ENDOD1 | 23052 | 4.158 | 1.12E-04 | 4.216 | 1.69E-03 | 2.965 | 2.72E-01 |
| P06576 | ATP5B | NA | 4.015 | 7.14E-03 | 5.187 | 2.54E-03 | 4.844 | 2.57E-02 |
| Q07617 | SPAG1 | 6674 | 3.980 | 8.30E-02 | 0.000 | NA | 22.885 | 6.32E-05 |
| P11166 | SLC2A1 | 6513 | 3.715 | 2.83E-06 | 3.650 | 2.08E-03 | 3.044 | 1.57E-02 |
| P01701 | IGLV1-51 | 28820 | 3.304 | 1.91E-04 | 2.687 | 1.85E-01 | 2.461 | 5.63E-01 |
| Q14847-2 | LASP1 | 3927 | 3.271 | 4.96E-04 | -3.825 | 2.17E-01 | -4.506 | 1.86E-01 |
| Q02985-1 | CFHR3 | 10878 | 3.089 | 6.90E-05 | 2.441 | 1.09E-01 | 1.855 | 8.40E-01 |
| P05164-2 | MPO | 4353 | 3.049 | 3.75E-05 | 0.954 | 7.93E-02 | -0.698 | 8.36E-01 |
| P50991 | CCT4 | 10575 | 2.735 | 8.37E-04 | -5.942 | 4.44E-02 | -0.698 | 6.80E-01 |
| P50990 | CCT8 | 10694 | 2.657 | 1.50E-05 | -0.705 | 9.28E-01 | 0.766 | 4.42E-01 |
| Q9NP58-4 | ABCB6 | 10058 | 2.494 | 1.27E-01 | 9.980 | 2.12E-04 | 2.253 | 2.86E-01 |
| P04792 | HSPB1 | 3315 | 2.463 | 9.91E-03 | 2.046 | 3.47E-01 | 1.796 | 5.35E-01 |
| Q14254 | FLOT2 | 2319 | 2.349 | 2.19E-06 | 2.301 | 4.01E-04 | 2.026 | 4.97E-03 |
| P02724 | GYPA | 2993 | 2.342 | 6.19E-03 | 2.755 | 2.68E-02 | 0.230 | 4.92E-01 |
| Q8NF91-7 | SYNE1 | 23345 | 2.339 | 1.04E-05 | -0.596 | 1.57E-01 | -0.377 | 1.29E-01 |
| P61981 | YWHAG | 7532 | 2.049 | 1.09E-02 | 0.630 | 2.68E-01 | -3.202 | 2.85E-02 |
| P22352 | GPX3 | 2878 | 2.038 | 3.77E-03 | 1.692 | 5.95E-02 | -4.156 | 6.92E-02 |
| P61006 | RAB8A | 4218 | 1.897 | 6.87E-06 | 1.581 | 5.46E-04 | 1.203 | 3.52E-02 |
| Q09666-1 | AHNAK | 79026 | 1.841 | 1.55E-06 | 0.396 | 1.57E-01 | 0.864 | 4.09E-02 |
| Q9HDC9 | APMAP | 57136 | 1.740 | 1.04E-06 | 1.135 | 7.93E-04 | 1.231 | 4.37E-03 |
| Q5VST9-6 | OBSCN | 84033 | 1.699 | 5.42E-07 | 0.781 | 2.20E-04 | 0.056 | 8.36E-01 |
| Q03164 | KMT2A | 4297 | 1.676 | 2.03E-04 | -0.115 | 9.51E-01 | -4.906 | 6.50E-01 |
| P69905 | HBA2 | 3040 | 1.609 | 1.77E-05 | 1.165 | 2.66E-02 | 0.792 | 2.84E-02 |
| P06733-1 | ENO1 | 2023 | 1.597 | 7.97E-06 | 1.026 | 6.15E-03 | 0.637 | 5.22E-02 |
| P28072 | PSMB6 | 5694 | 1.590 | 7.85E-03 | -0.416 | 8.80E-01 | 1.986 | 4.09E-02 |
| P0DJI9 | SAA2 | 6289 | 1.585 | 2.20E-04 | -0.303 | 9.75E-01 | 0.177 | 2.00E-01 |
| P04899-1 | GNAI2 | 2771 | 1.395 | 1.26E-06 | 0.620 | 1.26E-02 | 0.161 | 3.86E-01 |
| Q02413 | DSG1 | 1828 | 1.337 | 8.05E-04 | 0.484 | 9.04E-02 | 0.238 | 4.33E-01 |
| Q9BZQ6 | EDEM3 | 80267 | 1.318 | 7.55E-01 | 12.710 | 4.21E-04 | 8.424 | 2.48E-02 |
| P05107 | ITGB2 | 3689 | 1.306 | 2.66E-04 | -0.408 | 6.51E-01 | -0.181 | 7.17E-01 |
| P48735 | IDH2 | 3418 | 1.294 | 2.50E-03 | -0.696 | 6.76E-03 | -0.314 | 8.28E-02 |
| A0A0B4J1X5 | IGHV3-74 | 28408 | 1.227 | 1.63E-03 | 0.245 | 4.49E-01 | -1.126 | 2.59E-02 |
| Q13418 | ILK | 3611 | 1.208 | 1.90E-04 | 1.447 | 1.04E-05 | 0.651 | 6.20E-02 |
| P01031 | C5 | 727 | 1.181 | 9.97E-05 | 0.071 | 4.78E-01 | 0.027 | 6.79E-01 |
| Q8IZT6-1 | ASPM | 259266 | 1.155 | 3.78E-03 | 1.850 | 7.14E-05 | 0.042 | 8.36E-01 |
| P05556-5 | ITGB1 | 3688 | 1.153 | 7.75E-04 | 0.330 | 6.51E-01 | 0.288 | 7.72E-01 |
| P05556-1 | ITGB1 | 3688 | 1.153 | 7.75E-04 | 0.330 | 6.51E-01 | 0.288 | 7.72E-01 |
| P67936 | TPM4 | 7171 | 1.119 | 3.61E-04 | 1.114 | 1.88E-03 | 0.579 | 2.06E-01 |
| P67936-2 | TPM4 | 7171 | 1.119 | 3.61E-04 | 1.114 | 1.88E-03 | 0.579 | 2.06E-01 |
| Q8IY51 | TIGD4 | 201798 | 1.096 | 3.53E-04 | 0.248 | 2.67E-01 | 0.364 | 2.82E-01 |
| P29375-1 | KDM5A | 5927 | 1.071 | 3.35E-03 | 0.899 | 6.46E-02 | 1.194 | 2.57E-03 |
| Q12789-2 | GTF3C1 | 2975 | 1.062 | 4.96E-04 | -0.280 | 1.48E-01 | 1.501 | 3.26E-04 |
| P19013 | KRT4 | 3851 | 1.035 | 2.88E-04 | -0.395 | 5.58E-02 | 0.085 | 8.04E-01 |
| P15144 | ANPEP | 290 | 1.020 | 1.76E-03 | 0.663 | 7.93E-02 | 0.614 | 1.37E-01 |
| Q8NGP6 | OR5M8 | 219484 | 0.976 | 3.20E-01 | 16.238 | 6.75E-05 | 12.228 | 1.14E-03 |
| P61160-1 | ACTR2 | 10097 | 0.911 | 2.85E-01 | 6.555 | 9.78E-04 | 4.243 | 4.32E-01 |
| P02549-1 | SPTA1 | 6708 | 0.902 | 1.26E-05 | 1.089 | 6.18E-04 | 0.686 | 3.12E-02 |
| Q9NQC3 | RTN4 | 57142 | 0.609 | 3.91E-03 | 1.086 | 2.81E-04 | 1.014 | 5.29E-05 |
| P02775 | PPBP | 5473 | 0.567 | 3.45E-02 | 2.530 | 4.50E-04 | 1.201 | 2.15E-01 |
| P11171-2 | EPB41 | 2035 | 0.540 | 6.51E-02 | 1.171 | 2.08E-03 | 1.505 | 3.18E-04 |
| P11171-4 | EPB41 | 2035 | 0.540 | 6.51E-02 | 1.171 | 2.08E-03 | 1.505 | 3.18E-04 |
| P61026 | RAB10 | 10890 | 0.417 | 1.01E-02 | 1.152 | 8.84E-04 | 0.811 | 3.52E-02 |
| A0A0B4J1V0 | IGHV3-15 | 28448 | 0.254 | 4.19E-01 | 1.158 | 1.37E-03 | -0.355 | 2.83E-01 |
| O00468-5 | AGRN | 375790 | 0.022 | 4.47E-01 | 1.567 | 1.43E-04 | -0.677 | 3.51E-02 |
| P13646 | KRT13 | 3860 | -0.148 | 5.36E-01 | -1.052 | 1.43E-04 | -0.021 | 1.00E+00 |
| P13987 | CD59 | 966 | -0.524 | 8.12E-01 | 1.224 | 1.23E-03 | 0.367 | 1.16E-01 |
| P14625 | HSP90B1 | 7184 | -0.992 | 1.27E-04 | -1.431 | 1.53E-04 | -1.223 | 7.94E-04 |
| P00742 | F10 | 2159 | -1.017 | 3.24E-03 | -0.396 | 1.09E-01 | -0.725 | 2.54E-03 |
| Q08554-2 | DSC1 | 1823 | -1.043 | 1.57E-05 | -0.293 | 2.42E-01 | -1.810 | 3.26E-04 |
| O60281-1 | ZNF292 | 23036 | -1.059 | 3.98E-04 | -1.272 | 5.52E-05 | 0.318 | 8.26E-02 |
| P06331 | IGHV4-34 | 28395 | -1.068 | 8.49E-03 | -0.281 | 3.25E-01 | -0.271 | 5.35E-01 |
| P98160 | HSPG2 | 3339 | -1.130 | 1.30E-04 | -0.032 | 9.02E-01 | -0.325 | 2.30E-02 |
| P00338-1 | LDHA | 3939 | -1.135 | 1.09E-03 | -0.498 | 7.41E-02 | -0.136 | 5.63E-01 |
| P78371-1 | CCT2 | 10576 | -1.198 | 1.85E-03 | -0.110 | 7.35E-01 | -0.416 | 2.38E-01 |
| Q92743 | HTRA1 | 5654 | -1.207 | 5.52E-03 | -0.467 | 7.82E-01 | -0.590 | 8.36E-01 |
| O75083 | WDR1 | 9948 | -1.219 | 1.07E-02 | -0.208 | 5.59E-01 | 0.081 | 4.92E-01 |
| Q8WUA8 | TSKU | 25987 | -1.237 | 1.34E-03 | -0.437 | 4.06E-01 | 0.111 | 6.79E-01 |
| Q08431 | MFGE8 | 4240 | -1.270 | 2.00E-02 | -1.382 | 6.66E-05 | -0.865 | 2.24E-03 |
| A0A075B6S2 | IGKV2D-29 | 28882 | -1.274 | 8.23E-03 | 0.250 | 3.47E-01 | -0.422 | 2.38E-01 |
| Q5T7N2 | L1TD1 | 54596 | -1.298 | 8.82E-01 | -15.042 | 5.32E-05 | -0.768 | 1.37E-01 |
| P02741-1 | CRP | 1401 | -1.305 | 6.43E-02 | -4.245 | 1.23E-03 | -4.239 | 1.95E-03 |
| Q13200 | PSMD2 | 5708 | -1.311 | 3.46E-04 | -0.528 | 1.66E-01 | 0.109 | 8.36E-01 |
| P48594 | SERPINB4 | 6318 | -1.354 | 3.05E-05 | -1.281 | 1.69E-03 | -1.313 | 5.03E-04 |
| Q9NQ36-1 | SCUBE2 | 57758 | -1.357 | 3.19E-03 | -0.200 | 5.18E-01 | -0.578 | 2.57E-02 |
| P27918 | CFP | 5199 | -1.400 | 6.92E-03 | -0.935 | 6.15E-03 | -0.313 | 3.02E-01 |
| P13639 | EEF2 | 1938 | -1.453 | 4.12E-07 | -1.480 | 1.48E-06 | 0.076 | 9.68E-01 |
| P00739-1 | HPR | 3250 | -1.507 | 3.00E-04 | -0.463 | 8.45E-02 | 0.094 | 1.00E+00 |
| Q5T749 | KPRP | 448834 | -1.519 | 9.11E-07 | -0.727 | 4.98E-04 | -0.261 | 3.86E-01 |
| P07737 | PFN1 | 5216 | -1.564 | 2.46E-03 | 0.189 | 5.25E-01 | 0.003 | 9.34E-01 |
| P15169 | CPN1 | 1369 | -1.590 | 9.03E-03 | -0.924 | 3.17E-02 | -0.351 | 6.00E-01 |
| Q9UQB8-3 | BAIAP2 | 10458 | -1.662 | 2.83E-03 | 0.071 | 1.00E+00 | -0.507 | 1.17E-01 |
| P08319-2 | ADH4 | 127 | -1.663 | 1.22E-03 | -0.408 | 7.93E-02 | -0.469 | 7.26E-03 |
| P04217 | A1BG | 1 | -1.725 | 6.95E-07 | -0.326 | 6.88E-02 | -0.427 | 6.92E-02 |
| P20742 | PZP | 5858 | -1.767 | 3.48E-06 | -0.511 | 8.80E-03 | -0.733 | 7.18E-03 |
| Q9BWP8-9 | COLEC11 | 78989 | -1.793 | 7.23E-03 | -0.250 | 3.39E-01 | -0.659 | 2.52E-03 |
| P55056 | APOC4 | 346 | -1.934 | 5.81E-04 | -0.545 | 5.79E-01 | -1.763 | 7.26E-03 |
| P0CW18 | PRSS56 | 646960 | -1.952 | 9.74E-03 | -0.199 | 4.78E-01 | -0.406 | 5.19E-02 |
| P07359 | GP1BA | 2811 | -2.028 | 1.83E-04 | 0.014 | 9.51E-01 | -0.650 | 4.09E-02 |
| Q86YA3-1 | ZGRF1 | 55345 | -2.112 | 1.57E-03 | -0.012 | 9.75E-01 | -0.612 | 1.32E-02 |
| O75882-1 | ATRN | 8455 | -2.239 | 8.62E-03 | 0.008 | 1.00E+00 | -0.011 | 7.17E-01 |
| P16671 | CD36 | 948 | -2.271 | 7.37E-03 | -0.399 | 2.18E-01 | -0.394 | 2.72E-01 |
| P29144 | TPP2 | 7174 | -2.280 | 2.29E-03 | -2.021 | 1.43E-04 | -1.032 | 6.20E-02 |
| P08174-5 | CD55 | 1604 | -2.363 | 5.26E-04 | -0.873 | 1.03E-01 | 0.320 | 4.83E-01 |
| Q04917 | YWHAH | 7533 | -2.395 | 2.93E-03 | 0.536 | 5.18E-01 | 0.371 | 9.68E-01 |
| P23083 | IGHV1OR15-1 | 388077 | -2.427 | 1.66E-03 | -0.458 | 5.66E-01 | -0.429 | 3.95E-01 |
| Q8N960 | CEP120 | 153241 | -2.454 | 1.77E-07 | -1.364 | 1.31E-01 | -0.216 | 2.82E-01 |
| A0A075B6K4 | IGLV3-10 | 28803 | -2.546 | 7.76E-04 | -0.357 | 2.29E-01 | -0.185 | 6.49E-01 |
| P16070-12 | CD44 | 960 | -2.779 | 6.16E-05 | -0.918 | 1.00E+00 | -0.178 | 3.02E-01 |
| Q8IU54 | IL29 | NA | -2.902 | 6.86E-01 | 8.355 | 2.47E-03 | -2.986 | 3.59E-01 |
| P00441 | SOD1 | 6647 | -3.419 | 9.68E-06 | -0.282 | 1.48E-01 | -0.709 | 4.97E-03 |
| P04430 | IGKV1-16 | 28938 | -3.559 | 4.64E-01 | -7.948 | 1.52E-03 | 0.443 | 4.82E-01 |
| Q01546 | KRT76 | 51350 | -3.864 | 9.88E-07 | -0.276 | 3.40E-01 | -0.135 | 9.68E-01 |
| P62805 | HIST1H4A | NA | -4.041 | 1.10E-02 | -3.072 | 7.92E-02 | -0.775 | 3.95E-01 |
| P00352 | ALDH1A1 | 216 | -4.136 | 5.42E-03 | -10.624 | 1.85E-03 | 2.053 | 7.78E-01 |
| Q8TEC5 | SH3RF2 | 153769 | -4.146 | 3.91E-04 | -0.215 | 5.79E-01 | 0.791 | 3.63E-01 |
| Q9NRY5 | FAM114A2 | 10827 | -4.217 | 7.50E-05 | 0.646 | 9.72E-03 | 0.174 | 8.04E-01 |
| O14950 | MYL12A | 10627 | -4.726 | 4.84E-03 | -0.467 | 4.60E-01 | 1.136 | 1.48E-02 |
| P34932 | HSPA4 | 3308 | -4.982 | 2.88E-06 | -0.060 | 5.25E-01 | -0.696 | 5.74E-02 |
| Q6ZN30 | BNC2 | 54796 | -5.024 | 1.48E-03 | -2.933 | 7.93E-02 | 1.351 | 2.93E-03 |
| Q9NQ79 | CRTAC1 | 55118 | -5.074 | 2.18E-04 | -0.512 | 7.41E-02 | -0.003 | 1.00E+00 |
| P09382 | LGALS1 | 3956 | -5.407 | 2.93E-03 | -4.930 | 1.34E-01 | -0.312 | 9.68E-01 |
| Q96FQ6 | S100A16 | 140576 | -5.420 | 8.18E-03 | -1.372 | 2.80E-01 | 3.372 | 6.79E-01 |
| Q5J8M3-1 | EMC4 | 51234 | -5.508 | 1.50E-04 | -1.220 | 1.29E-03 | -0.392 | 1.29E-01 |
| P01766 | IGHV3-13 | 28449 | -5.560 | 8.92E-03 | -6.157 | 1.23E-01 | -6.308 | 7.12E-02 |
| P48163 | ME1 | 4199 | -5.565 | 4.67E-04 | 1.564 | 4.49E-01 | 2.384 | 3.95E-01 |
| P47929 | LGALS7 | 3963 | -5.733 | 6.33E-03 | -1.301 | 6.92E-02 | -1.127 | 2.65E-01 |
| Q9UP83-3 | COG5 | 10466 | -5.806 | 4.15E-02 | -13.040 | 5.32E-05 | -0.107 | 5.08E-01 |
| Q14134-1 | TRIM29 | 23650 | -6.072 | 1.09E-02 | -6.798 | 4.10E-02 | 0.500 | 8.36E-01 |
| P84095 | RHOG | 391 | -6.233 | 3.88E-03 | -1.632 | 9.26E-01 | 0.622 | 9.04E-02 |
| Q9H3S7 | PTPN23 | 25930 | -6.340 | 3.30E-03 | -4.891 | 5.55E-03 | -0.121 | 5.06E-02 |
| A0A0B4J1X8 | IGHV3-43 | 28426 | -6.402 | 6.00E-03 | -5.283 | 7.79E-02 | -3.656 | 6.27E-02 |
| Q9Y6U3 | SCIN | 85477 | -6.465 | 6.52E-04 | -16.004 | 4.33E-05 | -16.427 | 2.90E-04 |
| P30153 | PPP2R1A | 5518 | -6.499 | 8.87E-05 | -1.944 | 2.30E-01 | -3.412 | 3.60E-03 |
| A0A0C4DH25 | IGKV3D-20 | 28874 | -6.507 | 6.01E-03 | -0.174 | 3.40E-01 | 0.364 | 4.08E-01 |
| P01718 | IGLV3-27 | 28791 | -6.613 | 4.14E-05 | -0.320 | 6.95E-01 | -1.430 | 1.09E-01 |
| P03950 | ANG | 283 | -6.681 | 8.35E-03 | -3.347 | 5.16E-01 | -3.866 | 9.01E-01 |
| P51659-1 | HSD17B4 | 3295 | -6.695 | 6.34E-03 | -2.652 | 6.48E-02 | -1.610 | 7.78E-01 |
| Q86XI8 | ZSWIM9 | 374920 | -6.821 | 5.43E-05 | -0.885 | 8.89E-03 | -0.416 | 8.28E-02 |
| Q9Y613 | FHOD1 | 29109 | -6.826 | 5.70E-03 | -13.662 | 5.26E-05 | -3.137 | 7.56E-02 |
| P08670 | VIM | 7431 | -7.062 | 1.86E-03 | 1.070 | 6.94E-02 | 1.485 | 2.65E-01 |
| A0A075B6R2 | IGHV4-4 | 28401 | -7.235 | 3.54E-04 | -1.195 | 7.41E-03 | -0.553 | 4.09E-02 |
| Q9HCY8 | S100A14 | 57402 | -7.406 | 1.60E-06 | -2.234 | 2.88E-04 | -2.602 | 3.18E-04 |
| P23219 | PTGS1 | 5742 | -7.455 | 3.96E-05 | 0.818 | 3.92E-02 | 0.927 | 3.28E-02 |
| P19971 | TYMP; SCO2 | NA | -7.471 | 6.82E-03 | -13.147 | 4.12E-04 | -0.066 | 9.08E-02 |
| Q92508 | PIEZO1 | 9780 | -7.574 | 1.75E-03 | 1.820 | 5.59E-01 | -1.055 | 2.72E-01 |
| A0A075B6H9 | IGLV4-69 | 28784 | -7.705 | 9.09E-06 | -2.830 | 1.86E-03 | 0.463 | 6.20E-01 |
| Q86VP6-1 | CAND1 | 55832 | -7.749 | 2.15E-04 | -6.137 | 6.74E-03 | -4.211 | 1.32E-02 |
| Q9UI42-1 | CPA4 | 51200 | -8.570 | 4.56E-05 | -1.805 | 7.17E-05 | -0.399 | 3.10E-01 |
| P06748 | NPM1 | 4869 | -8.682 | 1.96E-05 | -0.450 | 3.72E-01 | -0.351 | 3.42E-01 |
| Q9NP80-1 | PNPLA8 | 50640 | -8.787 | 1.62E-03 | -1.704 | 6.51E-01 | -8.138 | 1.84E-02 |
| P23284 | PPIB | 5479 | -9.112 | 1.64E-03 | -1.660 | 2.88E-01 | 0.231 | 4.82E-01 |
| P28066-1 | PSMA5 | 5686 | -9.770 | 5.26E-05 | -0.333 | 7.12E-01 | -0.657 | 3.89E-02 |
| Q9NUD7 | C20orf96 | 140680 | -10.254 | 8.83E-03 | -1.131 | 1.34E-01 | 0.027 | 8.40E-01 |
| P01706 | IGLV2-11 | 28816 | -10.256 | 3.84E-04 | -0.330 | 1.90E-01 | -0.354 | 5.45E-01 |
| Q68DK7 | MSL1 | 339287 | -10.572 | 9.78E-04 | -0.152 | 6.51E-01 | 0.166 | 3.95E-01 |
| Q9H444 | CHMP4B | 128866 | -10.596 | 8.05E-03 | 2.579 | 2.42E-01 | 2.978 | 4.30E-02 |
| Q96M02-1 | C10orf90 | 118611 | -11.335 | 1.47E-04 | -11.011 | 1.12E-03 | -7.923 | 5.65E-03 |
| A0A075B6I9 | IGLV7-46 | 28775 | -12.198 | 4.96E-05 | -0.589 | 8.78E-01 | -0.179 | 8.36E-01 |
| Q14574-2 | DSC3 | 1825 | -12.866 | 2.85E-06 | -17.043 | 8.53E-05 | -0.965 | 2.03E-02 |
| Q8N0S2-1 | SYCE1 | 93426 | -13.047 | 2.28E-07 | -6.615 | 2.35E-03 | 1.318 | 1.29E-01 |
| P01704 | IGLV2-14 | 28815 | -13.369 | 2.74E-07 | -1.706 | 2.42E-01 | -0.734 | 5.06E-02 |
| Q9C075 | KRT23 | 25984 | -13.400 | 4.42E-08 | -12.925 | 2.87E-03 | -0.380 | 3.28E-02 |
| Q6DHV5 | CC2D2B | 387707 | -13.603 | 1.08E-07 | -0.124 | 3.40E-01 | -0.142 | 1.86E-01 |
| Legend: List of 207 EV proteins used for principal component analysis. These proteins are significantly altered in at least one of the pancreas diseases compared to controls. | | | | | | | | |
