## Supplementary Table 6 for "A Large-Scale Proteomics Resource of Circulating Extracellular Vesicles for Biomarker Discovery in Pancreatic Cancer"

**Supplementary Table 6. List of EV proteins that are significantly alter metastatic versus non-metastatic diseases**

| ProteinID | GeneName | log2 Fold Change* | pValue |
|---|---|---|---|
| Q7Z5J8-1 | ANKAR | 9.46 | 3.06E-03 |
| P00480 | OTC | 8.73 | 8.00E-03 |
| Q92820 | GGH | 8.31 | 6.43E-03 |
| Q86W25 | NLRP13 | 8.12 | 8.06E-03 |
| Q5IJ48 | CRB2 | 7.56 | 8.00E-03 |
| Q8IZ83 | ALDH16A1 | 6.50 | 1.76E-02 |
| P35900 | KRT20 | 6.44 | 2.19E-02 |
| P07738 | BPGM | 6.21 | 2.19E-02 |
| P28070 | PSMB4 | 6.06 | 1.42E-02 |
| Q69YZ2 | TMEM200B | 6.00 | 2.90E-02 |
| Q6UB99 | ANKRD11 | 5.95 | 2.81E-02 |
| O14523 | C2CD2L | 5.94 | 1.78E-02 |
| P28906 | CD34 | 5.84 | 4.72E-02 |
| P98088 | MUC5AC | 5.74 | 4.88E-02 |
| Q13277-1 | STX3 | 5.69 | 2.54E-02 |
| Q8NI99 | ANGPTL6 | 5.62 | 8.00E-03 |
| Q9Y230 | RUVBL2 | 5.59 | 1.21E-02 |
| Q5JXB2 | UBE2NL | 5.33 | 7.02E-03 |
| P31025 | LCN1 | 5.29 | 4.65E-02 |
| P10619 | CTSA | 5.04 | 6.06E-03 |
| O60664-1 | PLIN3 | 4.69 | 4.88E-02 |
| Q9P0V3-1 | SH3BP4 | 4.63 | 2.80E-02 |
| P03973 | SLPI | 4.52 | 4.88E-02 |
| Q13296-1 | SCGB2A2 | 4.51 | 4.95E-03 |
| Q12913-1 | PTPRJ | 4.03 | 3.30E-02 |
| O75223-1 | GGCT | 3.46 | 3.06E-03 |
| O60832-1 | DKC1 | 3.17 | 4.72E-02 |
| Q6E0U4-6 | DMKN | 3.09 | 5.06E-03 |
| P48643 | CCT5 | 3.07 | 4.88E-02 |
| P13798 | APEH | 2.84 | 1.63E-02 |
| Q15485-1 | FCN2 | 2.44 | 5.71E-03 |
| O95810 | SDPR | 1.96 | 4.72E-02 |
| P25786-1 | PSMA1 | 1.93 | 5.71E-03 |
| P0DOY2 | IGLC2 | 1.90 | 1.76E-02 |
| P15311 | EZR | 1.72 | 6.60E-03 |
| Q7L576-1 | CYFIP1 | 1.59 | 3.70E-02 |
| O00294-2 | TULP1 | 1.35 | 3.06E-03 |
| Q96L93-1 | KIF16B | 1.28 | 3.30E-02 |

| | | | |
|---|---|---|---|
| P04070-2 | PROC | 1.25 | 6.60E-03 |
| Q8WWZ8-1 | OIT3 | 1.17 | 6.69E-03 |
| P07195 | LDHB | 1.13 | 4.03E-02 |
| Q08830 | FGL1 | 1.07 | 4.98E-02 |
| O00592-2 | PODXL | 1.05 | 1.76E-02 |
| Q8WUA8 | TSKU | -1.01 | 7.96E-03 |
| P07996 | THBS1 | -1.03 | 2.90E-03 |
| P78371-1 | CCT2 | -1.06 | 3.86E-02 |
| P04196 | HRG | -1.07 | 2.52E-03 |
| P08575-2 | PTPRC | -1.11 | 5.71E-03 |
| P61626 | LYZ | -1.19 | 7.85E-03 |
| Q9NQ36-1 | SCUBE2 | -1.21 | 1.58E-02 |
| P23229-6 | ITGA6 | -1.23 | 3.06E-03 |
| P11226 | MBL2 | -1.27 | 1.05E-02 |
| Q6ICB0 | DESI1 | -1.28 | 6.06E-03 |
| Q13200 | PSMD2 | -1.36 | 3.30E-02 |
| P08174-5 | CD55 | -1.37 | 1.76E-02 |
| P13987 | CD59 | -1.38 | 8.24E-03 |
| P50991 | CCT4 | -1.48 | 4.32E-02 |
| P08319-2 | ADH4 | -1.52 | 3.75E-02 |
| Q8NF91-7 | SYNE1 | -1.53 | 3.54E-03 |
| P07359 | GP1BA | -1.63 | 5.71E-03 |
| P27918 | CFP | -1.82 | 4.33E-03 |
| Q04917 | YWHAH | -1.98 | 6.60E-03 |
| Q86YA3-1 | ZGRF1 | -2.01 | 9.06E-03 |
| P16671 | CD36 | -2.02 | 1.63E-02 |
| Q6AHZ1-1 | ZNF518A | -2.17 | 5.71E-03 |
| P61981 | YWHAG | -2.17 | 1.42E-02 |
| O14950 | MYL12A | -2.33 | 3.13E-02 |
| A0A075B6K4 | IGLV3-10 | -2.49 | 6.53E-03 |
| Q8TEC5 | SH3RF2 | -2.76 | 2.66E-02 |
| P11169 | SLC2A3 | -2.82 | 6.53E-03 |
| Q86TH1 | ADAMTSL2 | -3.11 | 7.96E-03 |
| P61225 | RAP2B | -3.42 | 2.66E-02 |
| Q7Z745-1 | HEATR7B2 | -3.45 | 3.30E-02 |
| P34932 | HSPA4 | -3.49 | 7.85E-03 |
| A0A0J9YX35 | IGHV3-64D | -3.57 | 1.08E-02 |
| Q9NRY5 | FAM114A2 | -4.05 | 5.49E-03 |
| P23284 | PPIB | -4.78 | 2.29E-02 |
| A0A0C4DH25 | IGKV3D-20 | -4.89 | 1.63E-02 |
| P54920 | NAPA | -4.90 | 3.23E-02 |
| P17900 | GM2A | -5.75 | 4.72E-02 |
| Q9HCH0 | NCKAP5L | -6.06 | 1.56E-02 |

| | | | |
|---|---|---|---|
| O15440 | ABCC5 | -6.22 | 1.76E-02 |
| P01704 | IGLV2-14 | -6.38 | 2.80E-02 |
| A0A075B6H9 | IGLV4-69 | -7.06 | 5.71E-03 |
| Q8IW75 | SERPINA12 | -7.20 | 4.28E-03 |

*positive value represents over expression in metastatic group compared to non-metas

**ed in patients with**

static group
