## Supplementary Table 7 for "A Large-Scale Proteomics Resource of Circulating Extracellular Vesicles for Biomarker Discovery in Pancreatic Cancer"

| **Supplementary Table 7. Support Vector Machine Prediction of 16 individual genes in Discovery Test and Validation Cohorts** | | | | | | | | |
| --- | --- | --- | --- | --- | --- | --- | --- | --- |
| **GeneName** | **Discovery Test** | | | | **Validation Cohort** | | | |
|  | **Sensitivity** | **Specificity** | **Precision** | **AUC** | **Sensitivity** | **Specificity** | **Precision** | **AUC** |
| PDCD6IP | 0.89 | 0.83 | 0.96 | 0.93 | 0.92 | 0.50 | 0.79 | 0.86 |
| RUVBL2 | 1.00 | 0.00 | 0.82 | 0.82 | 1.00 | 0.00 | 0.67 | 0.86 |
| CCT4 | 1.00 | 0.00 | 0.82 | 0.89 | 1.00 | 0.00 | 0.67 | 0.82 |
| ATP5F1 | 1.00 | 0.00 | 0.82 | 0.71 | 1.00 | 0.00 | 0.67 | 0.76 |
| PSME1 | 1.00 | 0.00 | 0.82 | 0.82 | 1.00 | 0.00 | 0.67 | 0.69 |
| PFKL | 1.00 | 0.00 | 0.82 | 0.93 | 1.00 | 0.00 | 0.67 | 0.68 |
| CTSA | 1.00 | 0.00 | 0.82 | 0.56 | 1.00 | 0.00 | 0.67 | 0.68 |
| DLD | 1.00 | 0.00 | 0.82 | 0.56 | 1.00 | 0.00 | 0.67 | 0.68 |
| GGH | 1.00 | 0.00 | 0.82 | 0.56 | 1.00 | 0.00 | 0.67 | 0.68 |
| KRT20 | 1.00 | 0.00 | 0.82 | 0.56 | 1.00 | 0.00 | 0.67 | 0.68 |
| SCIN | 1.00 | 0.17 | 0.84 | 0.17 | 0.54 | 0.33 | 0.62 | 0.55 |
| SERPINA12 | 1.00 | 0.00 | 0.82 | 0.17 | 1.00 | 0.00 | 0.67 | 0.55 |
| ATP5H | 1.00 | 0.00 | 0.82 | 0.71 | 1.00 | 0.00 | 0.67 | 0.51 |
| SLPI | 1.00 | 0.00 | 0.82 | 0.67 | 1.00 | 0.00 | 0.67 | 0.49 |
| LASP1 | 0.89 | 0.00 | 0.80 | 0.74 | 0.54 | 0.33 | 0.62 | 0.48 |
| WFDC12 | 1.00 | 0.00 | 0.82 | 0.79 | 1.00 | 0.00 | 0.67 | 0.44 |

| **SVM weights of individual markers in 7-Protein Biomarker** | |
| --- | --- |
| **Gene Name** | **Weights** |
| SERPINA12 | 5.77 |
| KRT20 | 5.47 |
| RUVBL2 | 5.36 |
| PDCD6IP | 4.84 |
| CCT4 | 4.41 |
| DLD | 4.24 |
| ATP5F1 | 1.84 |
| SVM model prediction based on the 7Gene Biomarker | |

| **SVM model prediction based on the 7-Protein Biomarker** | | | | | | | | | |
| --- | --- | --- | --- | --- | --- | --- | --- | --- | --- |
| **Prediction in Discovery Test Cohort** | | | |  |  | **Prediction in Validation Cohort** | | | |
|  |  | **PREDICTED** | |  |  |  |  | **PREDICTED** | |
|  |  | **Benign** | **PDAC** |  |  |  |  | **Benign** | **PDAC** |
| **TRUE** | **Benign** | 6 | 0 |  |  | **TRUE** | **Benign** | 5 | 7 |
|  | **PDAC** | 0 | 27 |  |  |  | **PDAC** | 0 | 24 |
|  | Sensitivity = 1 | |  |  |  |  | Sensitivity = 1 | |  |
|  | Specificity = 1 | |  |  |  |  | Specificity = 0.42 | |  |
|  | Precision = 1 | |  |  |  |  | Precision = 0.77 | |  |
|  | Predicted AUC = 1 | | |  |  |  | Predicted AUC = 0.89 | |  |
