## Supplementary Table 8 for "A Large-Scale Proteomics Resource of Circulating Extracellular Vesicles for Biomarker Discovery in Pancreatic Cancer"

**Supplementary Table 8. List of 25 cEV proteins that met the eligibility criteria for validation studies.**

ARHGEF12
ATP5F1
ATP5H
CAST
CCT4
COL12A1
CRB2
CTSA
DLD
GGH
KRT20
LASP1
MAPRE2
OAF
PDCD6IP
PDIA6
PFKL
PLSCR1
PSME1
RUVBL2
SCIN
SERPINA12
SLPI
STX7
WFDC12
