## Supplementary Table 9 for "A Large-Scale Proteomics Resource of Circulating Extracellular Vesicles for Biomarker Discovery in Pancreatic Cancer"

| **Supplementary Table 9. Baseline characteristics of patients enrolled in the validation cohort.** | | | | |  |
| --- | --- | --- | --- | --- | --- |
|  | **Pancreatic Cancer** | | | **Chronic Pancreatitis** | **IPMN** |
|  | **I-II** | **III-IV** | **All Stages** |  |  |
| Number of subjects | 9 | 15 | 24 | 6 | 6 |
| Age (mean, range) | 70 (59-78) | 63.5 (35-87) | 66 (35-87) | 67.5 (45-86) | 67.3 (37-84) |
| Gender (F) | 44.4% | 60.0% | 54.1% | 16.6% | 50.0% |
| Baseline CA19-9 (mean, range) | 650 (3 -1,969) | 22,849 (3 -145,608) | 13,768 (3 -145,608) | - | - |
